# Transmission of COVID-19 in the state of Georgia, United States: Spatiotemporal variation and impact of social distancing

**DOI:** 10.1101/2020.10.22.20217661

**Authors:** Yuke Wang, Casey Siesel, Yangping Chen, Ben Lopman, Laura Edison, Michael Thomas, Carly Adams, Max Lau, Peter F.M. Teunis

**Affiliations:** Center for Global Safe WASH, Hubert Department of Global Health, Rollins School of Public Health, Emory University, Atlanta, Georgia 30322, USA; The Department of Epidemiology, Rollins School of Public Health, Emory University, Atlanta, Georgia 30322, USA; Centers for Disease Control and Prevention, Division of State and Local Readiness, Atlanta, Georgia 30322, USA; Georgia Department of Public Health, Atlanta, GA 30303; Department of Biostatistics and Bioinformatics, Rollins School of Public Health, Emory University, Atlanta, Georgia 30322, USA

**Keywords:** novel coronavirus disease, COVID-19, transmission, serial interval, reproduction number, shelter-in-place, Georgia

## Abstract

**Background:** Beginning in early February 2020, COVID-19 spread across the state of Georgia, leading to 258,354 cumulative cases as of August 25, 2020. The time scale of spreading (i.e., serial interval) and magnitude of spreading (i.e., *R*_*t*_ or reproduction number) for COVID-19, were observed to be heterogenous by demographic characteristics, region and time period. In this study, we examined the COVID-19 transmission in the state of Georgia, United States.

**Methods:** During February 1 to July 13, 2020, we identified 4080 transmission pairs using contact information from reports of COVID-19 cases from the Georgia Department of Public Health. We examined how various transmission characteristics were affected by disease symptoms, demographics (age, gender, and race), and time period (during shelter-in-place and after reopening). In addition, we estimated the time course of reproduction numbers during early February–mid-June for all 159 counties in the state of Georgia, using a total of 118,491 reported COVID-19 cases.

**Findings:** Over this period, the serial interval appeared to decrease from 5.97 days in February–April to 4.40 days in June–July. With regard to age, transmission was assortative and patterns of transmission changed over time. COVID-19 mainly spread from adults to all age groups; transmission among and between children and the elderly was found less frequently. Younger adults (20– 50 years old) were involved in the majority of transmissions occurring during or after reopening subsequent to the shelter-in-place period. By mid-July, two waves of COVID-19 transmission were apparent, separated by the shelter-in-place period in the state of Georgia. Counties around major cities and along interstate highways had more intense transmission.

**Interpretation:** The transmission of COVID-19 in the state of Georgia had been heterogeneous by area and changed over time. The shelter-in-place was not long enough to sufficiently suppress COVID-19 transmission in densely populated urban areas connected by major transportation links. Studying local transmission patterns may help in predicting and guiding states in prevention and control of COVID-19 according to population and region.

**Funding:** Emory COVID-19 Response Collaborative.

**Research in context:** 

**Evidence before this study:** The ongoing COVID-19 pandemic has caused 37,109,581 cases and 1,070,355 deaths worldwide as of October 11, 2020. We searched PubMed for articles published on and before October 11, 2020 using keywords “novel coronavirus”, “SARS–nCoV–2”, “COVID-19”, “transmission”, “serial interval”, “reproduction number”, and “shelter-in-place”. Few published studies have estimated the serial interval but no study was found that examined the time-varying serial interval. Few studies have examined the transmission patterns between groups with different characteristics. And no study has examined the timevarying reproduction number for COVID-19 and impact of shelter-in-place order at the county level in the United States.

**Added value of this study:** To our knowledge, this is the first study showing the multiple aspects of COVID-19 transmission, including serial interval, transmission patterns between age, gender, or race groups, and spatiotemporal patterns, based on data from 118,491 confirmed COVID-19 cases and 4080 tracked pairs of infector and infectee. We found that during February–July the serial interval for symptom onset shortened, and the major contribution to the spread of COVID-19 shifted to younger ages (from 40–70 years old in February–April to 20–50 years old in June–July). We also found three to four weeks of the shelter-in-place slowed transmission but was insufficient to prevent transmission into urban and peri-urban counties connected with major transportation.

**Implications of all the available evidence:** The contracting serial intervals and increasing spread by younger generation show the COVID-19 transmission at county level changes over time. The spatiotemporal patterns of transmission in county level further provide important evidence to guide effective COVID-19 prevention and control measures (e.g., shelter-in-place) in different areas.

## Introduction

Coronavirus disease 2019 (COVID-19) is an infectious disease caused by the severe acute respiratory syndrome coronavirus 2 (SARS-CoV-2). After the disease was first reported in Wuhan, China in December 2019, COVID-19 spread rapidly across the world to its currently ongoing global pandemic. As of October 22, 2020, the United States had the most COVID-19 confirmed cases (8,338,467) and deaths (222,220) globally [1], and the state of Georgia had reported 343,750 confirmed cases and 7704 deaths [2].

Transmission of COVID-19 has been shown to vary by region (states and counties), setting (long-term care facilities, prisons, churches, and factories), population demographics (age, sex, and race), and even among individuals (by physiologic and behavioral differences). During the early phases of transmission in the United States, new cases were mainly imported by travelers and transmission was associated with human mobility [3]. Local transmission was more intense in regions with high population density and in populations with frequent social contacts [4, 5, 6]. When SARS-CoV-2 was introduced into high-risk settings such as long-term care facilities, transmission rates were observed to be high and the outcomes were often fatal [7].

To study transmission of COVID-19, two important characteristics, the time scale and magnitude of spreading can be defined. The time scale for spreading of infectious disease can be expressed by the serial interval for onset of symptoms defined as the time interval between a case-patient experiencing symptoms and a descendant case-patient, infected by the first case-patient, becoming symptomatic. The magnitude of spreading can be expressed as the reproduction number *R*_*t*_: the expected number of cases directly caused by any single infectious person. *R*_*t*_ has been shown to vary strongly with some case-patients leading to super-spreading events [8, 9]. Such heterogeneity influences the spread as well as the control of COVID-19, as documented by studies of non-pharmaceutical interventions in China [10, 11] and Europe [12] at country and province level.

After the first case of COVID-19 was reported in the state of Georgia on March 2, 2020, a series of transmission events and interventions occurred (Table S1). On March 14, a cumulative total of 1712 cases had been confirmed in Georgia and the governor declared a public health emergency. On March 24, large gatherings were banned, and on April 3 a shelter-in-place order was announced. Approximately three weeks later beginning on April 24, Georgia became the first U.S. state that allowed some businesses to reopen, and on April 30 the shelter-in-place order was lifted. On June 1, limits on the size of public gatherings were relaxed, bars and nightclubs could reopen, sport events could resume, and summer schools and camps were allowed to begin sessions [13]. During June–July 2020, as new COVID-19 cases continued to surge in Georgia and other states, it was critical to understand how shelter-in-place and the subsequent reopening events impacted the transmission of COVID-19 in different regions.

In this study, we examined the characteristics of COVID-19 transmission in the state of Georgia by estimating serial intervals and *R*_*t*_ using epidemiological data based on confirmed cases. Using these data, we sought to characterize the spread of COVID-19 among various subpopulations. Identifying a large number of linked cases, including the infector and infectee, enabled us to estimate the distribution of the serial interval for COVID-19 symptoms. Using the serial interval distribution, we can estimate the time-varying effective reproduction number (*R*_*t*_) using the methods developed by Wallinga and Teunis [14, 15]. With *R*_*t*_s over time, we can study the spatial distribution of transmission across all 159 Georgia counties, as well as the effects of shelter-in-place and subsequent gradual reopening.

## Methods

### Data Source

The Georgia Department of Public Health (GDPH) provided data from 118,491 confirmed COVID-19 cases in all 159 counties during February 1–July 13, 2020. Available data included demographic characteristics (age, sex, and race), clinical characteristics (dates of symptom onset, recorded symptoms, hospitalization, and ventilator usage), and social contacts (contact with confirmed cases and participant in a confirmed cluster) (Table 1). With regard to events possibly driving transmission, time periods were categorized as early transmission and shelter-in-place (February to April), after reopening (May), and further reopening (June to July). In this study, a COVID-19 case is defined as laboratory confirmation of SARS-CoV-2 infection by reverse transcriptase polymerase chain reaction (RT-PCR) in a person irrespective of clinical signs and symptoms.

**Table 1:** Descriptive statistics of demographic and clinical information for people with confirmed COVID-19 cases in the state of Georgia in three time periods (February–April, May, and June–July 2020).

|  |  | February–April | May | June–July | Total |
| --- | --- | --- | --- | --- | --- |
| Total | N | 31,575 | 19,270 | 67,646 | 118,491 |
| Age | median (Q1-Q3) | 51 (37-65) | 43 (28-59) | 34 (23-50) | 40 (26-56) |
| Sex | Male | 13,770 (43.6%) | 9,142 (47.4%) | 30,247 (44.7%) | 53,159 (44.9%) |
|  | Female | 17,308 (54.8%) | 9,747 (50.6%) | 33,828 (50%) | 60,883 (51.4%) |
|  | Missing | 497 (1.6%) | 381 (2.0%) | 3,571 (5.3%) | 4,449 (3.7%) |
| Race | Black | 13,010 (41.2%) | 4,639 (24.1%) | 13,878 (20.5%) | 31,527 (26.6%) |
|  | White | 11,418 (36.2%) | 7,168 (37.2%) | 17,500 (25.9%) | 36,086 (30.5%) |
|  | Other | 2,818 (8.9%) | 2,029 (10.5%) | 6,158 (9.1%) | 11,005 (9.3%) |
|  | Missing | 4,329 (13.7%) | 5,434 (28.2%) | 30,110 (44.5%) | 39,873 (33.6%) |
| Hospitalized | Yes | 6,714 (21.3%) | 2,099 (10.9%) | 4,523 (6.7%) | 13,336 (11.3%) |
|  | No | 15,627 (49.5%) | 10,729 (55.7%) | 27,926 (41.3%) | 54,282 (45.8%) |
|  | Missing | 9,234 (29.2%) | 6,442 (33.4%) | 35,197 (52%) | 50,873 (42.9%) |
| Ventilator use | Yes | 1,046 (3.3%) | 184 (1.0%) | 258 (0.4%) | 1,488 (1.3%) |
|  | No | 12,188 (38.6%) | 8,404 (43.6%) | 19,313 (28.6%) | 39,905 (33.7%) |
|  | Missing | 18,341 (58.1%) | 10,682 (55.4%) | 48,075 (71.1%) | 77,098 (65.0%) |
| Abnormal Chest X-ray | Yes | 2,602 (8.2%) | 494 (2.6%) | 742 (1.1%) | 3,838 (3.2%) |
|  | No | 10,151 (32.1%) | 8,081 (41.9%) | 18,246 (27.0%) | 36,478 (30.8%) |
|  | Missing | 18,822 (59.6%) | 10,695 (55.5%) | 48,658 (71.9%) | 78,175 (66.0%) |
| Death | Yes | 2,127 (6.7%) | 558 (2.9%) | 320 (0.5%) | 3,005 (2.5%) |
|  | No | 15,766 (49.9%) | 10,183 (52.8%) | 26,304 (38.9%) | 52,253 (44.1%) |
|  | Missing | 13,682 (43.3%) | 8,529 (44.3%) | 41,022 (60.6%) | 63,233 (53.4%) |
| Fever | Yes | 10,094 (32.0%) | 4,005 (20.8%) | 11,787 (17.4%) | 25,886 (21.8%) |
|  | No | 8,489 (26.9%) | 7,951 (41.3%) | 19,655 (29.1%) | 36,095 (30.5%) |
|  | Missing | 12,992 (41.1%) | 7,314 (38%) | 36,204 (53.5%) | 56,510 (47.7%) |
| Cough | Yes | 12,417 (39.3%) | 4,992 (25.9%) | 15,319 (22.6%) | 32,728 (27.6%) |
|  | No | 6,462 (20.5%) | 7,059 (36.6%) | 16,434 (24.3%) | 29,955 (25.3%) |
|  | Missing | 12,696 (40.2%) | 7,219 (37.5%) | 35,893 (53.1%) | 55,808 (47.1%) |
| Short of Breath | Yes | 8,504 (26.9%) | 2,952 (15.3%) | 7,325 (10.8%) | 18,781 (15.9%) |
|  | No | 9,807 (31.1%) | 8,960 (46.5%) | 23,542 (34.8%) | 42,309 (35.7%) |
|  | Missing | 13,264 (42%) | 7,358 (38.2%) | 36,779 (54.4%) | 57,401 (48.4%) |
| Diarrhea | Yes | 4,410 (14%) | 1,971 (10.2%) | 6,072 (9.0%) | 12,453 (10.5%) |
|  | No | 12,718 (40.3%) | 9,589 (49.8%) | 23,936 (35.4%) | 46,243 (39.0%) |
|  | Missing | 14,447 (45.8%) | 7,710 (40.0%) | 37,638 (55.6%) | 59,795 (50.5%) |

### Tracked Pairs: Serial Intervals and Characteristics of Transmission

Based on reported contacts with confirmed cases, pairs of primary case-patients (infectors) and their secondary case-patients (infectees) were identified, assuming the symptom onset of any infector occurred before the symptom onset of his/her infectees. The serial interval for symptom onset was defined as the number of days between symptom onset for a primary case and an associated secondary case. Thus, serial intervals were assumed to be always positive. Transmission pairs with serial intervals longer than 15 days were dropped as such long intervals are unlikely, as shown in previous studies [16, 17]. We modeled the serial interval as a gamma distribution and maximum likelihood estimators of shape and scale parameters were obtained. Furthermore, we explored whether the duration of the serial interval varied by demographic characteristics, various disease symptoms, and time periods. For example, do infectious cases with specific symptoms (e.g., coughing) lead to a shorter serial interval when infecting a descendant case compared to cases without those specific symptoms? The large numbers of linked cases enabled us to examine the variation in transmission within and between different groups by age, sex, and race.

### Confirmed Cases: Reproduction Numbers

We estimated probabilities of transmission between any pair of case-patients in an outbreak, as proposed by Teunis et al [15]. For an outbreak with *n* observed case-patients, a transmission probability matrix 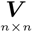 can be defined with any element *v*_*i,j*_ representing the probability that case-patient *i* was infected by case-patient *j*. When two cases are linked by their serial interval, the likelihood of transmission between these two case-patients can be calculated using the serial interval distribution as a kernel density [15]. Additional information at an individual level (e.g., evidence of social contact between case-patients *i* and *j*) is accounted for by a *n* × *n* weighting matrix [15]. The transmission probability matrix ***V*** can be estimated in a Markov chain Monte Carlo procedure [15, 4].

When the transmission probability matrix is known, it can be used to calculate reproduction numbers. Elements of row *i* show the probabilities of case-patient *i* having received their infection from any other case-patient in the observed population. Rows of ***V*** must therefore add to 1. Likewise, elements of column *j* show the probabilities that case-patient *j* has transmitted their infection to any other case-patient in the observed population. Columns of ***V*** therefore add to an estimate of the number of cases infected by case-patient *j*: its reproduction number.

Using GDPH data on cases confirmed with COVID-19 during February 1– July 13, 2020, we estimated effective reproduction numbers (*R*_*t*_) by date, using dates of symptoms onset and social contact information (wherever available) in each county independently by estimating the transmission probability matrix. Among 118,491 confirmed cases, 48,887 (41.3%) had a missing date of symptom onset. These missing symptom onset dates were imputed based on dates of first specimen collection when available, or dates of laboratory report otherwise. The number of days between symptom onset and date of first specimen collected (or date of laboratory report) was modeled using negative binomial regression with the date of first specimen collected (or date of laboratory report) as the predictor.

Estimation of *R*_*t*_ approaching present date (i.e., nowcasting) was not feasible because not all contributing cases had become symptomatic and been reported. To avoid this right censoring issue, the most recent four weeks (June 16–July 13) were removed from the analysis. It is likely to have multiple waves of COVID-19 transmission in the state of Georgia with considerable variation among Georgia counties. The time-varying average *R*_*t*_ estimations were smoothed using LOESS regression and local maximum/minimum were identified for each individual county.

We defined five transmission pattern categories based on reviewing the epidemic curves and *R*_*t*_ curves. Transmission patterns for individual counties were categorized as: consistent spreading, two strong waves, strong first wave, strong second wave, and small case number. Maps were generated to spatially examine the spreading of COVID-19 first wave and transmission patterns. The impact of shelter-in-place, reopening, and further reopening were evaluated by the trend of reproduction numbers before and after those events in different regions in the state of Georgia.

The GDPH Institutional Review Board has determined that this analysis is exempt from the requirement for IRB review and approval and informed consent was not required. This activity was reviewed by the Centers for Disease Control and Prevention (CDC) and was conducted consistent with applicable federal law and CDC policy^6^.

### Role of the funding source

The funder of this study had no role in study design, data collection, data analysis, data interpretation, or writing the report. All authors had full access to all the data in the study, and the corresponding author had final responsibility for the decision to submit for publication.

## Results

### Tracked Pairs: Serial Interval

Based on 4080 tracked pairs of primary case-patients (infectors) and their secondary case-patients (infectees) in the state of Georgia (Table S2), the serial interval distribution in Georgia was estimated as a gamma distribution with shape parameter 2.00 and scale parameter 2.49. Table S3 shows the variation in serial interval distribution by subgroup, for clinical and demographic categories. Generally, the serial interval was longer when primary case-patients had severe clinical outcomes, such as hospitalization, undergoing ventilation, having an abnormal chest X-ray result, or death. Specific symptoms in the primary cases including fever, cough, shortness of breath, or diarrhea did not shorten serial intervals. Serial intervals were not differentiated across demographic categories including age, sex, race, or location. Figure 1 shows estimated serial interval distributions for the three different time periods (February–April, May, and June–July). The average serial interval became shorter over time: from 5.97 days in February–April, to 5.03 days in May, and then to 4.40 days in June–July.

**Figure 1:**
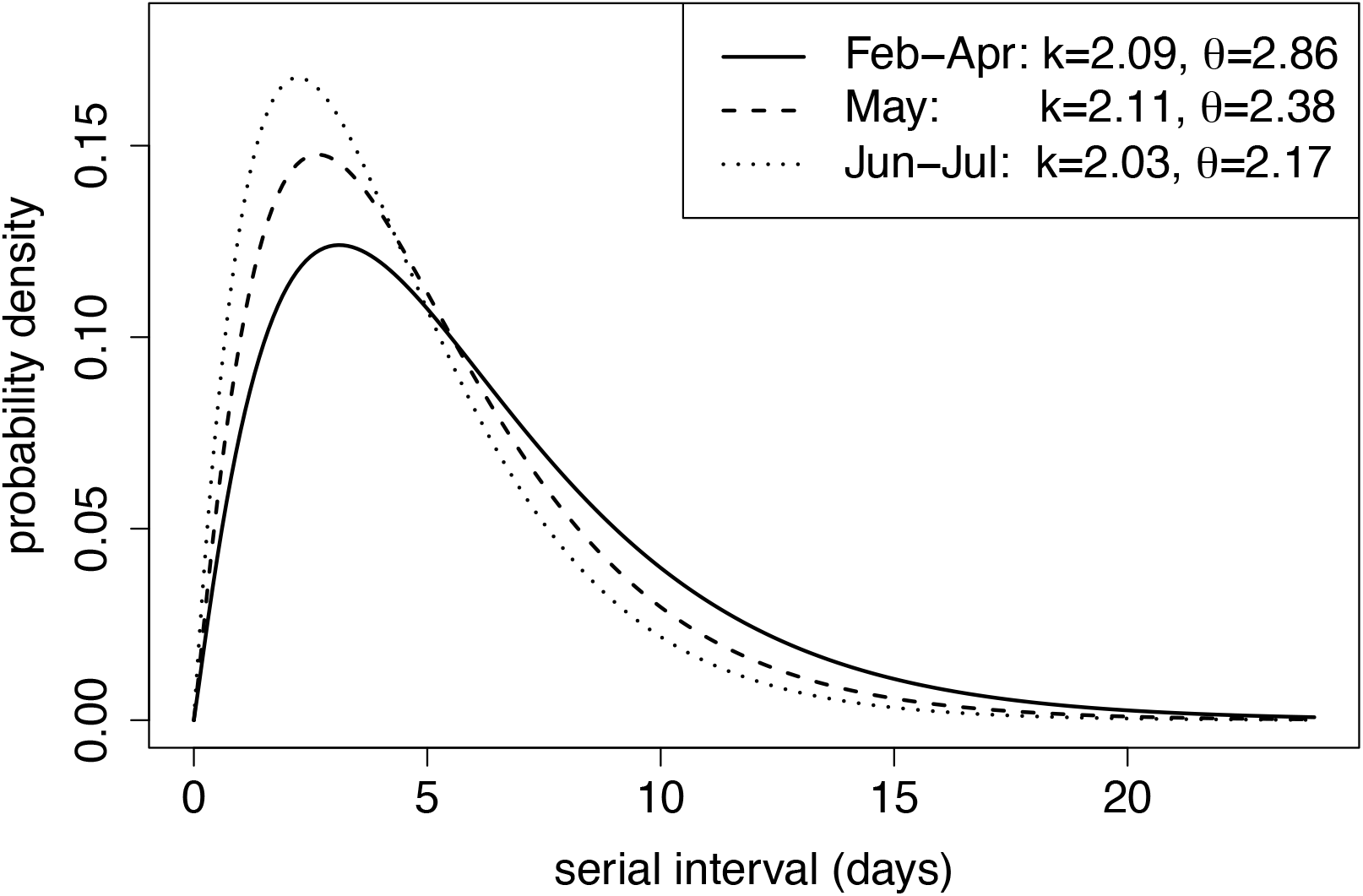
The estimated serial interval distribution for three time periods: Early transmission and shelter-in-place (Feb-Apr); after reopening (May); further reopening (Jun-Jul). *k* and *θ* are the scale and shape parameters for the gamma distribution. The y-axis represents the probability density of having certain serial interval. The unit of probability density is one per unit of time.

### Tracked Pairs: Characteristics of Transmission

To study the variation in transmission by age, the frequencies of tracked links can be shown in an age matrix. Figure 2(a)-2(b) show the disease transmission within and between age, sex, and race groups. Males were twice as likely to infect a female as to infect a male, while females were equally as likely to infect a male or a female. Transmission between races was strongly assortative. Compared with people of other races, white people were 4.4 times as likely to infect white people and black people were 5.6 times as likely to infect black people.

**Figure 2:**
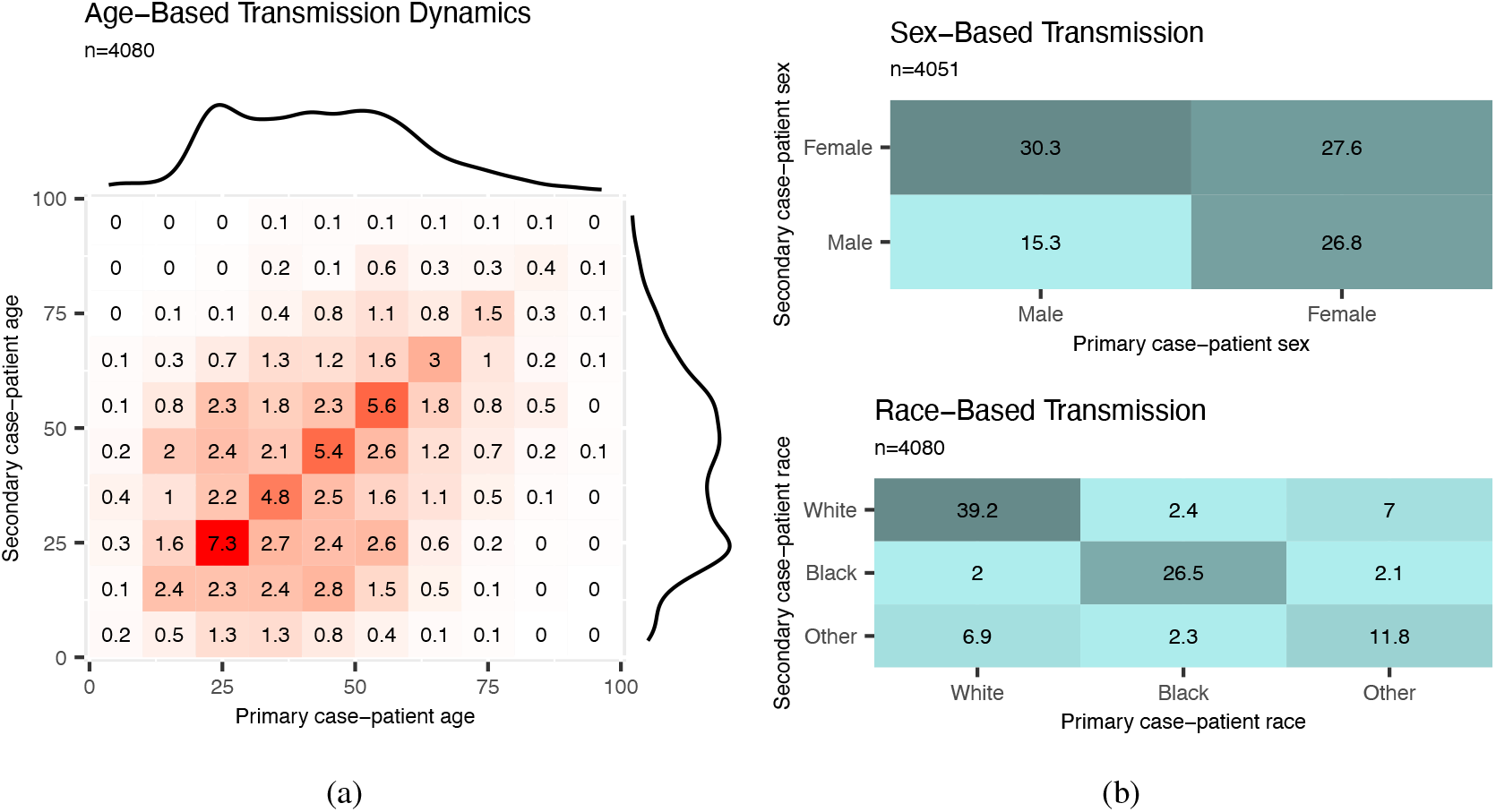
Patterns of COVID-19 disease transmission by age, sex, and race based on 4080 tracked pairs of COVID-19 cases from Georgia, U.S. over the time period of February–July 2020. The matrix graphs show numbers of transmission pairs as a percentage of the total, with primary case-patients (infectors) as columns and their secondary case-patients (infectees) as rows. Darker colors indicate a higher percentages of fraction of tracked pairs observed. Marginal totals are shown as density curves to illustrate the age distribution of case-patients in (a).

Based on these data, COVID-19 seemed to mainly spread from adults aged 20–60 years during February– July; transmission among and between children (*<*20 years old) and the elderly (*>*60 years old) was observed less often, suggesting that transmission occurred more frequently between people of similar ages. Figure 3(a)-3(d) further shows the transmission by age given sex of primary case-patient and secondary case-patient. Transmission between sexes happened mainly within the same age group. Cases in persons aged 10–30 years were associated with the majority of transmission pairs of the same sex. Over the study period, the majority of transmission pairs shifted from ages 40–70 years (median age for primary case-patients was 52 years, and for secondary case-patients was 50 years) in February–April to 20–50 years old (median age for primary case-patients was 36 years, and for secondary case-patients was 34 years) in June–July (Figure 4).

**Figure 3:**
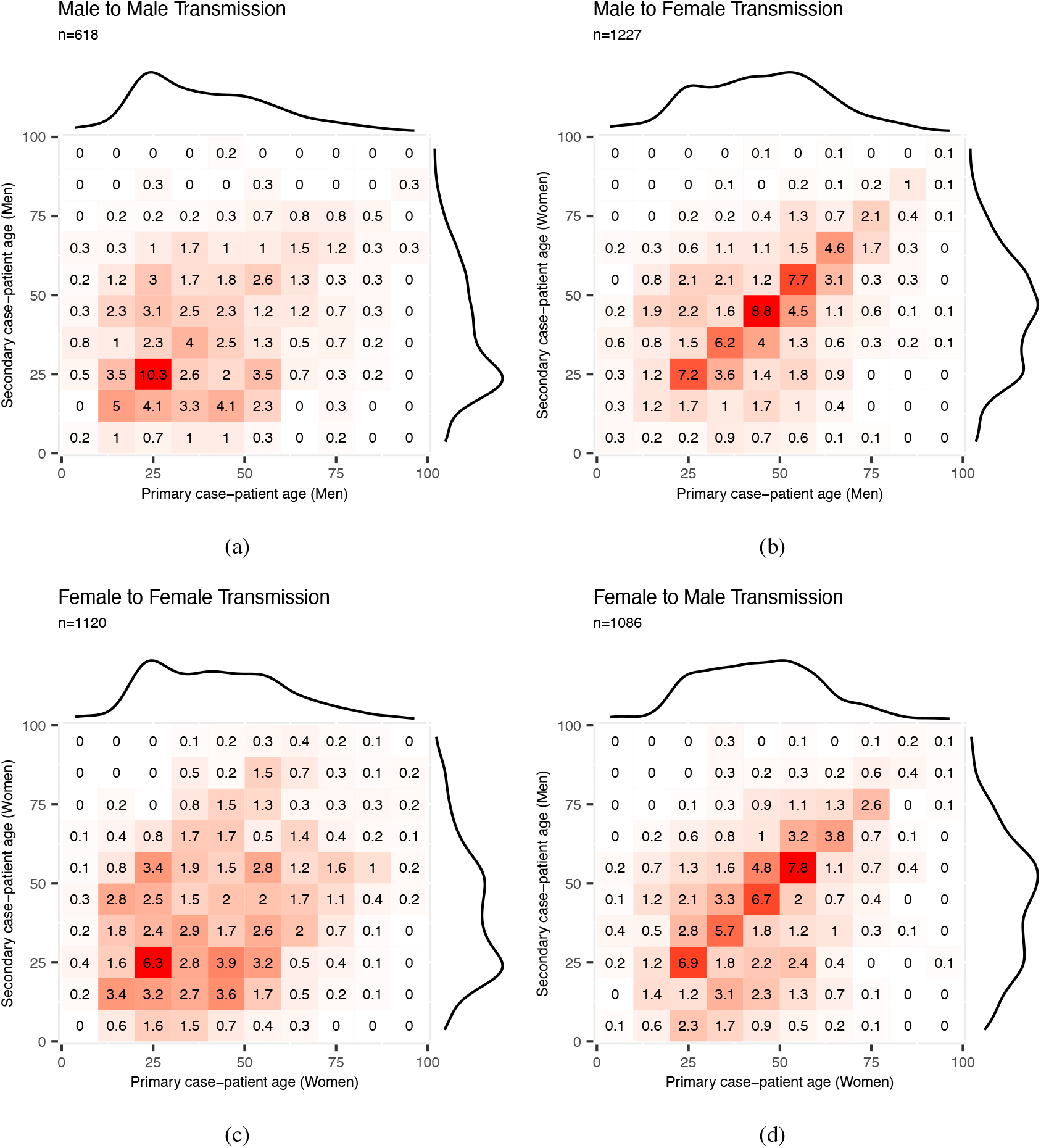
Patterns of COVID-19 disease transmission according to sex and age based on 4080 tracked pairs of COVID-19 cases over the time period of February–July 2020. The matrix graphs show numbers of transmission pairs as a percentage of the total, with primary case-patients (infectors) as columns and their secondary case-patients (infectees) as rows. Darker colors indicate a higher percentages of fraction of tracked pairs observed. Marginal totals are shown as density curves to illustrate the age distribution of case-patients.

**Figure 4:**
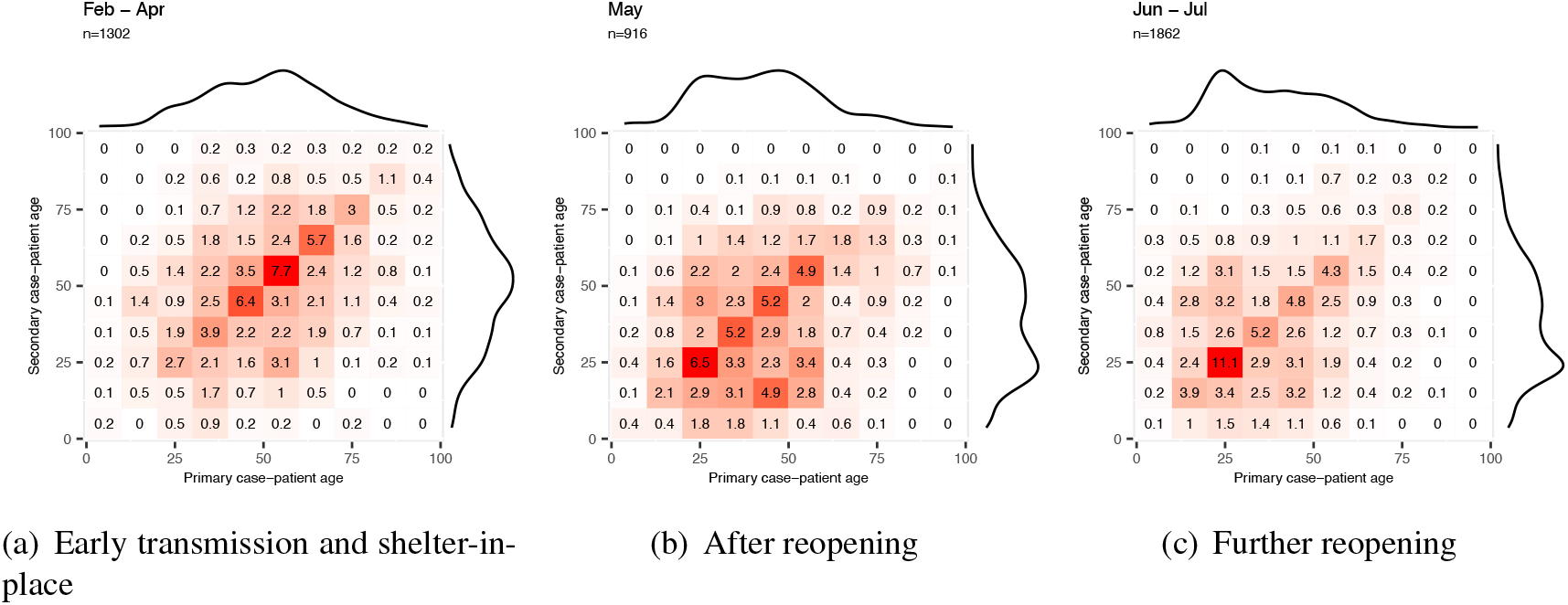
Patterns of COVID-19 disease transmission by age group in three successive time periods. Matrix graphs as in Figure 2. The time periods were divided as early transmission and shelter-in-place (February–April), after reopening (May), further reopening (June–July).

### Temporal and Spatial Patterns of Transmission

During February and March, reproduction numbers started higher than 1 and then decreased until late April and early May, which can be considered as the “first wave” in Georgia. *R*_*t*_ usually decreased to a (mathematical) local minimum during the shelter-in-place period but started to increase again as the “second wave” took off. As was observed during the first wave, the *R*_*t*_s peaked and then started to decrease (Figure 5). Although the number of reported cases was lower in first wave, the *R*_*t*_ (around 3) was much higher compared with the second wave.

**Figure 5:**
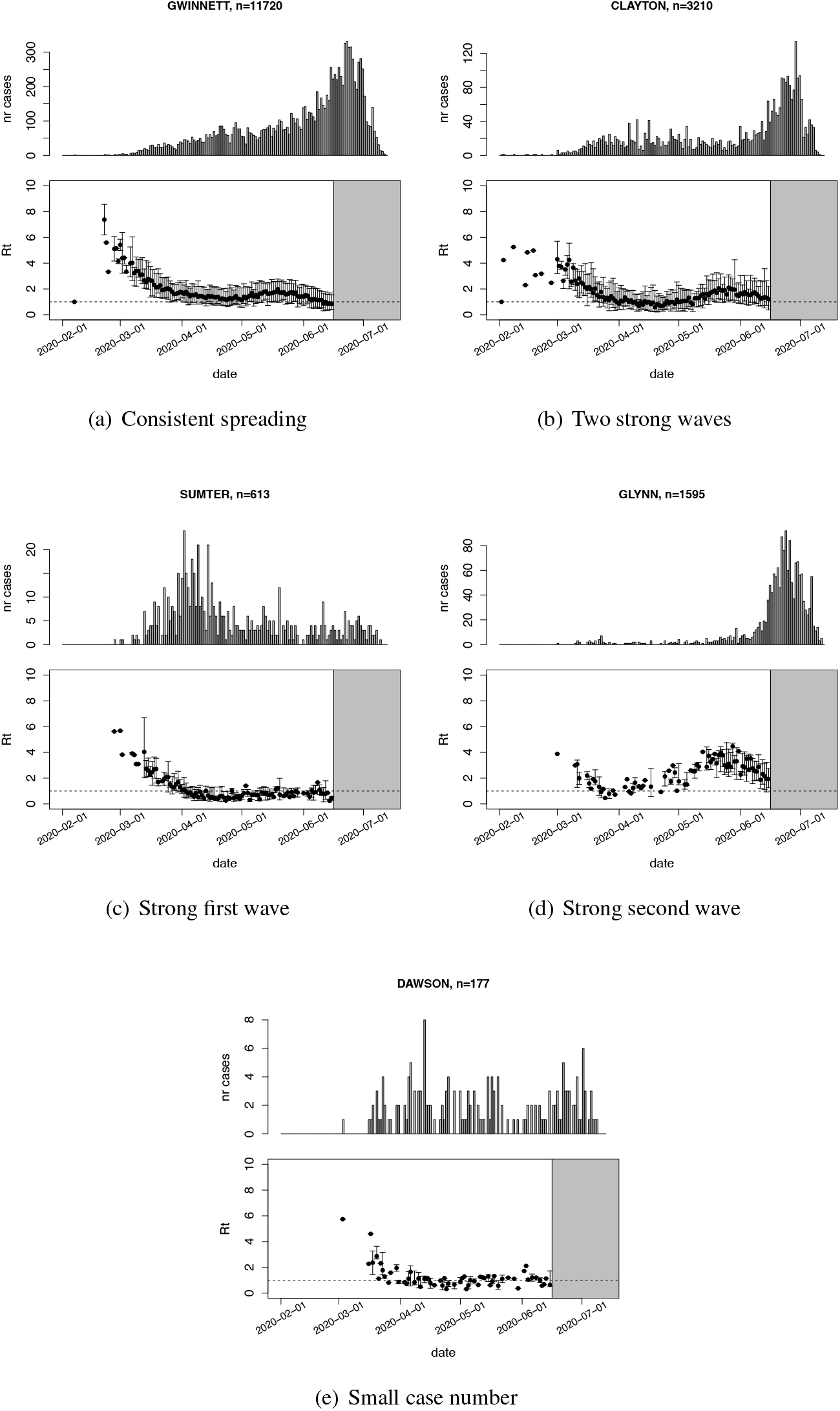
Epidemic curves from the start of the outbreak until July 13, 2020 and reproduction number estimates until June 15, 2020 in Gwinnett, Clayton, Sumter, Glynn, and Dawson counties. The x-axis represents the date of symptom onset. The y-axis in the top plot shows the number of cases while the y-axis in the bottom plot shows the estimated median reproduction numbers. The error bars represent 2.5th-97.5th percentile ranges of reproduction numbers. The grey area shows how *R*_*t*_ estimations were trunc2a0ted at June 15.

Although the general pattern of COVID-19 transmission was similar across all counties, the dates of local maxima/minima (i.e., first peak, local minimum, and second peak) and the magnitude of *R*_*t*_ at these extremes varied among counties. Figure 6(a) shows the peak dates for the first wave in counties with cumulative case numbers higher than 200 cases by July 13, 2020. At that time, counties with high numbers of COVID-19 cases were located around cities and along highways. Starting in early February, COVID-19 spread radially and along the interstate highway from Atlanta and Albany, the two initial outbreak sources. Other cities, including Augusta and Savannah had outbreaks later. Figure 7 shows that 74.7% (65/87) of counties with more than 200 cumulative cases by July 13th reached a local minimum in *R*_*t*_ during the shelter-in-place period (April 3–April 30). After reopening, many counties experienced a strong second wave with increased numbers of COVID-19 cases reported.

**Figure 6:**
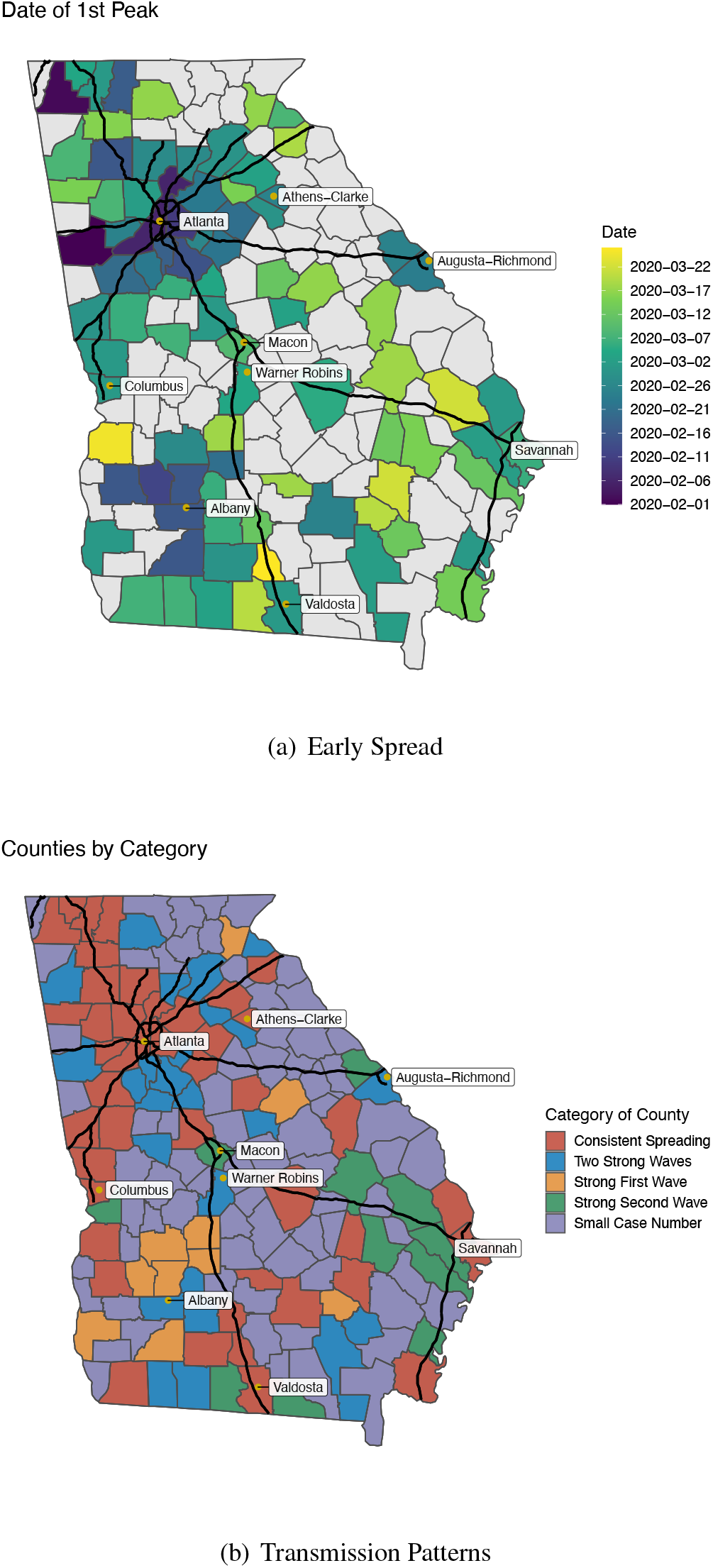
Spatial patterns of transmission of COVID-19. (a) date of reaching the peak (local maximum of *R*_*t*_) for the first wave. (b) spatial distribution of the five categories of transmission patterns of COVID-19 in Georgia by June 15, 2020. The black lines represent the interstate highways.

**Figure 7:**
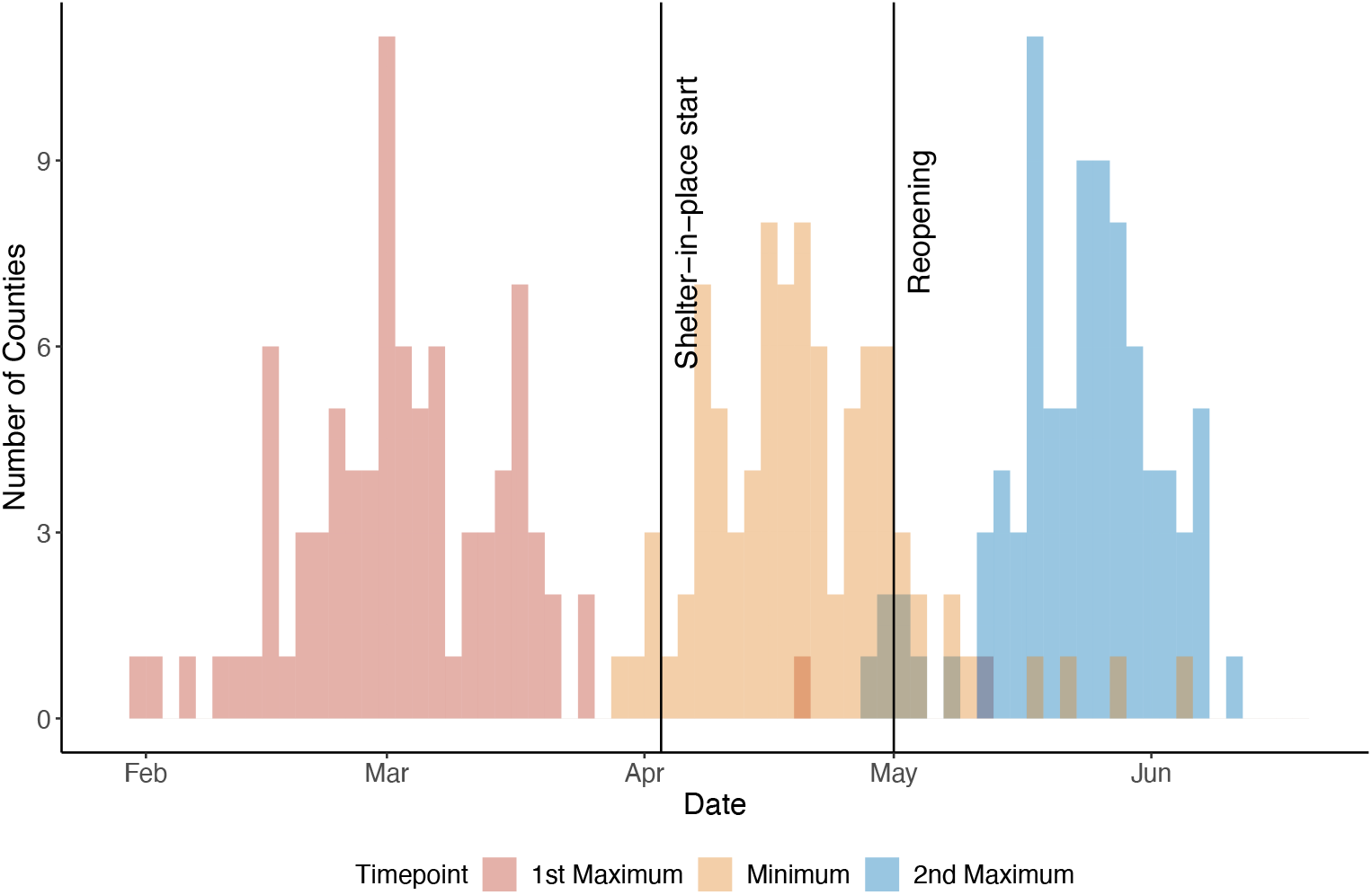
Distributions of estimated dates of first maximum, minimum, and second maximum in *R*_*t*_ for 87 counties with cumulative 200 cases by July 13, 2020, and key events possibly driving COVID-19 transmission.

Based on the magnitude of *R*_*t*_ at first peak, local minimum, and second peak, we categorized case data into the following patterns (Figure 5):

- **Consistent spreading**: sustained transmission of COVID-19 (*R*_*t*_ *>* 1) during the shelter-in-place period. Consequently, numbers of cases were high and increased rapidly upon reopening.
- **Two strong waves**: a first wave of early transmission followed by a slowdown (*R*_*t*_ *<* 1) during the shelter-in-place period. After reopening, a new surge in cases (1 ≤ *R*_*t*_ *<* 2) appeared.
- **Strong first wave**: a considerable number of cases during the initial period of the outbreak. During the shelter-in-place period spreading was controlled and after reopening no new surge in cases occurred (*R*_*t*_ *<* 1).
- **Strong second wave**: there were few cases during the early transmission period, but a surge in new cases (*R*_*t*_ ≥ 2) after reopening;
- **Small case number (***n<* 200): COVID-19 transmission was rare.

Figure 6(b) shows the spatial distribution of these five patterns of COVID-19 transmission on a map of the state of Georgia. Consistent spreading occurred in counties around some major cities, such as Atlanta, Athens, Columbus, Savannah, and counties along interstate highways. In counties bordering those counties with consistent spreading, there were two strong waves or only a strong second wave. Counties around the city of Albany (Lee, Sumter, Terrell, Mitchell, Crisp, and Dooly) had an early intense first wave but did not have a strong second wave. Counties not connected by the interstate highways had fewer COVID-19 cases.

## Discussion

### Serial Interval Contraction

During February–July 2020, the estimated serial intervals for symptom onset of COVID-19 in the state of Georgia appeared to become shorter (Figure 1). Such a phenomenon was also observed in mainland China during January–February 2020[18]. Shorter serial intervals imply more rapid transmission, leading to a higher number of new cases. Cases with severe symptoms and in older people tended to have longer observed serial intervals. During February–July, the state of Georgia had increasing disease prevalence and by August 25, 2020 it had the fifth highest number of confirmed COVID-19 cases in the U.S. In 2008, Kenah et al. showed that as the local prevalence increases, the mean generation interval, which is time between the infection of infector and infectee, becomes shorter. As there are increasingly more infectious cases present in the local population, they are competing to infect susceptibles and the expected time until a new infection is shortened [19]. Additional causes of contracting serial intervals could be that people had more contacts after reopening, in particular younger people (20–50 years old) might play a more important role in COVID-19 transmission; or that, with improved testing capacity, symptomatic cases were tested more promptly and thus isolated more quickly, which led to fewer exposures during their infectious periods.

The serial interval estimation also could be affected by changing testing practices over the duration of the pandemic. Georgia had limited COVID-19 testing capacity and contact tracing ability during earlier stages of the pandemic, and thus identification of COVID-19 cases during this period skewed towards populations with known COVID-19 contacts or COVID-19 associated symptoms. Because contact tracing is voluntary, there are likely segments of the population that are not represented in the tracing data due to unwillingness or inability of some contact persons to respond to contact tracers. Therefore, during February–April, older cases with more severe symptoms may have been over-represented in the notified case data. More recent data collected when availability of testing has improved are less likely to have this bias. As Figure 1 shows, contraction of serial intervals continued into May–early July, so that the changes may still be explained at least partly by increased prevalence and increased contact rates.

### Transmission Patterns by Age, Gender, and Race

Transmission of a respiratory infection such as COVID-19 is dependent on behavioral factors, and in particular on social contacts. Studies of contact behavior have shown that people tend to have social contacts with peers of the same age and demographic backgrounds [20]. The traced transmission links in this study show that such assortative mixing also applies to COVID-19 transmission (Figure 2). The tracked case pairs in this study were more likely to be observed when case-patients knew each other (e.g., family members, friends, or colleagues), while transmissions in public spaces (e.g., stores or restaurants) usually could not be traced. Transmission occurs frequently among subjects of the same age group, and less likely among different age groups. Figure 3 shows that some off-diagonal (between different age groups) transmission occurred in Georgia, although this may have been across generations, for example, between parents and children, or grandparents and grandchildren [21].

As to differences in transmission by sex, a primary case-patient who was male was more likely to pass their infection to a female contact than to a male contact. Figure 3(b) also shows that female case-patients were infected by male case-patients across a wide range of ages, while Figure 3(a) shows that male case-patients were mainly infected by young male case-patients. A possible explanation may be that females tend to be caregivers, taking care of sick people in the household, while young males may be more likely to acquire infection from outside the household.

Like the serial interval, transmission patterns also changed as the pandemic continued. The major contribution to spreading COVID-19 shifted over time to younger generation. This could be caused by the elderly becoming more careful to protect themselves from infection by taking measures such as staying at home, wearing face masks in public spaces, and observing good hand hygiene. At the same time, younger people might have been less compliant with quarantine measures and more likely to attend indoor gatherings such as parties, or to have visited bars, gyms, and clubs while not wearing facemasks.

### Waves of COVID-19 and Impact of Shelter-in-place and Reopening

Previous pandemics, such as the 1918 Spanish Flu and the 2009 H1N1 Swine Flu caused multiple waves of infections [22]. In the state of Georgia, we have observed two waves of COVID-19 transmission separated by the shelter-in-place period. The COVID-19 cases of the first wave were first observed in Atlanta, the state capital with one of the busiest airports in US, and Albany, the eighth largest city in Georgia. The outbreak in Albany was sparked by two super-spreading funeral events. However, the connectivity of these two cities is different: Atlanta as a transportation hub connects multiple interstate highways while Albany has no interstate highways. During the first wave, COVID-19 spread radially from both cities to the surrounding areas. For Atlanta, cases also started to appear along the interstate highway (Figure 6(a)). Concentrations of increased transmission along highways, as links connecting population centers, suggests that commuter links might have been effective transmission links.

During the shelter-in-place period (April 3–April 30), COVID-19 transmission slowed and the reproduction numbers reached a local minimum in most counties. However, before reopening, the reproduction numbers were still above 1 in many counties even at the local minimum, indicating continued disease spreading (Figure 6(b)). After reopening, transmission increased again across the state of Georgia. These data suggest that the three or four weeks of shelter-in-place orders were not long enough to sufficiently suppress COVID-19 transmission (local and out-of-state imported) in densely populated urban areas connected by major transportation links (i.e., airports and interstate highways).

Thus far, the second wave has been heterogenous in time and magnitude in different counties. Local prevalence was different at the time of reopening, and counties with high prevalence (i.e., counties bordering cities and along interstate highways) experienced a stronger second wave. Counties (e.g., those counties around Albany) not connected by major transportation links often saw a second wave of COVID-19 cases as well, but on a relatively small scale. Finally, some counties (e.g., Lee, Sumter, Terrell, and Mitchell), that saw an early and intense first wave, did not experience an obvious second wave. Possibly, inhabitants of these counties were more compliant with the disease prevention and control measures.

## Limitations

Despite having information for more than one hundred thousand cases, a majority of the records had missing reported contacts with a confirmed case. The tracked pairs were missed not at random, since contact tracing is voluntary and its capacity was limited at the early stage of the pandemic. Also, tracked pairs were more likely to be recorded when they involved known contacts; and identifying transmission links in public spaces or in a cluster of cases is challenging. In addition, testing capacity of COVID-19 showed a spatiotemporal variation with only cases with severe symptoms detected during February–April.

## Data Availability

The raw data used in this study are available from the corresponding author upon reasonable request.

## Key Findings

In this study, we found the COVID-19 transmission changed over time during February–July 2020. The serial interval decreased from 5.97 days in February–April to 4.40 days in June–July. The younger population (20–50 years old) was involved in the majority of transmissions occurring during or after reopening subsequent to the shelter-in-place period. By mid-July, two waves of COVID-19 transmission were apparent, separated by the shelter-in-place period in the state of Georgia. Counties around major cities and along interstate highways had more intense transmission. These findings of transmission patterns can help in predicting and guiding states in prevention and control of COVID-19 according to population and region.

## Contributors

YW and PT conceived the study. YW and PT developed the methods. YW, CS, YC, and CA conducted literature review. YW, CS, and YC produced the estimates. YW, CS, and YC created figures and tables. YW and PT wrote the first draft of the manuscript and led the writing of subsequent drafts. BL, LE, MT, CS, CA, and ML provided critical feedback on the first draft and contributed to the writing of subsequent drafts of the manuscript. ML and BL acquired the funding for the research.

## Declaration of Interests

The authors declare no competing interests.

## Disclaimer

The findings and conclusions in this report are those of the authors and do not necessarily represent the official position of the Centers for Disease Control and Prevention.

## Acknowledgements

This study was funded by the Emory Covid-19 Response Collaborative. We are grateful to Dr. Hannah Cooper and Laura Donnelly from Emory University for their efforts in leading and coordinating the research partnership between GDPH and Emory University. And we thank Michael Bryan from the Georgia Department of Public Health for providing the data.

## Supplementary material

### A Supplemental Tables

**Table. S1:**
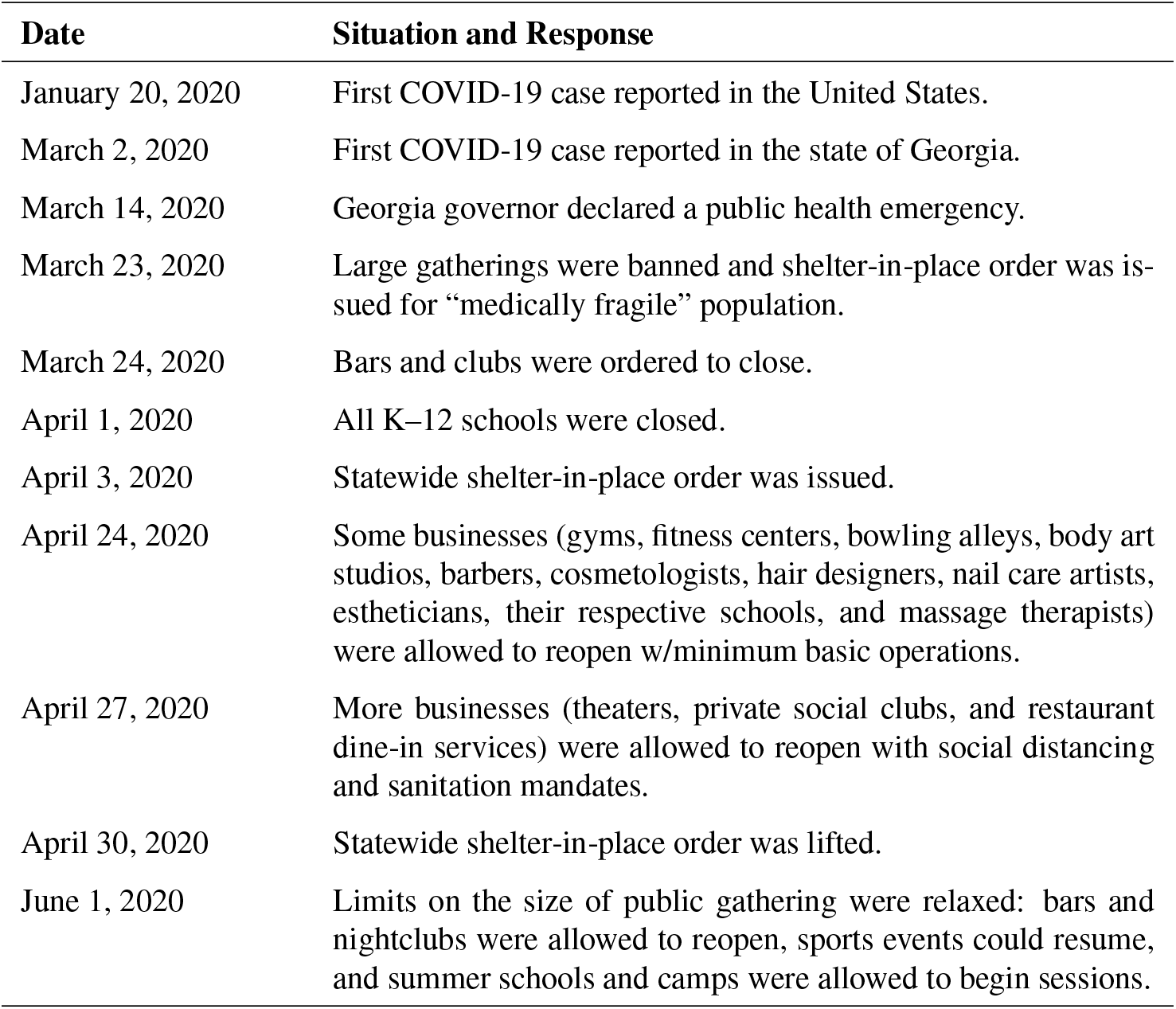
COVID-19 pandemic situation and state government responses.

| Date | Situation and Response |
| --- | --- |
| January 20, 2020 | First COVID-19 case reported in the United States. |
| March 2, 2020 | First COVID-19 case reported in the state of Georgia. |
| March 14, 2020 | Georgia governor declared a public health emergency. |
| March 23, 2020 | Large gatherings were banned and shelter-in-place order was issued for “medically fragile” population. |
| March 24, 2020 | Bars and clubs were ordered to close. |
| April 1, 2020 | All K–12 schools were closed. |
| April 3, 2020 | Statewide shelter-in-place order was issued. |
| April 24, 2020 | Some businesses (gyms, fitness centers, bowling alleys, body art studios, barbers, cosmetologists, hair designers, nail care artists, estheticians, their respective schools, and massage therapists) were allowed to reopen w/minimum basic operations. |
| April 27, 2020 | More businesses (theaters, private social clubs, and restaurant dine-in services) were allowed to reopen with social distancing and sanitation mandates. |
| April 30, 2020 | Statewide shelter-in-place order was lifted. |
| June 1, 2020 | Limits on the size of public gathering were relaxed: bars and nightclubs were allowed to reopen, sports events could resume, and summer schools and camps were allowed to begin sessions. |

**Table. S2:**
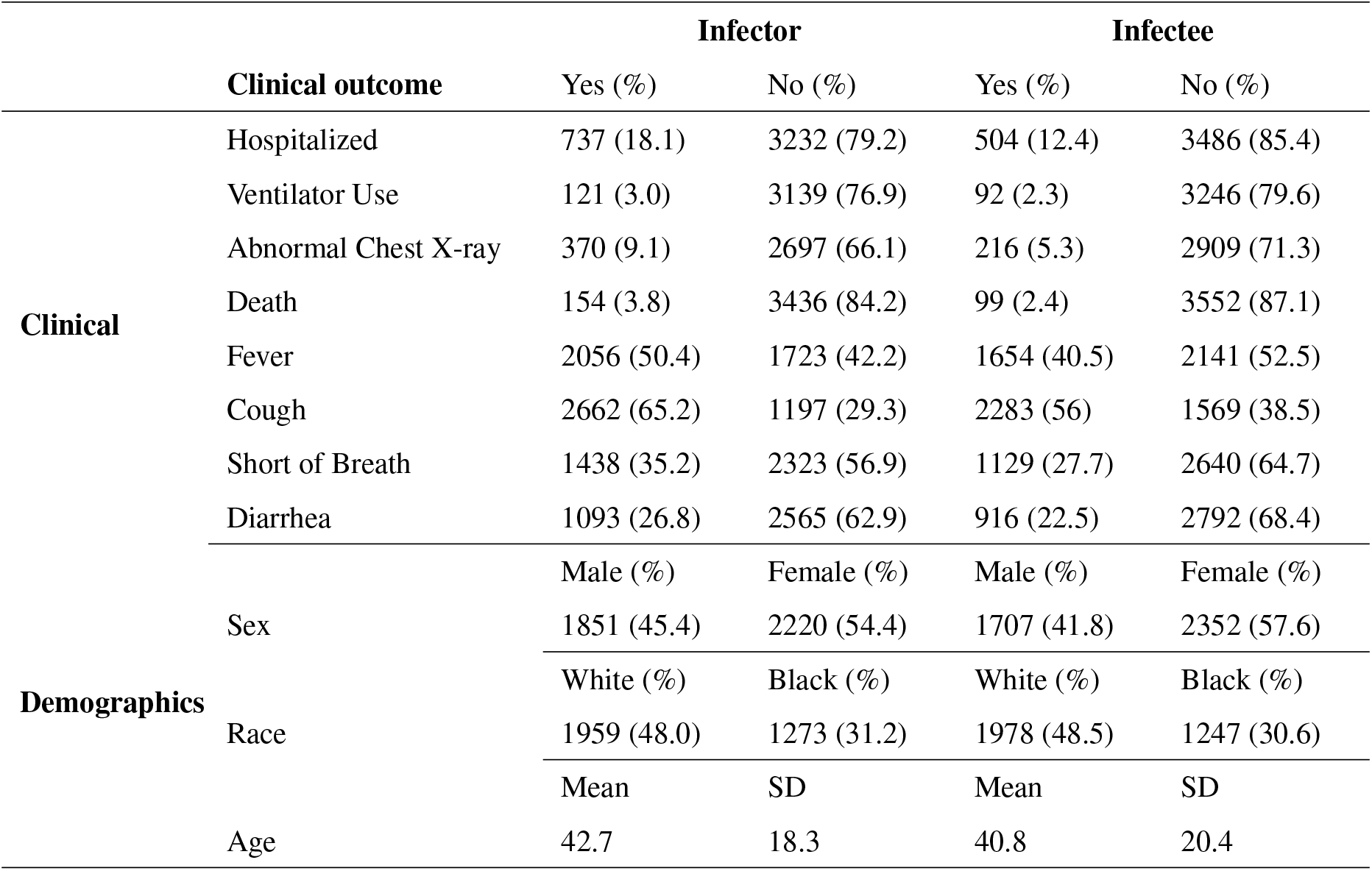
Clinical outcomes and demographic information for 4080 tracked pairs of primary case-patients (infectors) and secondary case-patients (infectees).

**Table. S3:** Serial interval by subgroup with different clinical characteristics and demographic characteristics

|  | Subgroup | n | Mean (day) | SD (day) | shape | scale |
| --- | --- | --- | --- | --- | --- | --- |
| **Clinical Info** | Hospitalized+ | 650 | 5.69 | 3.68 | 2.05 | 2.77 |
|  | Hospitalized- | 2986 | 4.84 | 3.43 | 2.00 | 2.42 |
|  | Ventilator+ | 106 | 6.28 | 3.81 | 2.21 | 2.84 |
|  | Ventilator- | 2877 | 4.93 | 3.43 | 2.02 | 2.44 |
|  | Abnormal Chest X-ray+ | 307 | 6.13 | 3.76 | 2.22 | 2.76 |
|  | Abnormal Chest X-ray- | 2492 | 4.84 | 3.37 | 2.04 | 2.37 |
|  | Death+ | 133 | 5.91 | 3.74 | 2.06 | 2.87 |
|  | Death- | 3153 | 4.95 | 3.45 | 2.03 | 2.44 |
|  | Fever+ | 1889 | 5.12 | 3.52 | 2.06 | 2.48 |
|  | Fever- | 1575 | 4.87 | 3.45 | 1.94 | 2.51 |
|  | Cough+ | 2435 | 5.09 | 3.51 | 2.05 | 2.49 |
|  | Cough- | 1096 | 4.79 | 3.45 | 1.92 | 2.50 |
|  | Short of Breath+ | 1302 | 5.25 | 3.51 | 2.07 | 2.54 |
|  | Short of Breath- | 2140 | 4.81 | 3.45 | 1.98 | 2.43 |
|  | Diarrhea+ | 993 | 5.22 | 3.71 | 1.88 | 2.77 |
|  | Diarrhea- | 2349 | 4.87 | 3.39 | 2.05 | 2.37 |
| **Age** | 20- | 412 | 4.10 | 3.06 | 1.96 | 2.09 |
|  | 20–40 | 1369 | 4.76 | 3.32 | 2.05 | 2.33 |
|  | 40–60 | 1330 | 5.29 | 3.61 | 2.05 | 2.58 |
|  | 60+ | 618 | 5.47 | 3.70 | 1.99 | 2.75 |
| **Sex** | Male | 1720 | 4.85 | 3.45 | 1.96 | 2.47 |
|  | Female | 2009 | 5.11 | 3.51 | 2.05 | 2.50 |
| **Race** | Black | 1147 | 5.39 | 3.63 | 2.04 | 2.64 |
|  | White | 1790 | 4.73 | 3.39 | 1.95 | 2.43 |
|  | Other | 801 | 4.99 | 3.49 | 2.00 | 2.49 |
| **Resident Area** | Metro Atlanta | 839 | 4.88 | 3.38 | 2.06 | 2.37 |
|  | Out of Metro Atlanta | 2899 | 5.02 | 3.52 | 1.99 | 2.53 |
| **Time Period** | Feb-Apr | 1070 | 5.97 | 3.84 | 2.09 | 2.86 |
|  | May | 850 | 5.03 | 3.44 | 2.11 | 2.38 |
|  | Jun-Jul | 1818 | 4.40 | 3.14 | 2.03 | 2.17 |
| **Total** |  | 3738 | 4.99 | 3.49 | 2.00 | 2.49 |

For clinical characteristics, “+” represents yes and “-” represents no. The shape and scale parameters of gamma distributions were estimated using maximum likelihood estimator.

## Footnotes

6 See e.g., 45 C.F.R part 46, 21 C.F.R part 56; 42 U.S.C. *§*241(d); 5 U.S.C. *§*552a; 44 U.S.C. *§*3501 et seq.

## Notes

### Competing Interest Statement

The authors have declared no competing interest.

### Author Declarations

The GDPH Institutional Review Board has determined that this analysis is exempt from the requirement for IRB review and approval and informed consent was not required. This activity was reviewed by the Centers for Disease Control and Prevention (CDC) and was conducted consistent with applicable federal law and CDC policy.

## References

1. Dong E, Du H, Gardner L. An interactive web-based dashboard to track covid-19 in real time. The Lancet infectious diseases 2020;20(5):533–534.

2. Georgia Department of Public Health. Georgia department of public health daily status report, 2020. URL https://dph.georgia.gov/covid-19-daily-status-report. [Online; accessed 22-October-2020].

3. Kraemer MU, Yang CH, Gutierrez B, et al. The effect of human mobility and control measures on the covid-19 epidemic in china. Science 2020;368(6490):493–497.http:

4. Wang Y, Teunis P. Strongly heterogeneous transmission of covid-19 in mainland china: Local and regional variation. Frontiers in Medicine 2020;7. doi:10.3389/fmed.2020.00329. URL http:dx.doi.org/10.3389/fmed.2020.00329

5. Rocklöv J, Sjödin H. High population densities catalyse the spread of covid-19. Journal of Travel Medicine 2020;27(3). doi:10.1093/jtm/taaa038. URL http://dx.doi.org/10.1093/jtm/taaa038.

6. Prem K, Liu Y, Russell TW, et al. The effect of control strategies to reduce social mixing on outcomes of the covid-19 epidemic in wuhan, china: a modelling study. The Lancet Public Health 2020; 5(5):e261–e270. doi:10.1016/S2468-2667(20)30073-6. URL https://doi.org/10.1016/S2468-2667(20)30073-6.

7. McMichael TM, Currie DW, Clark S, et al. Epidemiology of covid-19 in a long-term care facility in king county, washington. New England Journal of Medicine 2020;382(21):2005–2011. doi:10.1056/nejmoa2005412. URL http://dx.doi.org/10.1056/NEJMoa2005412.

8. Liu Y, Eggo RM, Kucharski AJ. Secondary attack rate and superspreading events for sars-cov-2. The Lancet 2020;395(10227):e47.

9. Lloyd-Smith JO, Schreiber SJ, Kopp PE, Getz WM. Superspreading and the effect of individual variation on disease emergence. Nature 2005;438(7066):355–359. doi:10.1038/nature04153. URL http://dx.doi.org/10.1038/nature04153

10. Lai S, Ruktanonchai NW, Zhou L, et al. Effect of non-pharmaceutical interventions to contain covid-19 in china. Nature 2020;585(7825):410–413. doi:10.1038/s41586-020-2293-x. URL https://doi.org/10.1038/s41586-020-2293-x

11. Cowling BJ, Ali ST, Ng TW, et al. Impact assessment of non-pharmaceutical interventions against coronavirus disease 2019 and influenza in hong kong: an observational study. The Lancet Public Health 2020;5(5):e279–e288. doi:10.1016/S2468-2667(20)30090-6. URL https://doi.org/10.1016/S2468-2667(20)30090-6.

12. Flaxman S, Mishra S, Gandy A, et al. Estimating the effects of non-pharmaceutical interventions on covid-19 in europe. Nature 2020;pages 1–5.

13. Wikipedia contributors. COVID-19 pandemic in Georgia (u.s. state), 2020. URL https://en.wikipedia.org/wiki/COVID-19_pandemic_in_Georgia_(U.S._state). [Online; accessed 25-August-2020].

14. Wallinga J, Teunis P. Different epidemic curves for severe acute respiratory syndrome reveal similar impacts of control measures. American Journal of epidemiology 2004;160(6):509–516.

15. Teunis P, Heijne JC, Sukhrie F, van Eijkeren J, Koopmans M, Kretzschmar M. Infectious disease transmission as a forensic problem: who infected whom? Journal of the Royal Society Interface 2013;10(81):20120955.

16. Nishiura H, Linton NM, Akhmetzhanov AR. Serial interval of novel coronavirus (covid-19) infections. International journal of infectious diseases 2020;93:284–286.

17. Du Z, Xu X, Wu Y, Wang L, Cowling BJ, Meyers LA. Serial interval of covid-19 among publicly reported confirmed cases. Emerging infectious diseases 2020;26(6):1341.

18. Ali ST, Wang L, Lau EH, et al. Serial interval of sars-cov-2 was shortened over time by nonpharmaceutical interventions. Science 2020;369(6507):1106–1109.

19. Kenah E, Lipsitch M, Robins JM. Generation interval contraction and epidemic data analysis. Mathematical Biosciences 2008;213(1):71–79.

20. Newman ME. Mixing patterns in networks. Physical review E 2003;67(2):026126.

21. Mossong J, Hens N, Jit M, et al. Social contacts and mixing patterns relevant to the spread of infectious diseases. PLoS Med 2008;5(3):e74.

22. Mummert A, Weiss H, Long LP, Amigú JM, Wan XF. A perspective on multiple waves of influenza pandemics. PloS one 2013;8(4):e60343.

