## Supplemental tables. for "Transmission of COVID-19 in the state of Georgia, United States: Spatiotemporal variation and impact of social distancing"

### B Supplemental Figures

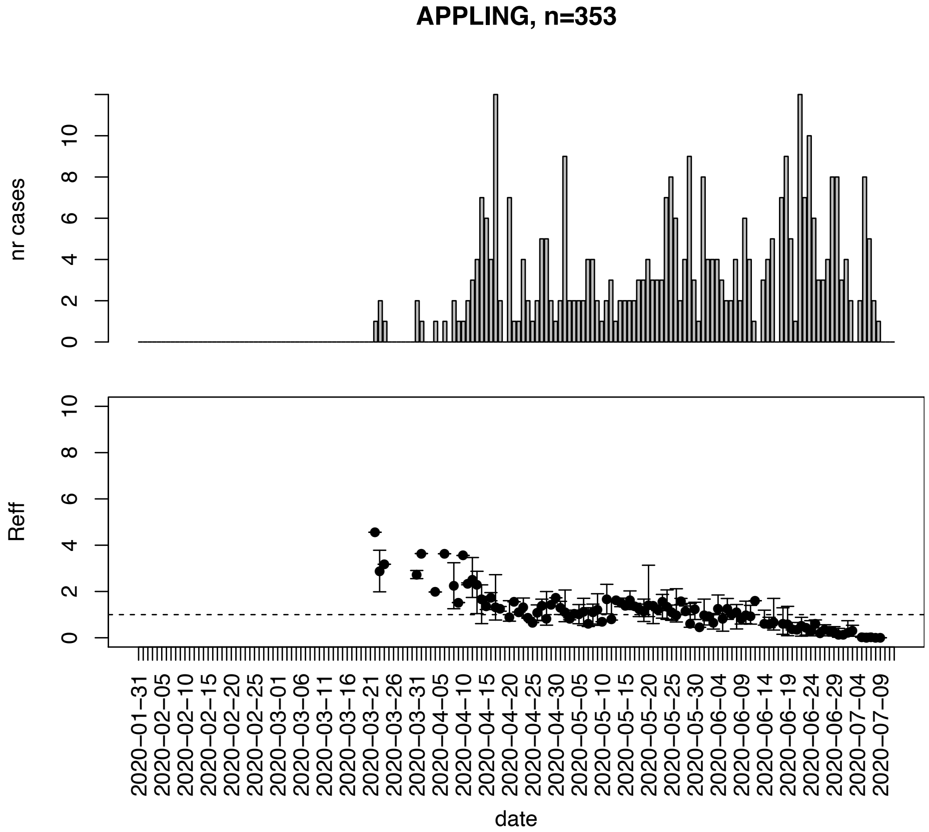


Figure. S1: Epidemic curves and reproduction number estimates until July 13th in Appling county.

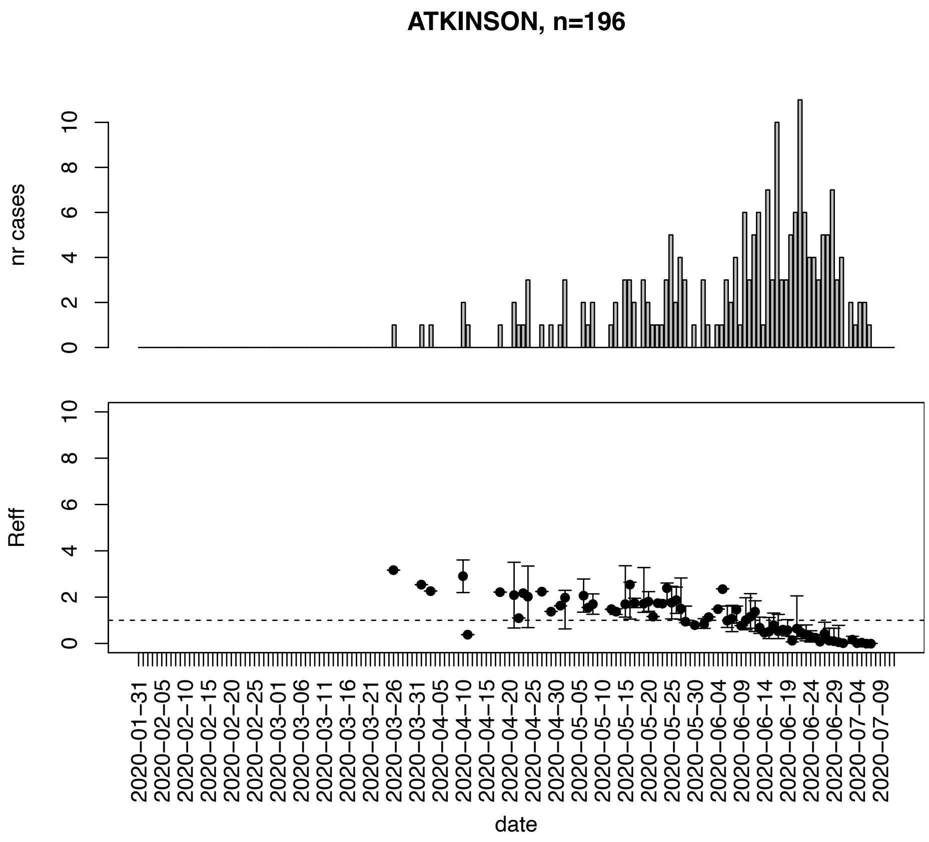


Figure. S2: Epidemic curves and reproduction number estimates until July 13th in Atkinson county.

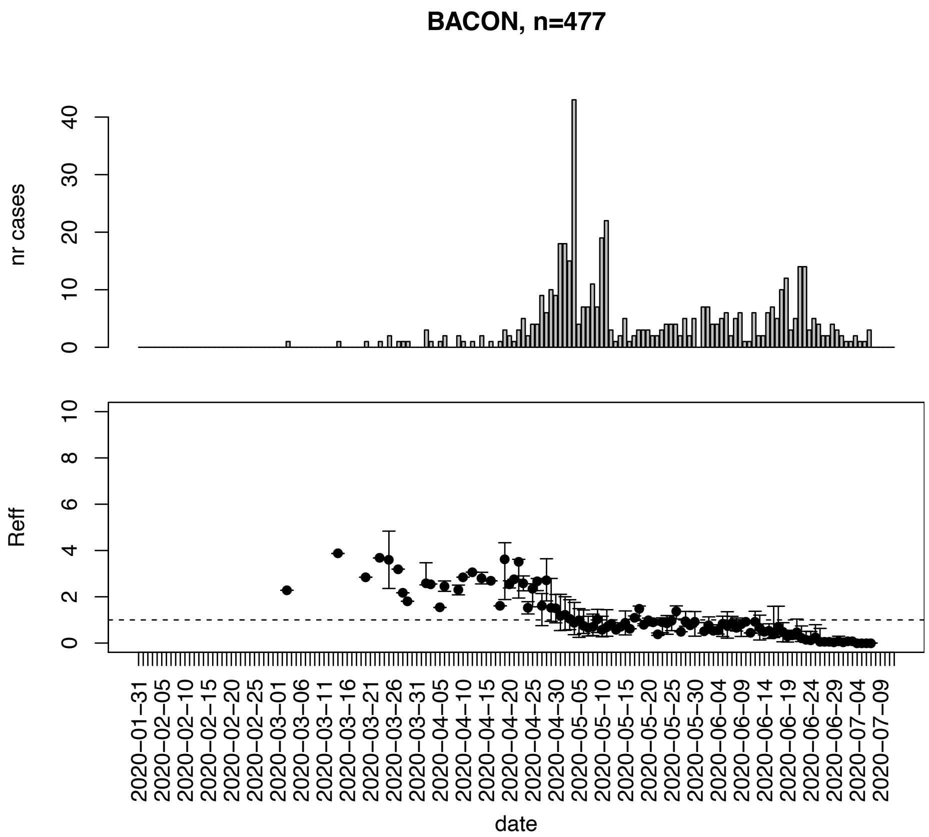


Figure. S3: Epidemic curves and reproduction number estimates until July 13th in Bacon county.

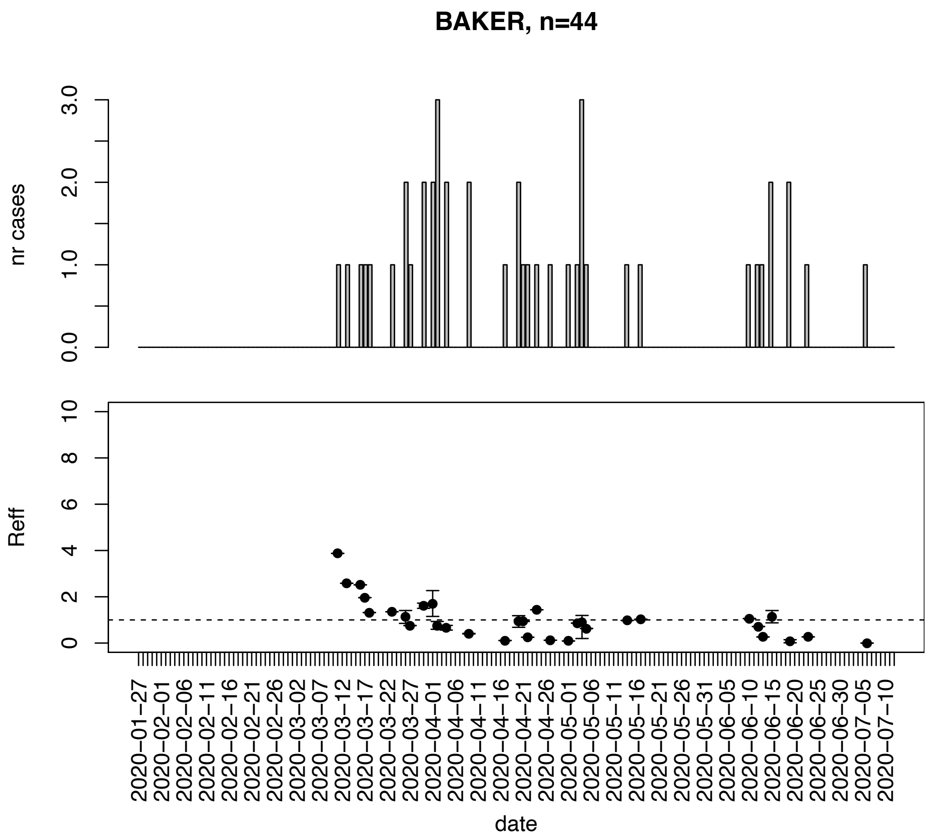


Figure. S4: Epidemic curves and reproduction number estimates until July 13th in Baker county.

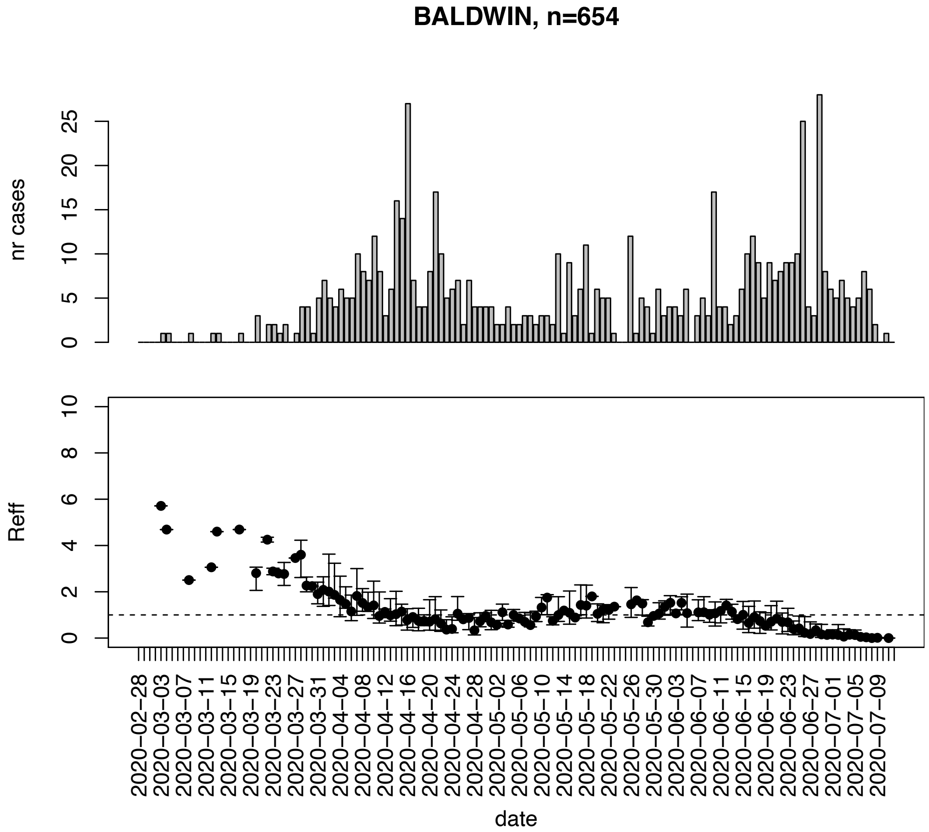


Figure. S5: Epidemic curves and reproduction number estimates until July 13th in Baldwin county.

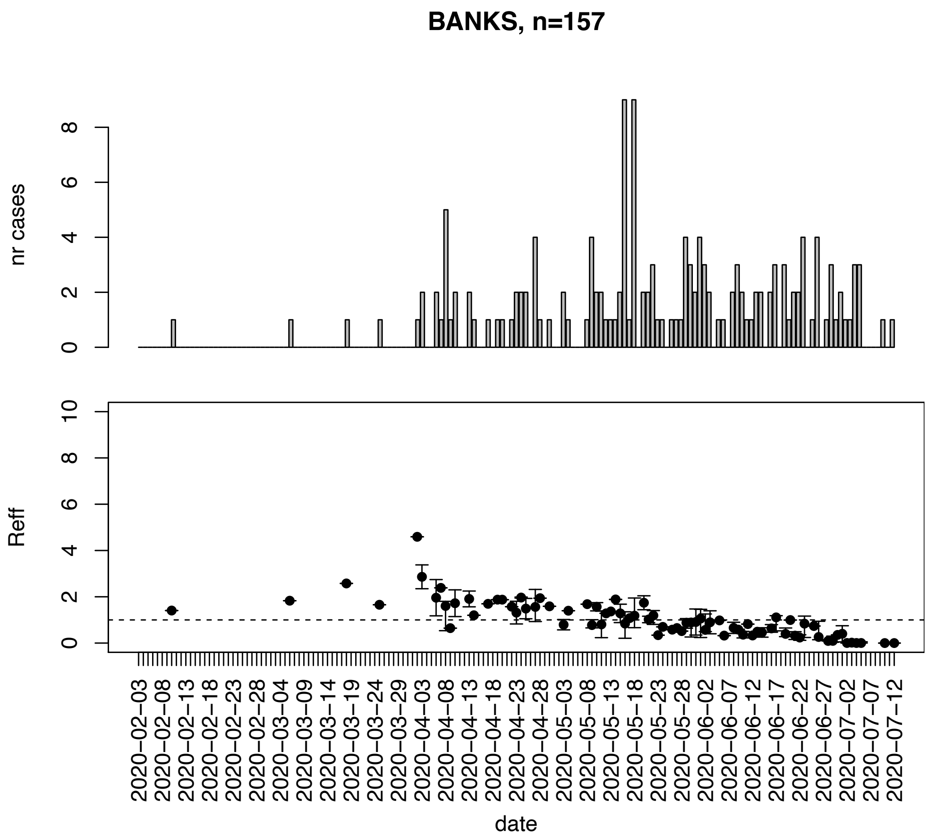


Figure. S6: Epidemic curves and reproduction number estimates until July 13th in Banks county.

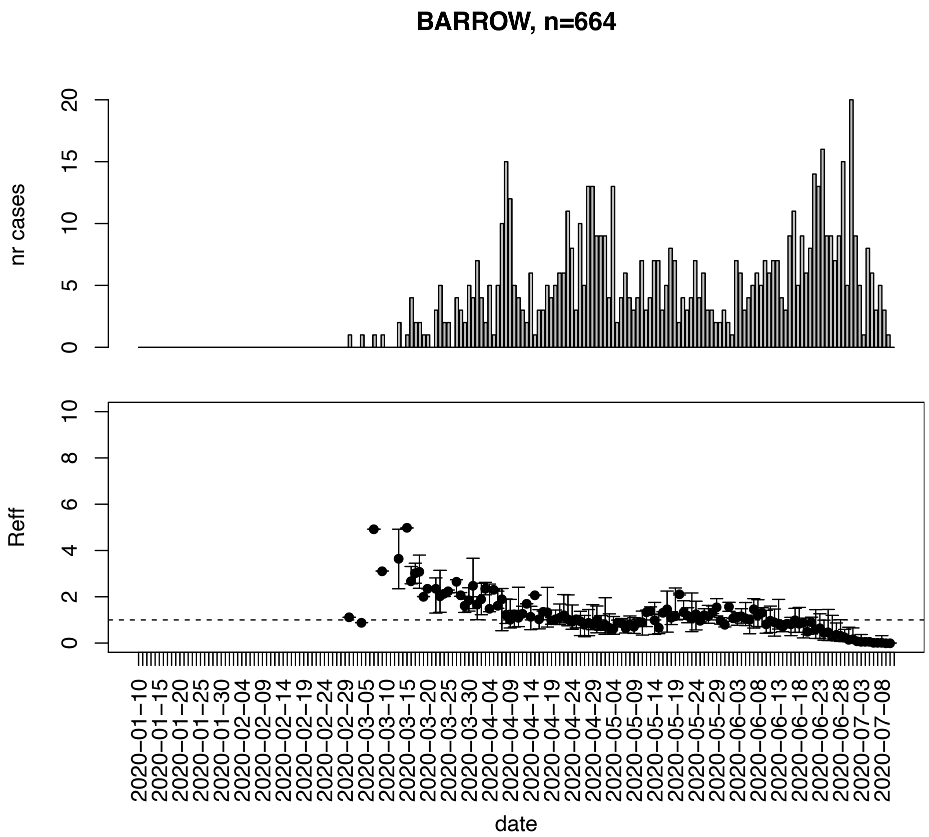


Figure. S7: Epidemic curves and reproduction number estimates until July 13th in Barrow county.

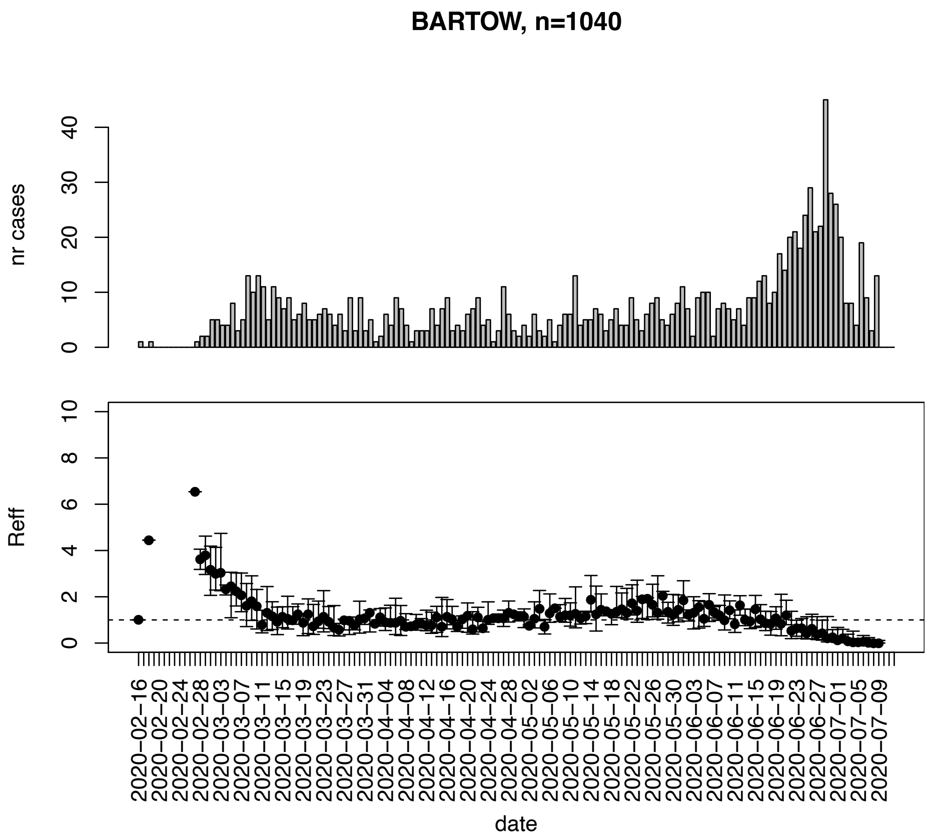


Figure. S8: Epidemic curves and reproduction number estimates until July 13th in Bartow county.

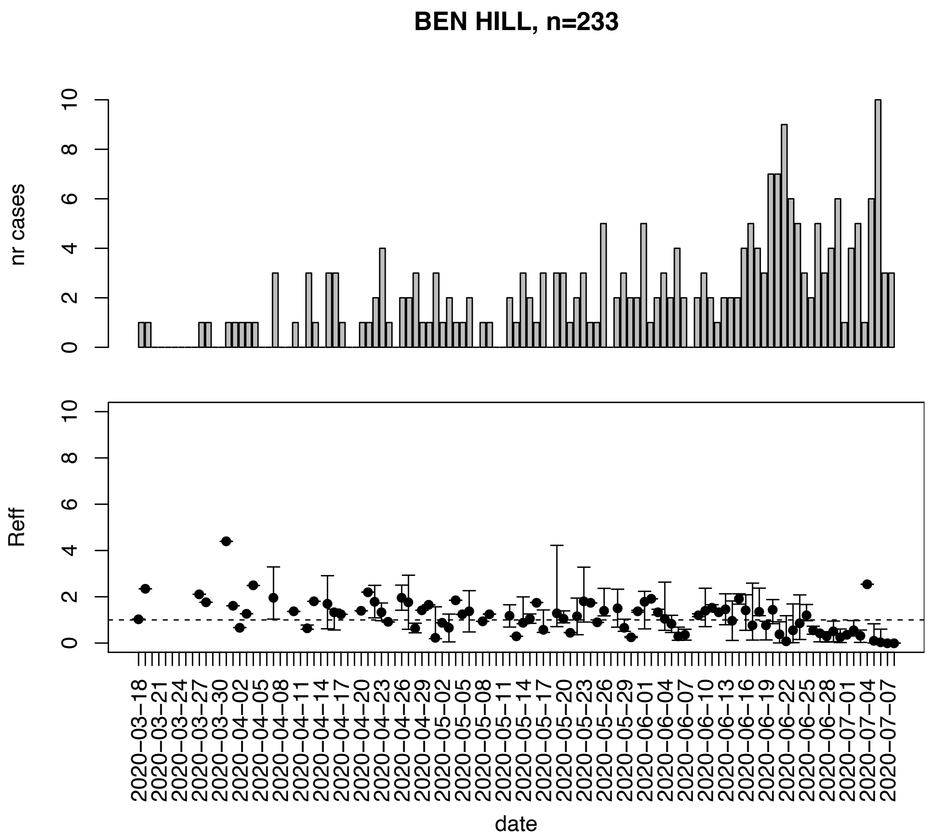


Figure. S9: Epidemic curves and reproduction number estimates until July 13th in Ben Hill county.

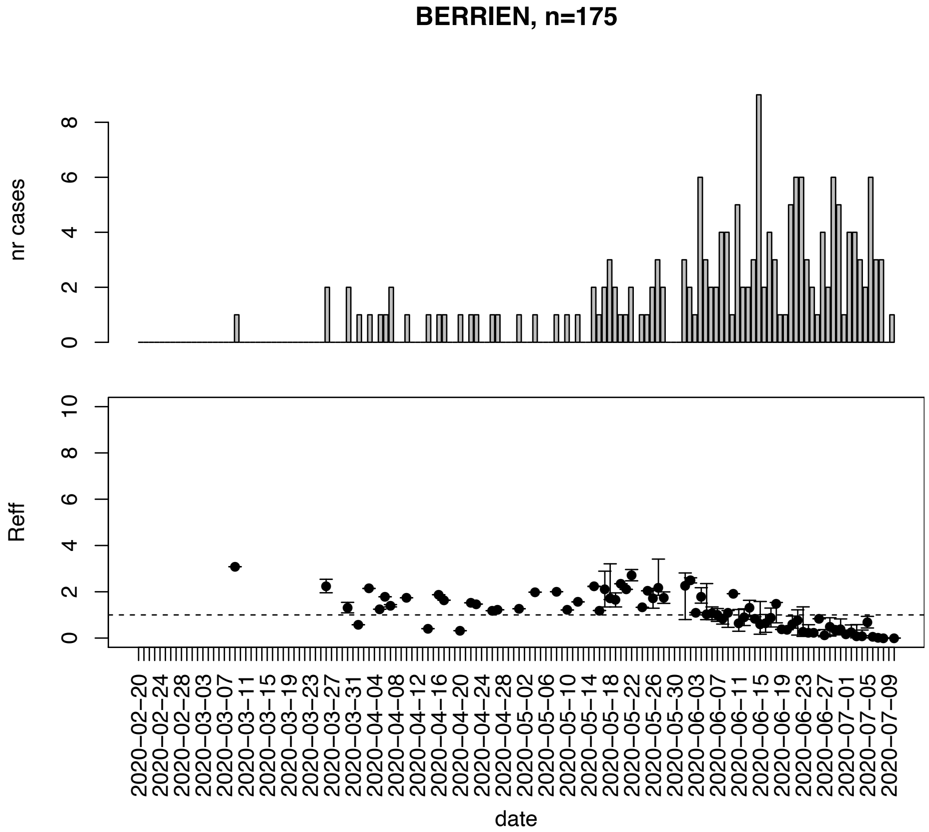


Figure. S10: Epidemic curves and reproduction number estimates until July 13th in Berrien county.

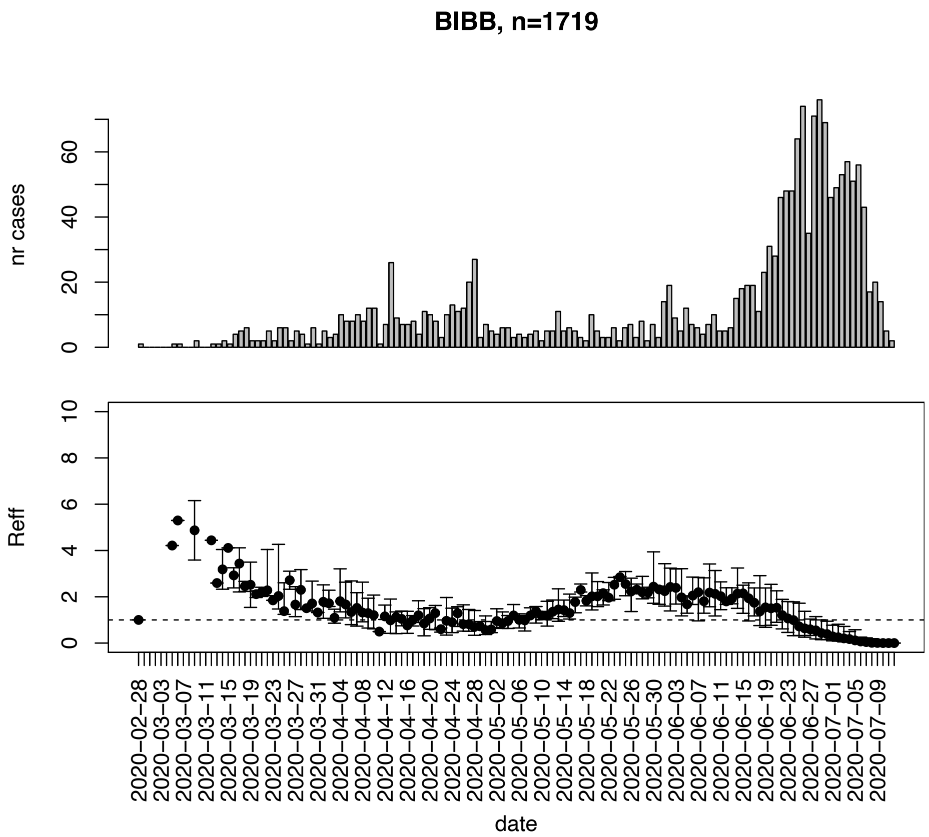


Figure. S11: Epidemic curves and reproduction number estimates until July 13th in Bibb county.

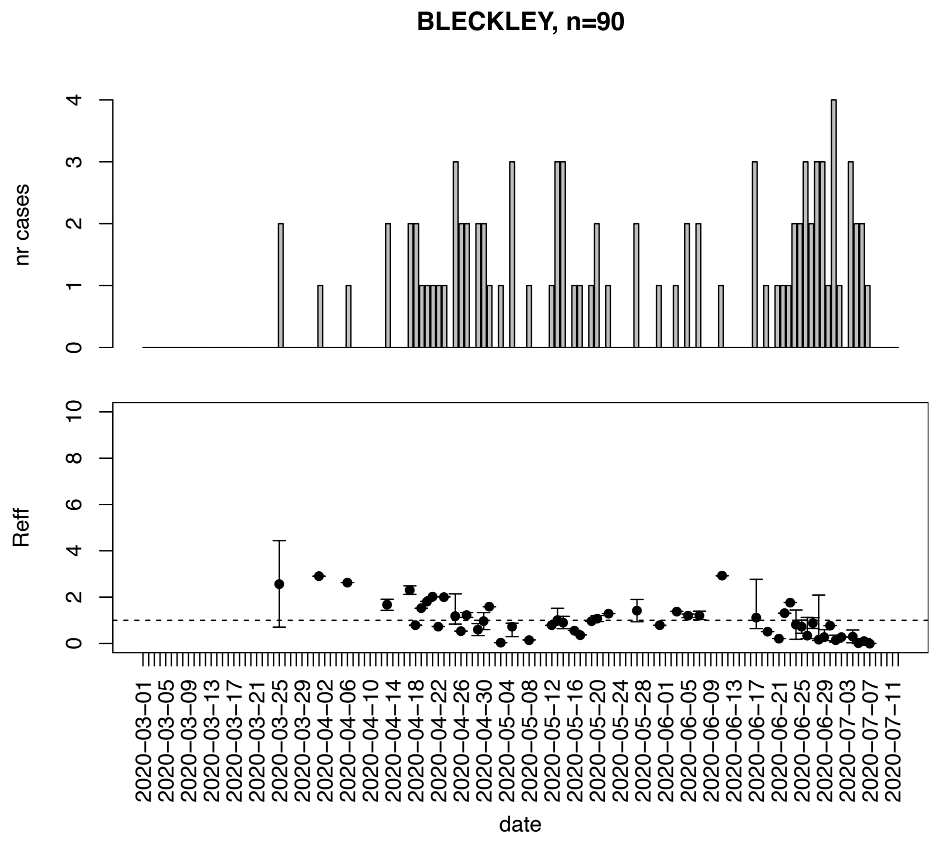


Figure. S12: Epidemic curves and reproduction number estimates until July 13th in Bleckley county.

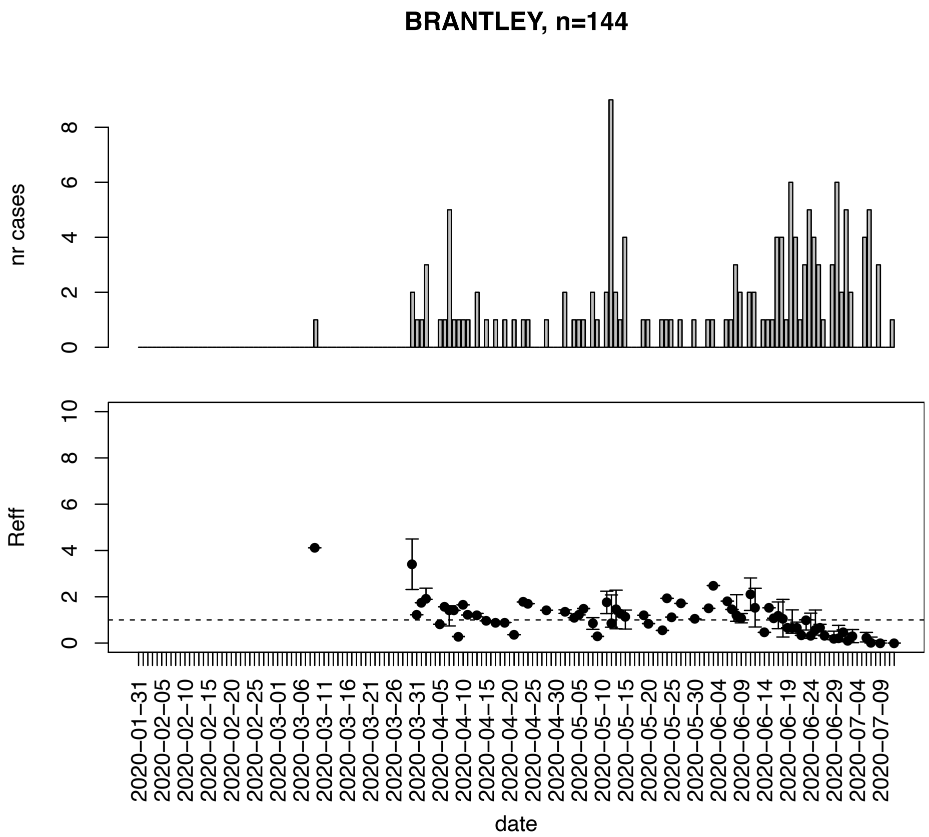


Figure. S13: Epidemic curves and reproduction number estimates until July 13th in Brantley county.

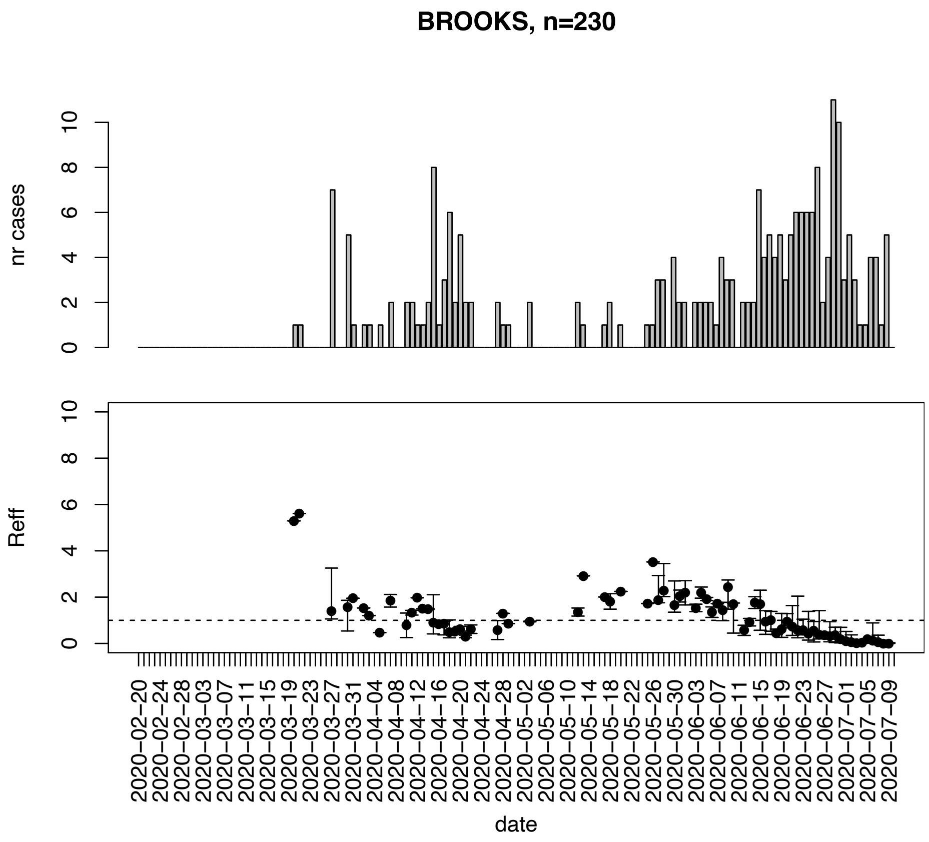


Figure. S14: Epidemic curves and reproduction number estimates until July 13th in Brooks county.

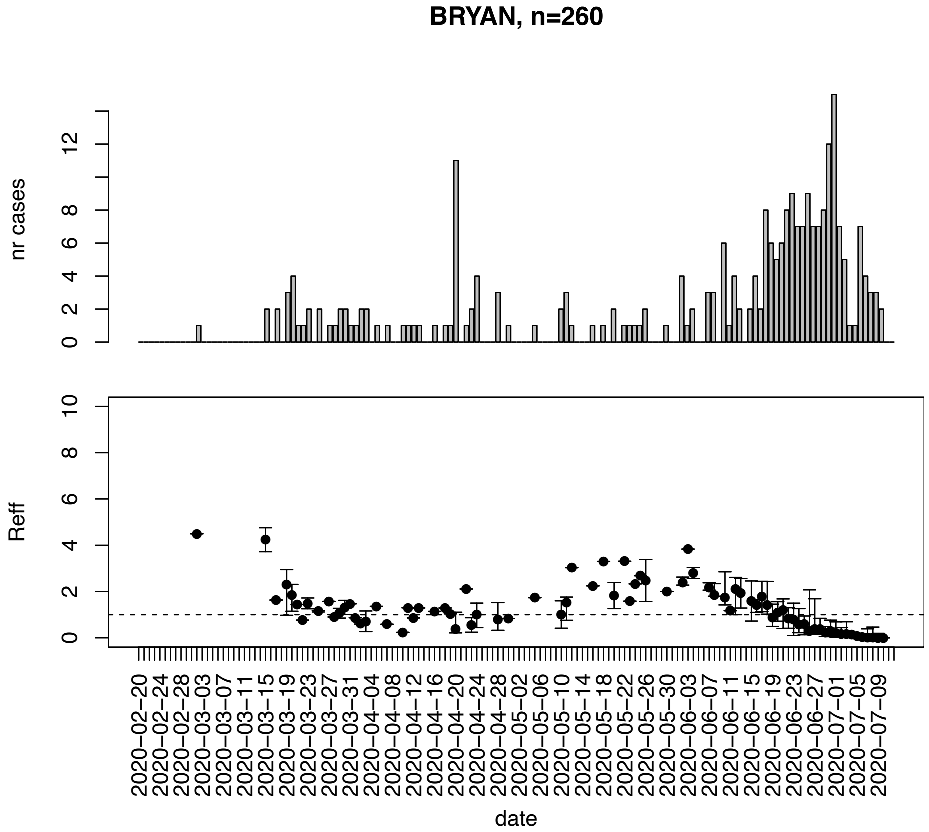


Figure. S15: Epidemic curves and reproduction number estimates until July 13th in Bryan county.

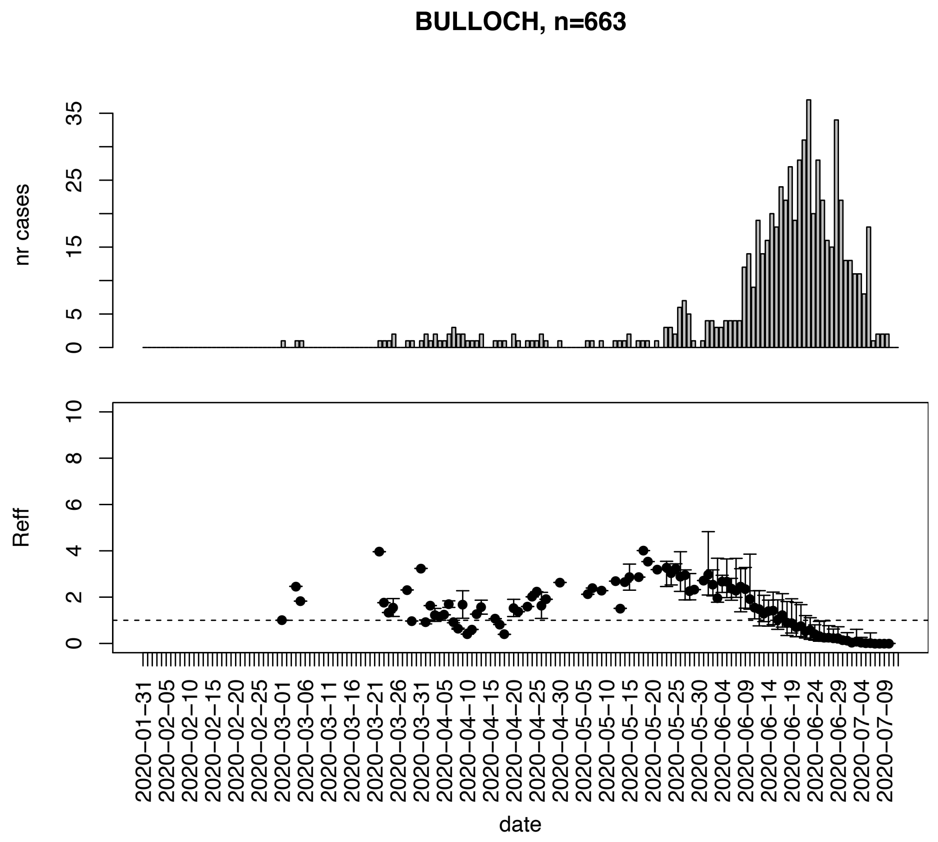


Figure. S16: Epidemic curves and reproduction number estimates until July 13th in Bulloch county.

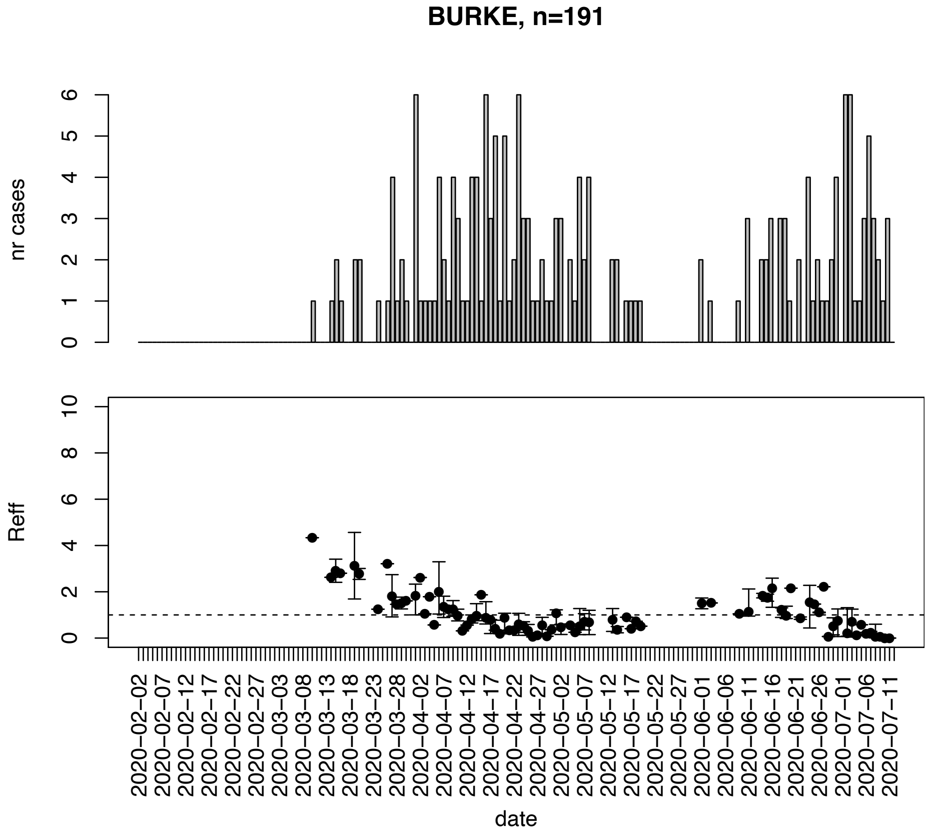


Figure. S17: Epidemic curves and reproduction number estimates until July 13th in Burke county.

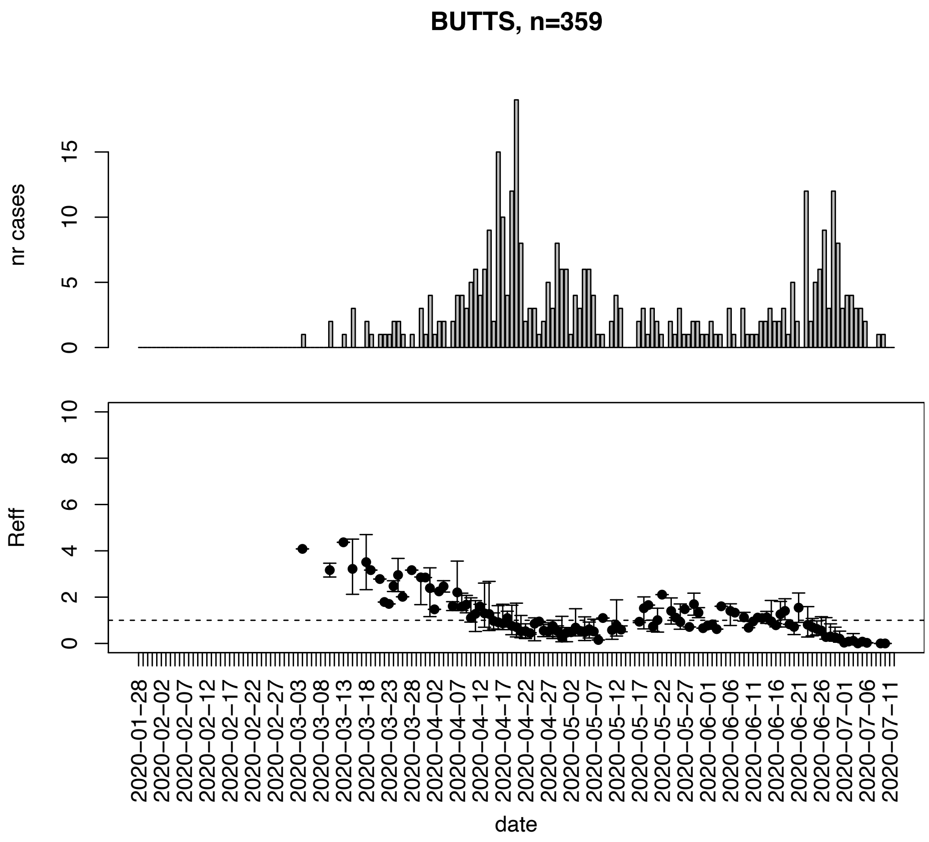


Figure. S18: Epidemic curves and reproduction number estimates until July 13th in Butts county.

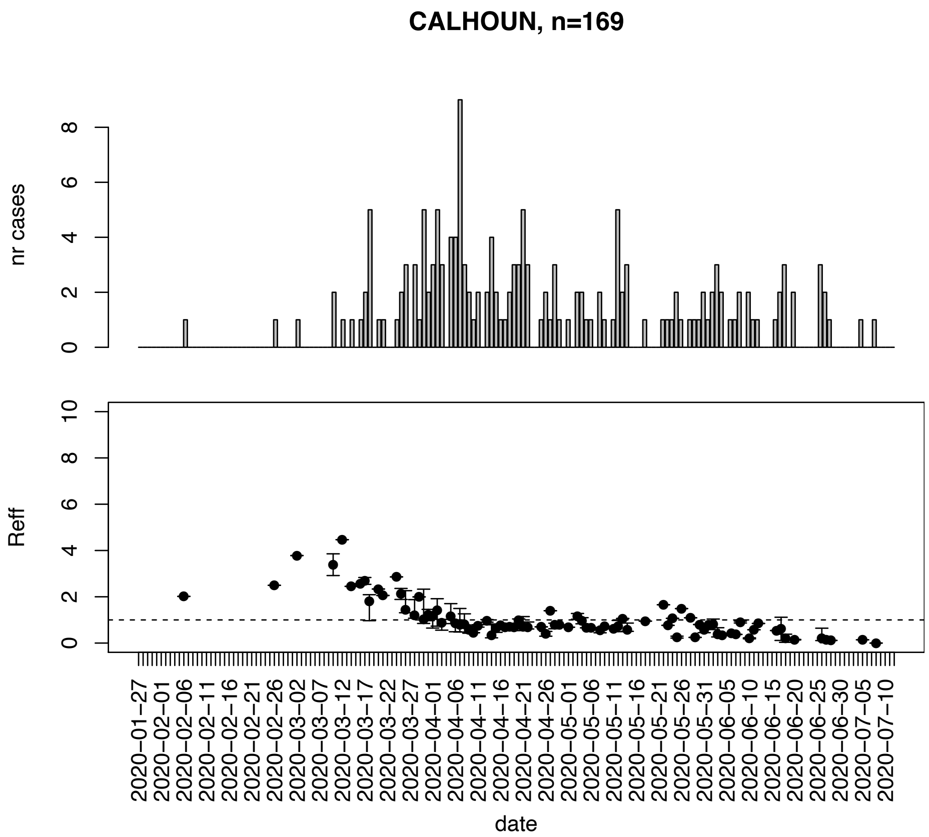


Figure. S19: Epidemic curves and reproduction number estimates until July 13th in Calhoun county.

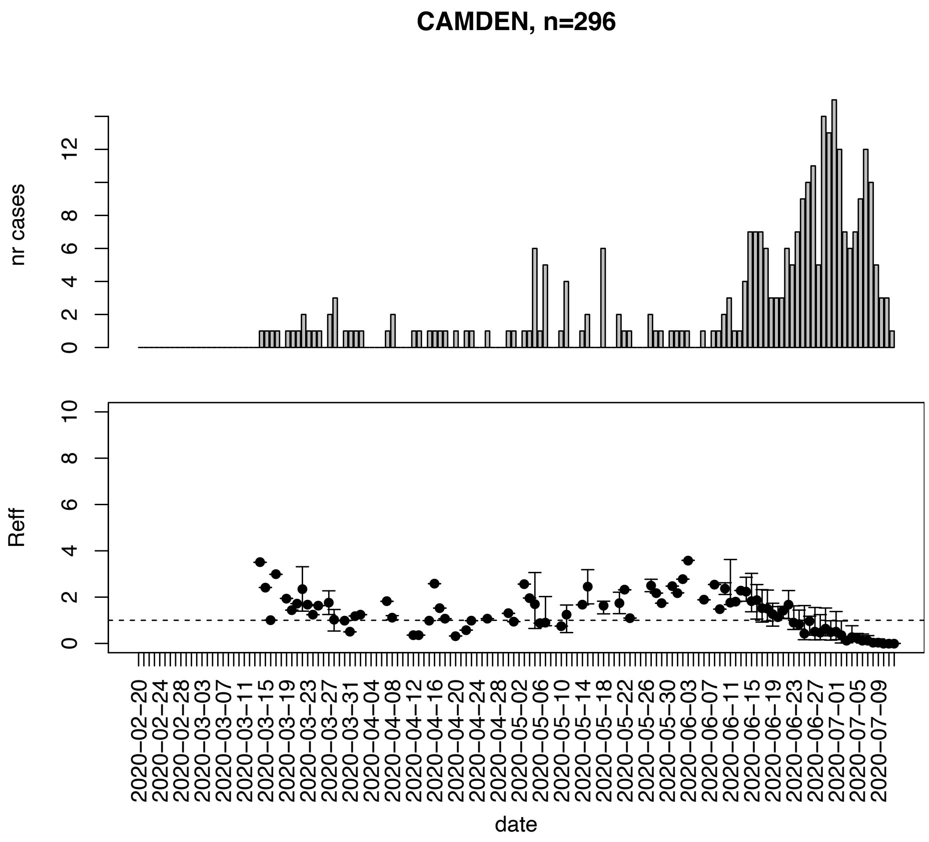


Figure. S20: Epidemic curves and reproduction number estimates until July 13th in Camden county.

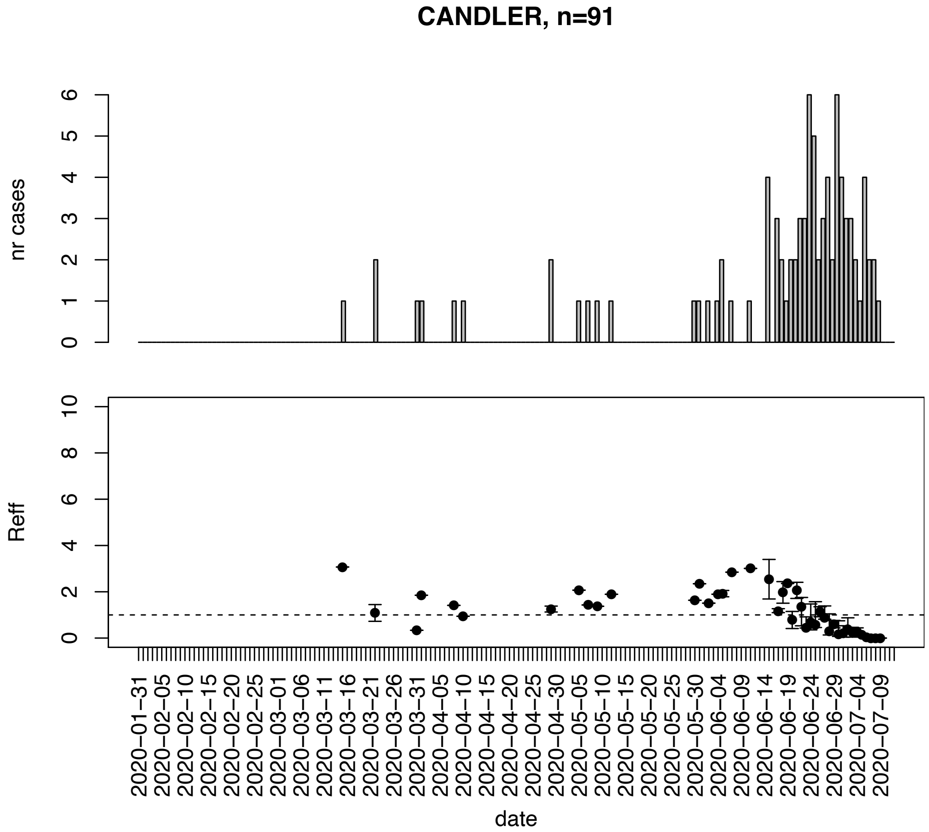


Figure. S21: Epidemic curves and reproduction number estimates until July 13th in Candler county.

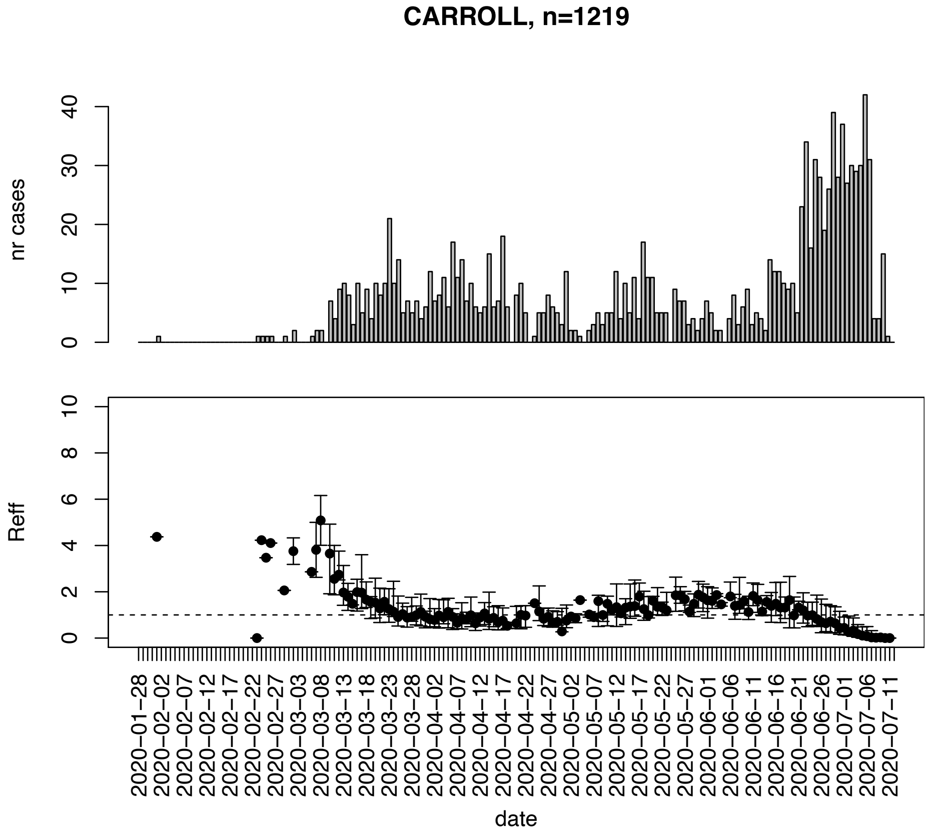


Figure. S22: Epidemic curves and reproduction number estimates until July 13th in Carroll county.

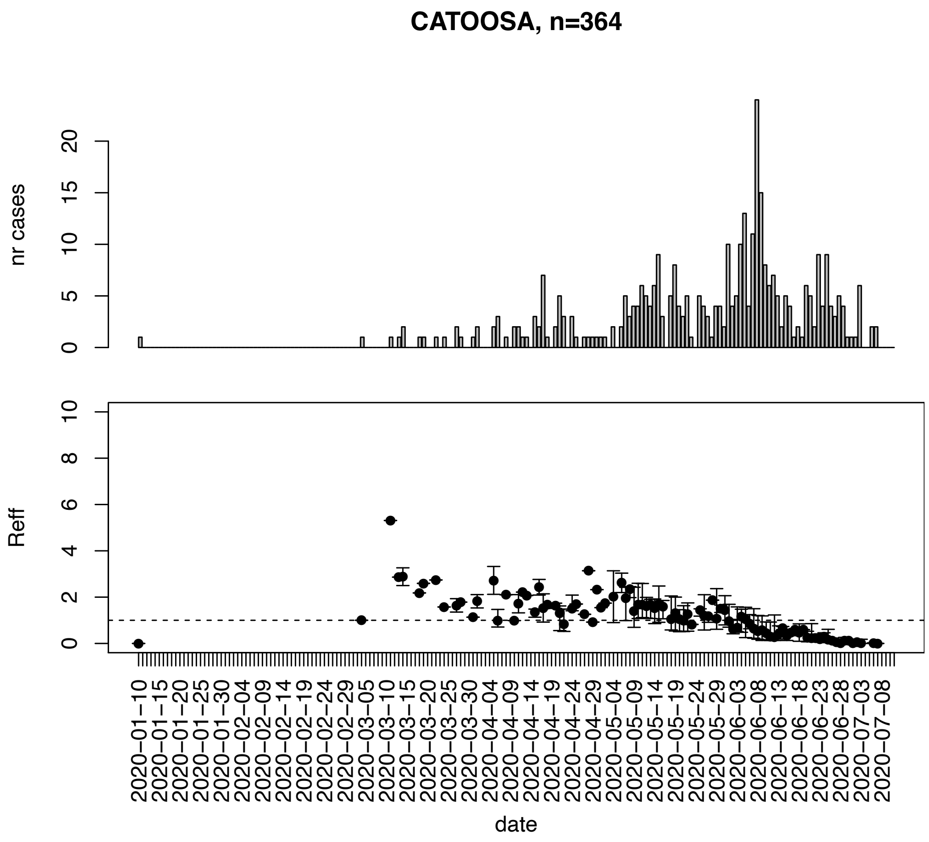


Figure. S23: Epidemic curves and reproduction number estimates until July 13th in Catoosa county.

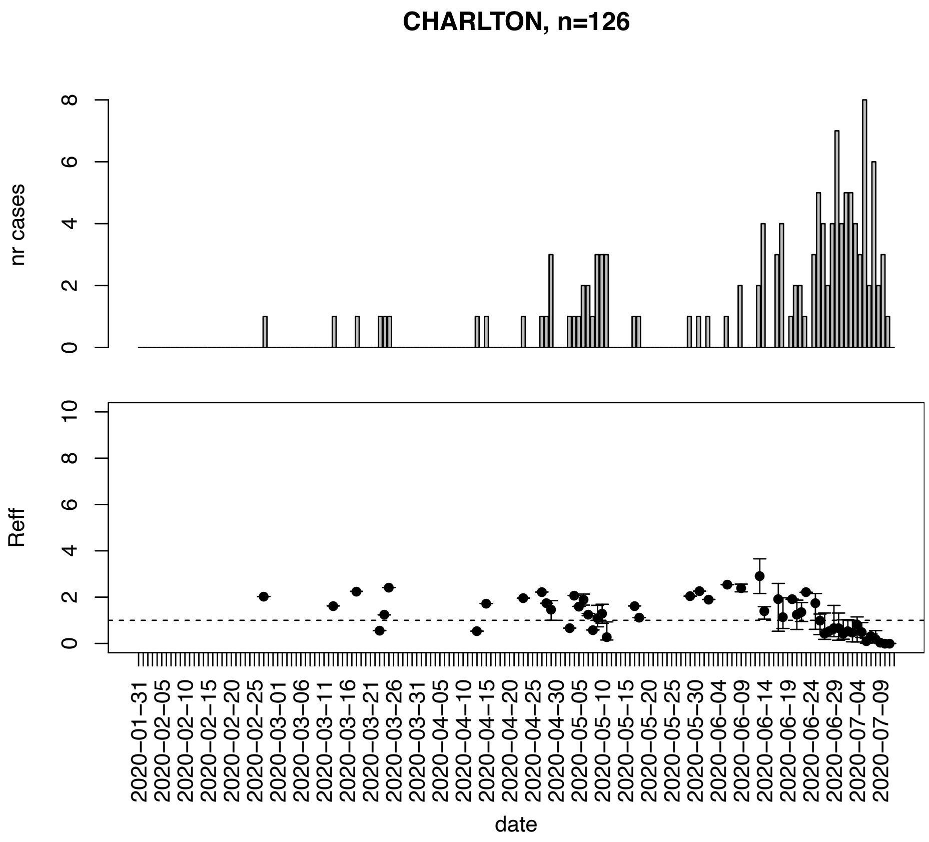


Figure. S24: Epidemic curves and reproduction number estimates until July 13th in Charlton county.

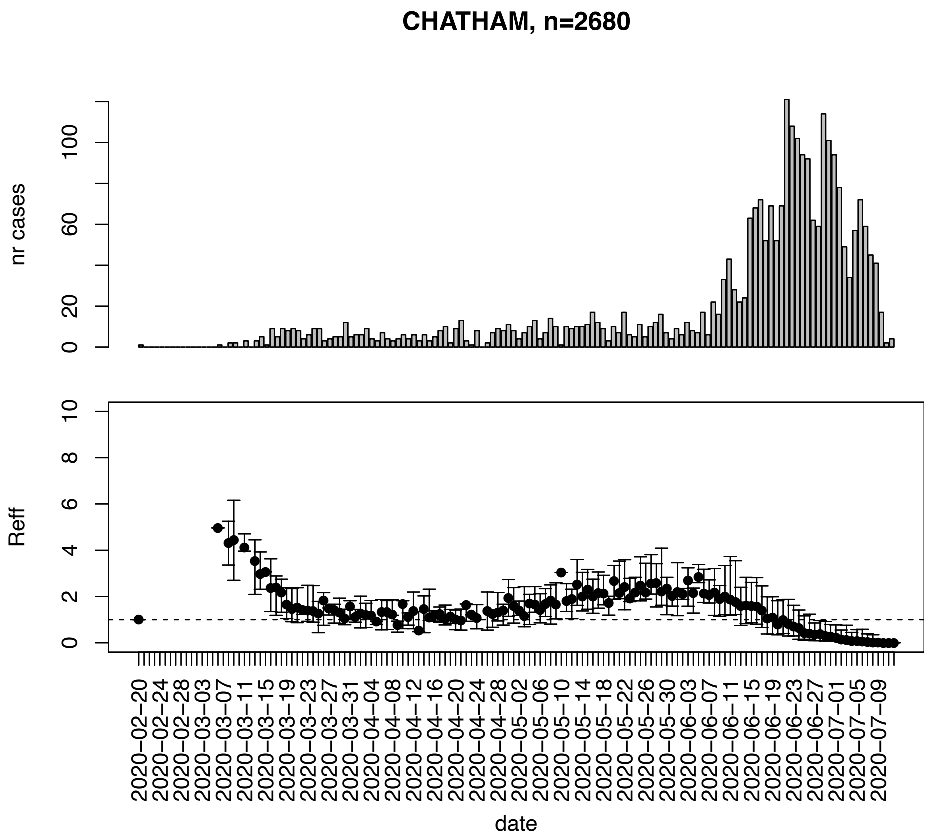


Figure. S25: Epidemic curves and reproduction number estimates until July 13th in Chatham county.

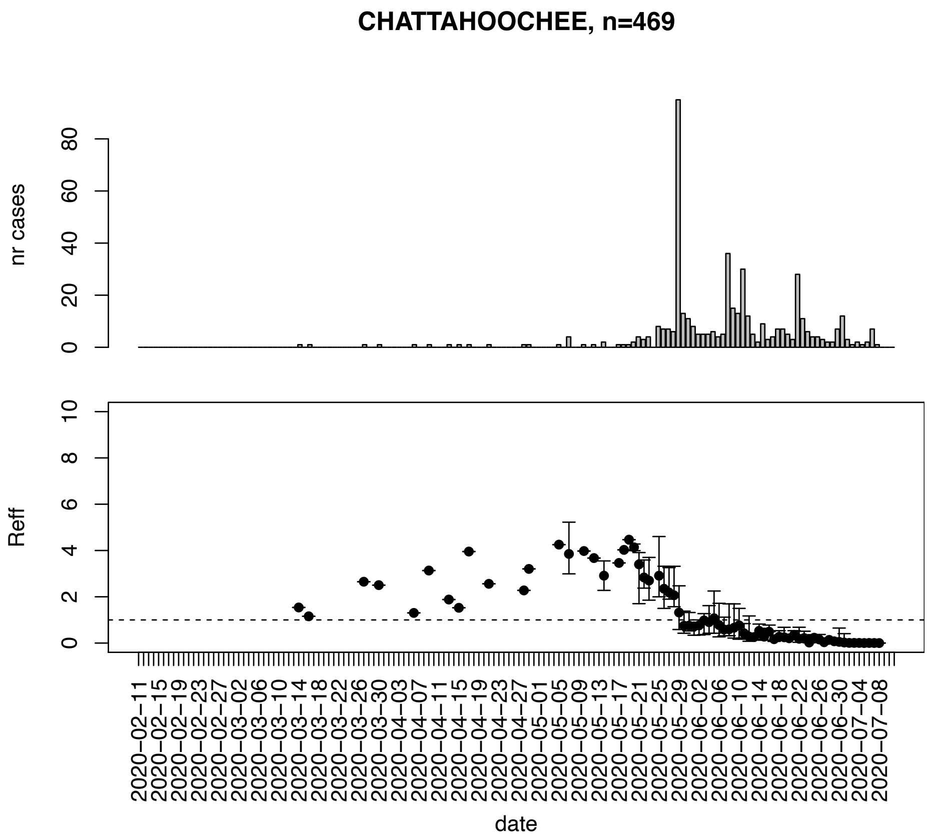


Figure. S26: Epidemic curves and reproduction number estimates until July 13th in Chattahoochee county.

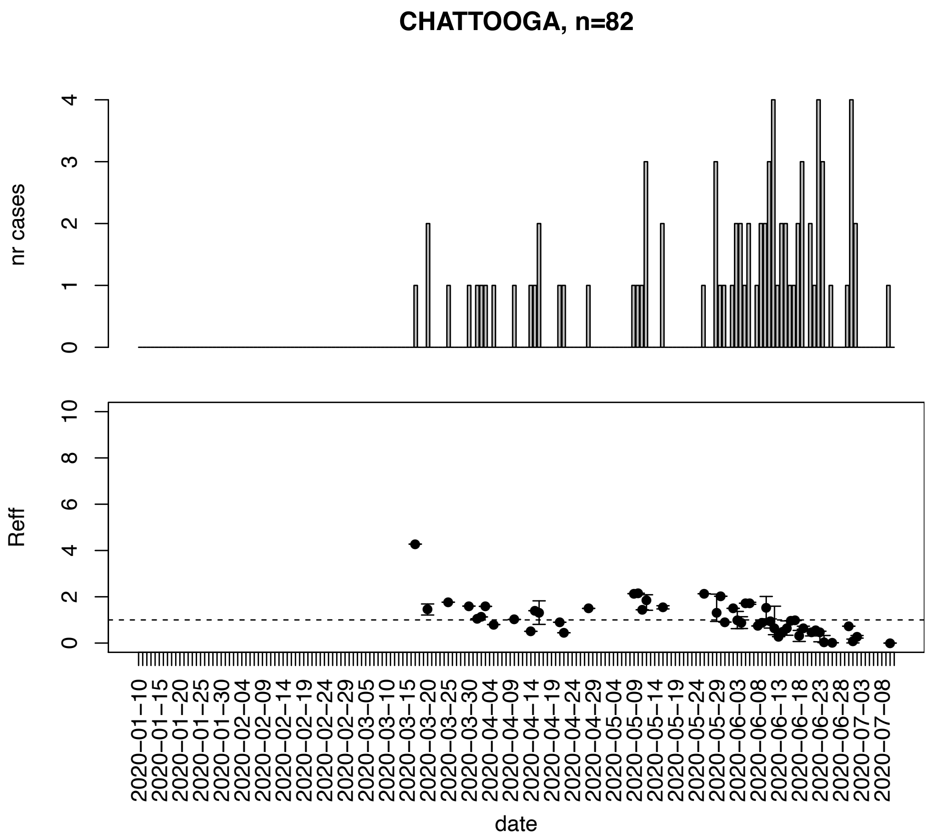


Figure. S27: Epidemic curves and reproduction number estimates until July 13th in Chattooga county.

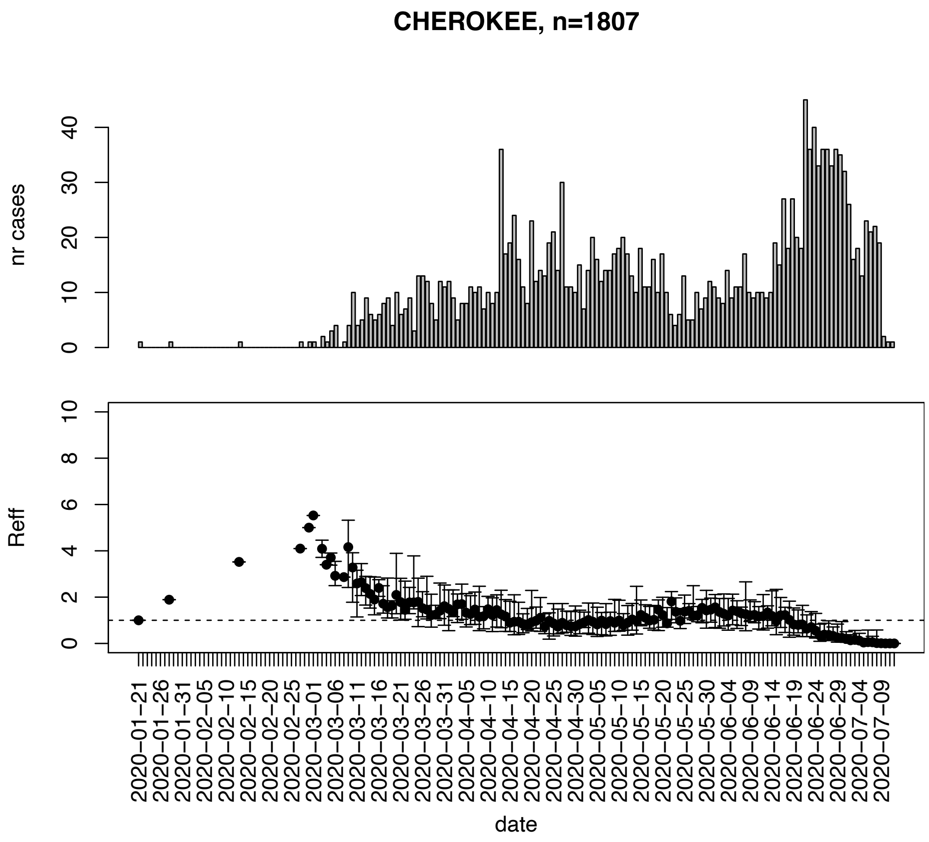


Figure. S28: Epidemic curves and reproduction number estimates until July 13th in Cherokee county.

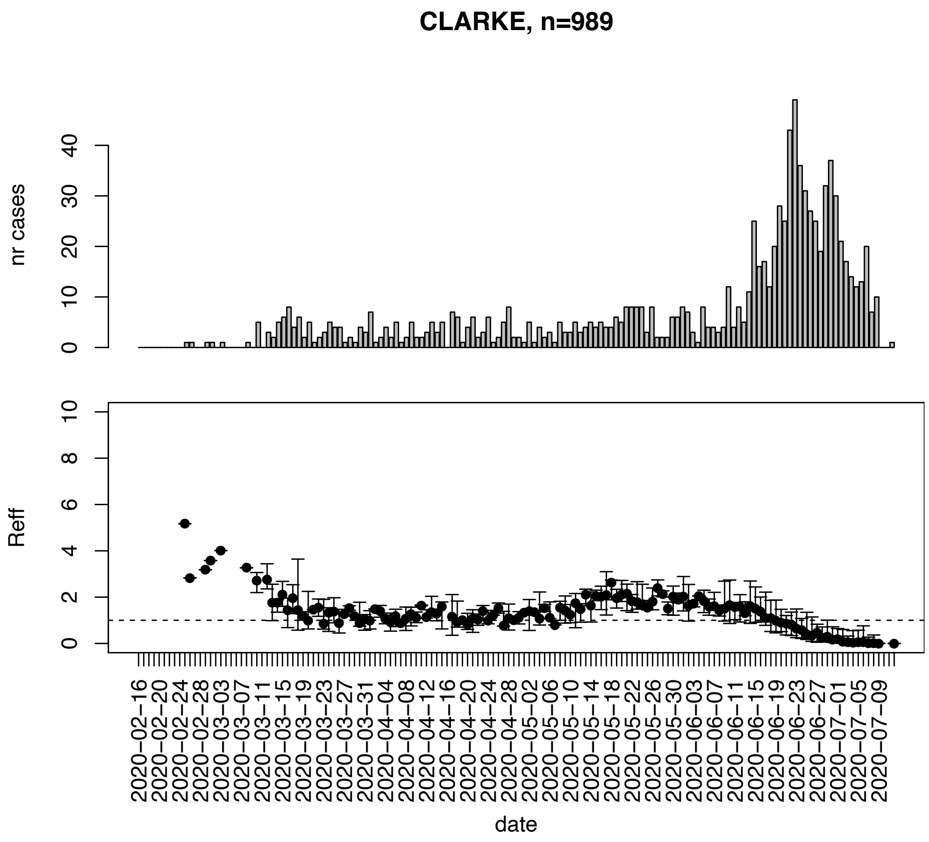


Figure. S29: Epidemic curves and reproduction number estimates until July 13th in Clarke county.

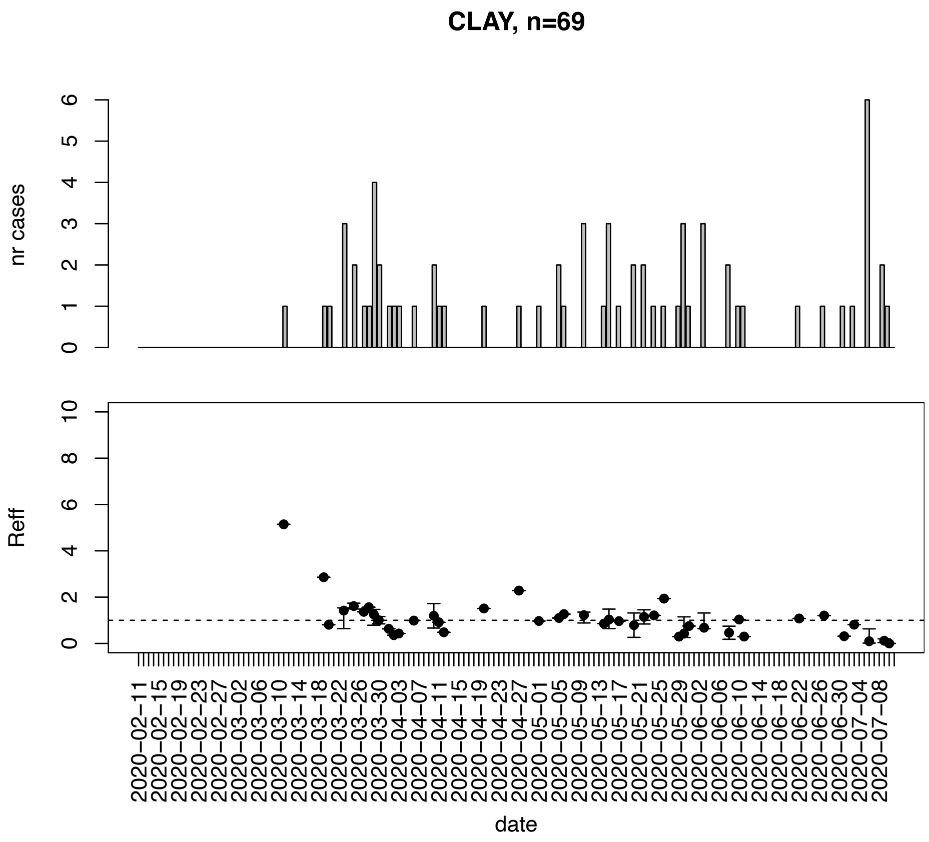


Figure. S30: Epidemic curves and reproduction number estimates until July 13th in Clay county.

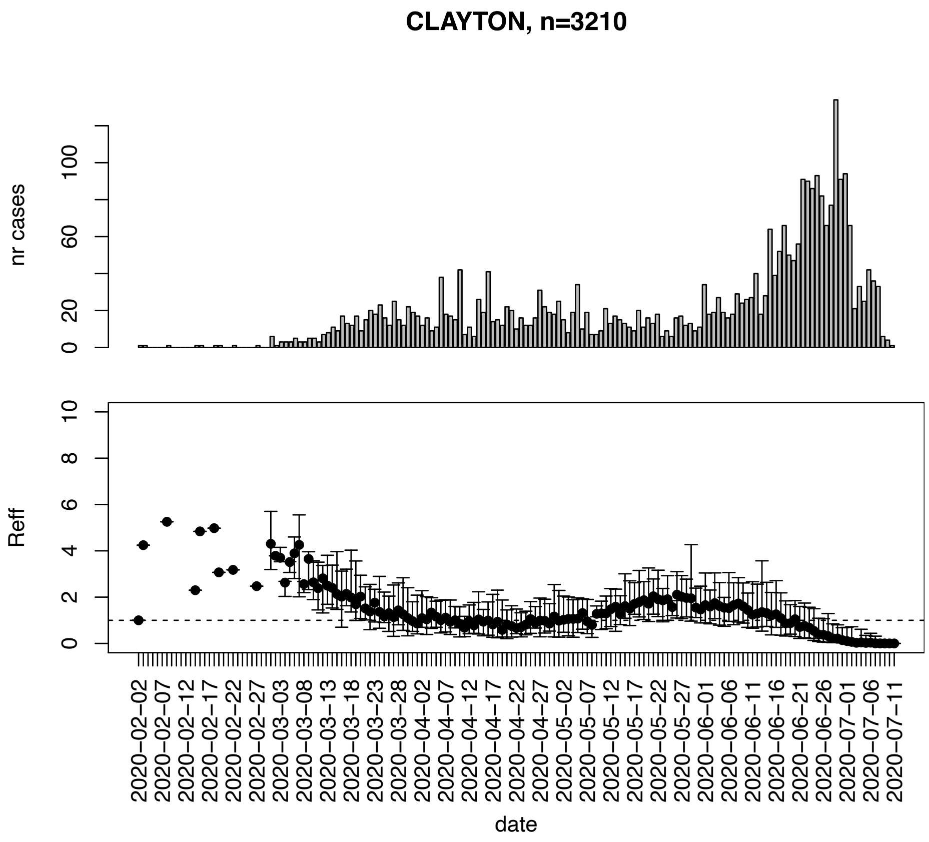


Figure. S31: Epidemic curves and reproduction number estimates until July 13th in Clayton county.

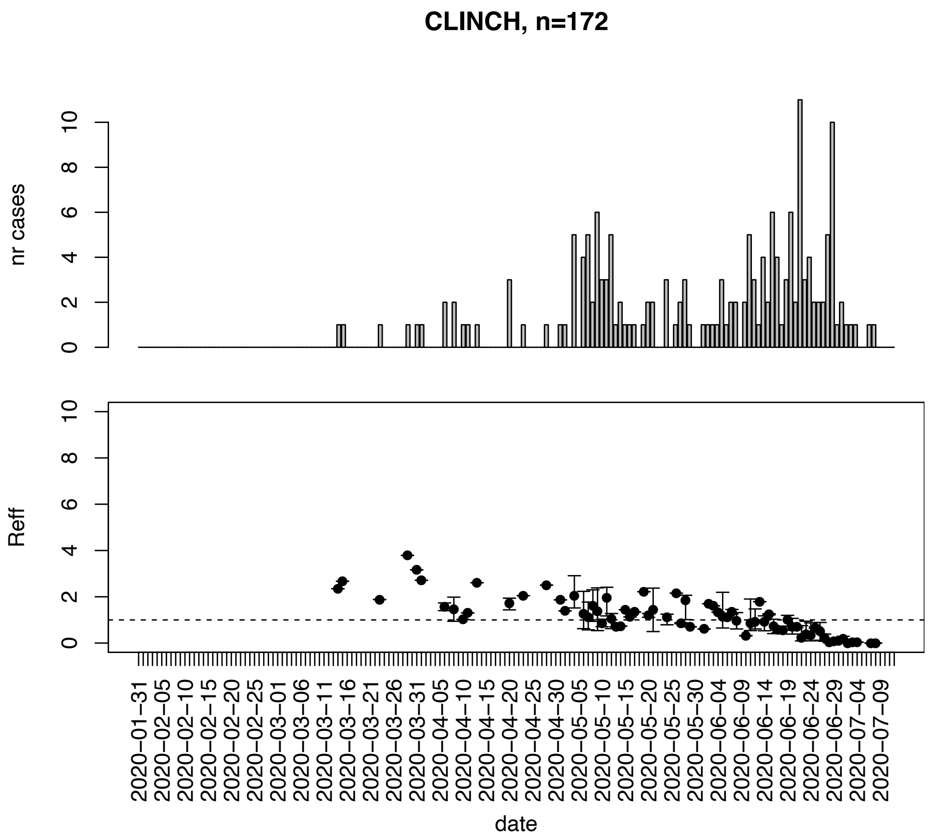


Figure. S32: Epidemic curves and reproduction number estimates until July 13th in Clinch county.

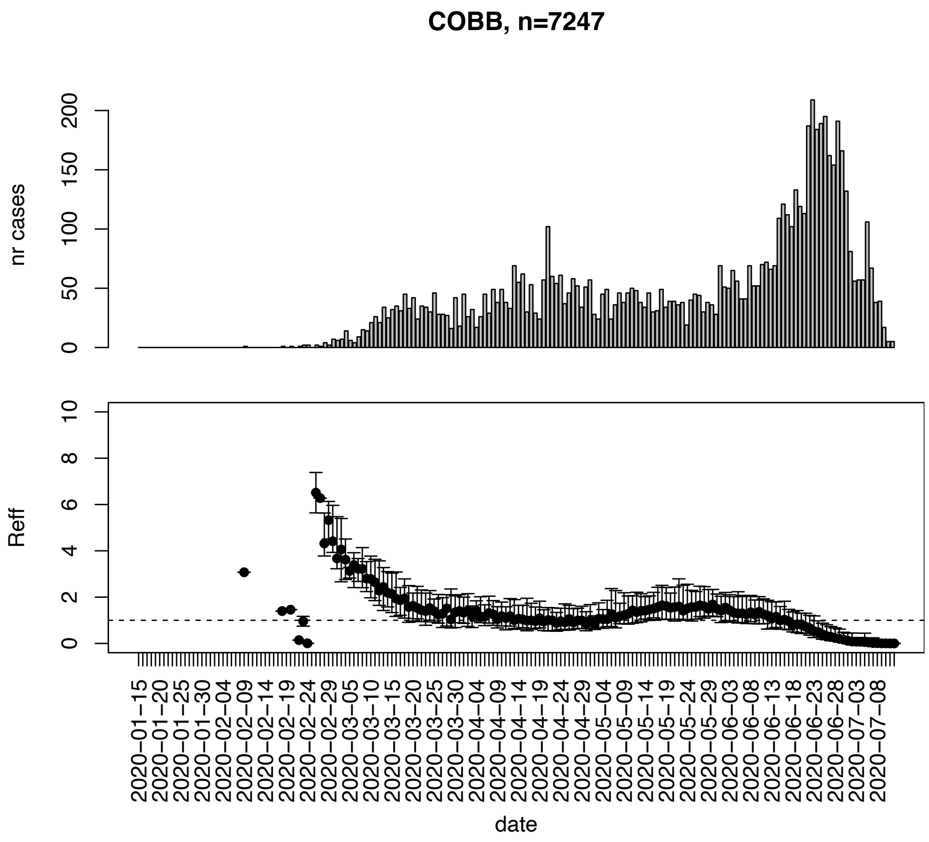


Figure. S33: Epidemic curves and reproduction number estimates until July 13th in Cobb county.

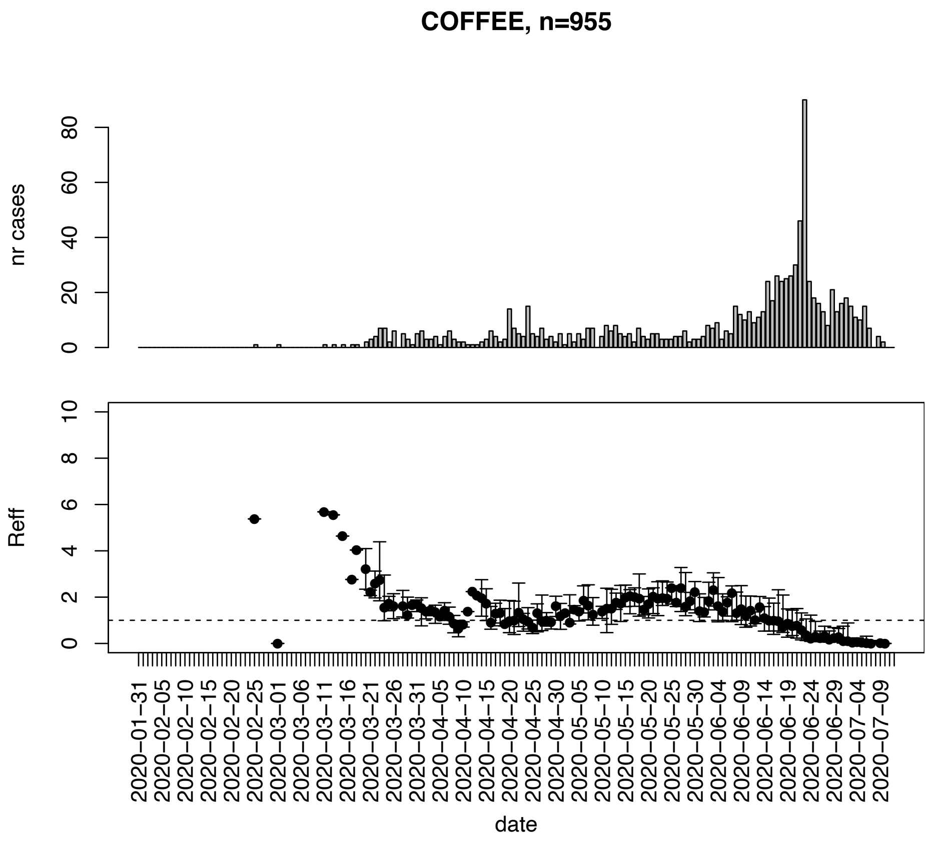


Figure. S34: Epidemic curves and reproduction number estimates until July 13th in Coffee county.

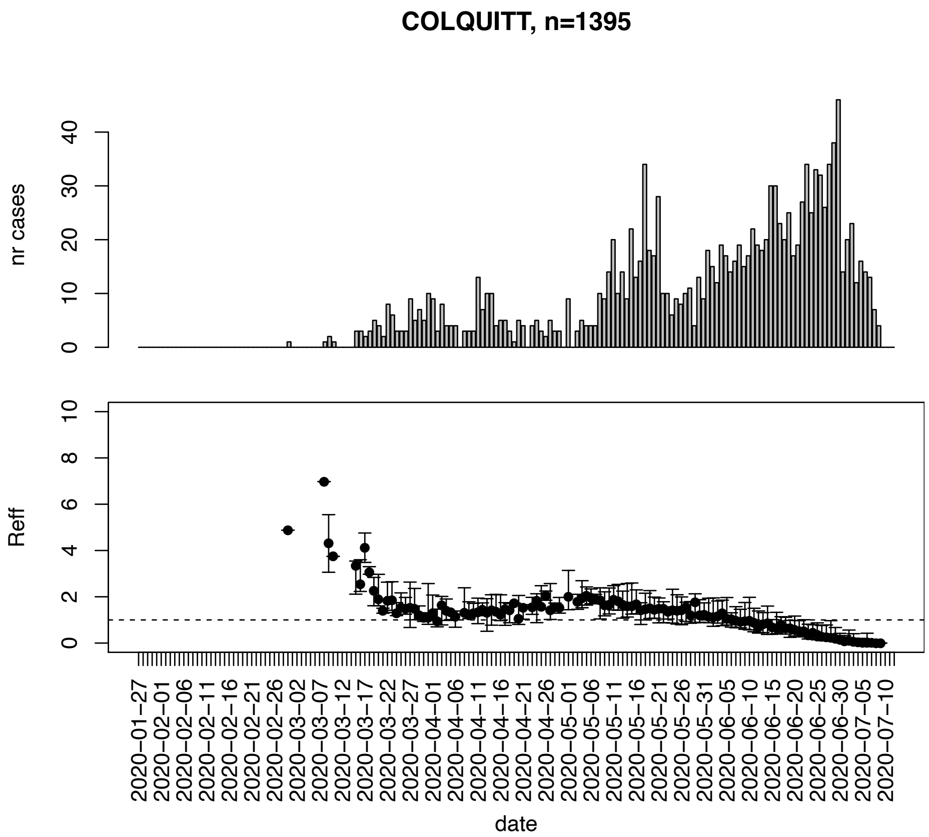


Figure. S35: Epidemic curves and reproduction number estimates until July 13th in Colquitt county.

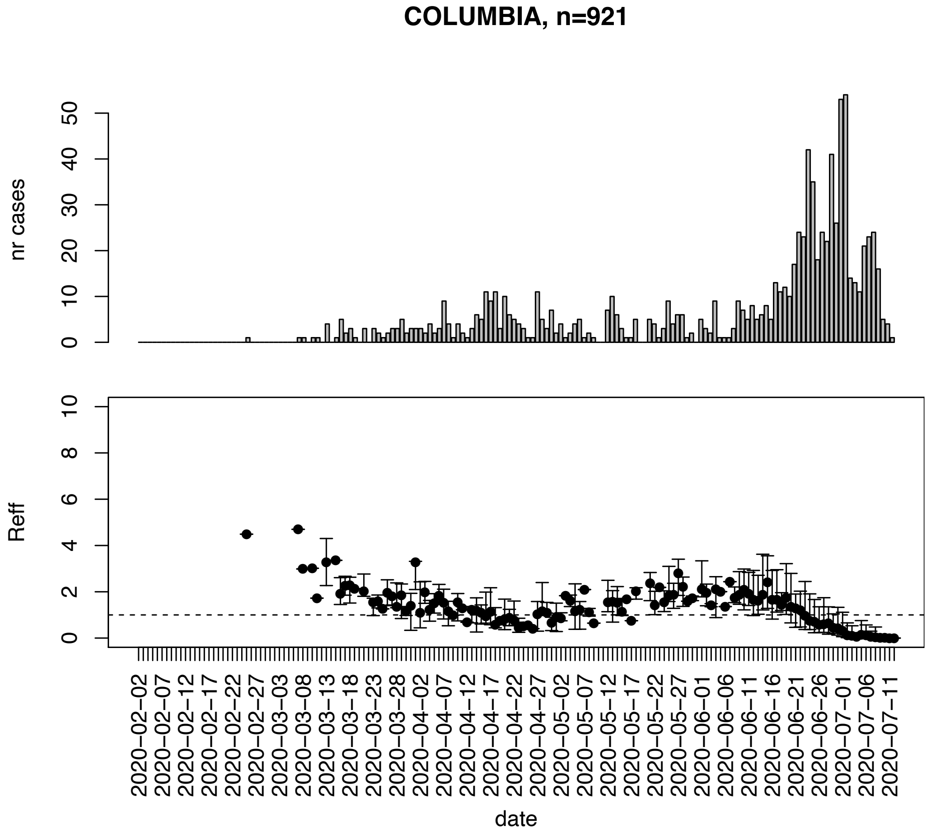


Figure. S36: Epidemic curves and reproduction number estimates until July 13th in Columbia county.

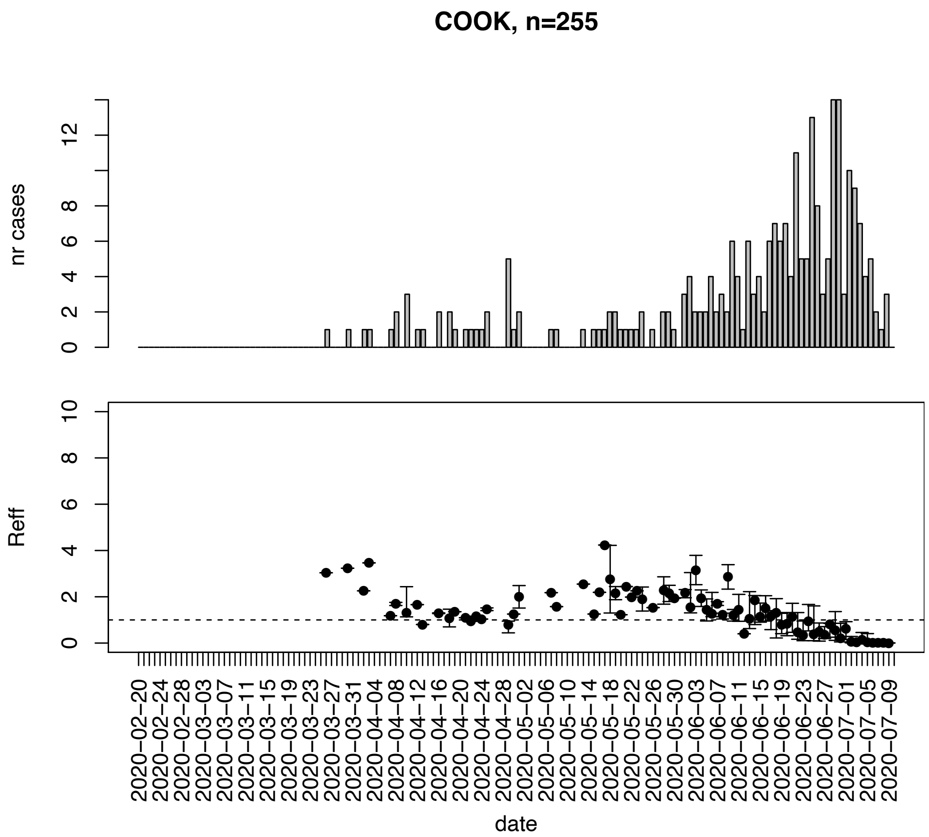


Figure. S37: Epidemic curves and reproduction number estimates until July 13th in Cook county.

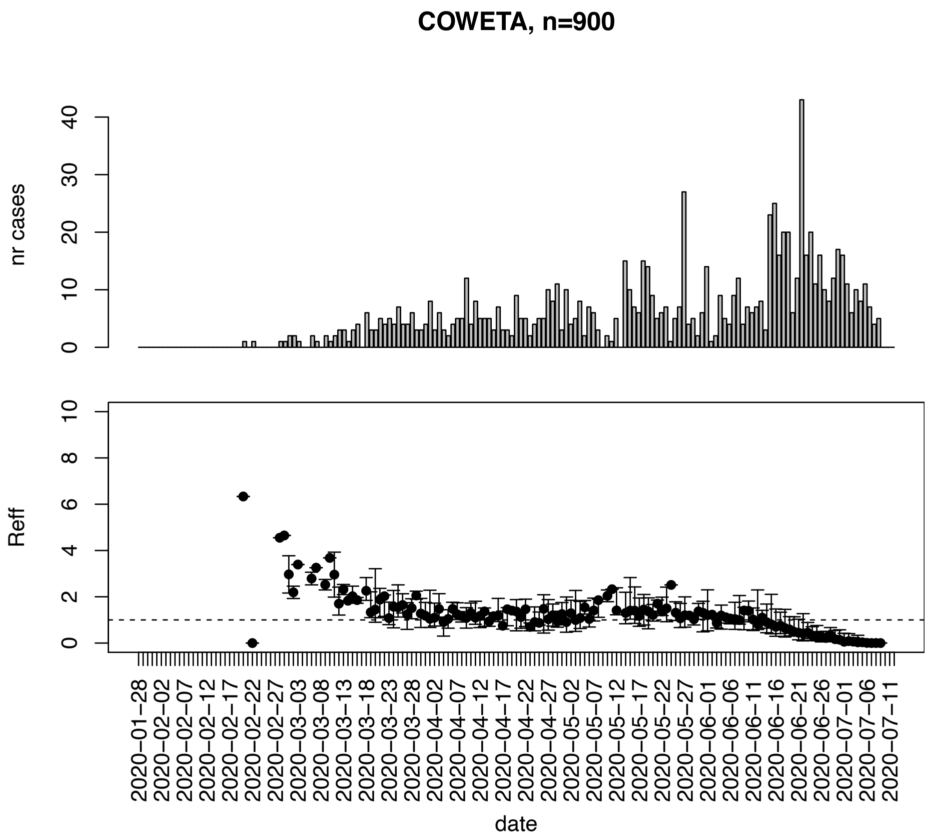


Figure. S38: Epidemic curves and reproduction number estimates until July 13th in Coweta county.

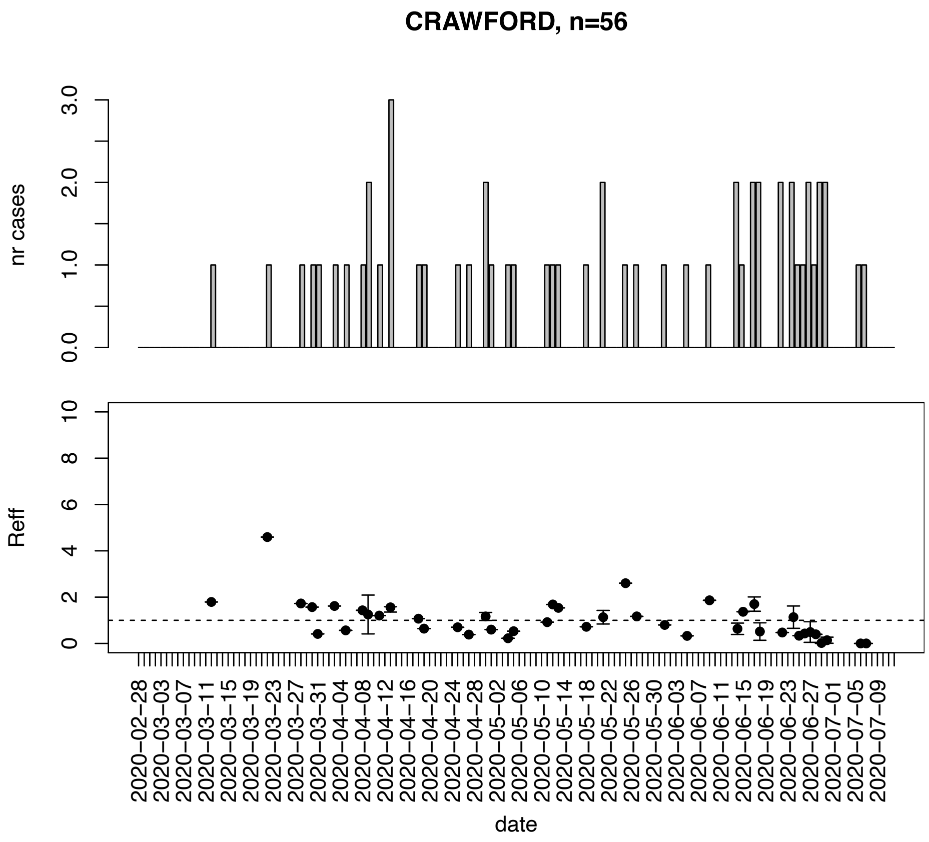


Figure. S39: Epidemic curves and reproduction number estimates until July 13th in Crawford county.

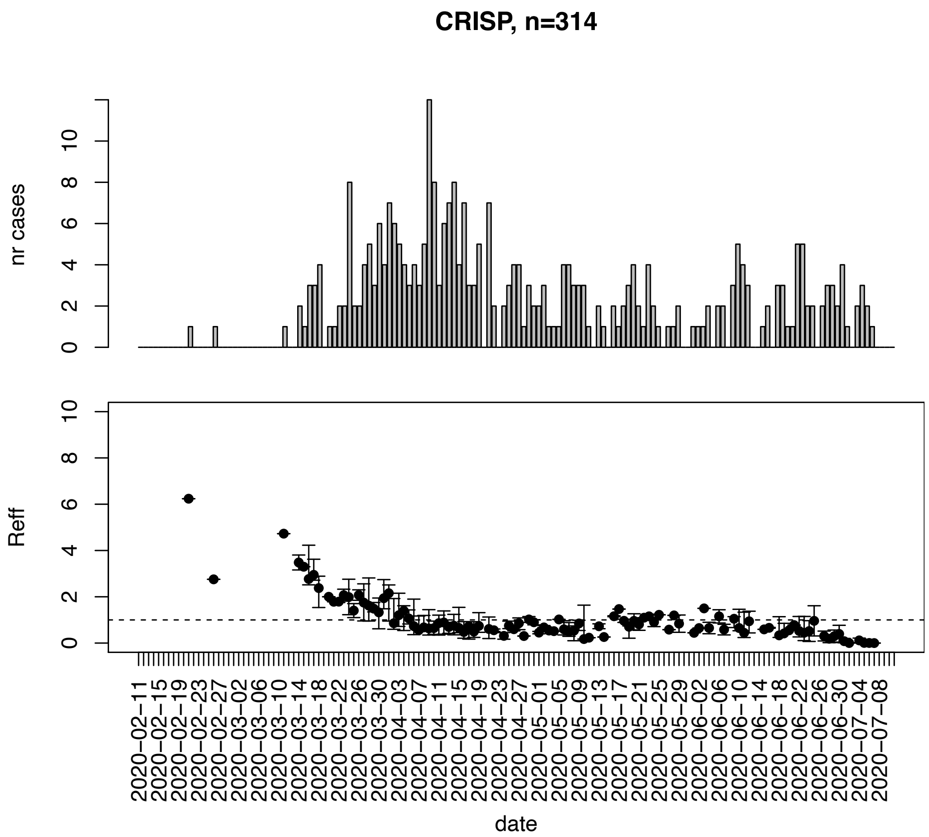


Figure. S40: Epidemic curves and reproduction number estimates until July 13th in Crisp county.

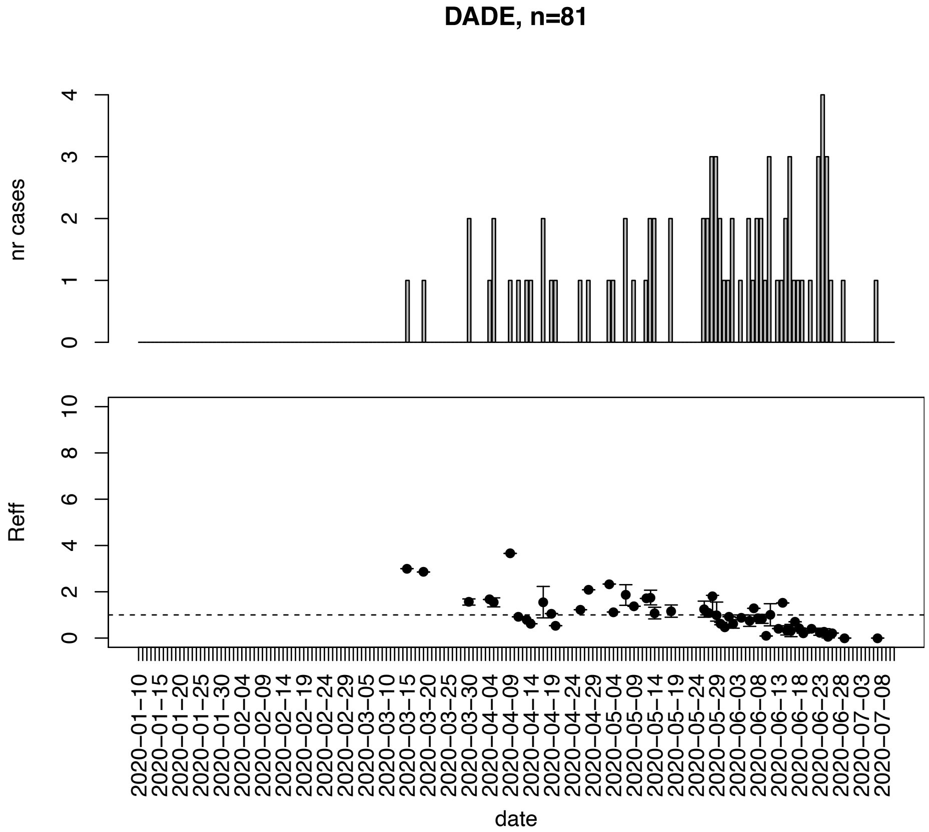


Figure. S41: Epidemic curves and reproduction number estimates until July 13th in Dade county.

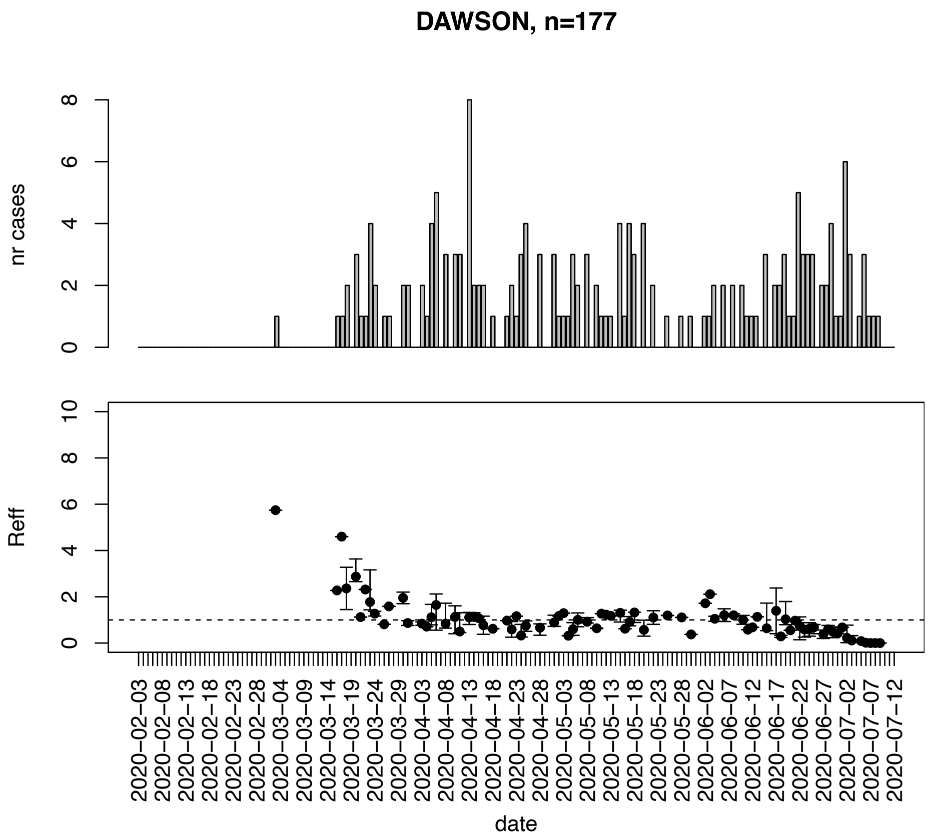


Figure. S42: Epidemic curves and reproduction number estimates until July 13th in Dawson county.

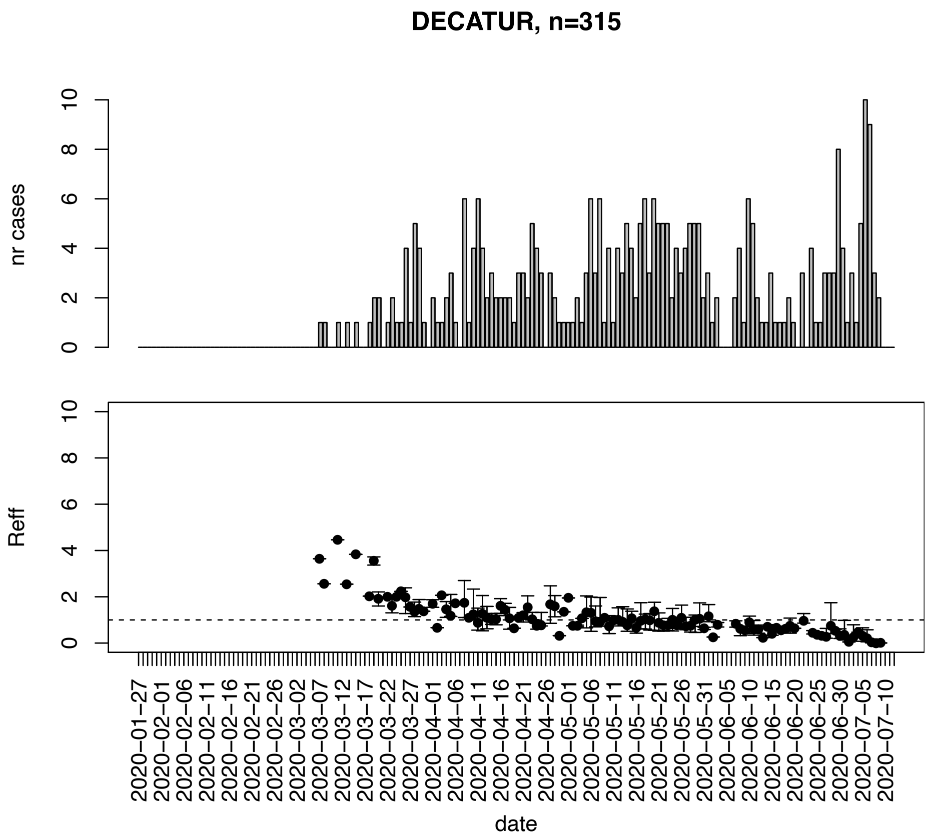


Figure. S43: Epidemic curves and reproduction number estimates until July 13th in Decatur county.

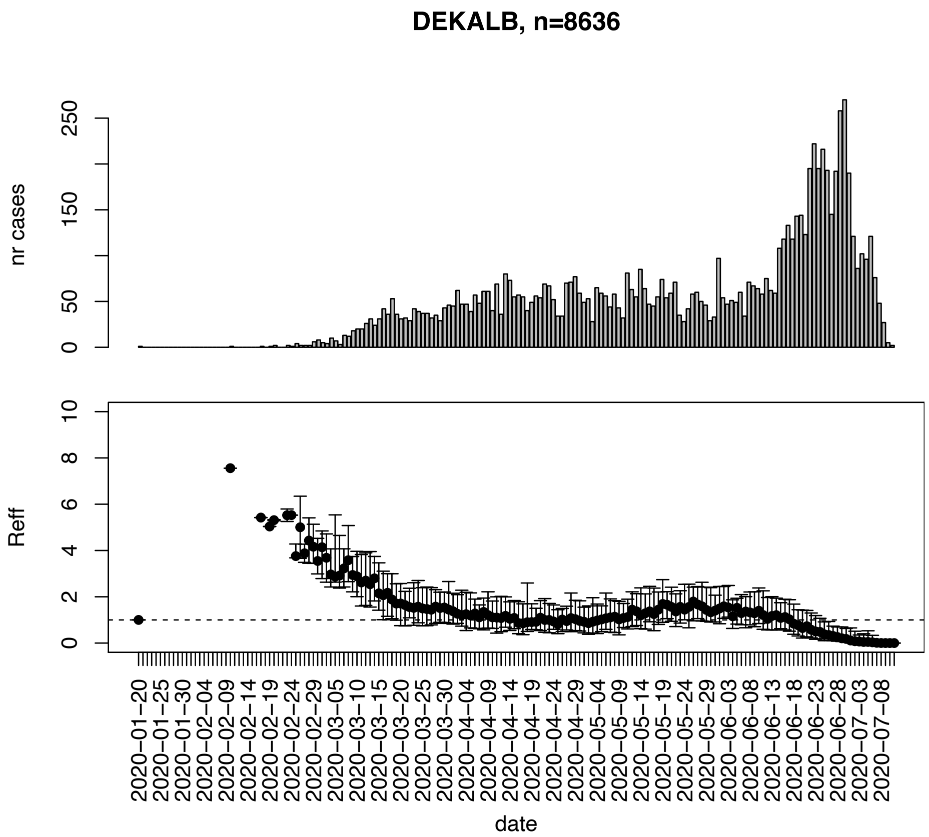


Figure. S44: Epidemic curves and reproduction number estimates until July 13th in Dekalb county.

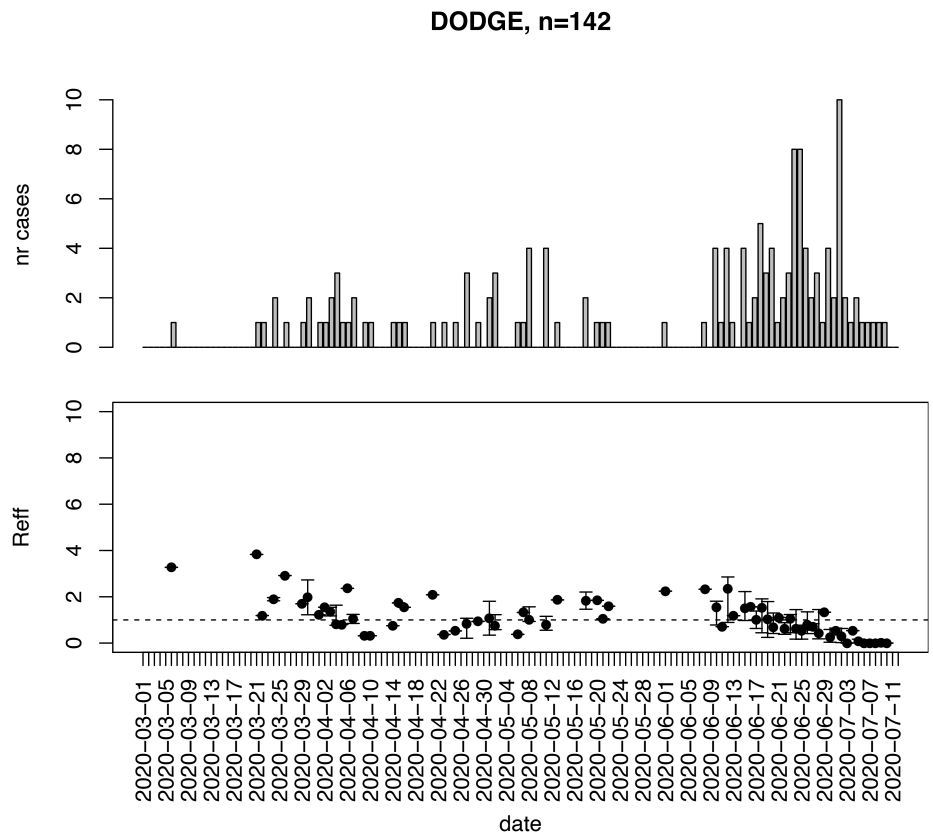


Figure. S45: Epidemic curves and reproduction number estimates until July 13th in Dodge county.

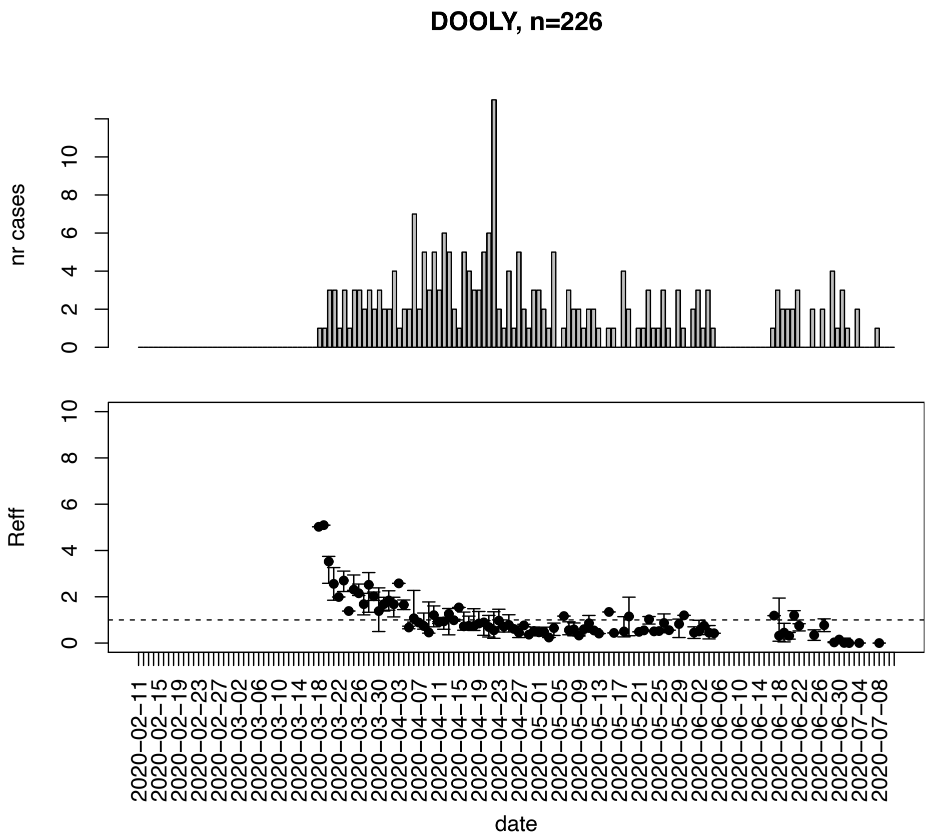


Figure. S46: Epidemic curves and reproduction number estimates until July 13th in Dooly county.

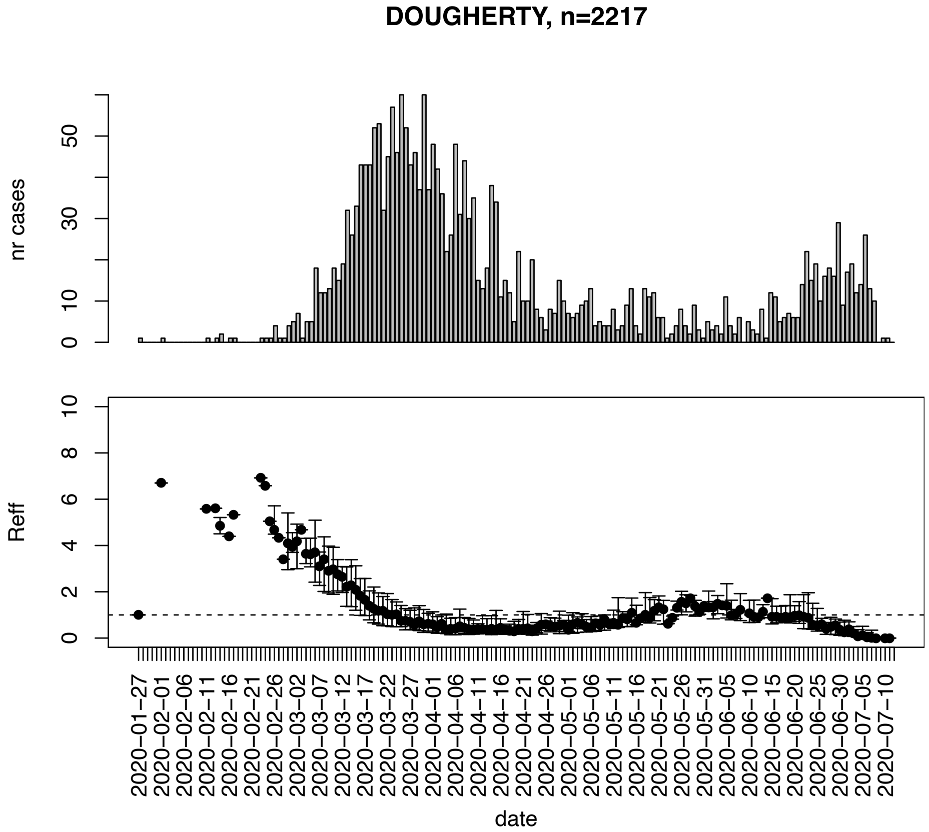


Figure. S47: Epidemic curves and reproduction number estimates until July 13th in Dougherty county.

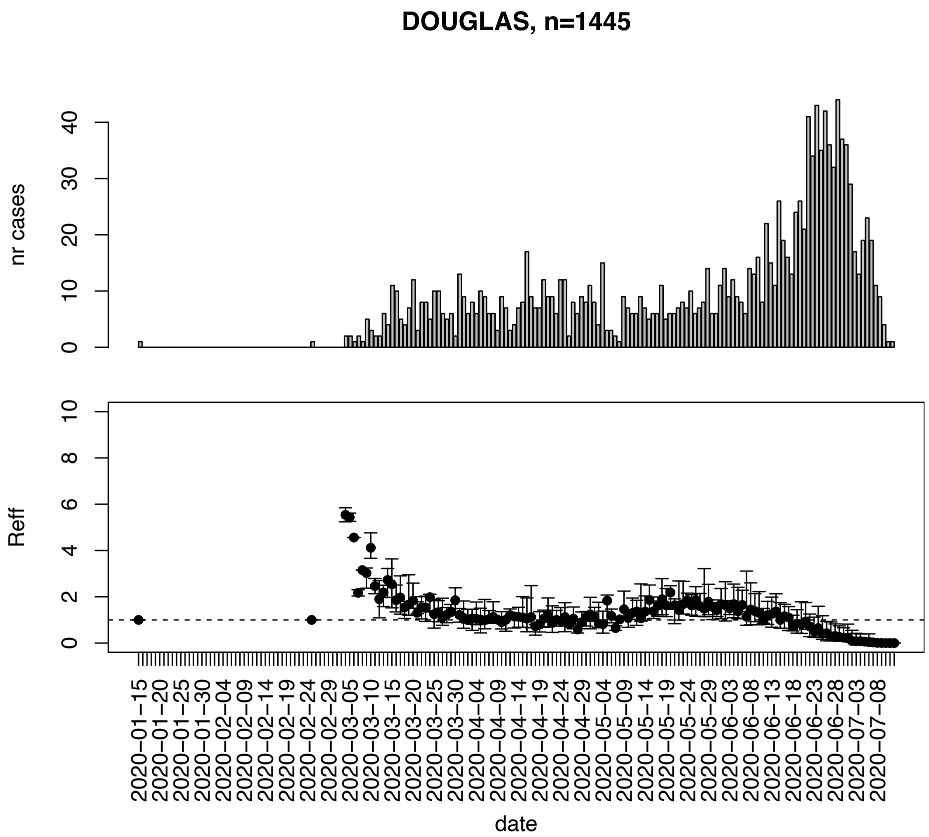


Figure. S48: Epidemic curves and reproduction number estimates until July 13th in Douglas county.

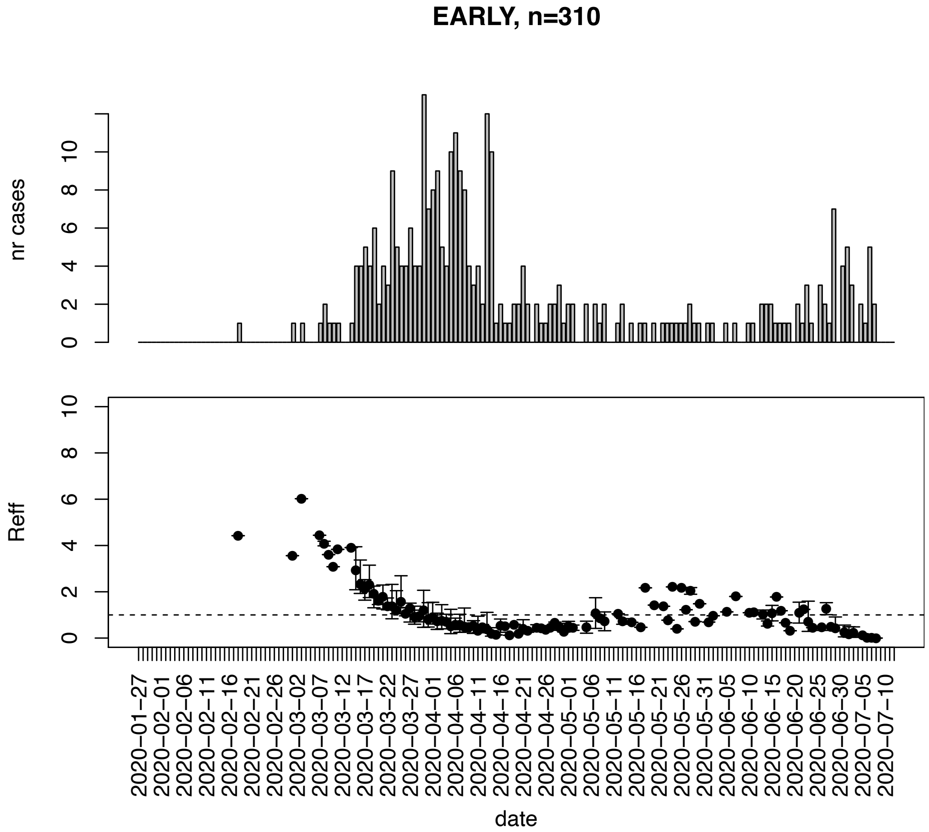


Figure. S49: Epidemic curves and reproduction number estimates until July 13th in Early county.

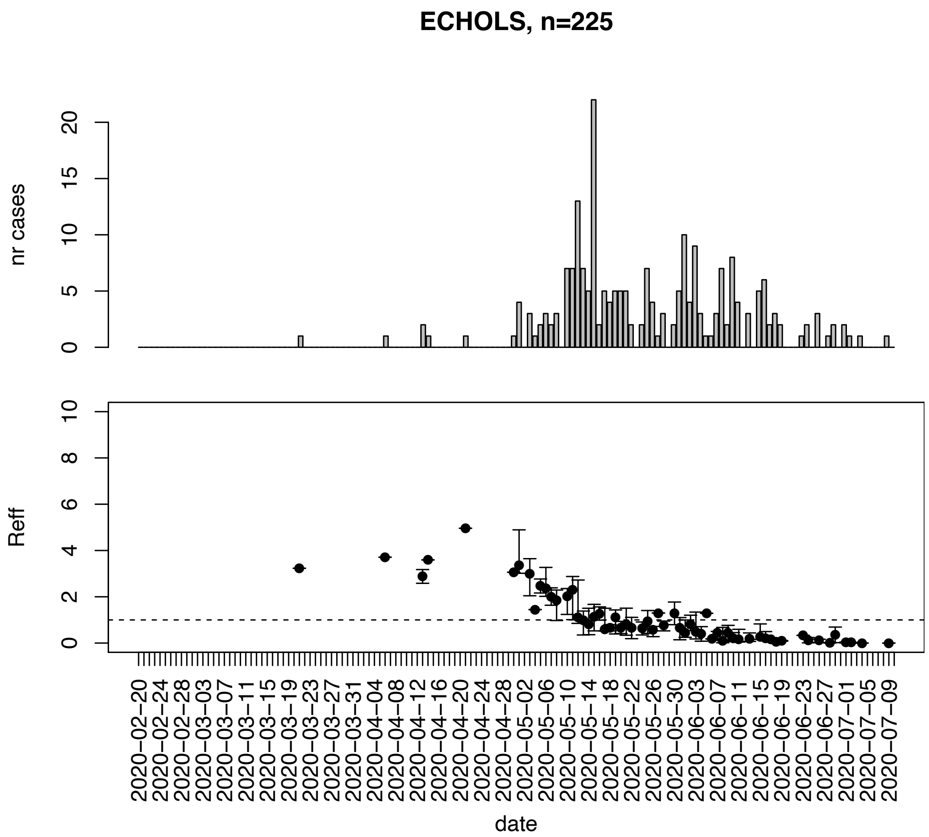


Figure. S50: Epidemic curves and reproduction number estimates until July 13th in Echols county.

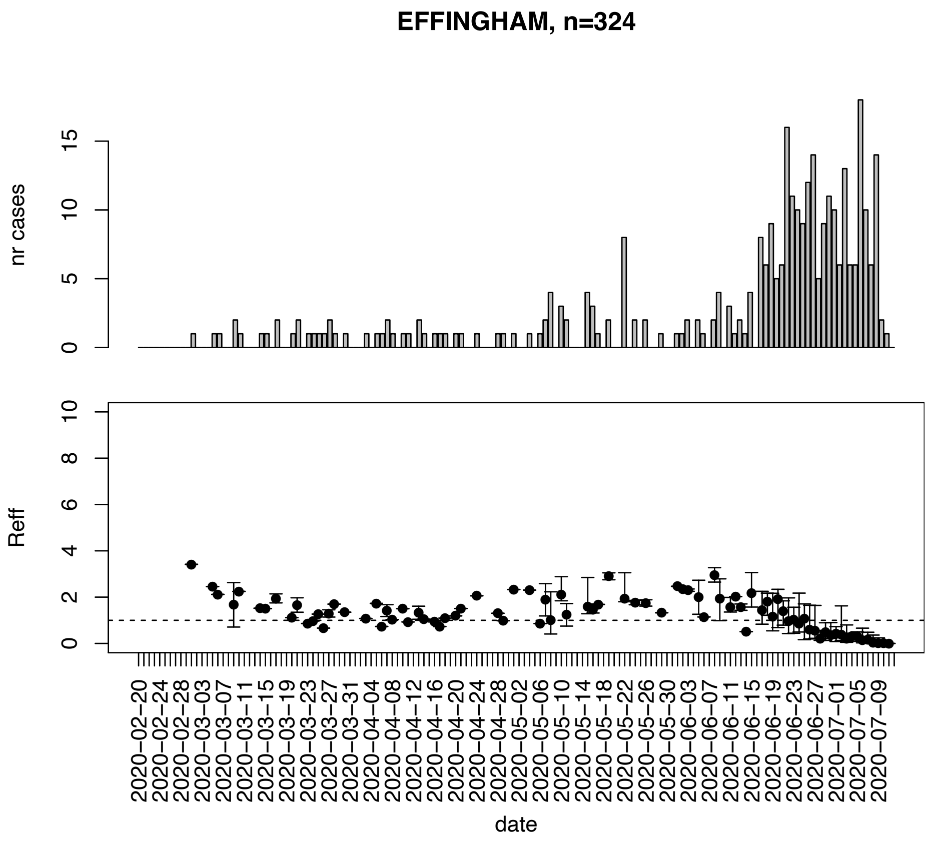


Figure. S51: Epidemic curves and reproduction number estimates until July 13th in Effingham county.

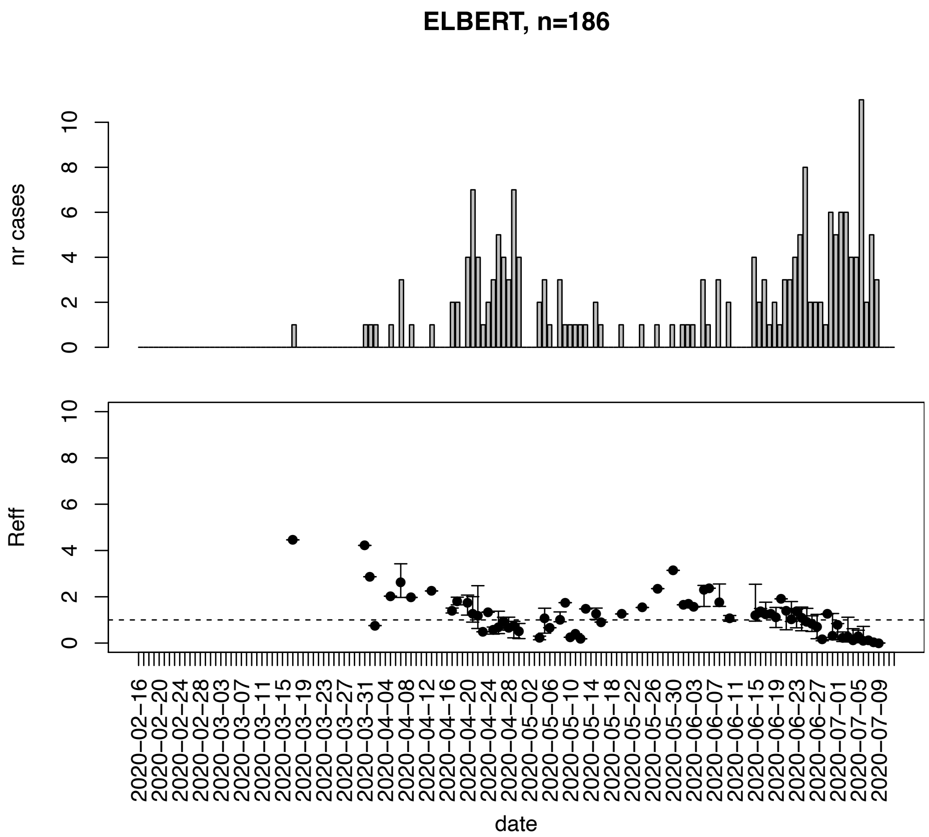


Figure. S52: Epidemic curves and reproduction number estimates until July 13th in Elbert county.

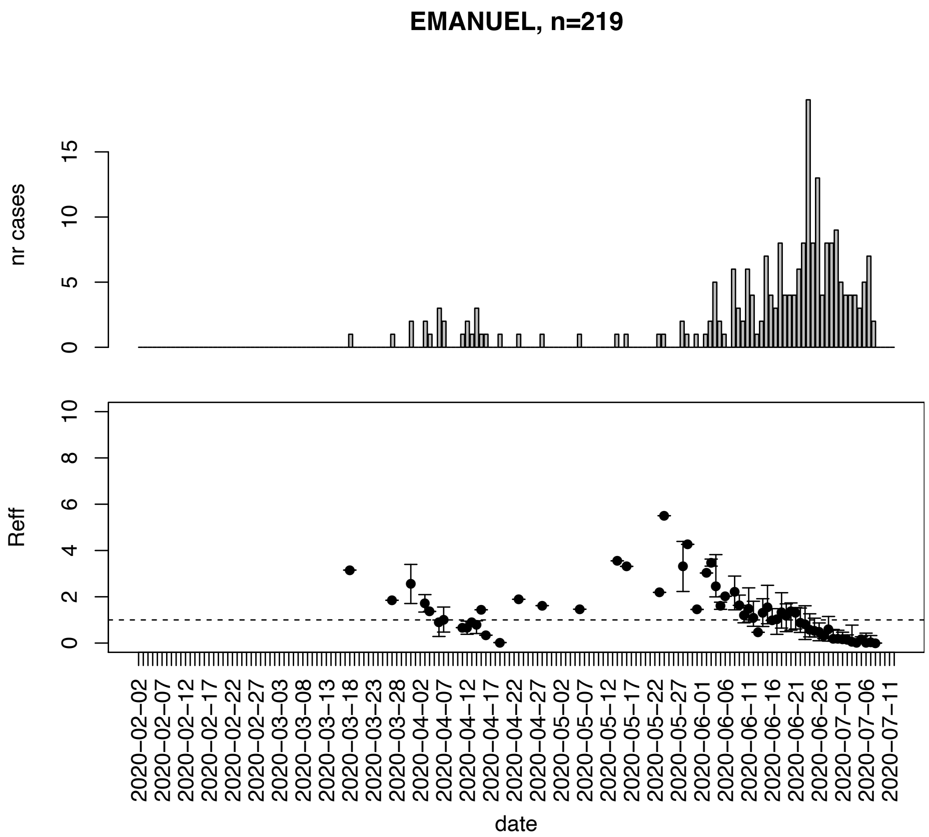


Figure. S53: Epidemic curves and reproduction number estimates until July 13th in Emanuel county.

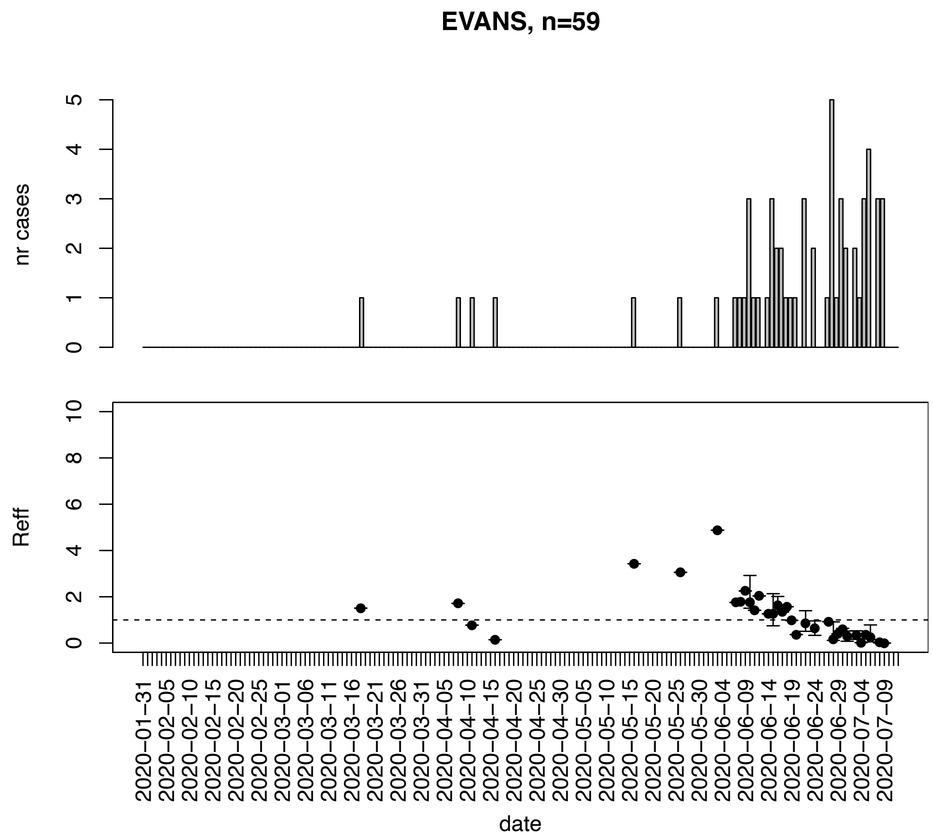


Figure. S54: Epidemic curves and reproduction number estimates until July 13th in Evans county.

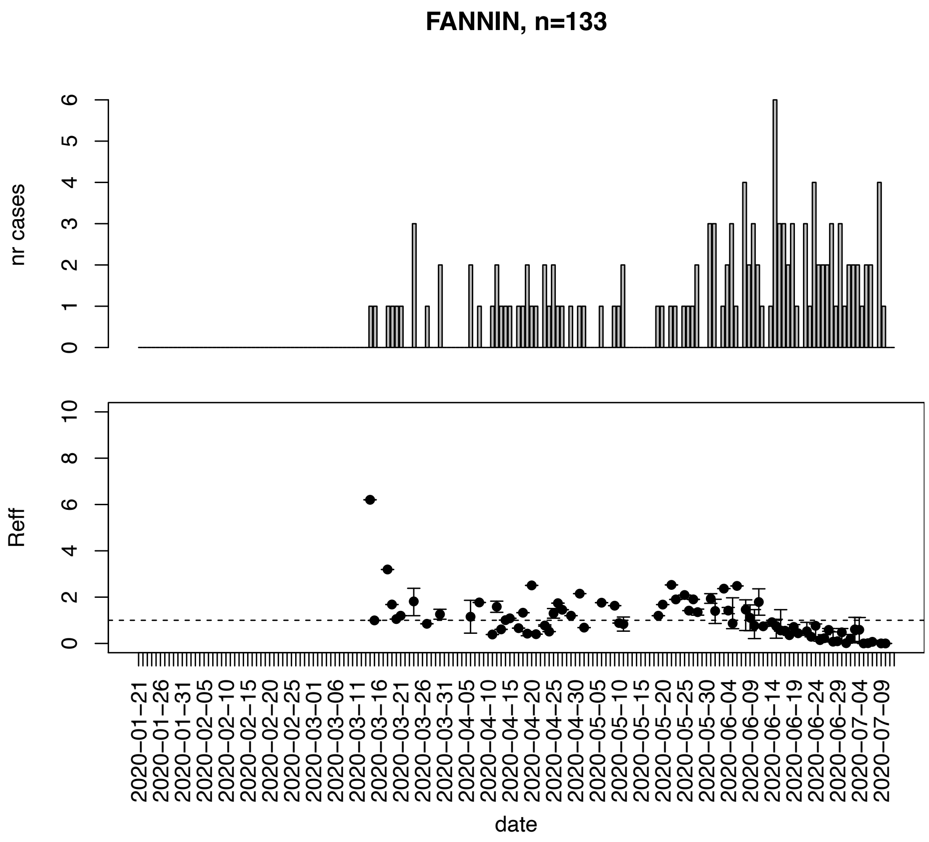


Figure. S55: Epidemic curves and reproduction number estimates until July 13th in Fannin county.

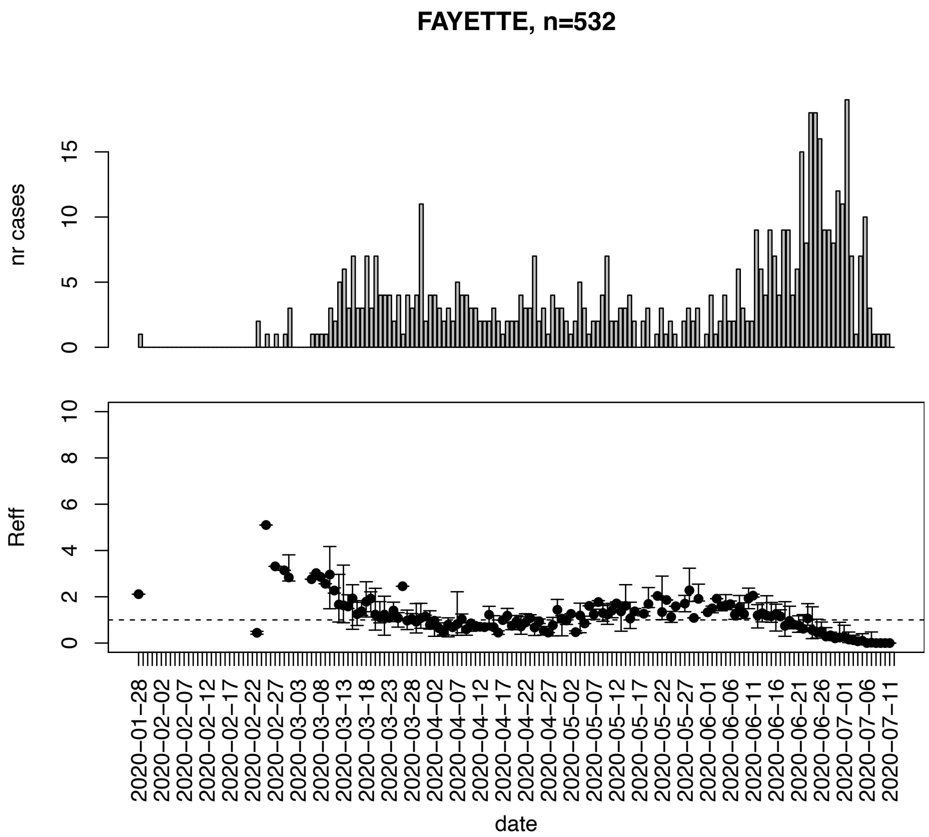


Figure. S56: Epidemic curves and reproduction number estimates until July 13th in Fayette county.

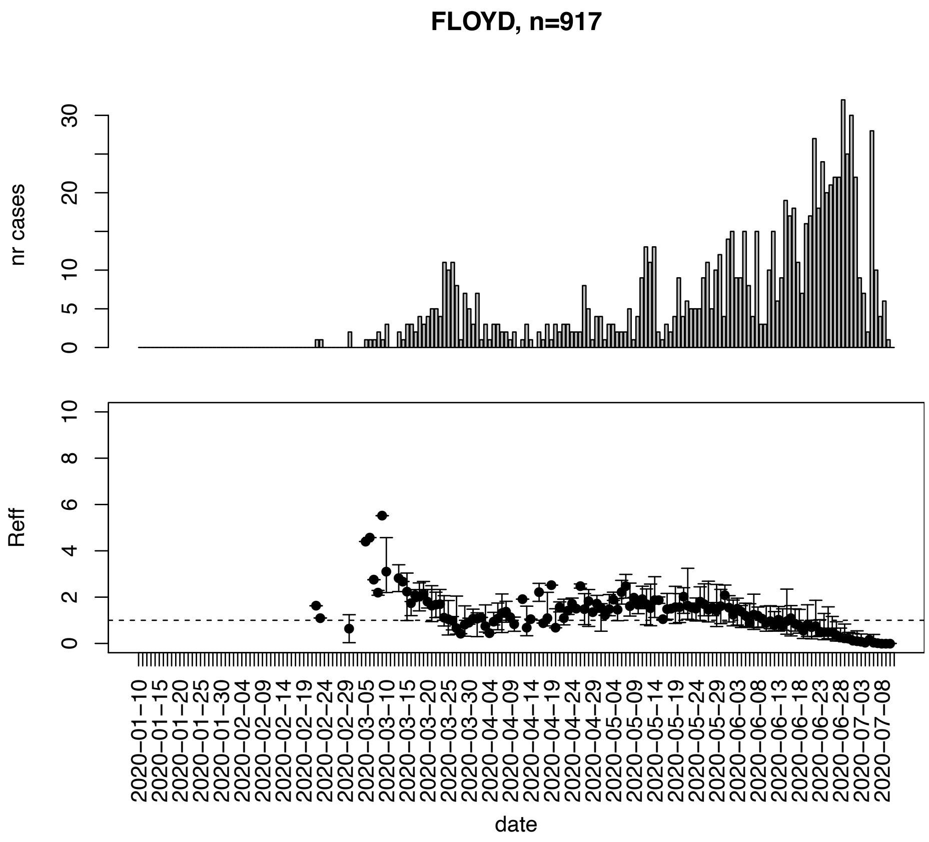


Figure. S57: Epidemic curves and reproduction number estimates until July 13th in Floyd county.

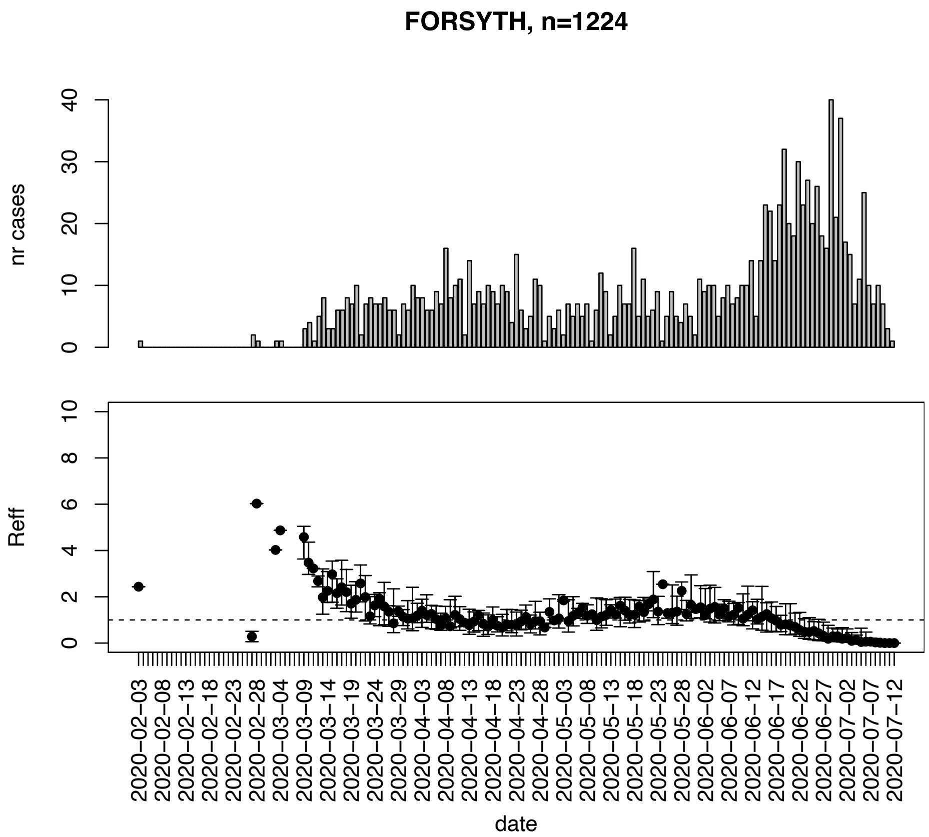


Figure. S58: Epidemic curves and reproduction number estimates until July 13th in Forsyth county.

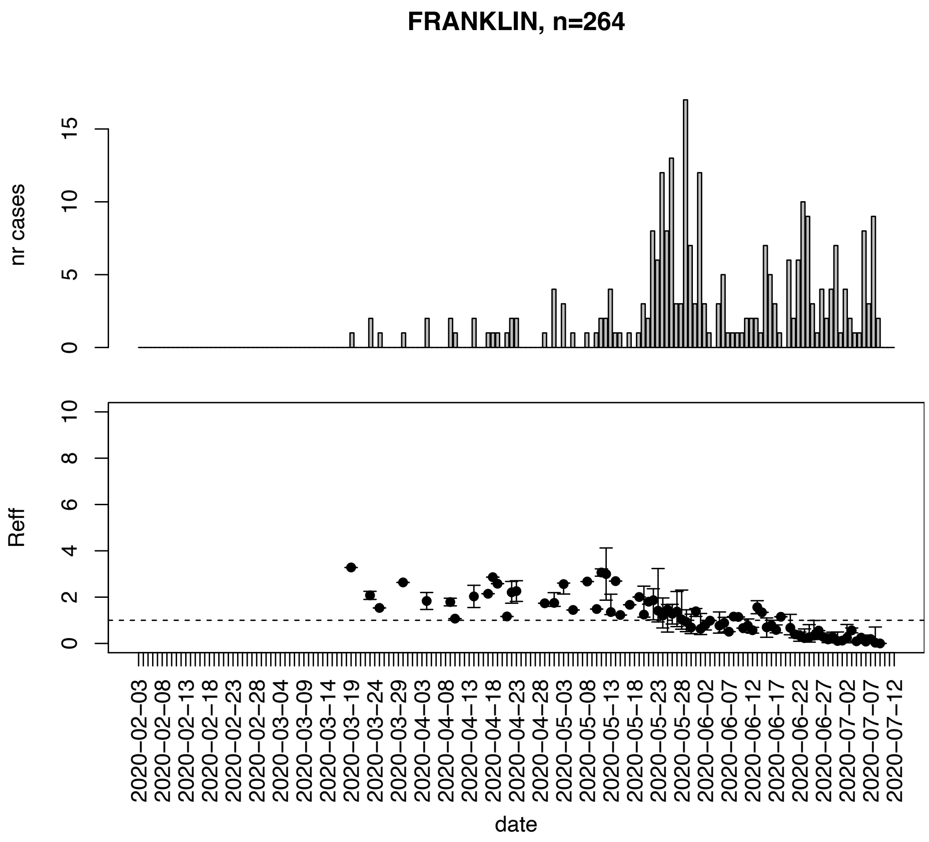


Figure. S59: Epidemic curves and reproduction number estimates until July 13th in Franklin county.

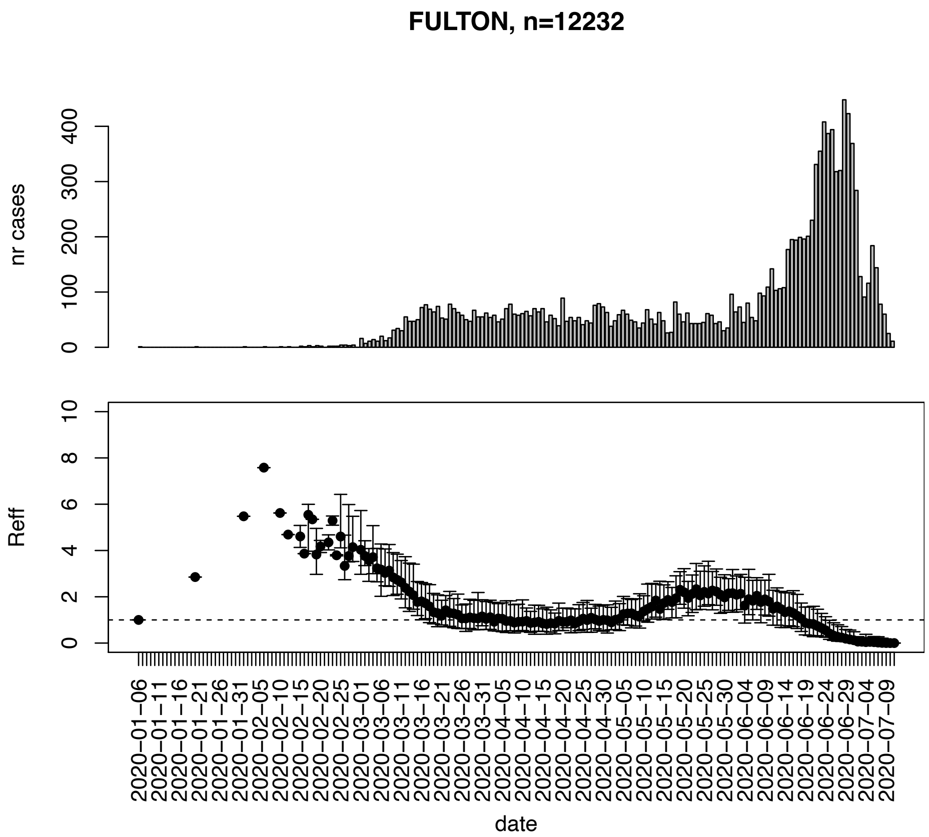


Figure. S60: Epidemic curves and reproduction number estimates until July 13th in Fulton county.

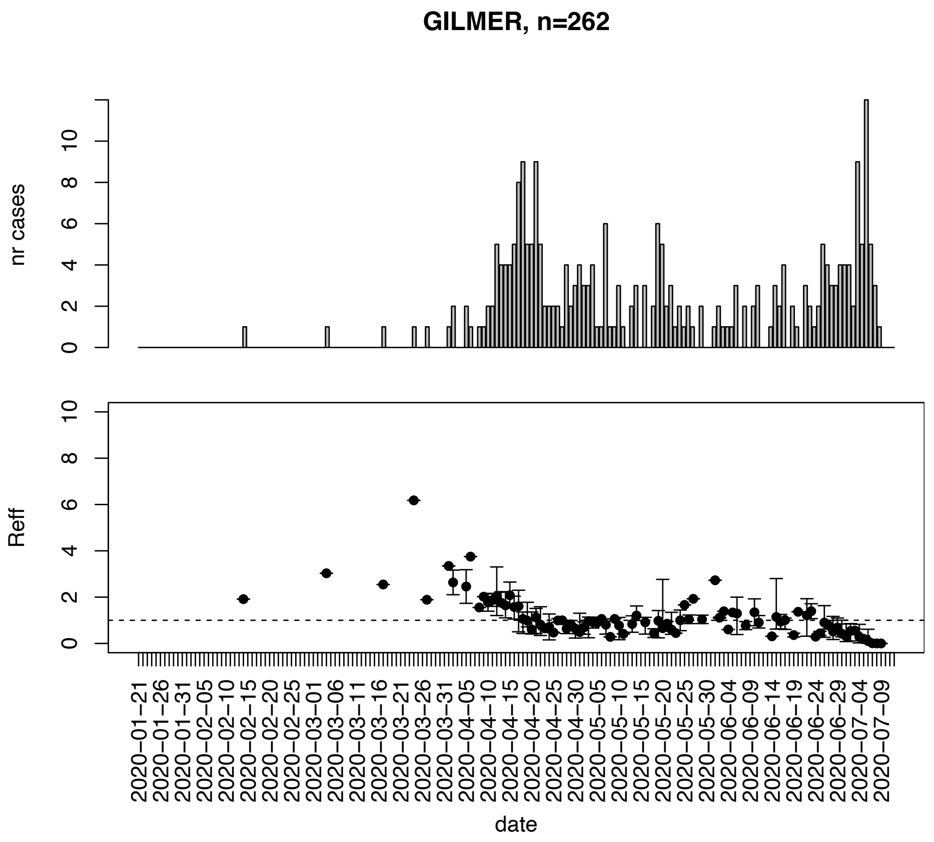


Figure. S61: Epidemic curves and reproduction number estimates until July 13th in Gilmer county.

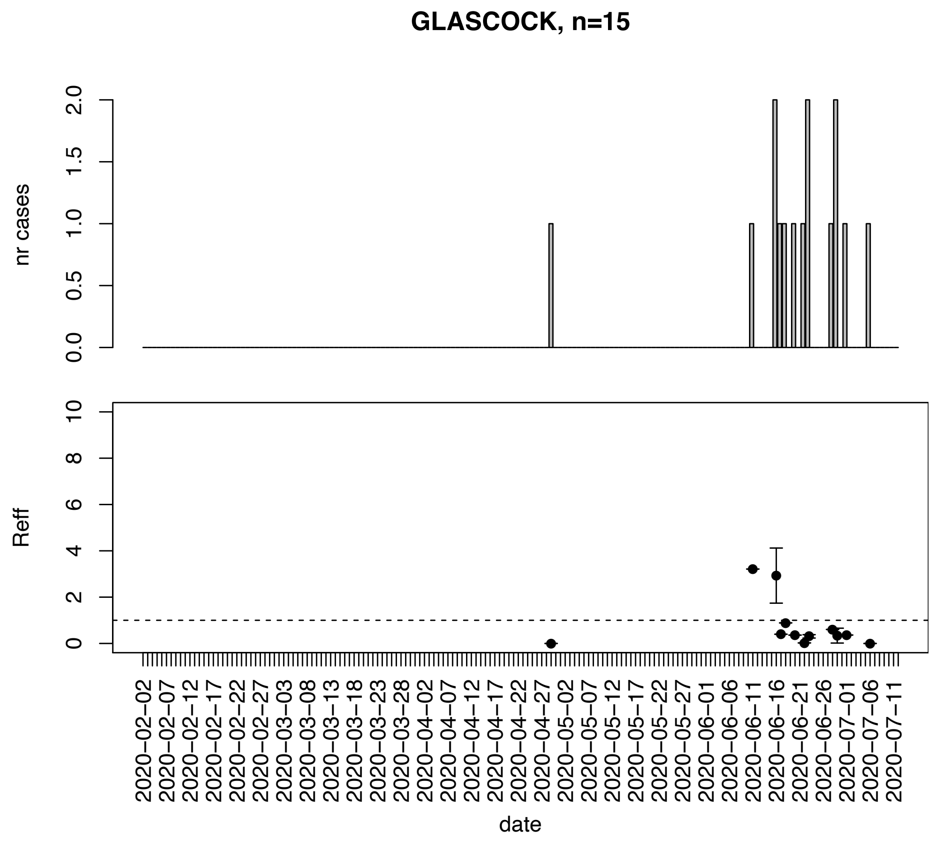


Figure. S62: Epidemic curves and reproduction number estimates until July 13th in Glascock county.

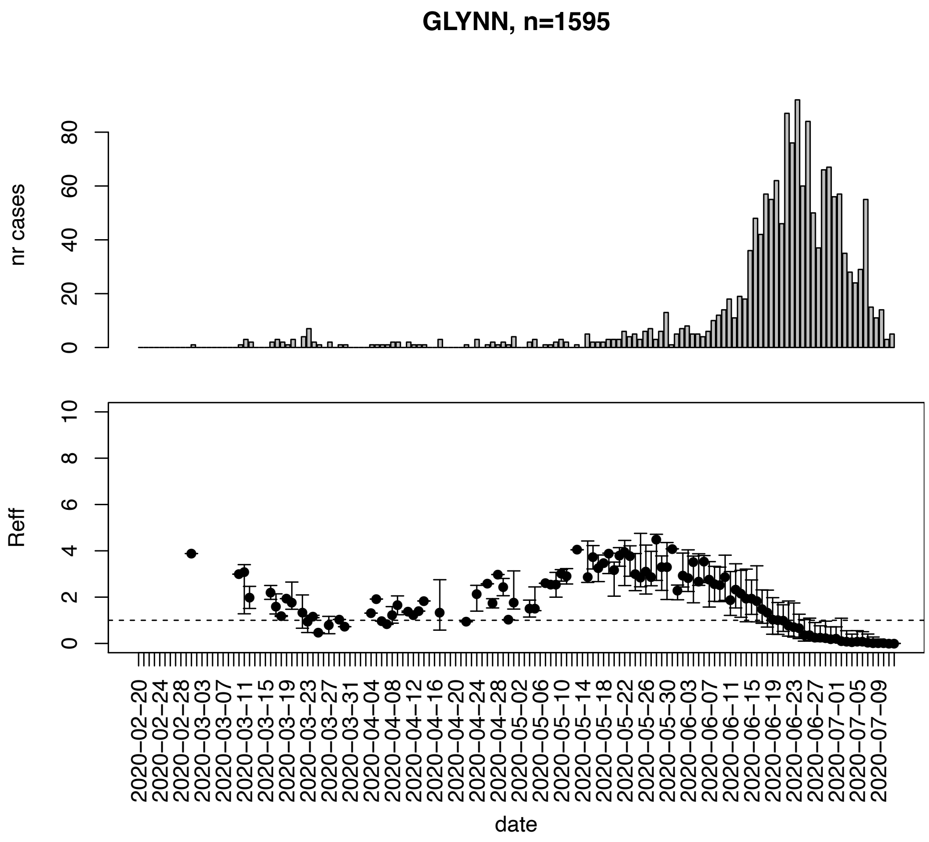


Figure. S63: Epidemic curves and reproduction number estimates until July 13th in Glynn county.

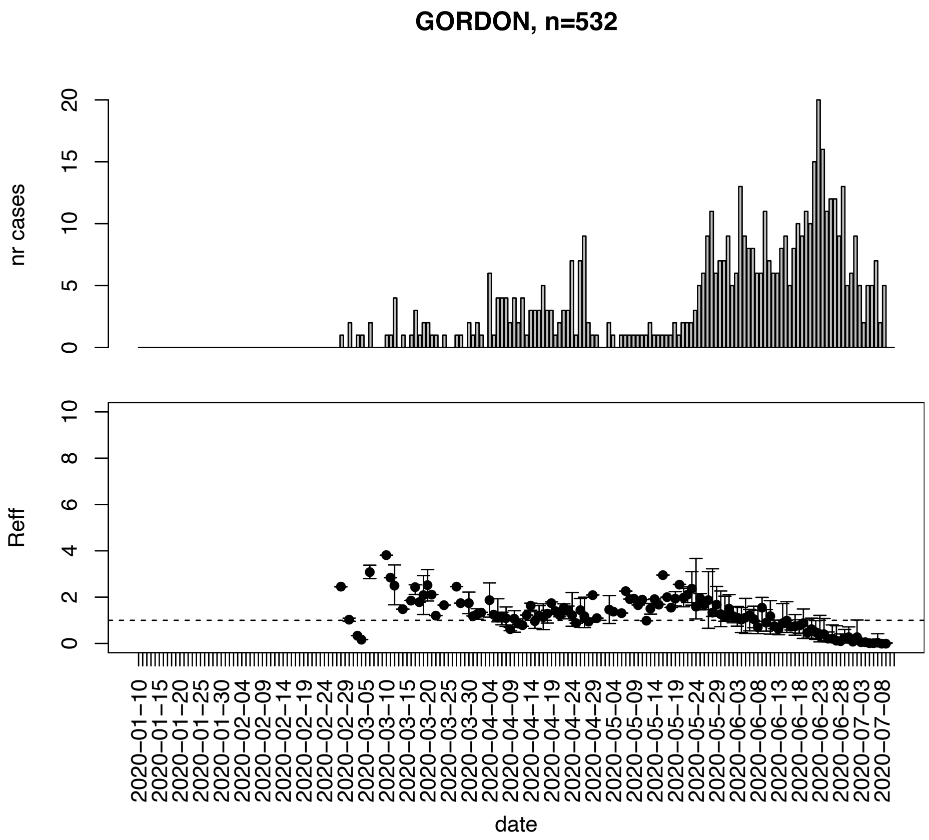


Figure. S64: Epidemic curves and reproduction number estimates until July 13th in Gordon county.

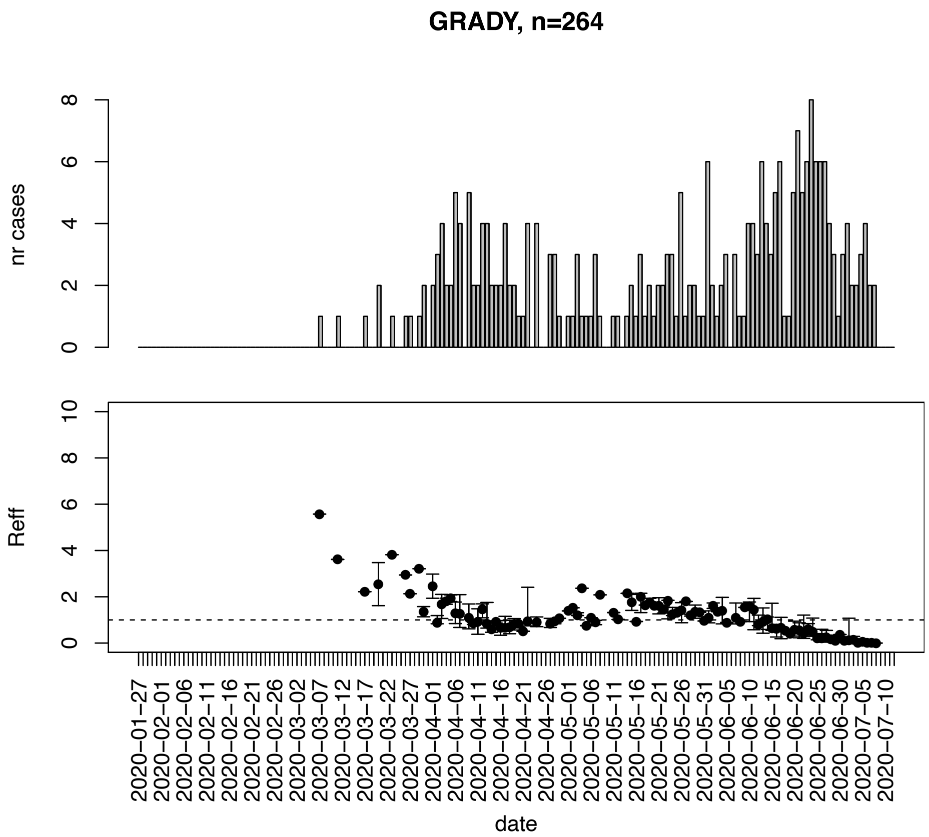


Figure. S65: Epidemic curves and reproduction number estimates until July 13th in Grady county.

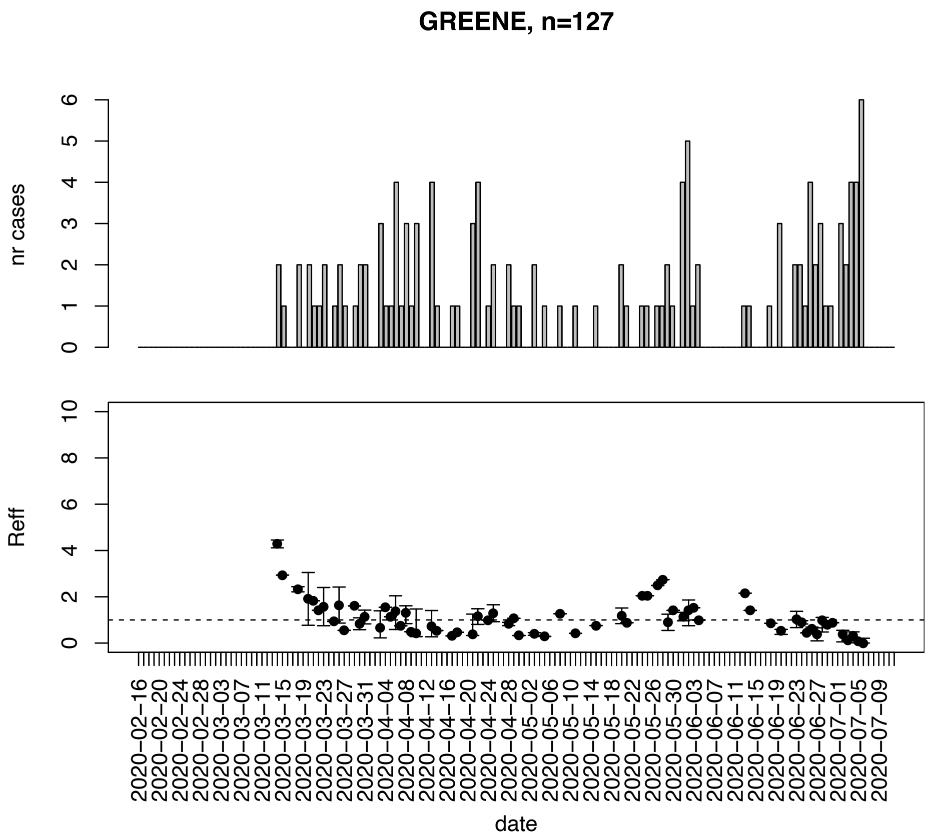


Figure. S66: Epidemic curves and reproduction number estimates until July 13th in Greene county.

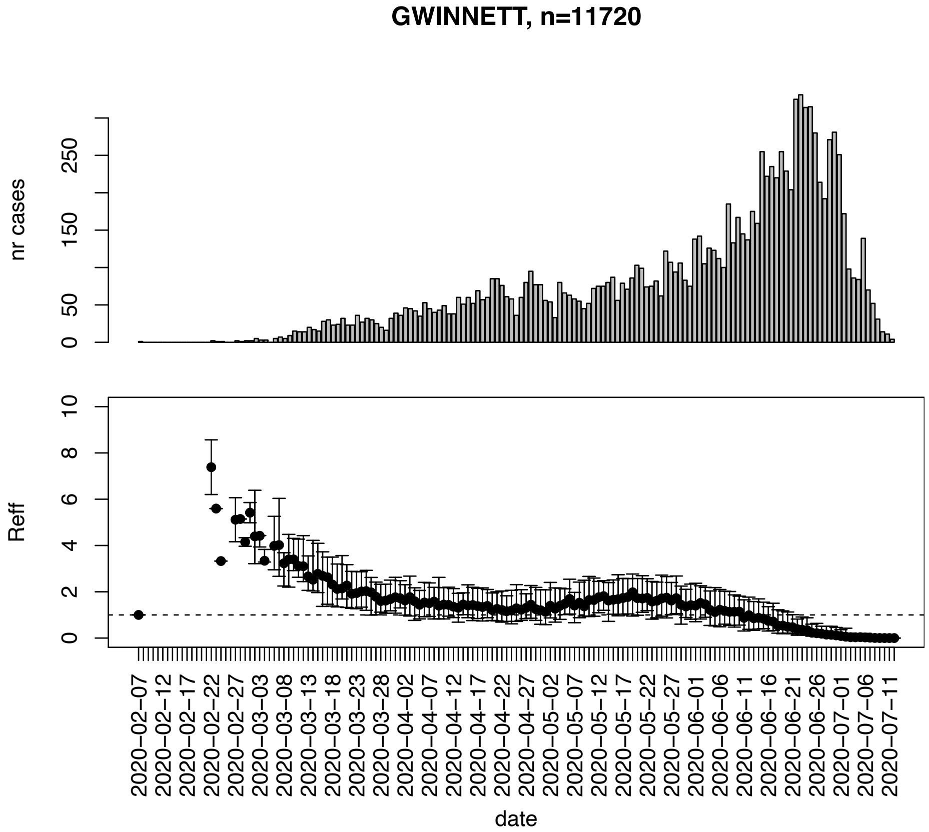


Figure. S67: Epidemic curves and reproduction number estimates until July 13th in Gwinnett county.

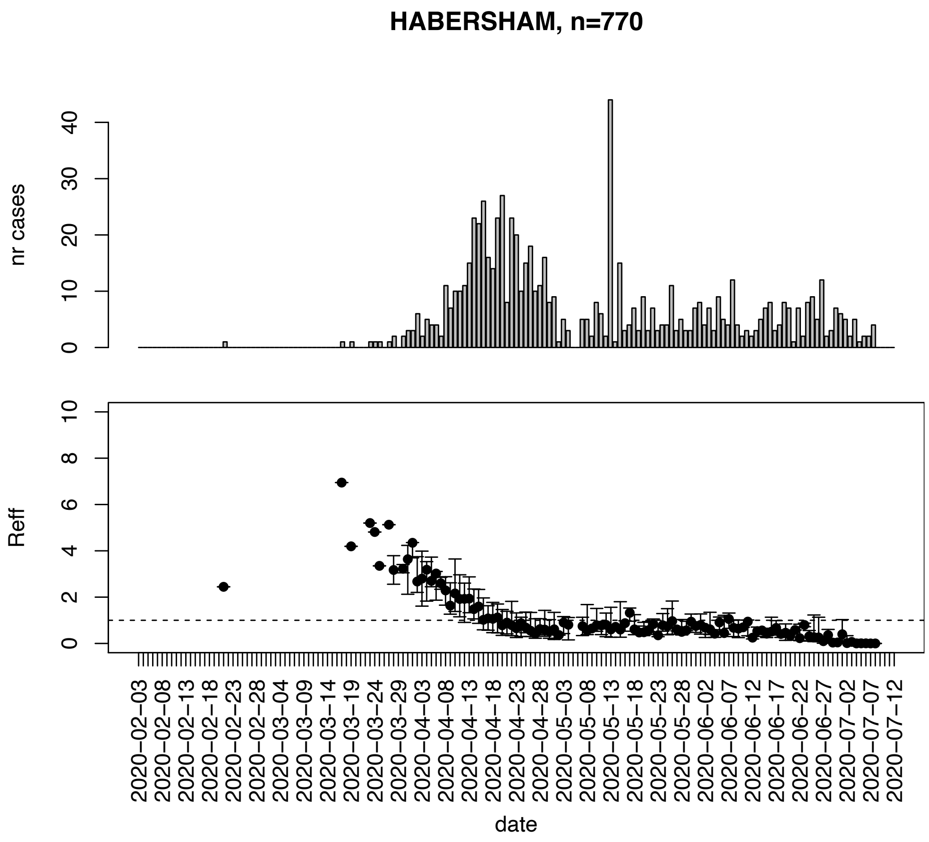


Figure. S68: Epidemic curves and reproduction number estimates until July 13th in Habersham county.

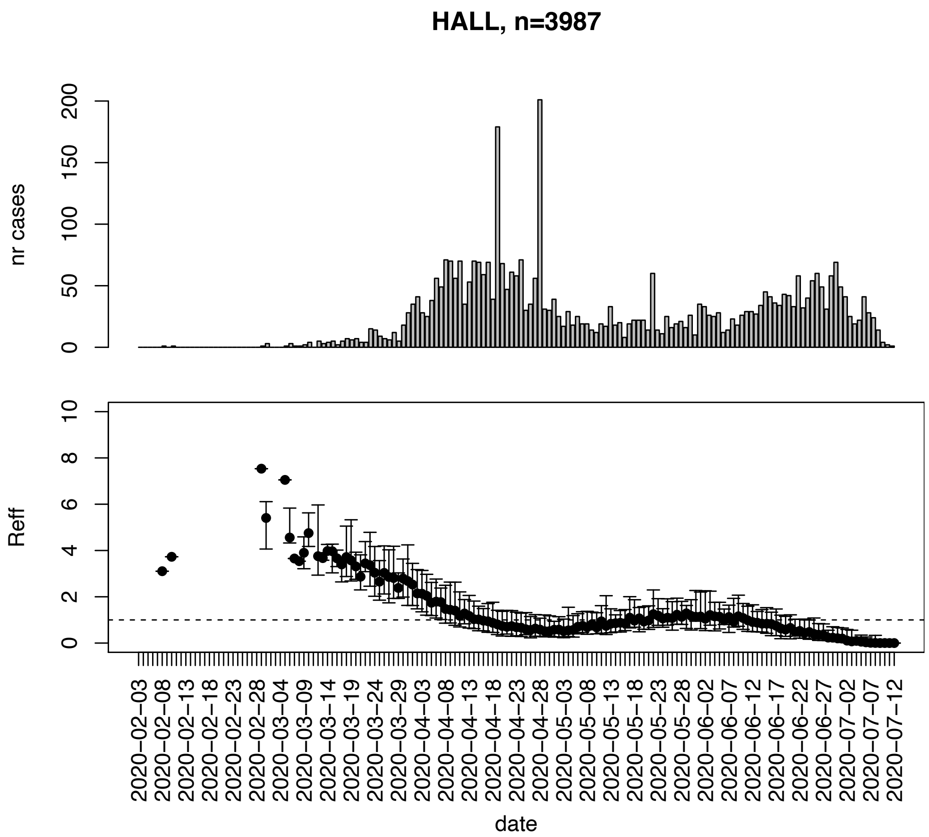


Figure. S69: Epidemic curves and reproduction number estimates until July 13th in Hall county.

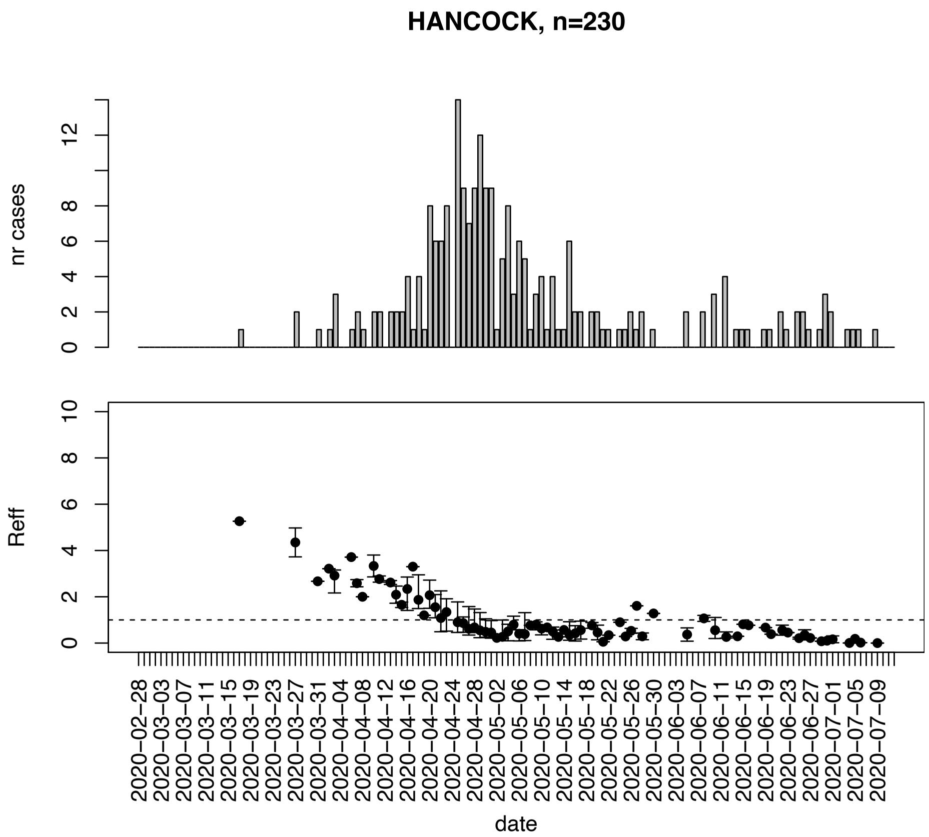


Figure. S70: Epidemic curves and reproduction number estimates until July 13th in Hancock county.

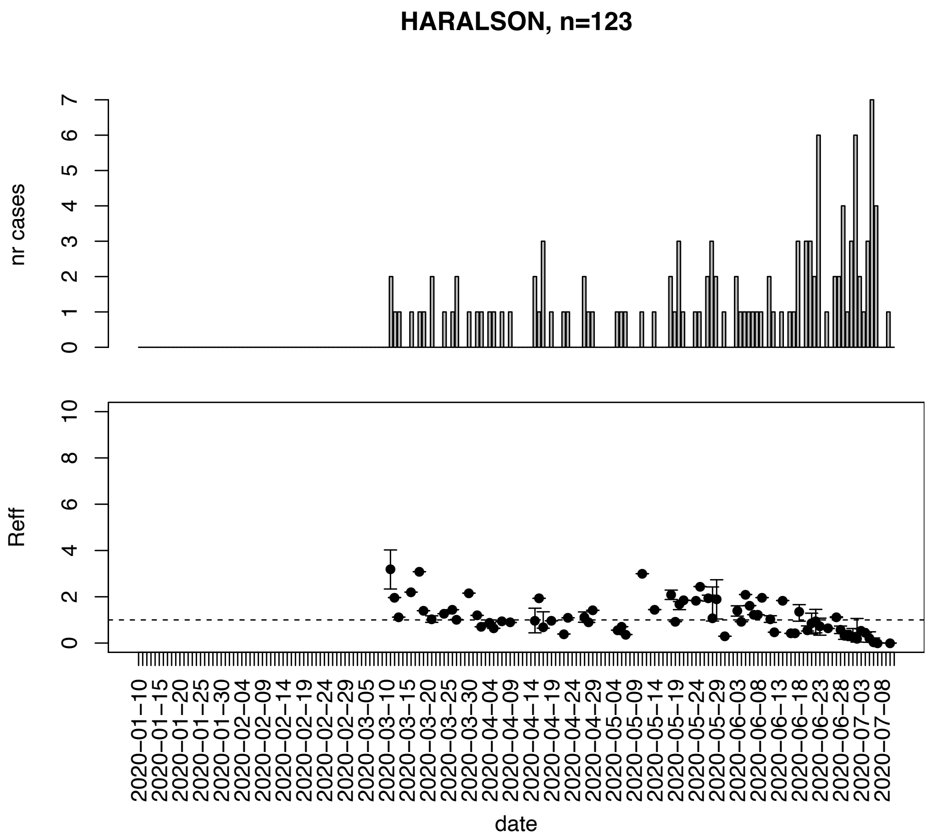


Figure. S71: Epidemic curves and reproduction number estimates until July 13th in Haralson county.

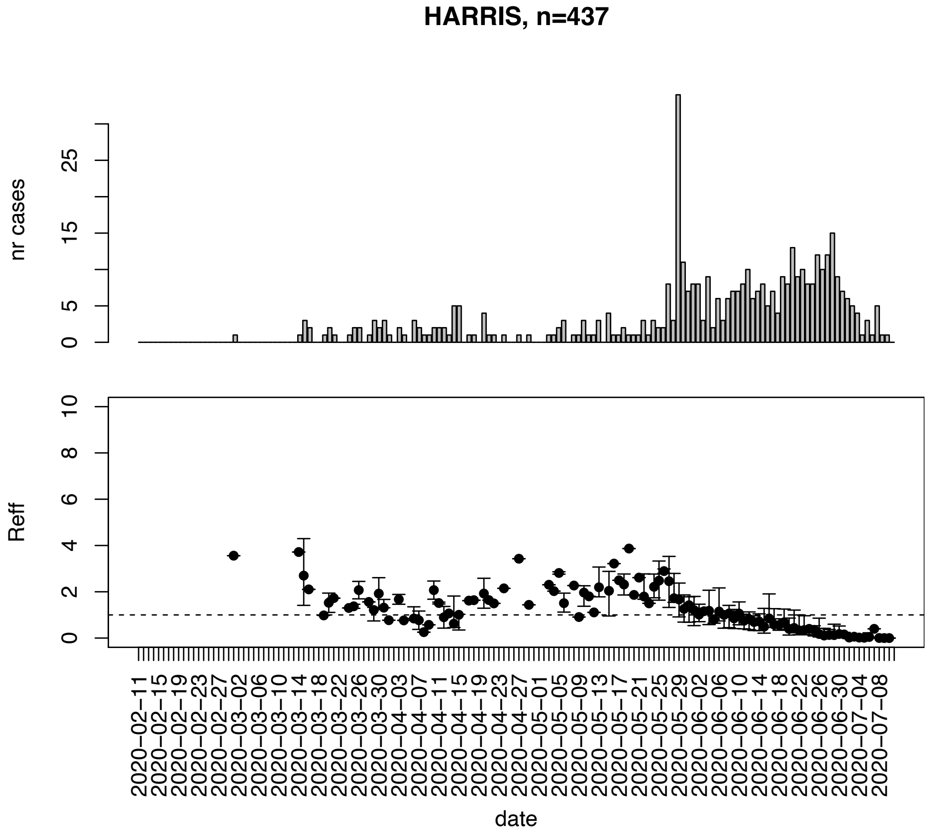


Figure. S72: Epidemic curves and reproduction number estimates until July 13th in Harris county.

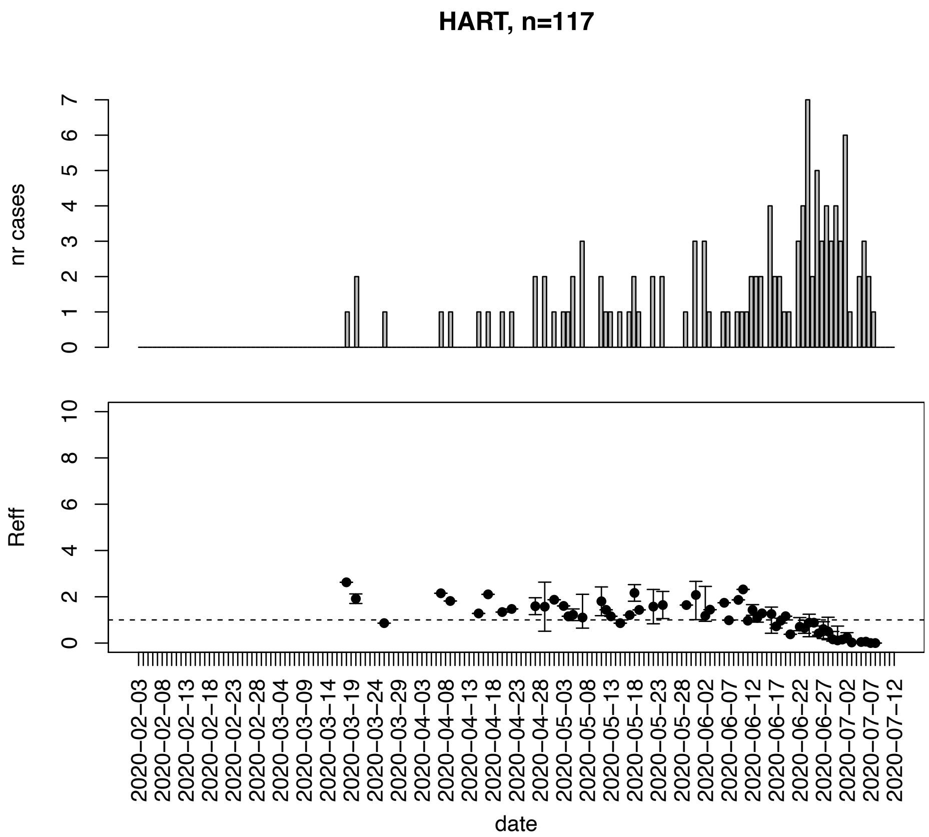


Figure. S73: Epidemic curves and reproduction number estimates until July 13th in Hart county.

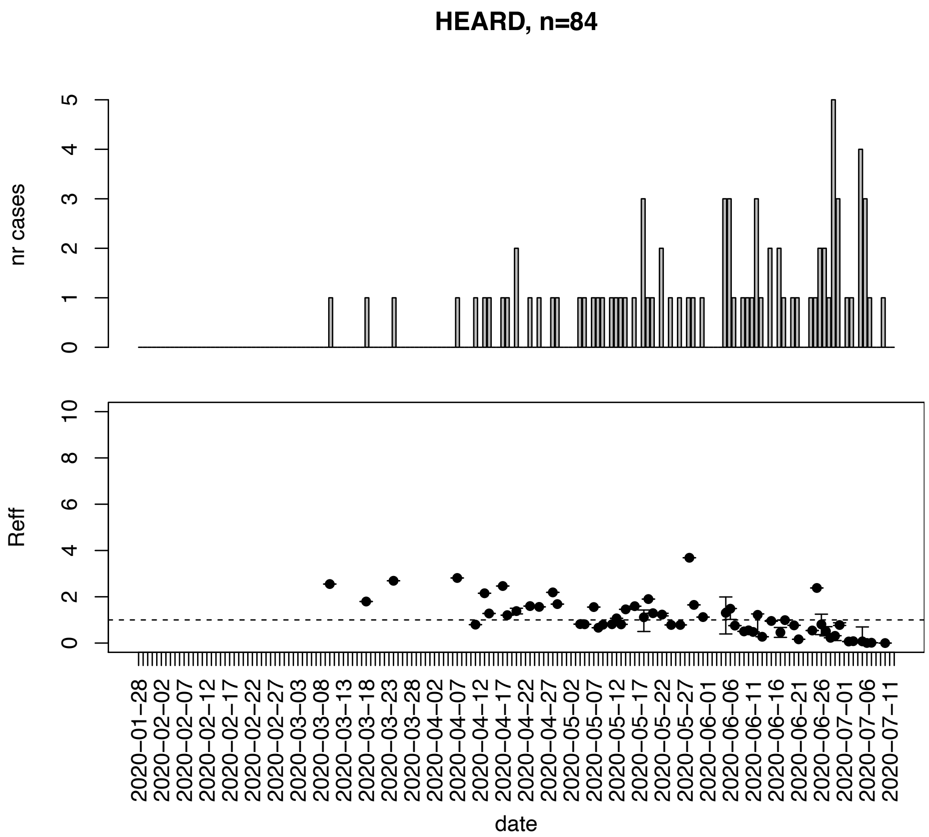


Figure. S74: Epidemic curves and reproduction number estimates until July 13th in Heard county.

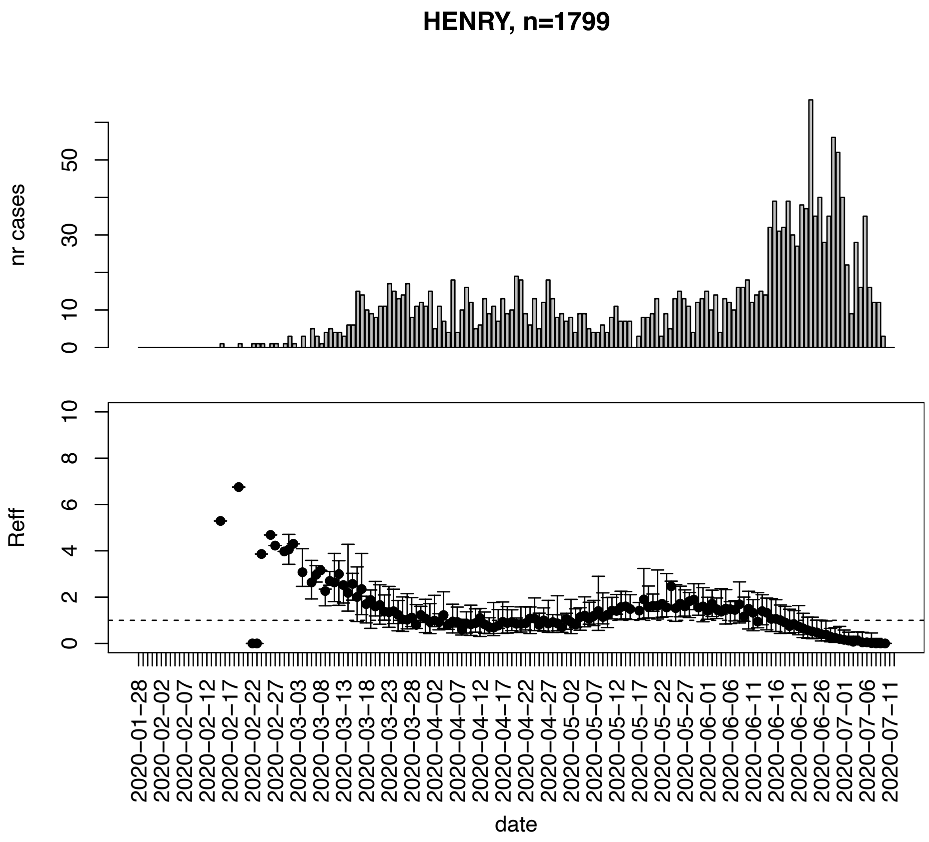


Figure. S75: Epidemic curves and reproduction number estimates until July 13th in Henry county.

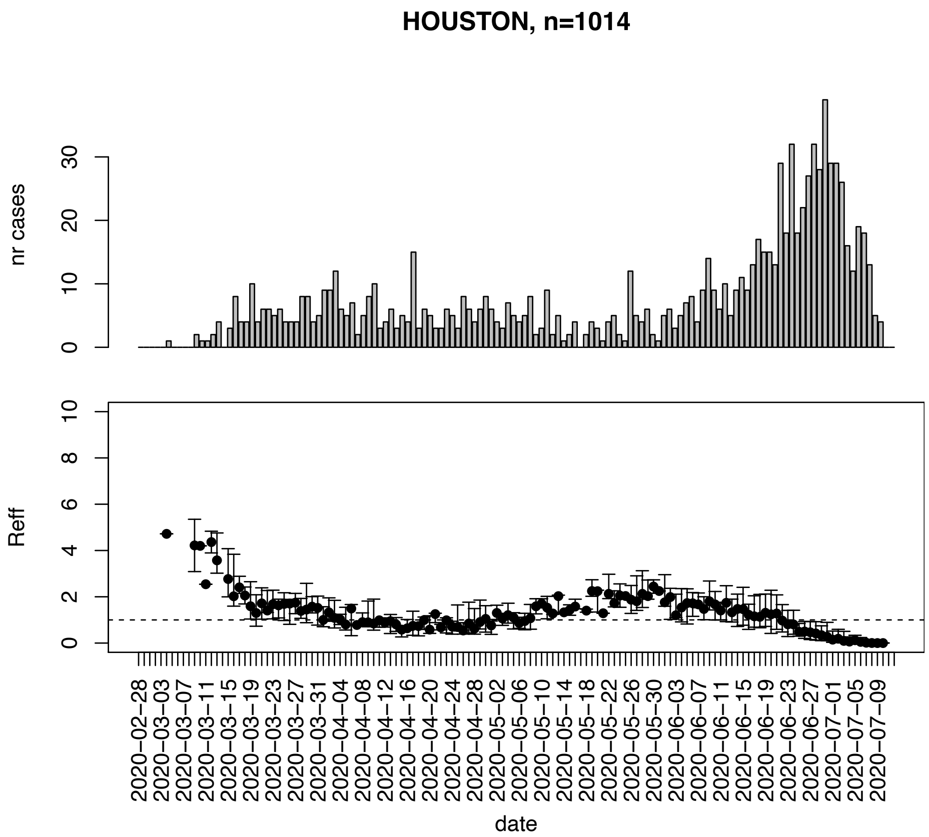


Figure. S76: Epidemic curves and reproduction number estimates until July 13th in Houston county.

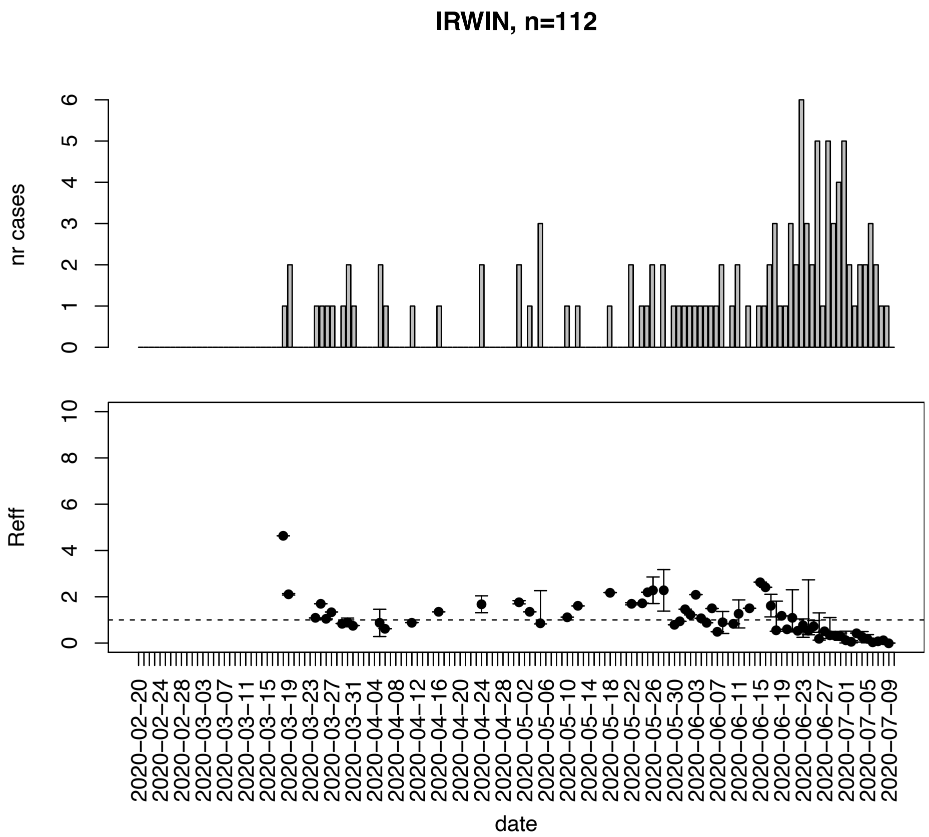


Figure. S77: Epidemic curves and reproduction number estimates until July 13th in Irwin county.

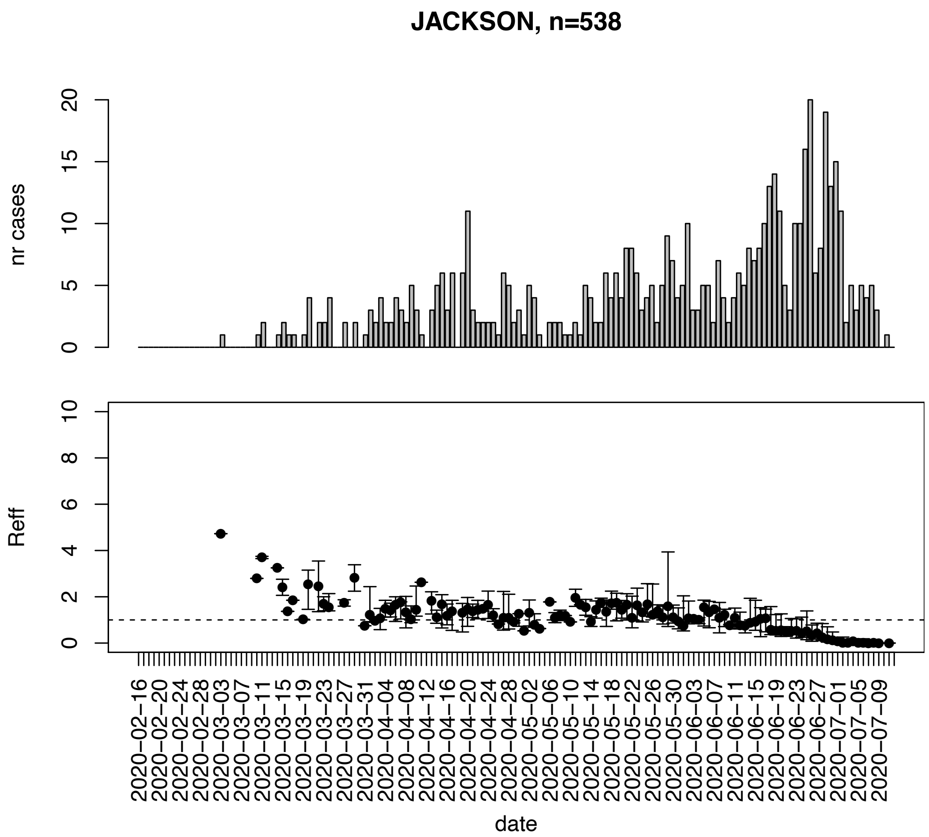


Figure. S78: Epidemic curves and reproduction number estimates until July 13th in Jackson county.

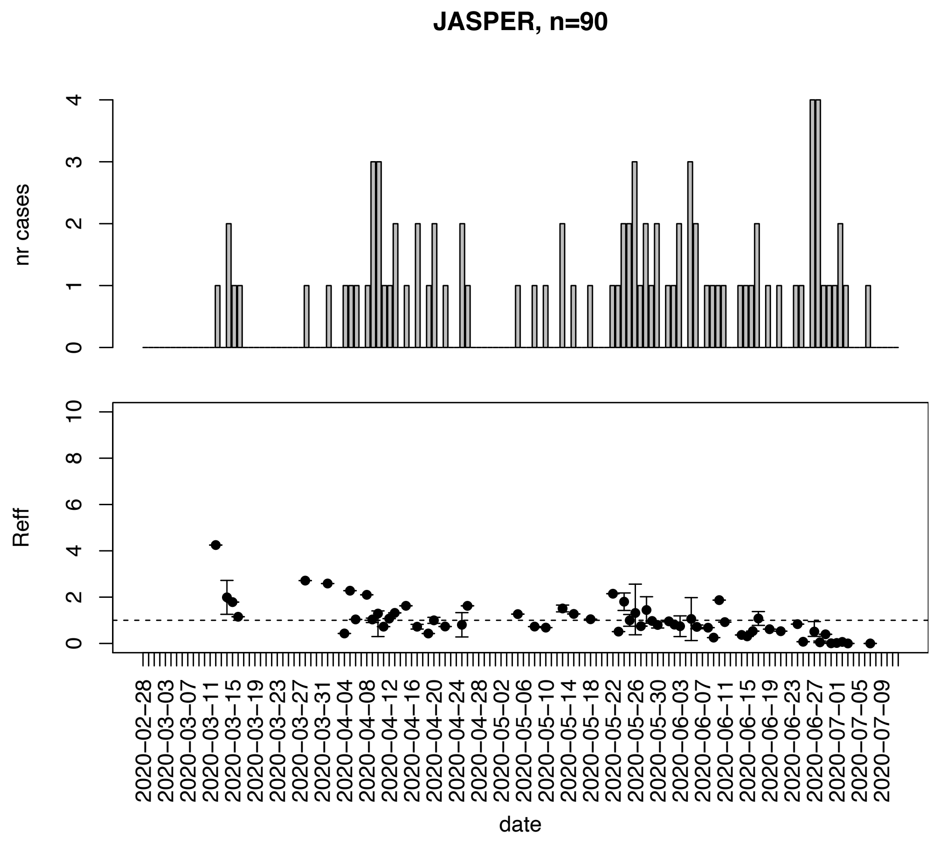


Figure. S79: Epidemic curves and reproduction number estimates until July 13th in Jasper county.

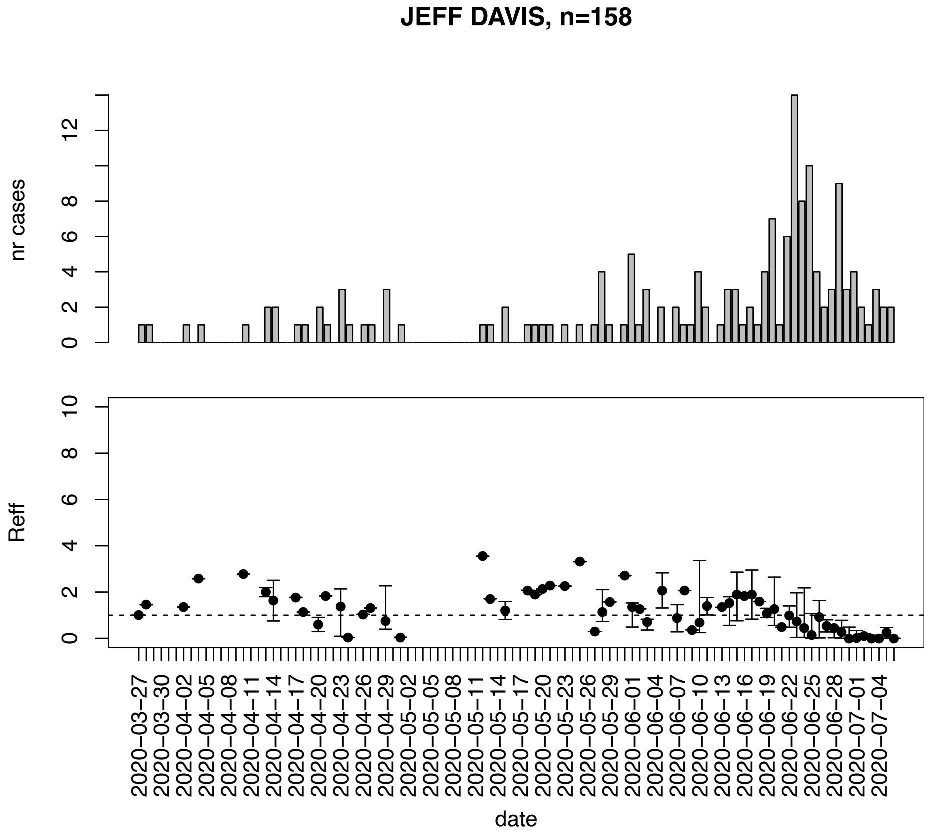


Figure. S80: Epidemic curves and reproduction number estimates until July 13th in Jeff Davis county.

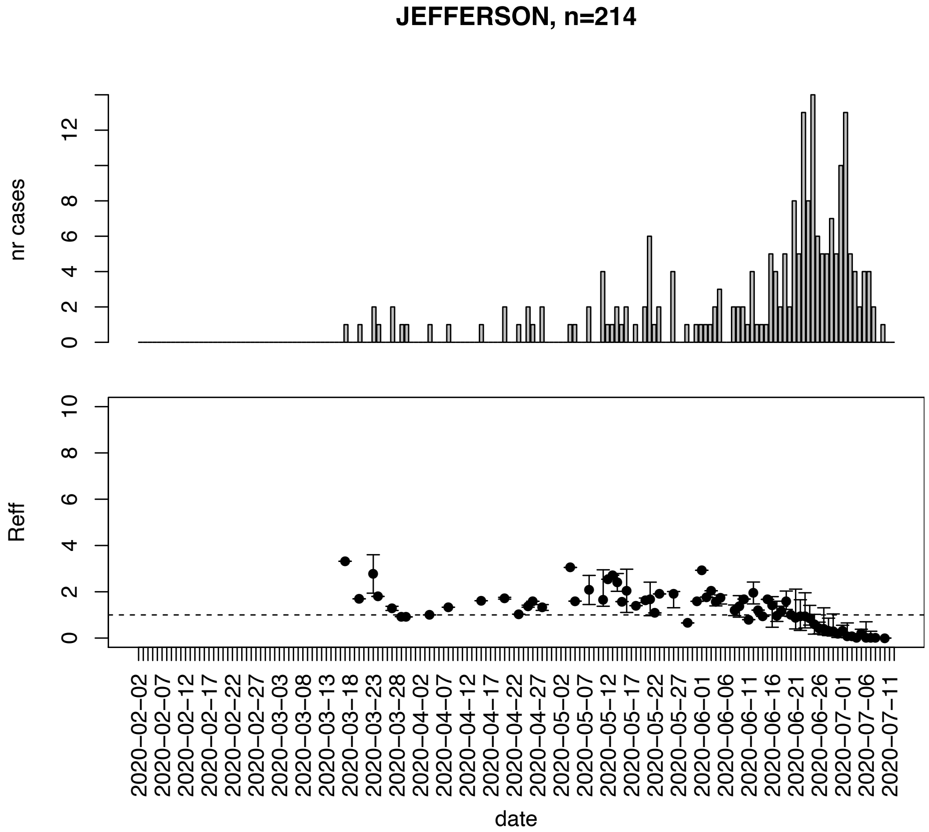


Figure. S81: Epidemic curves and reproduction number estimates until July 13th in Jefferson county.

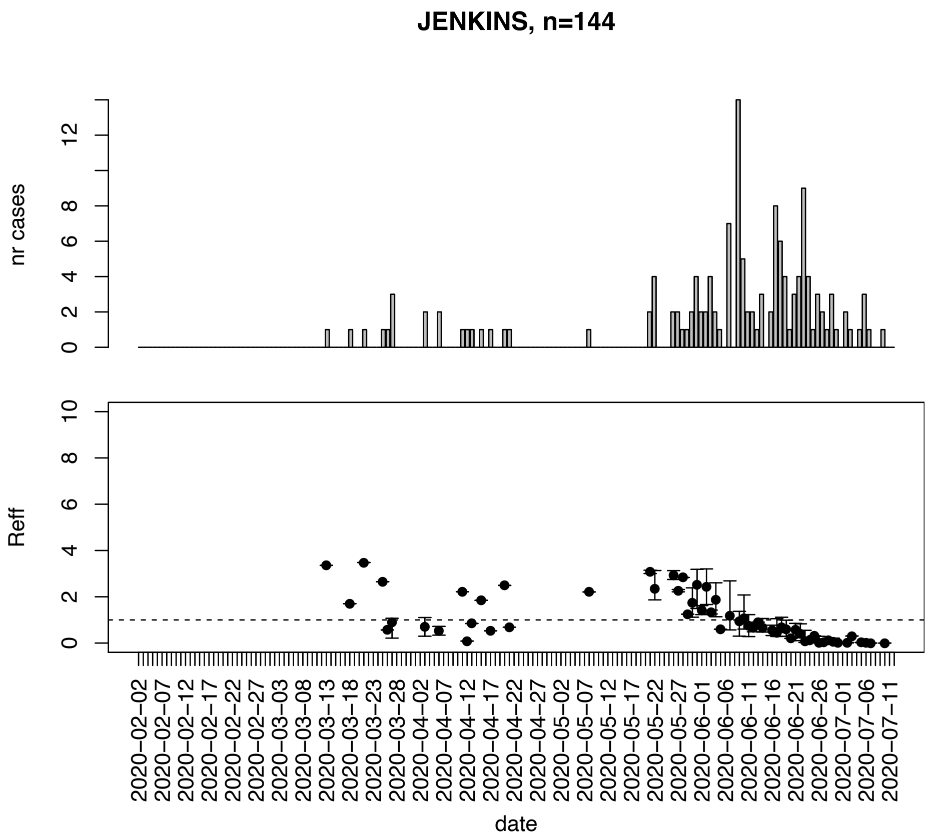


Figure. S82: Epidemic curves and reproduction number estimates until July 13th in Jenkins county.

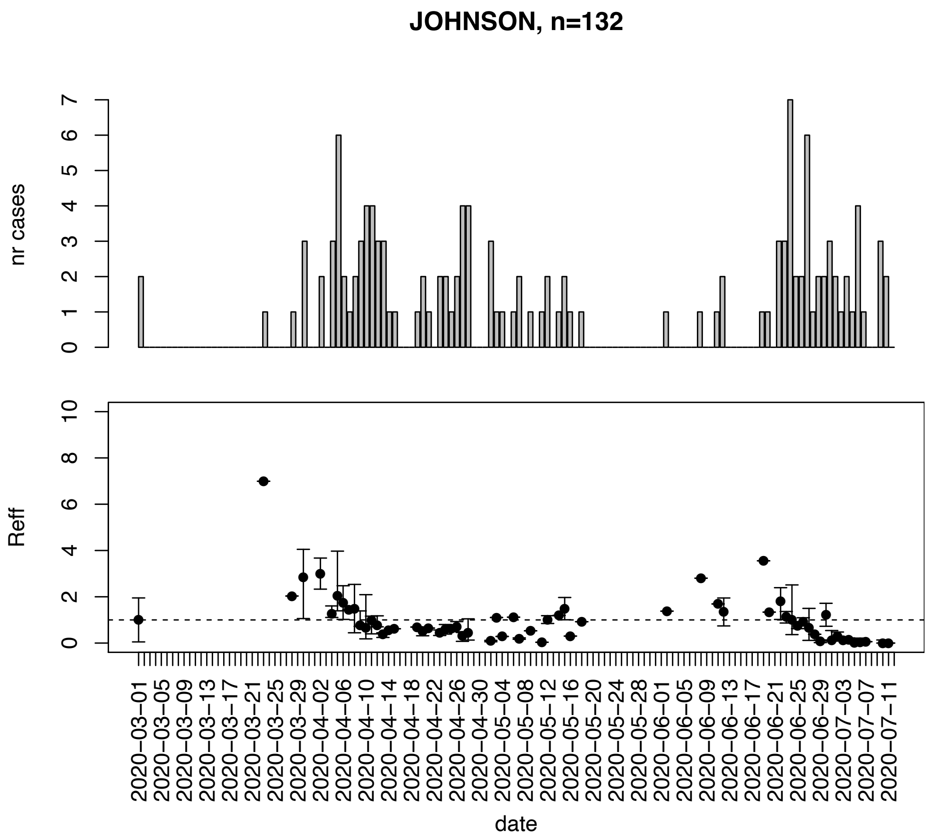


Figure. S83: Epidemic curves and reproduction number estimates until July 13th in Johnson county.

JONES, n=134

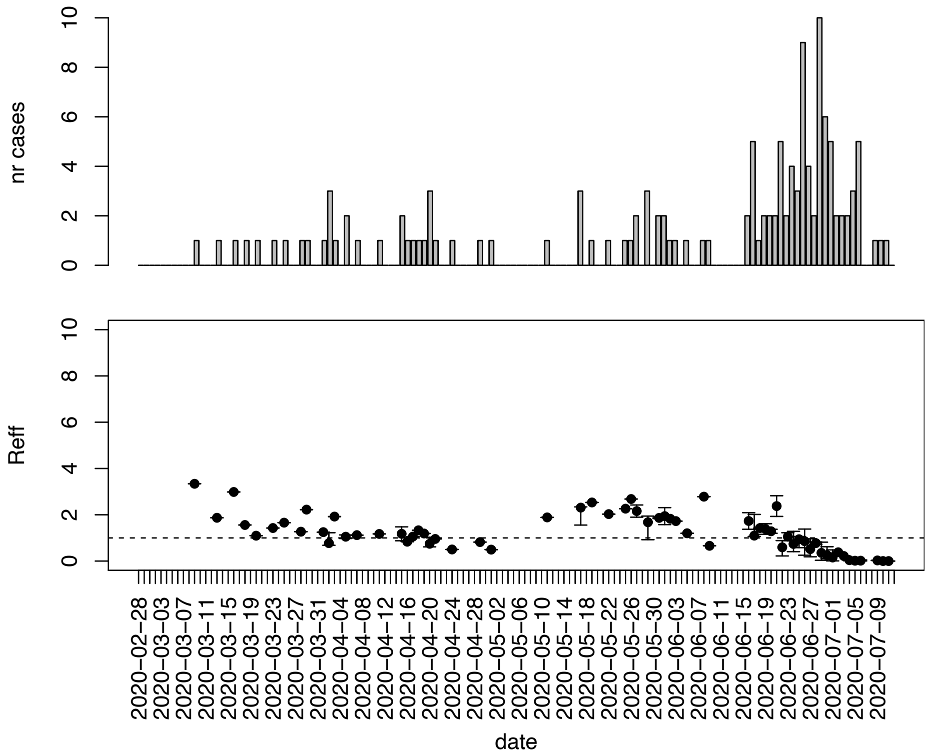


Figure. S84: Epidemic curves and reproduction number estimates until July 13th in Jones county.

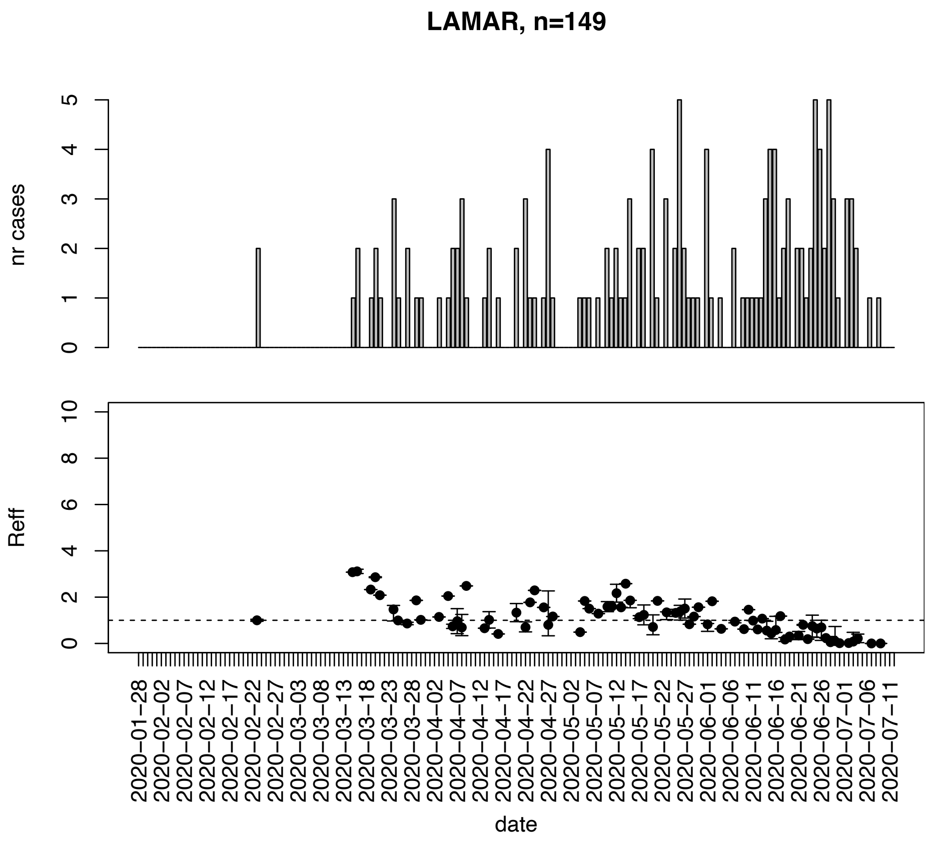


Figure. S85: Epidemic curves and reproduction number estimates until July 13th in Lamar county.

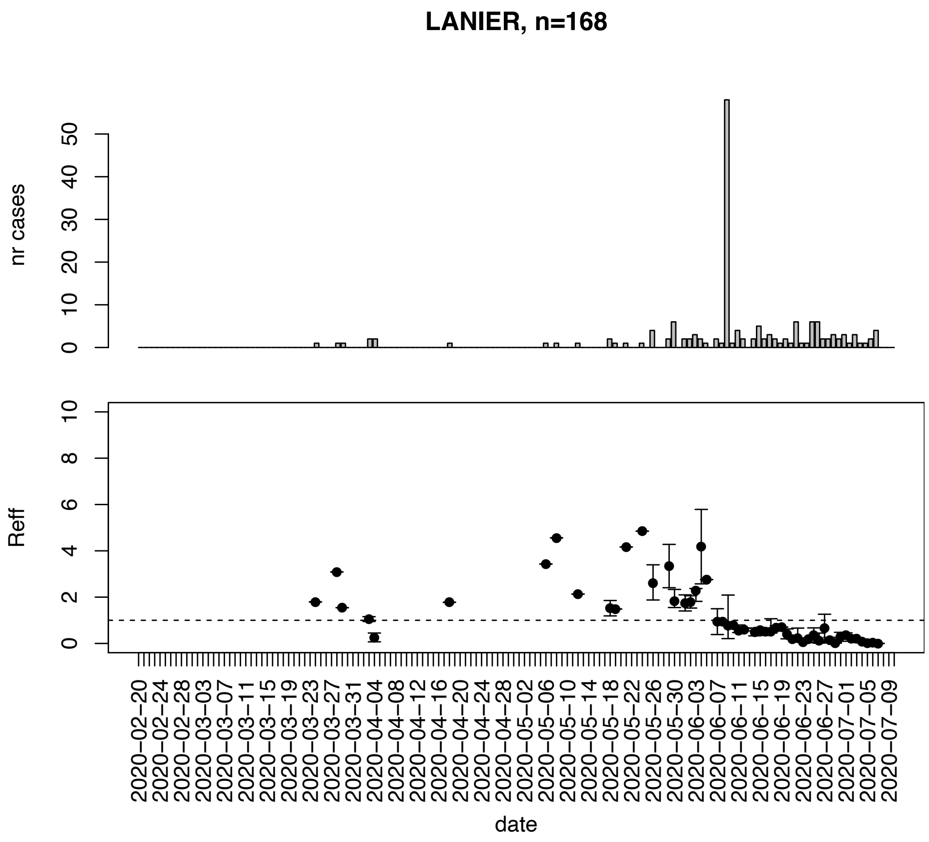


Figure. S86: Epidemic curves and reproduction number estimates until July 13th in Lanier county.

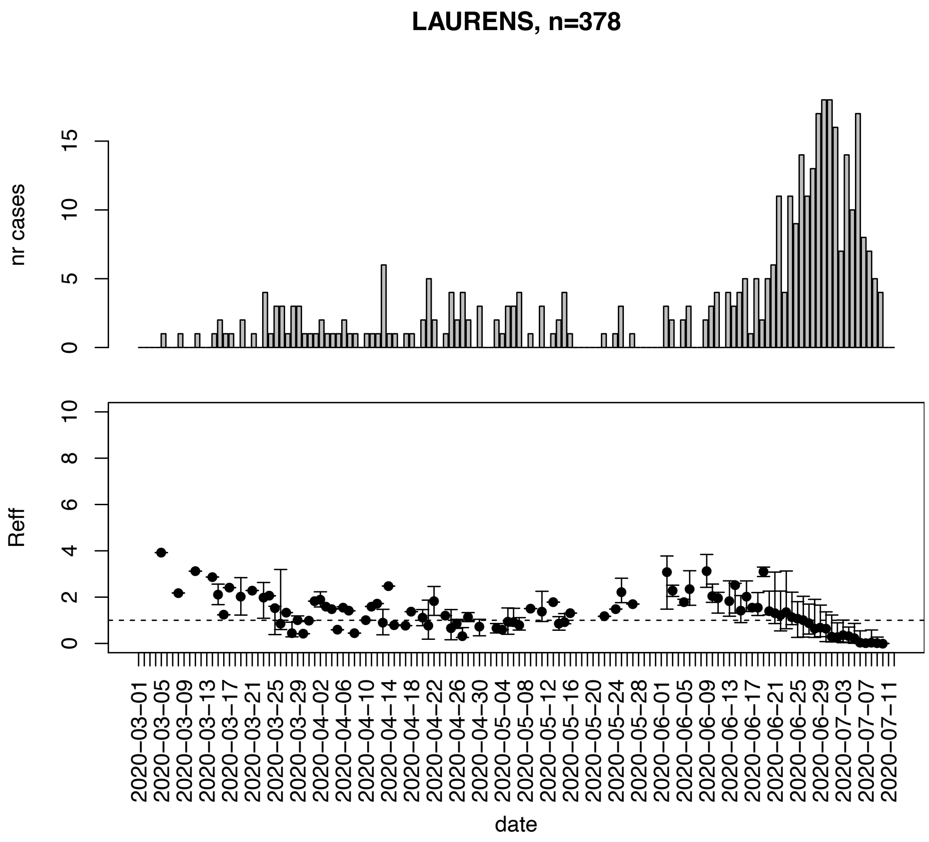


Figure. S87: Epidemic curves and reproduction number estimates until July 13th in Laurens county.

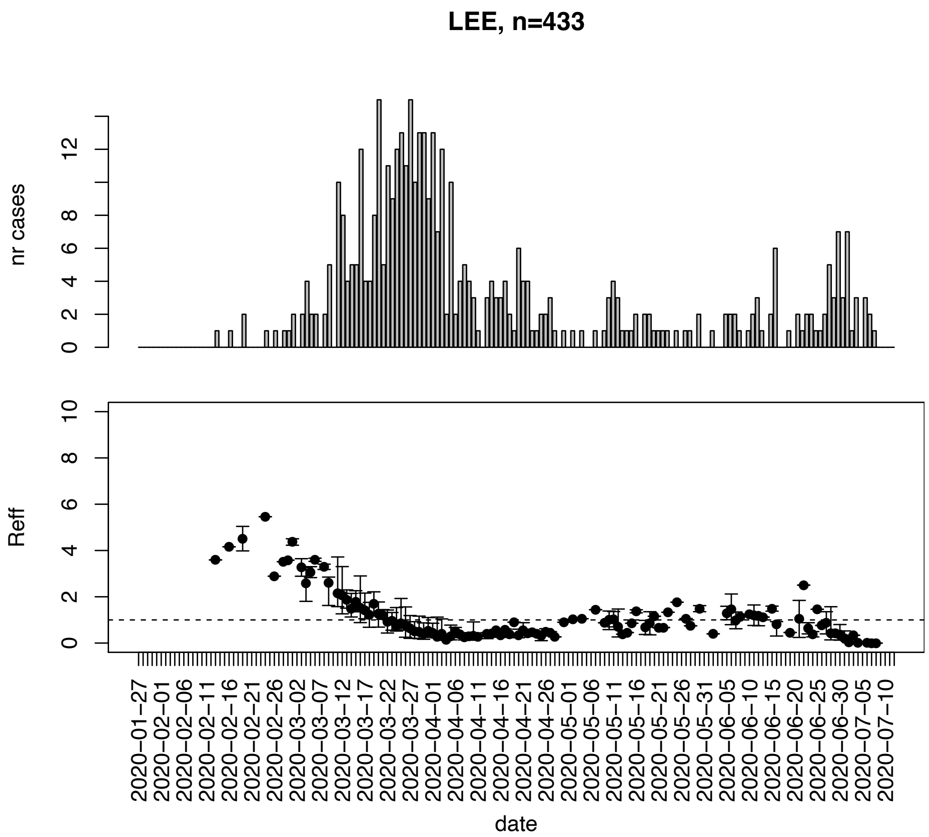


Figure. S88: Epidemic curves and reproduction number estimates until July 13th in Lee county.

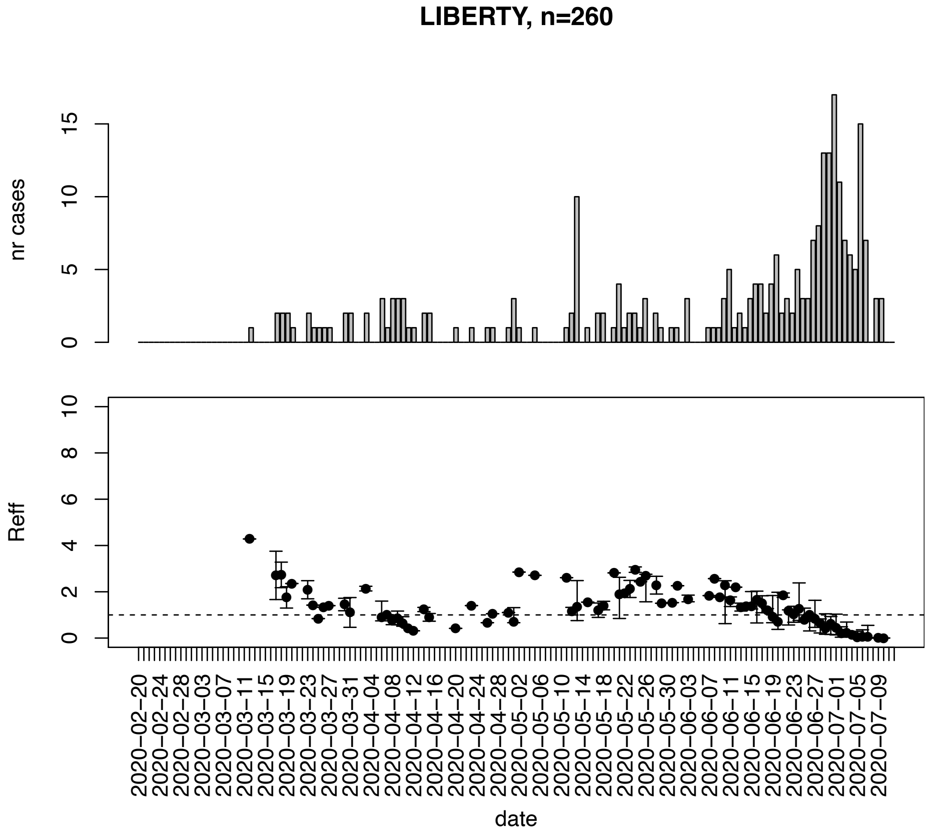


Figure. S89: Epidemic curves and reproduction number estimates until July 13th in Liberty county.

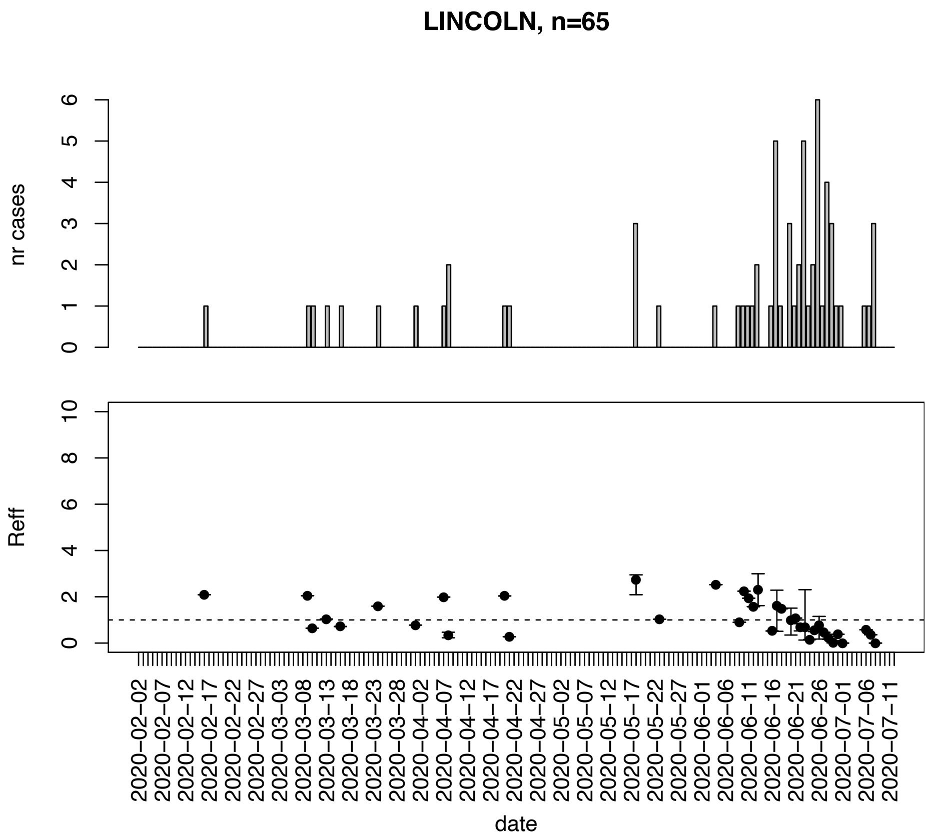


Figure. S90: Epidemic curves and reproduction number estimates until July 13th in Lincoln county.

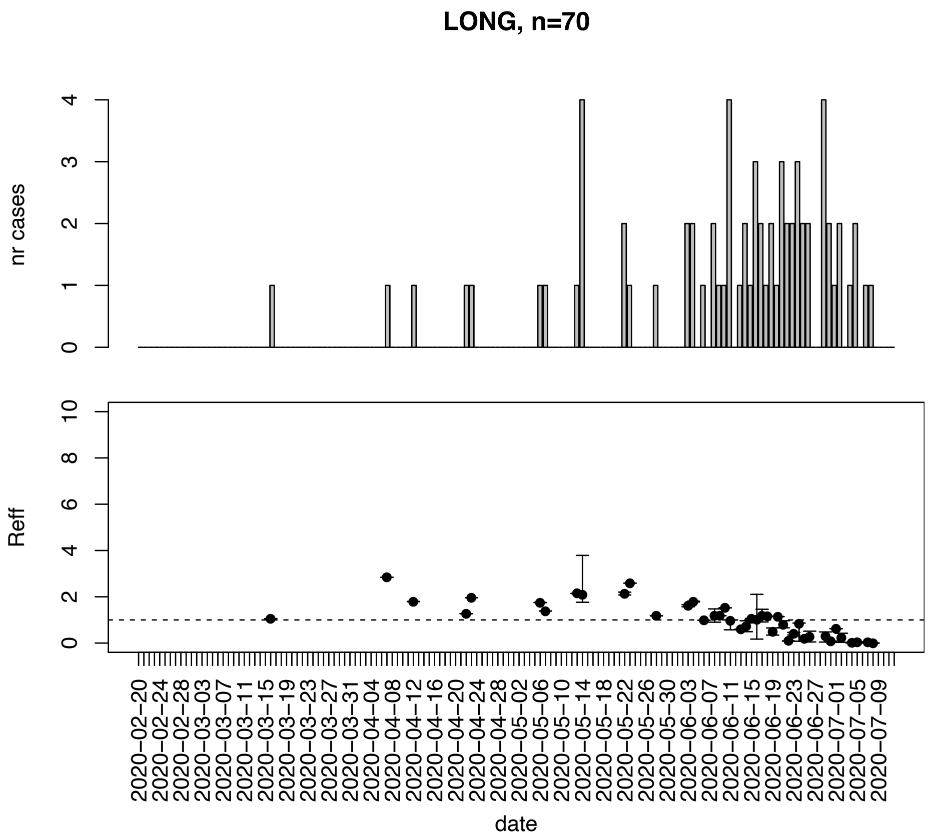


Figure. S91: Epidemic curves and reproduction number estimates until July 13th in Long county.

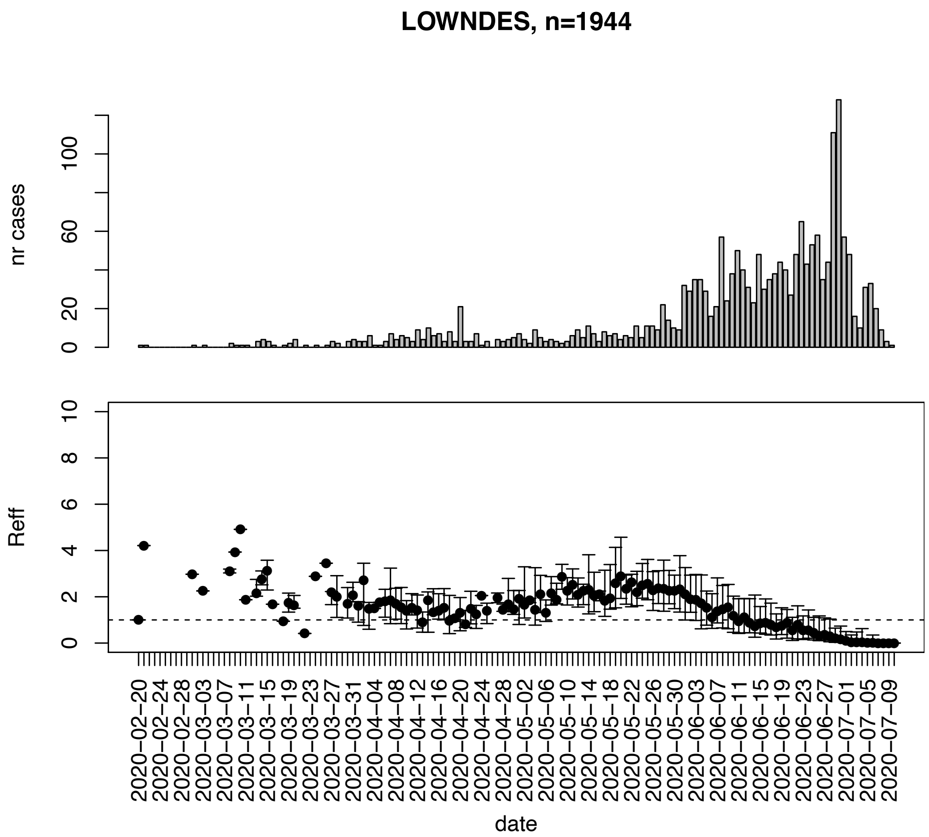


Figure. S92: Epidemic curves and reproduction number estimates until July 13th in Lowndes county.

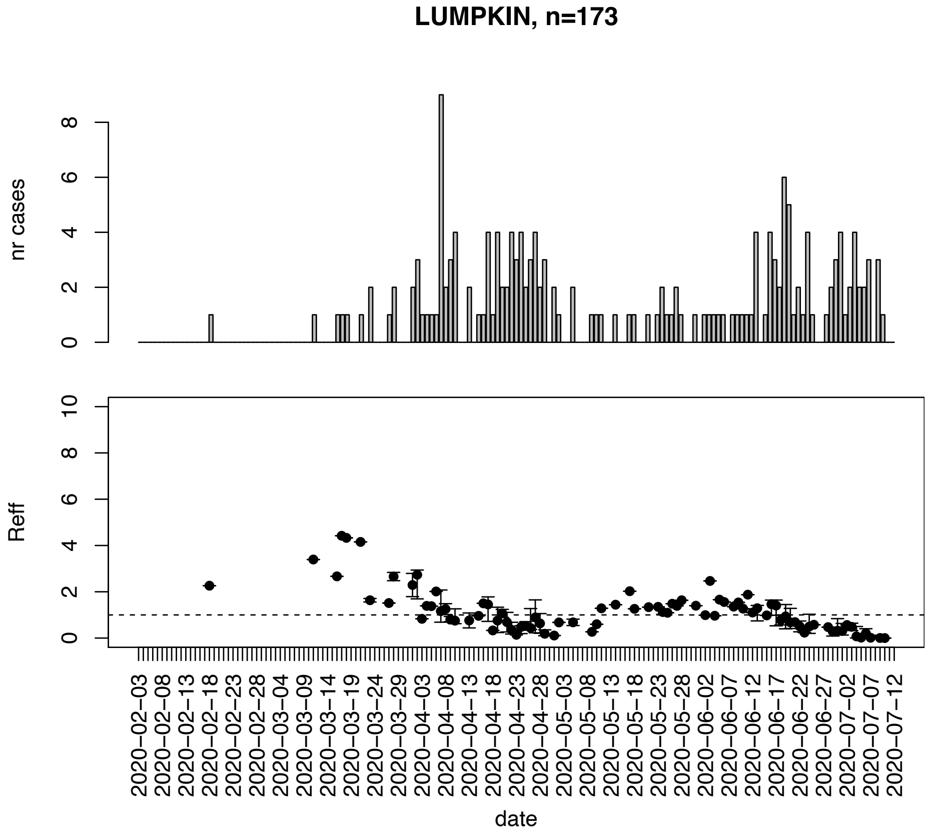


Figure. S93: Epidemic curves and reproduction number estimates until July 13th in Lumpkin county.

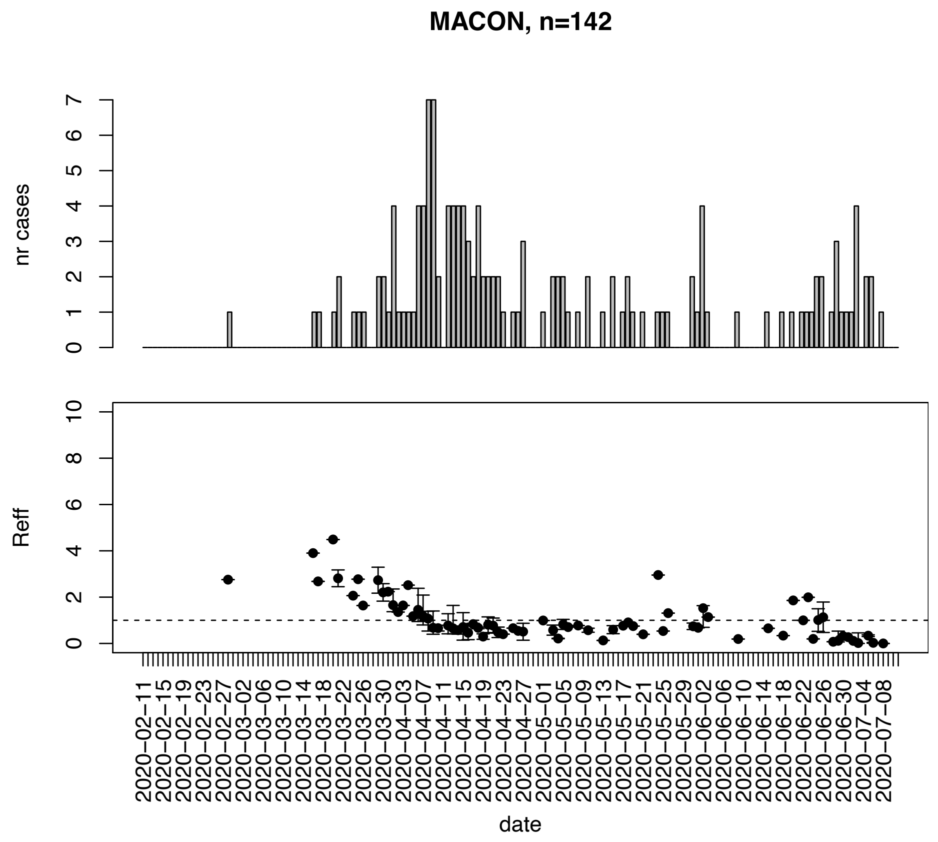


Figure. S94: Epidemic curves and reproduction number estimates until July 13th in Macon county.

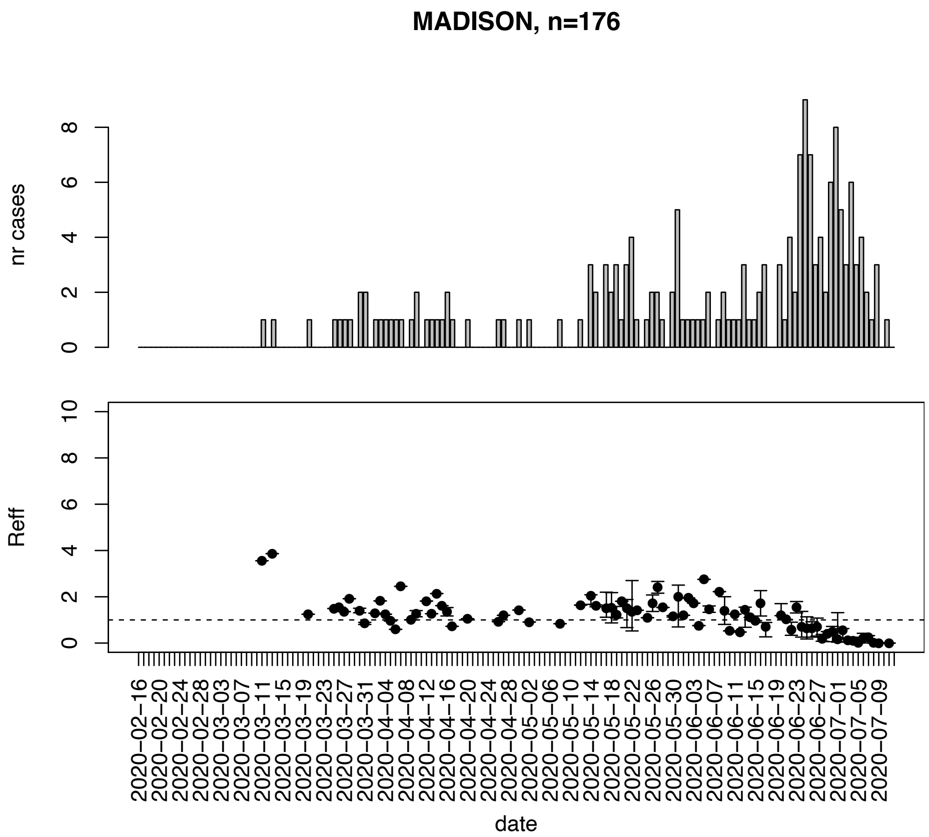


Figure. S95: Epidemic curves and reproduction number estimates until July 13th in Madison county.

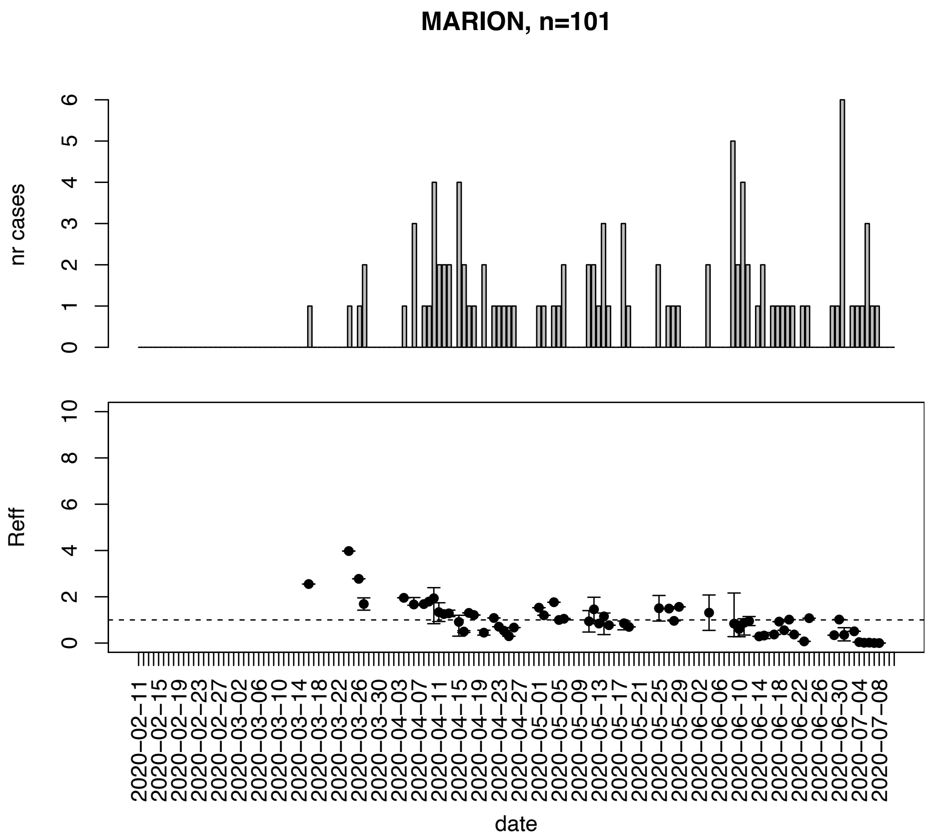


Figure. S96: Epidemic curves and reproduction number estimates until July 13th in Marion county.

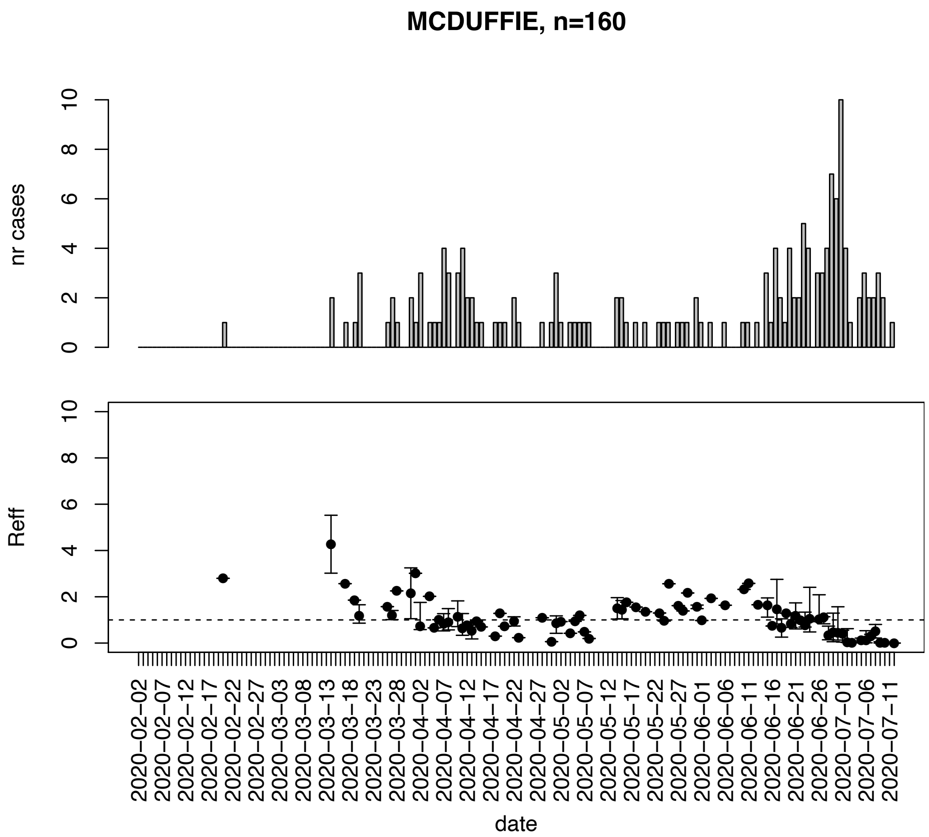


Figure. S97: Epidemic curves and reproduction number estimates until July 13th in Mcduffie county.

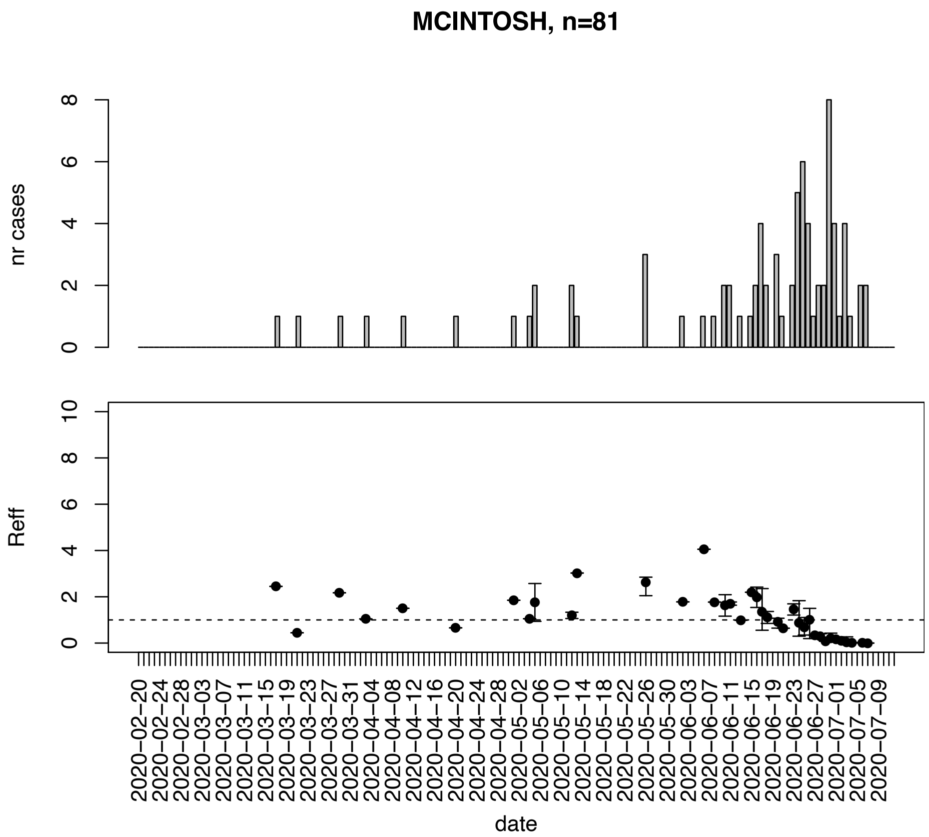


Figure. S98: Epidemic curves and reproduction number estimates until July 13th in Mcintosh county.

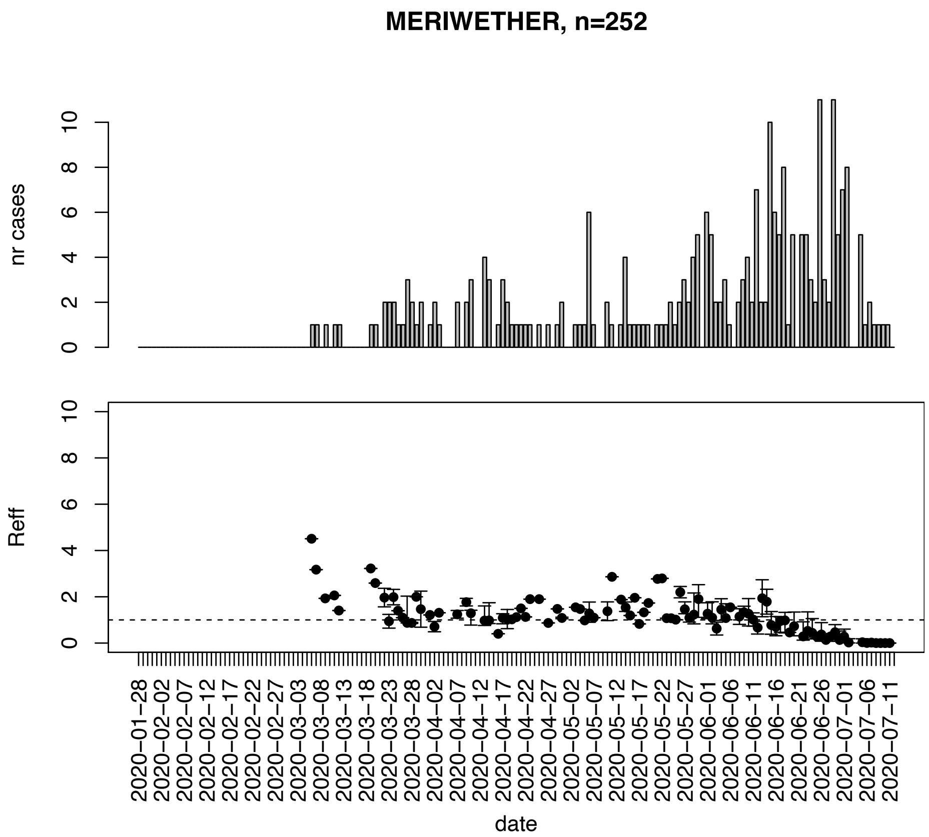


Figure. S99: Epidemic curves and reproduction number estimates until July 13th in Meriwether county.

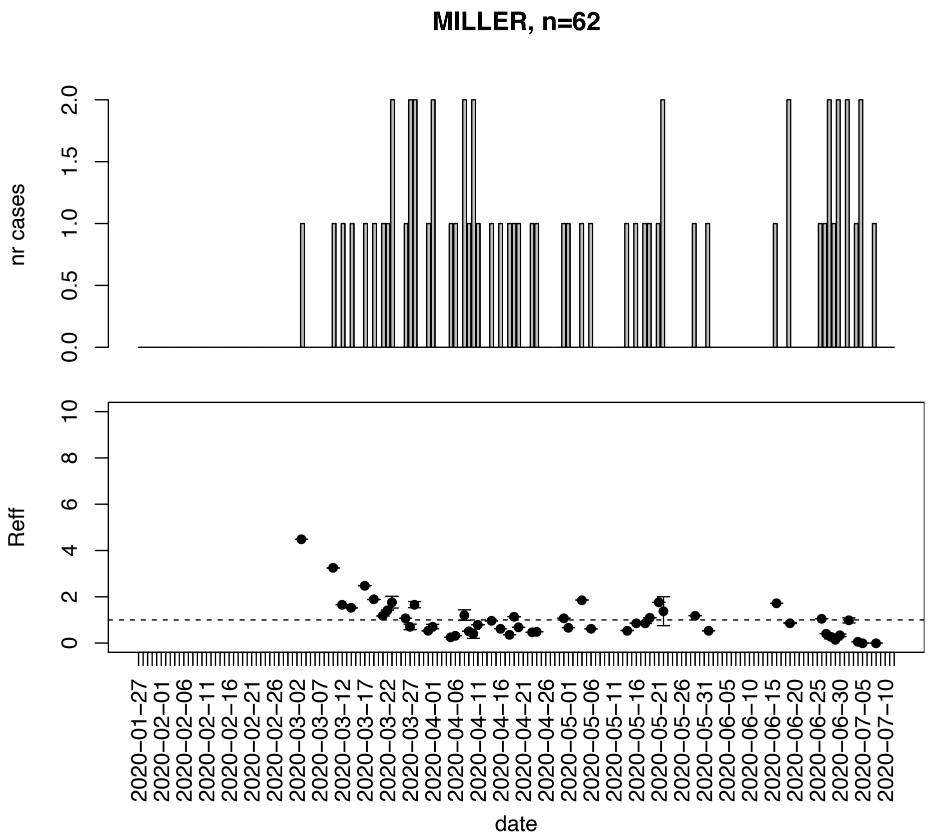


Figure. S100: Epidemic curves and reproduction number estimates until July 13th in Miller county.

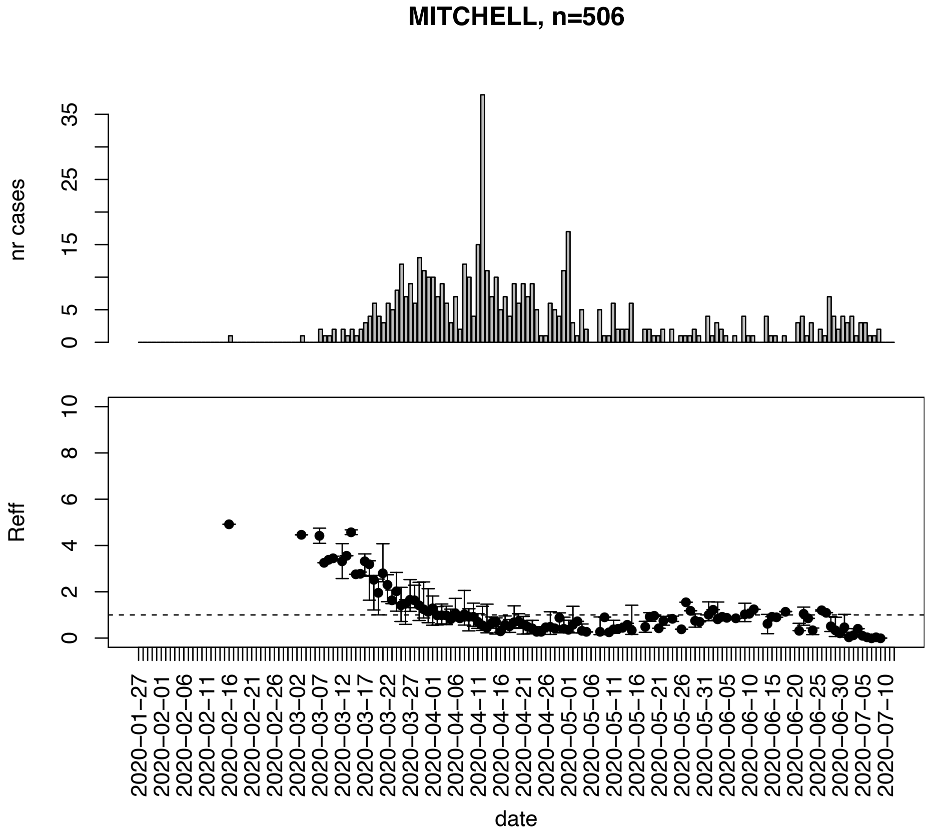


Figure. S101: Epidemic curves and reproduction number estimates until July 13th in Mitchell county.

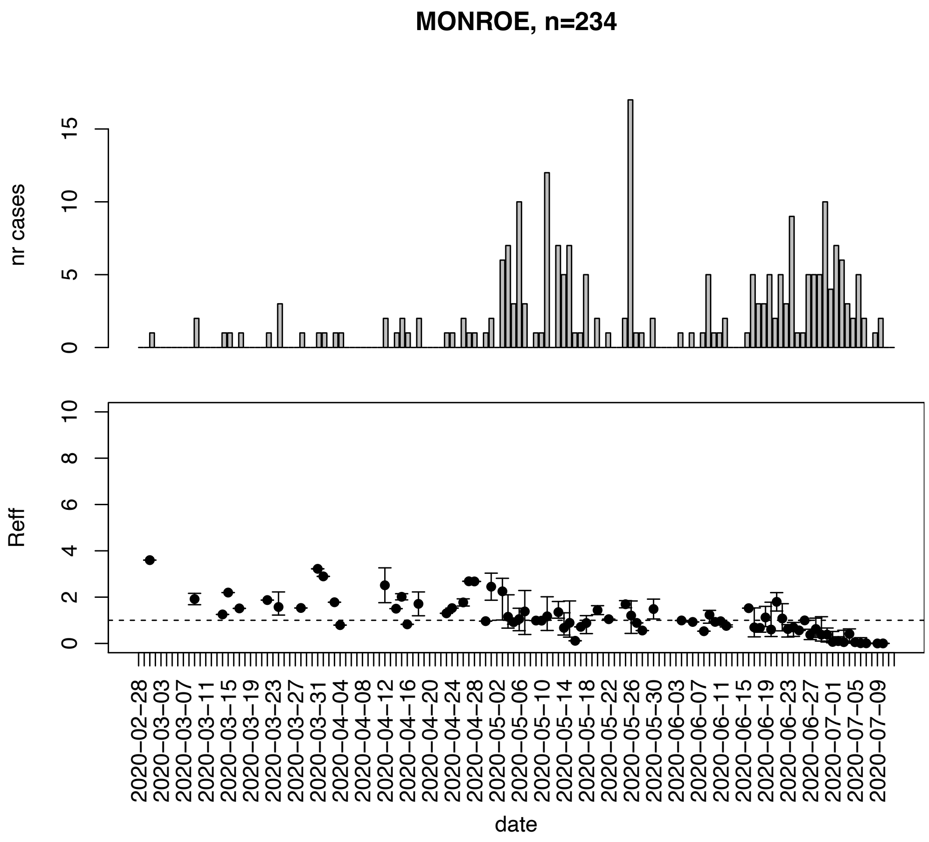


Figure. S102: Epidemic curves and reproduction number estimates until July 13th in Monroe county.

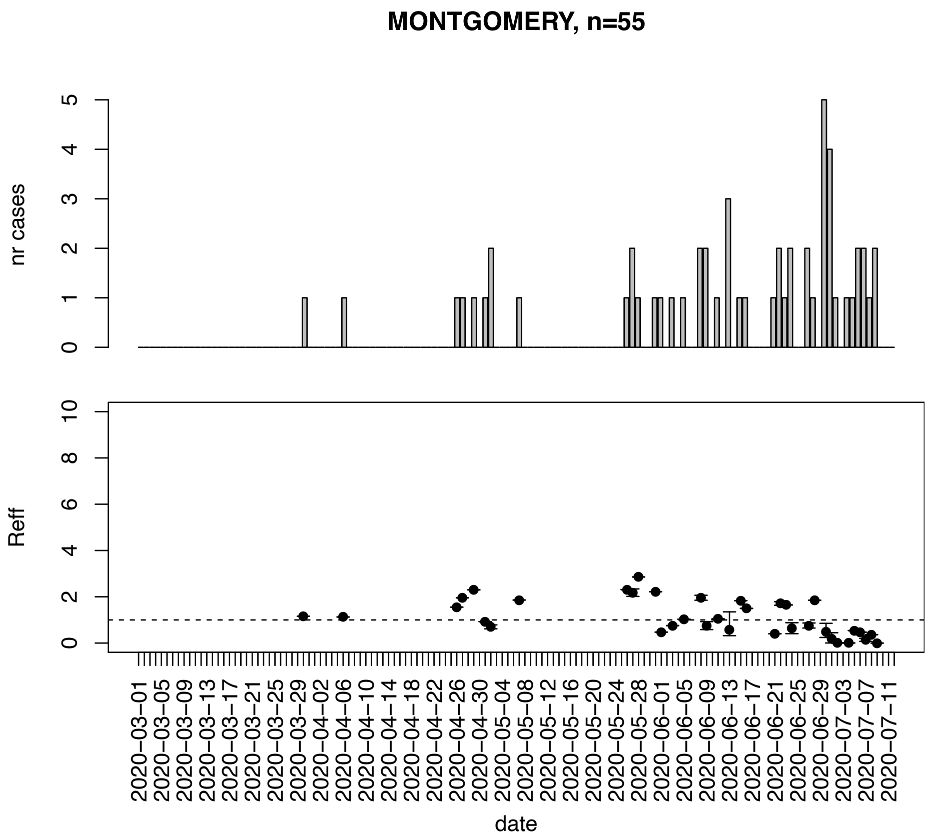


Figure. S103: Epidemic curves and reproduction number estimates until July 13th in Montgomery county.

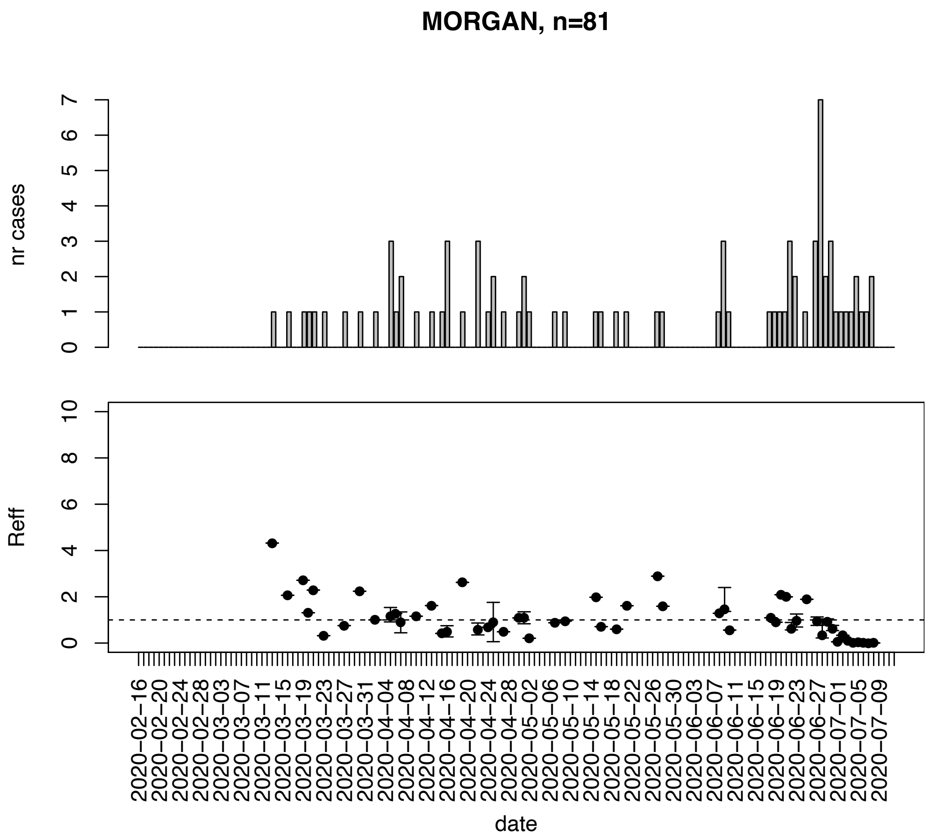


Figure. S104: Epidemic curves and reproduction number estimates until July 13th in Morgan county.

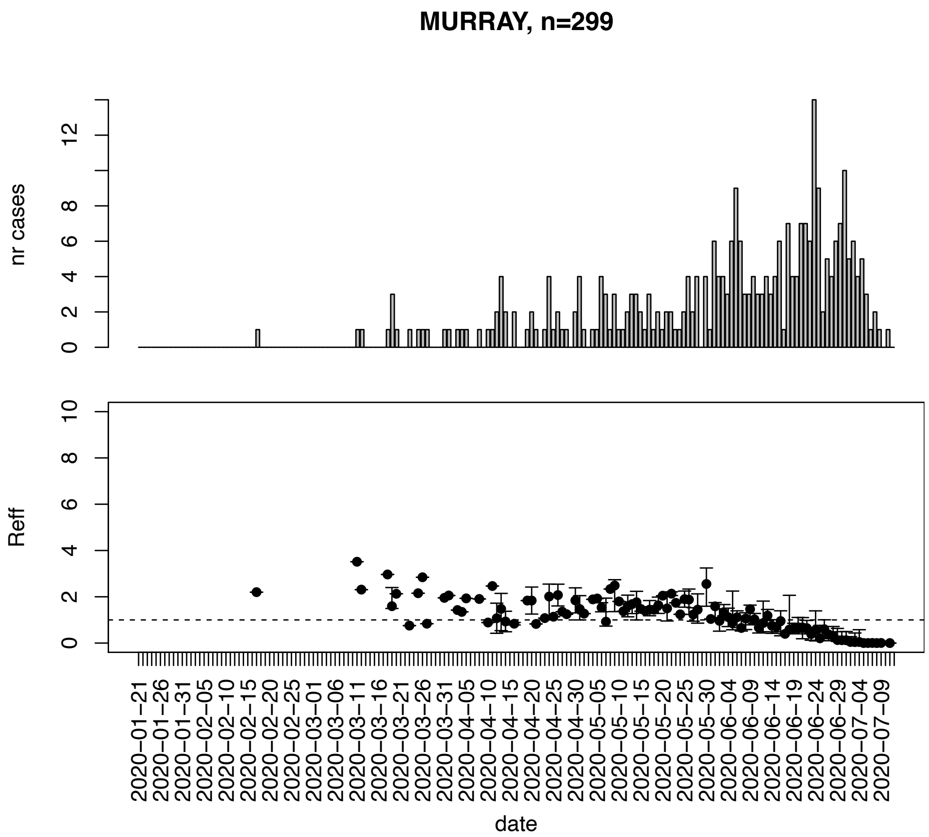


Figure. S105: Epidemic curves and reproduction number estimates until July 13th in Murray county.

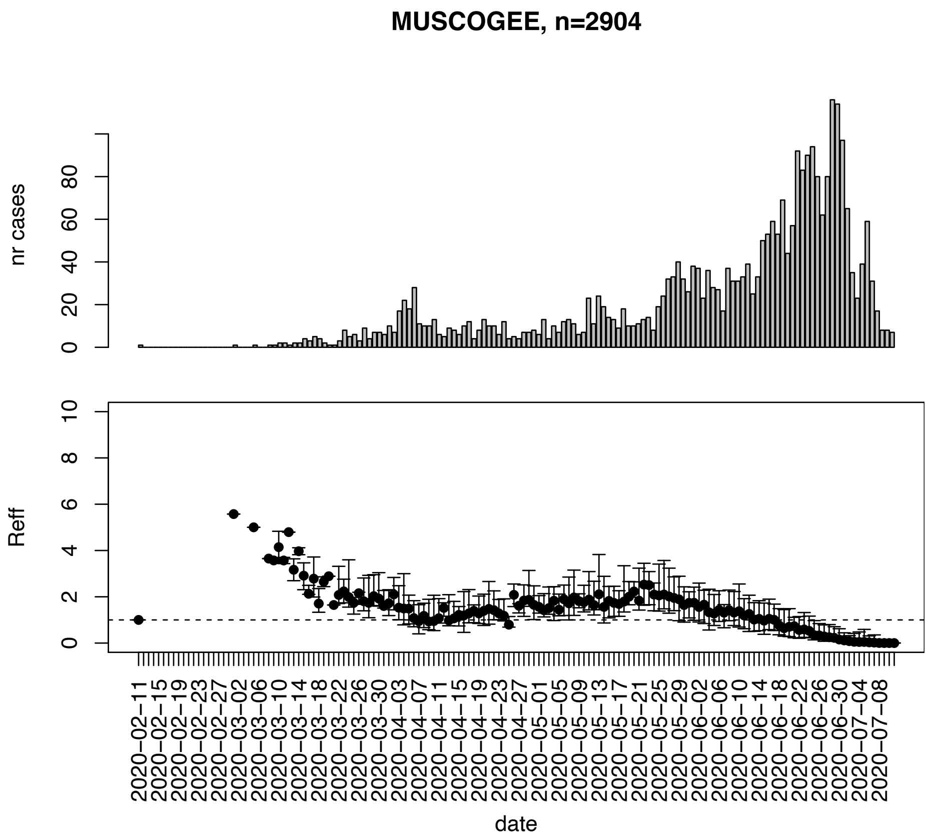


Figure. S106: Epidemic curves and reproduction number estimates until July 13th in Muscogee county.

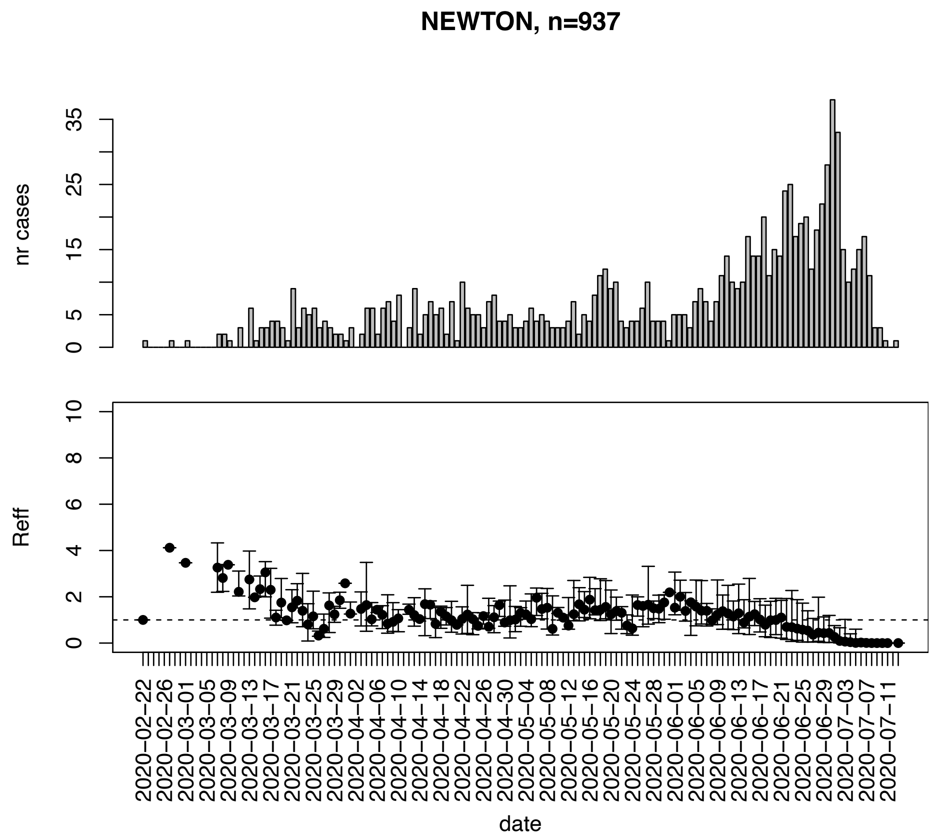


Figure. S107: Epidemic curves and reproduction number estimates until July 13th in Newton county.

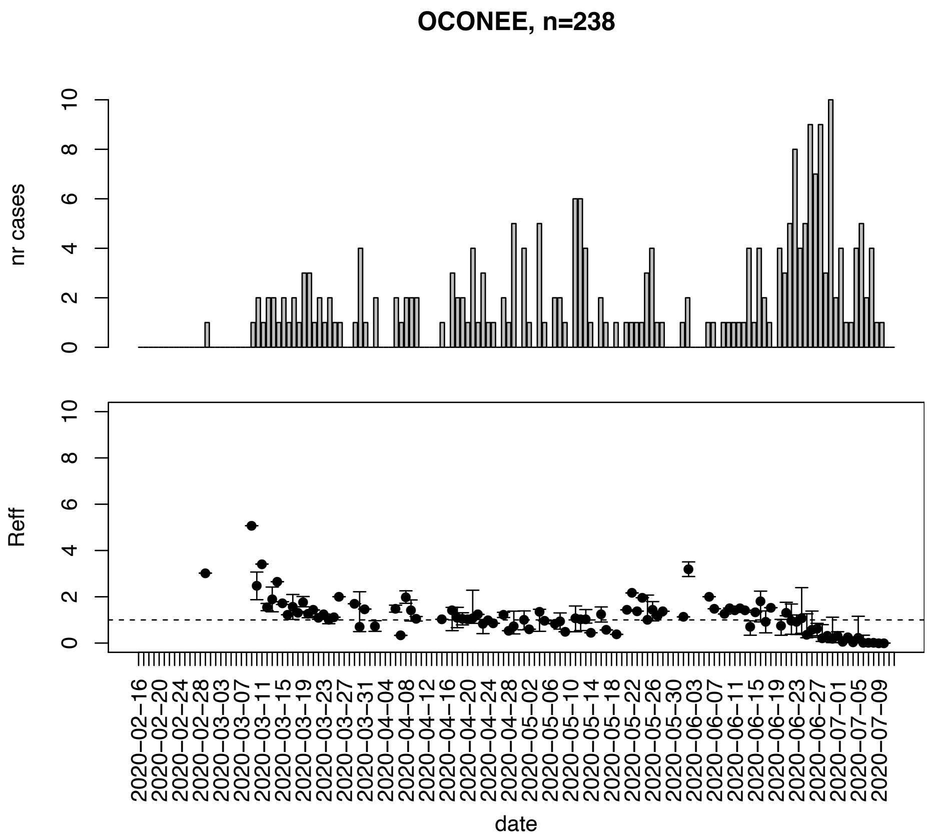


Figure. S108: Epidemic curves and reproduction number estimates until July 13th in Oconee county.

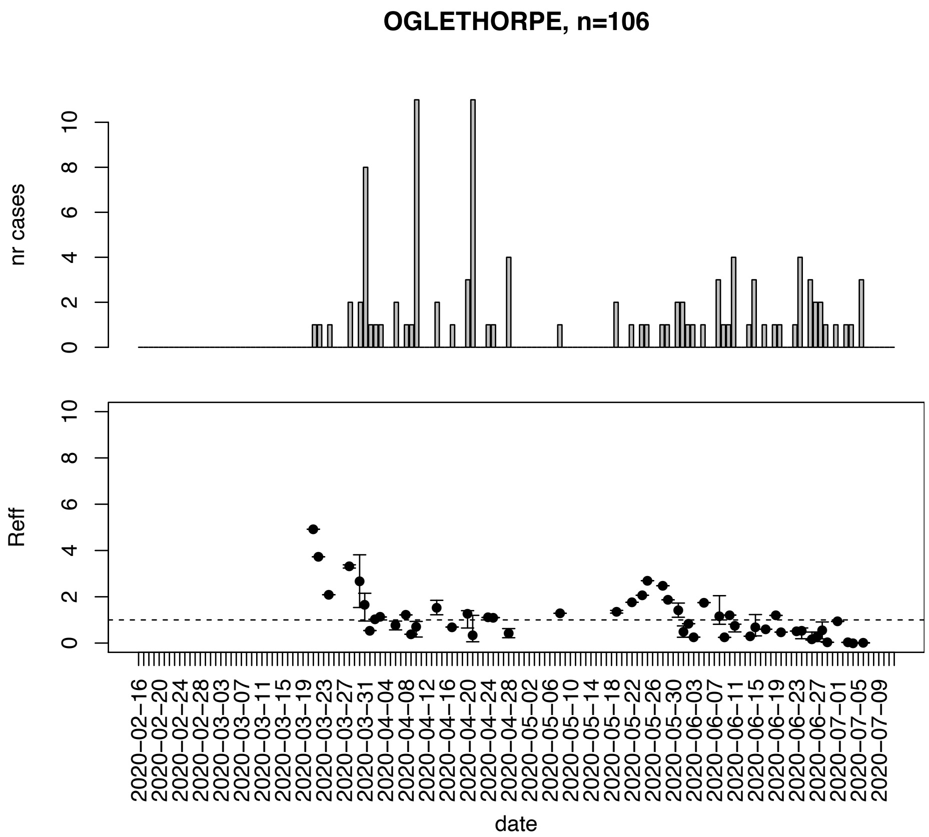


Figure. S109: Epidemic curves and reproduction number estimates until July 13th in Oglethorpe county.

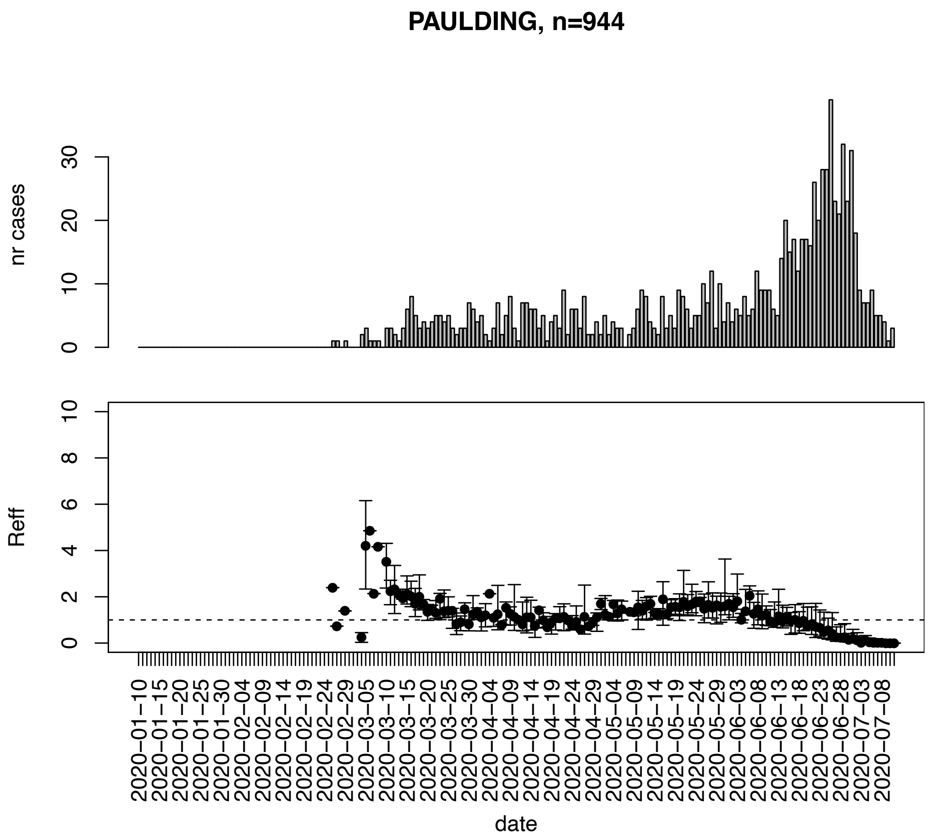


Figure. S110: Epidemic curves and reproduction number estimates until July 13th in Paulding county.

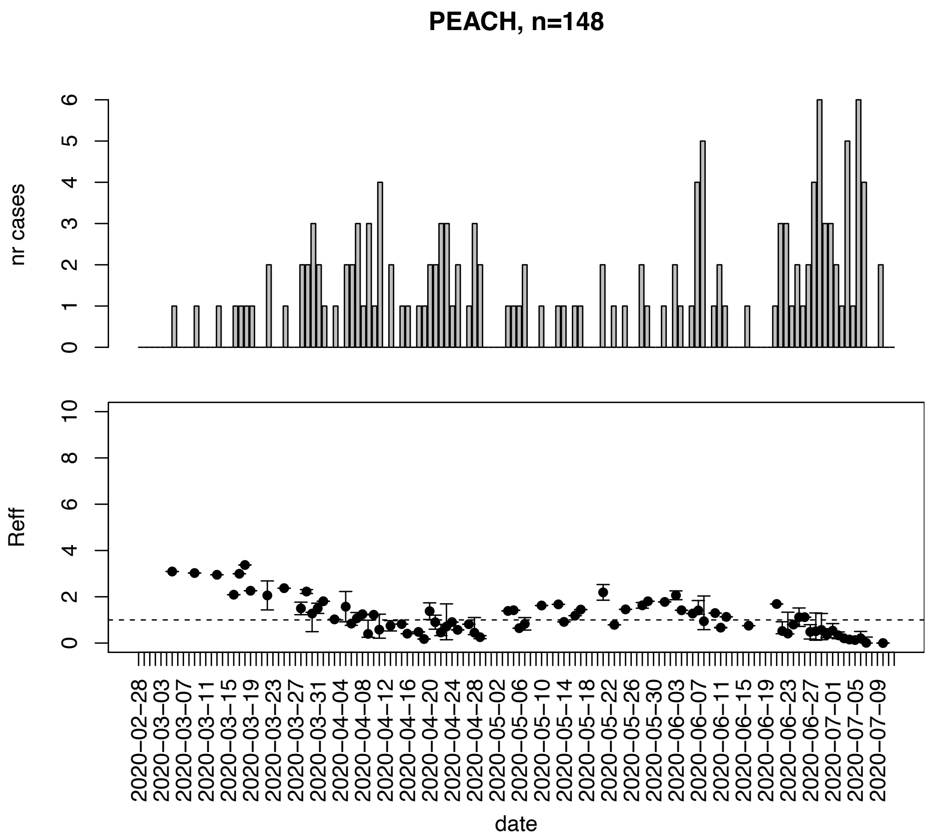


Figure. S111: Epidemic curves and reproduction number estimates until July 13th in Peach county.

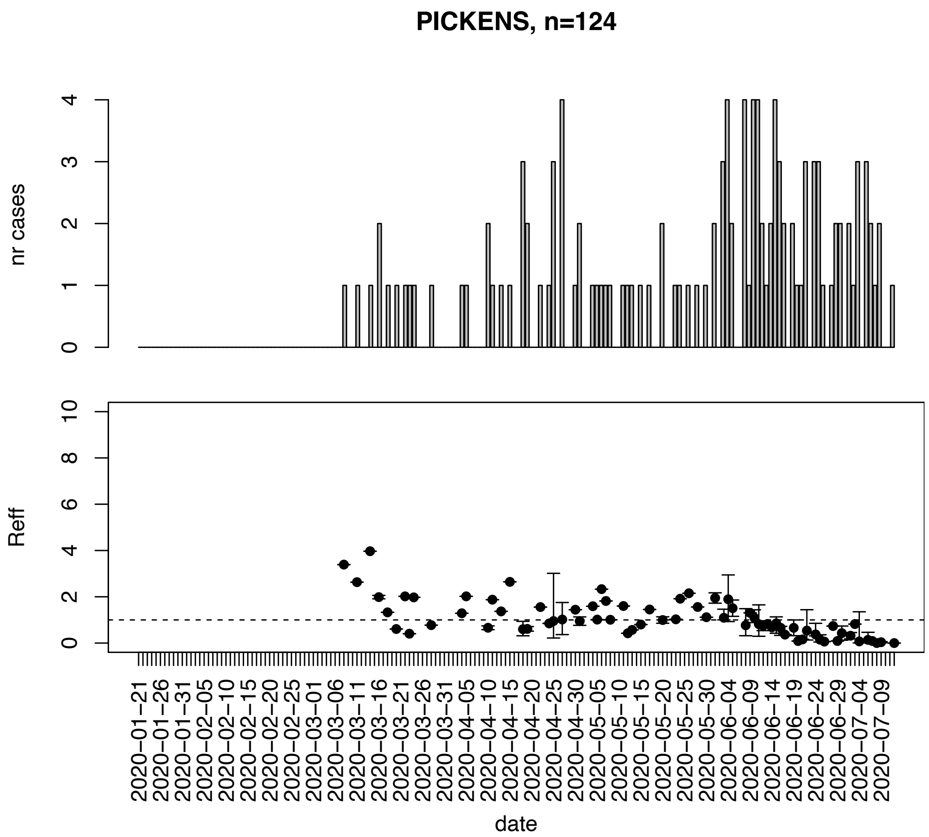


Figure. S112: Epidemic curves and reproduction number estimates until July 13th in Pickens county.

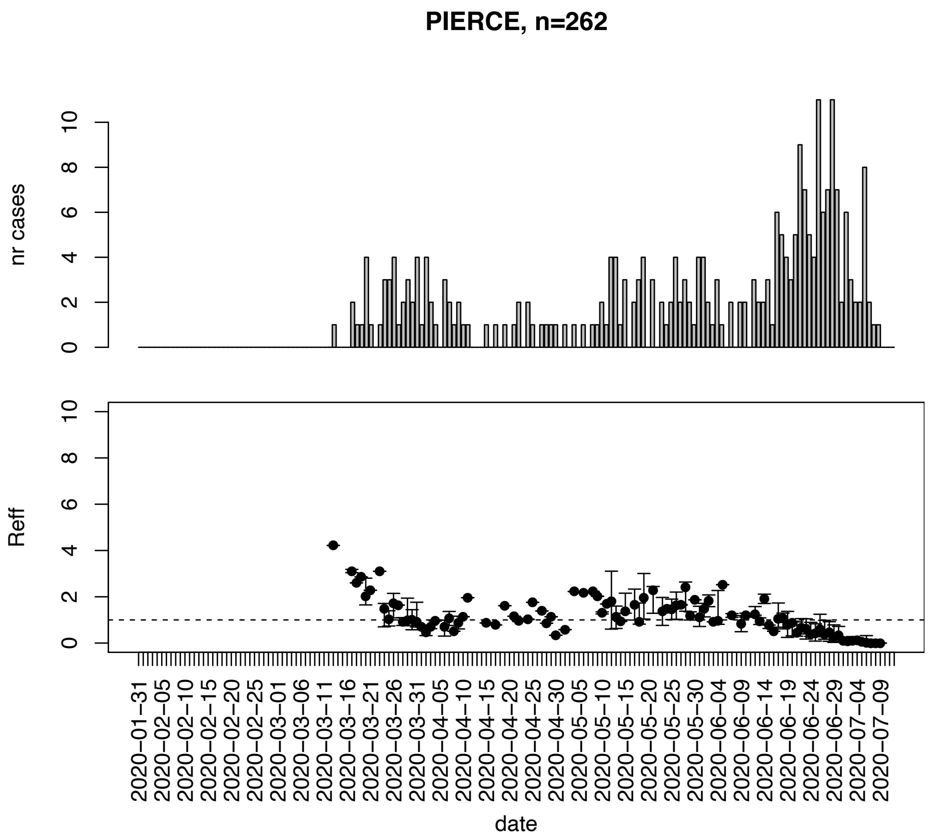


Figure. S113: Epidemic curves and reproduction number estimates until July 13th in Pierce county.

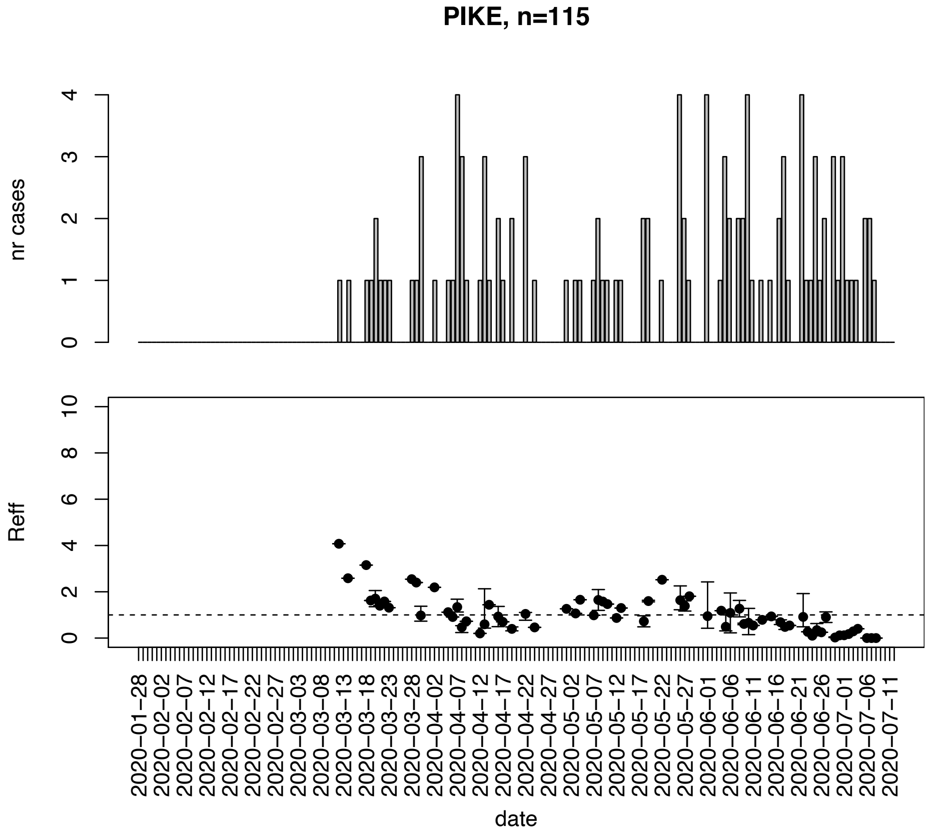


Figure. S114: Epidemic curves and reproduction number estimates until July 13th in Pike county.

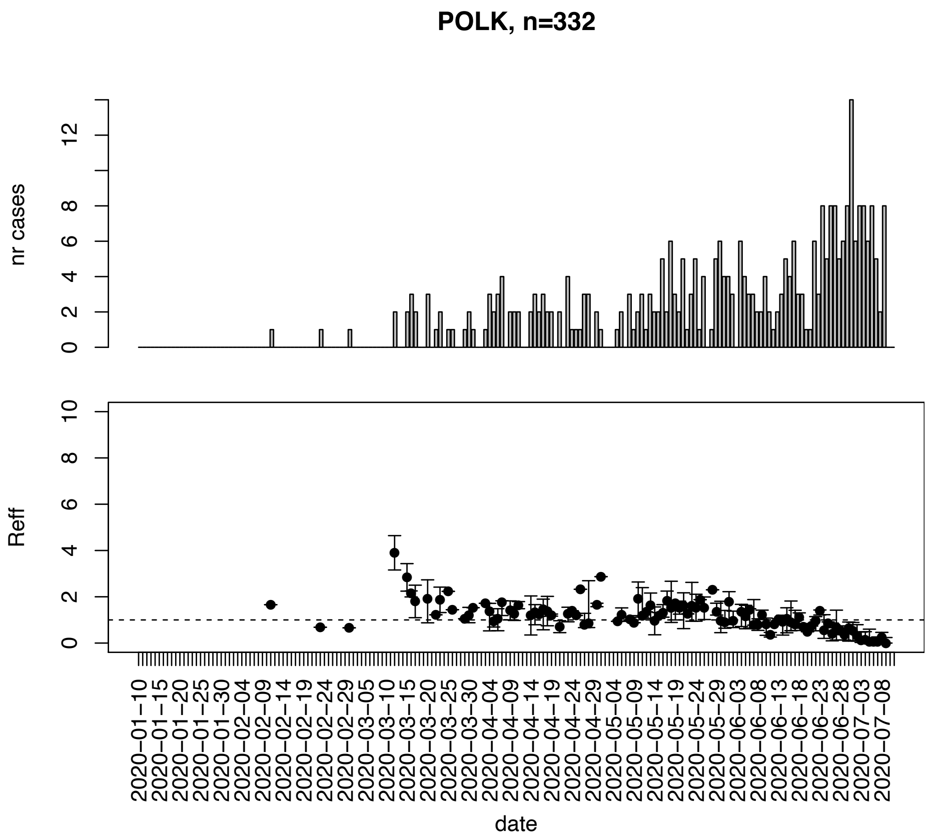


Figure. S115: Epidemic curves and reproduction number estimates until July 13th in Polk county.

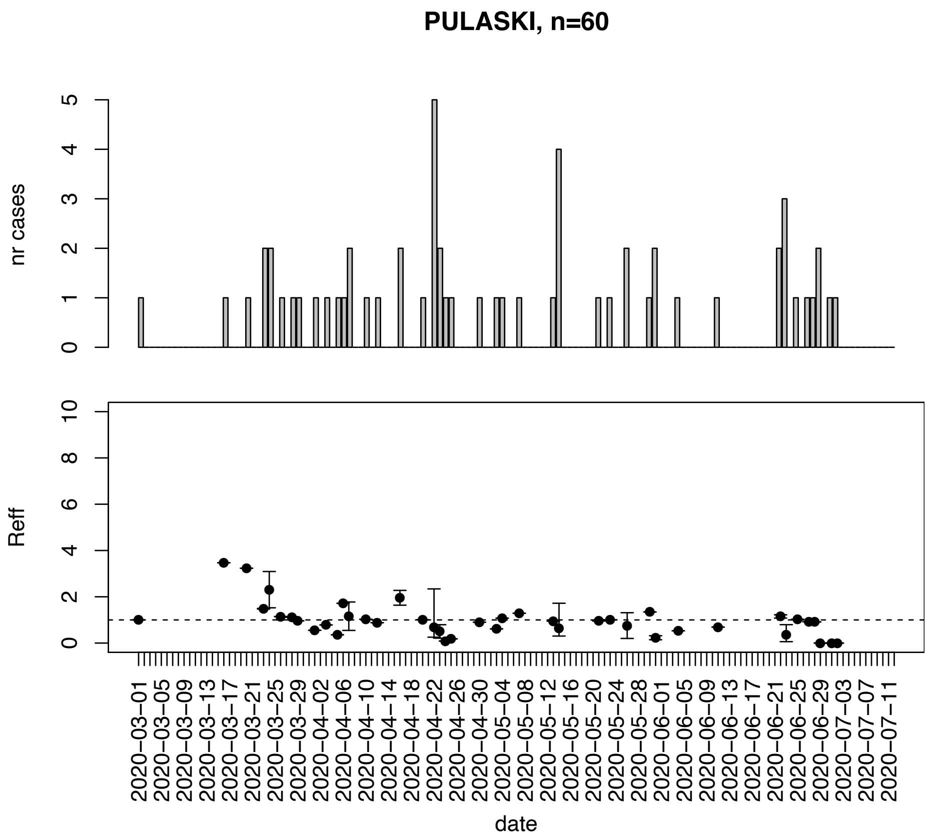


Figure. S116: Epidemic curves and reproduction number estimates until July 13th in Pulaski county.

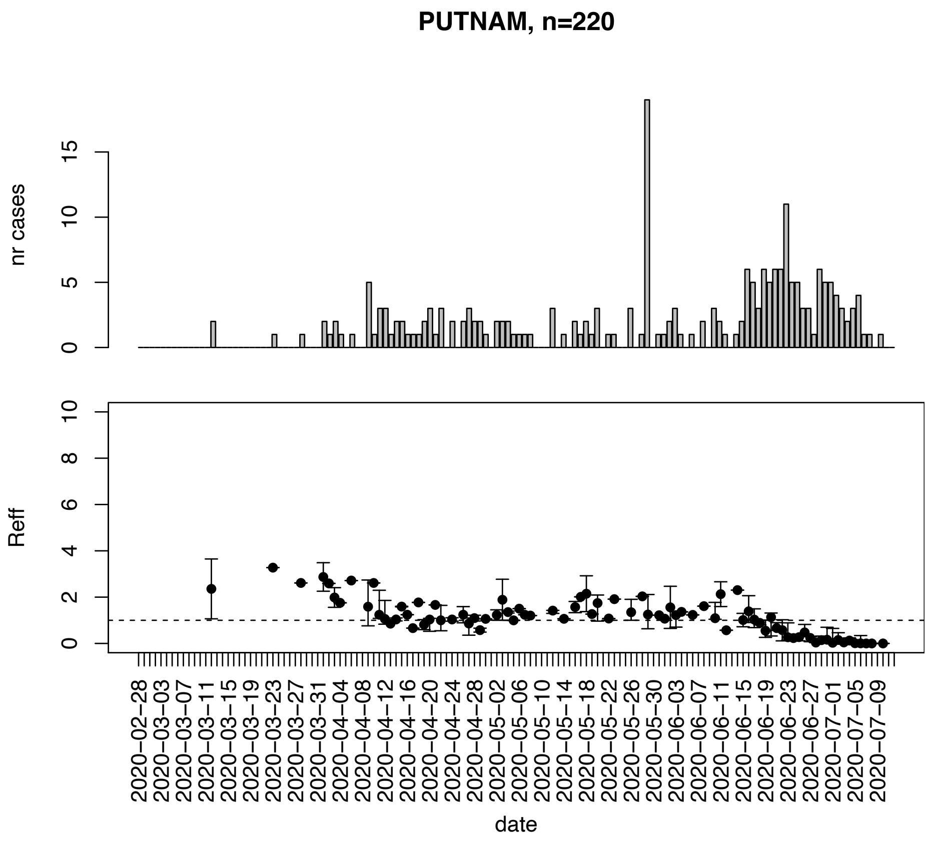


Figure. S117: Epidemic curves and reproduction number estimates until July 13th in Putnam county.

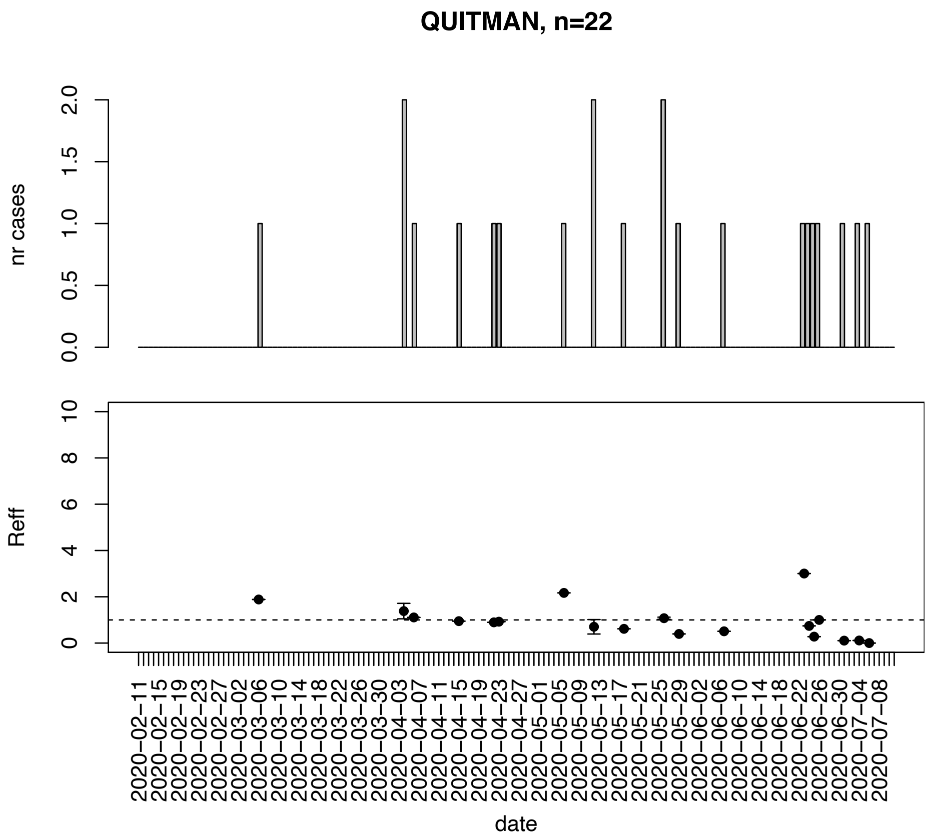


Figure. S118: Epidemic curves and reproduction number estimates until July 13th in Quitman county.

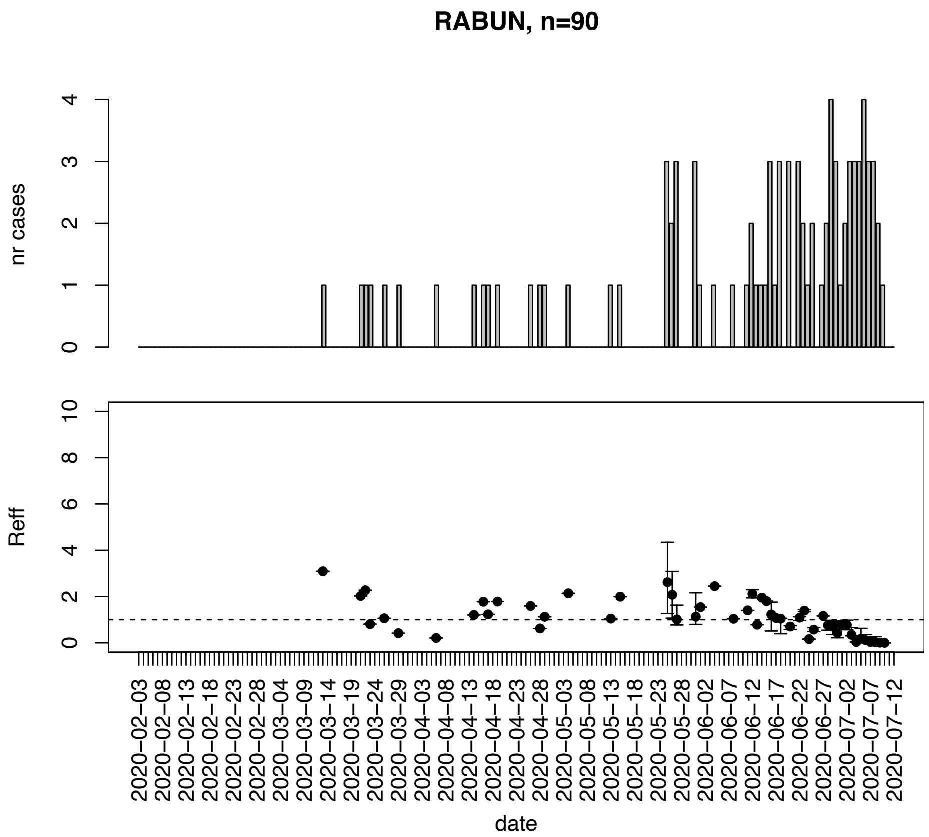


Figure. S119: Epidemic curves and reproduction number estimates until July 13th in Rabun county.

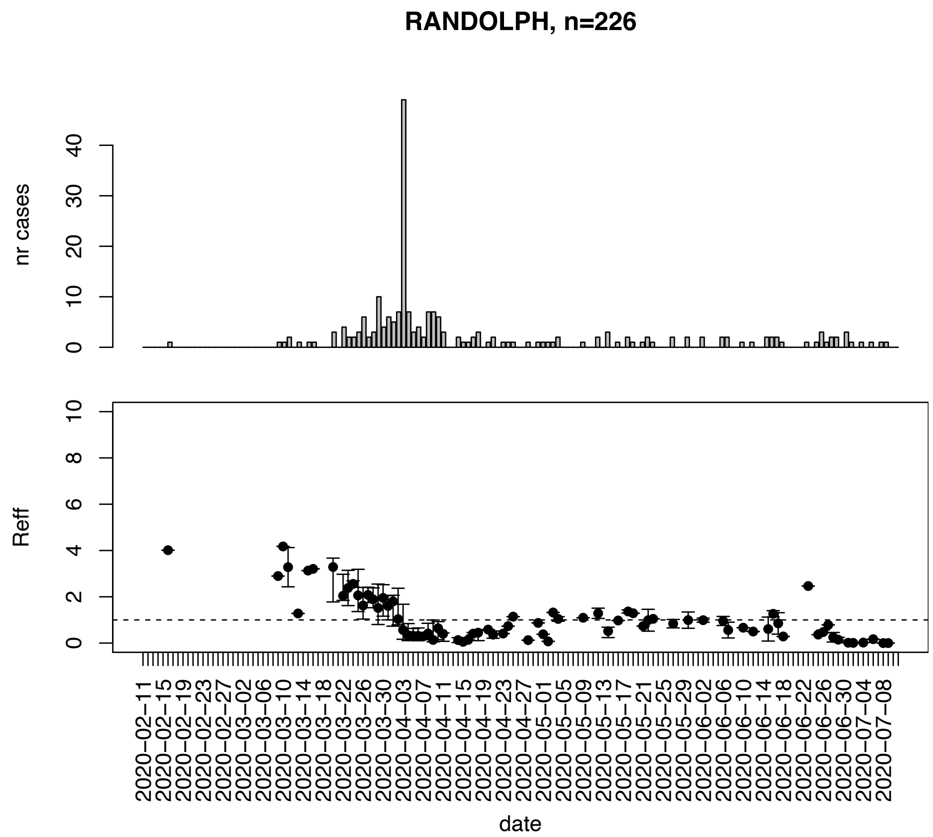


Figure. S120: Epidemic curves and reproduction number estimates until July 13th in Randolph county.

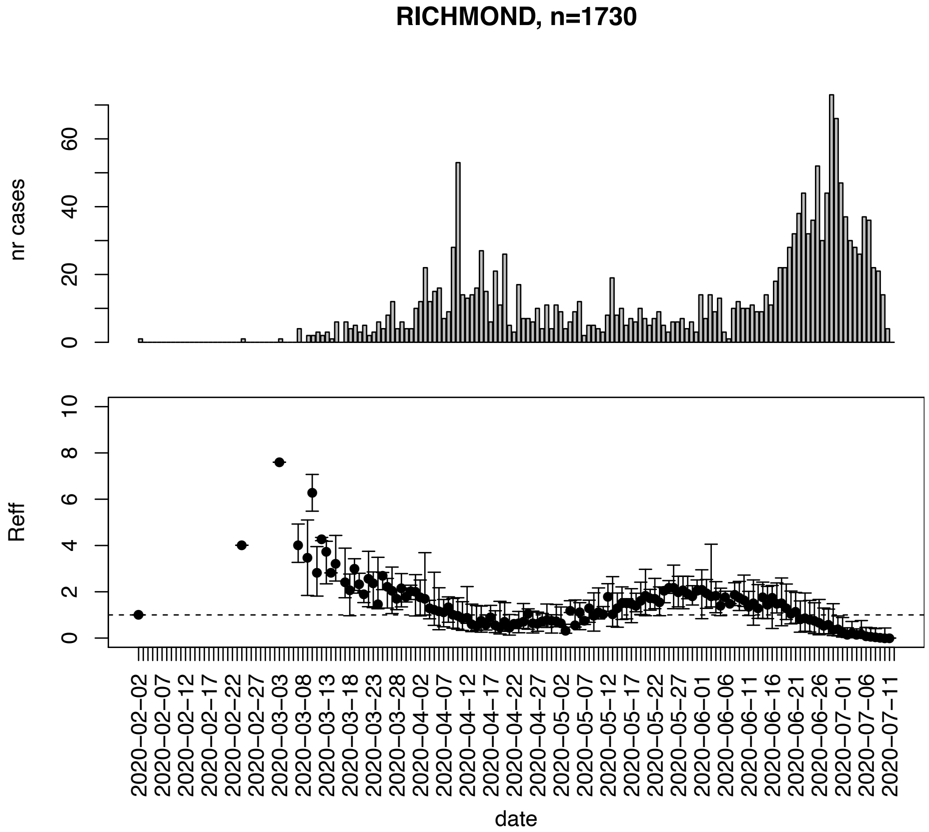


Figure. S121: Epidemic curves and reproduction number estimates until July 13th in Richmond county.

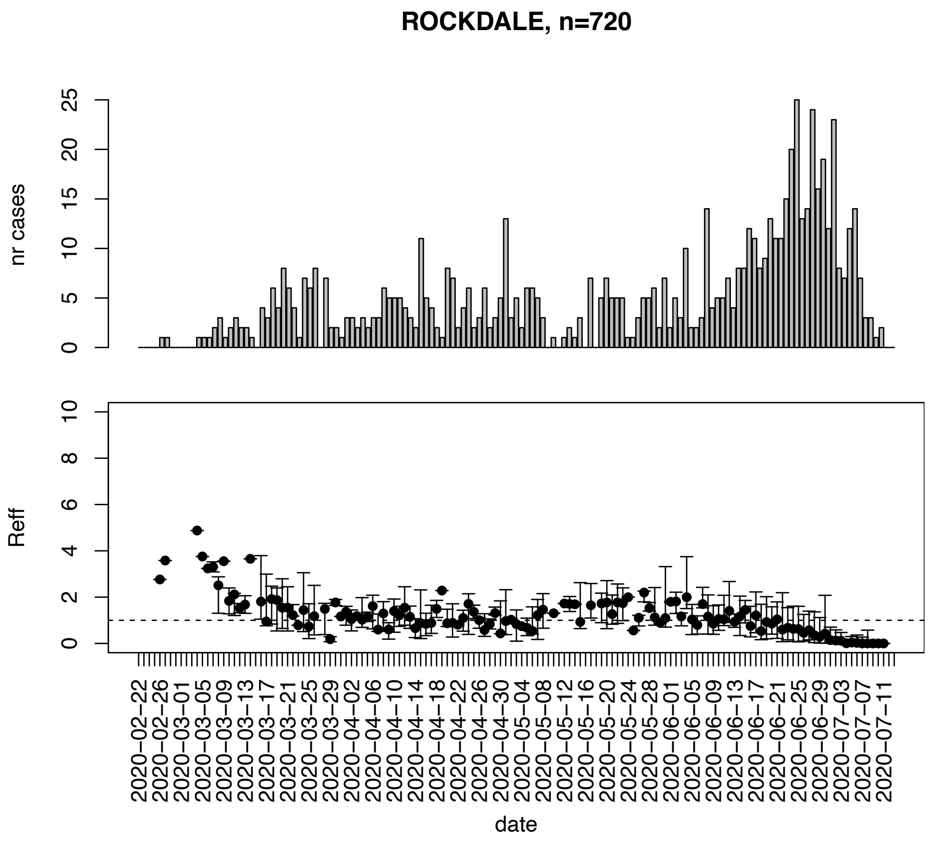


Figure. S122: Epidemic curves and reproduction number estimates until July 13th in Rockdale county.

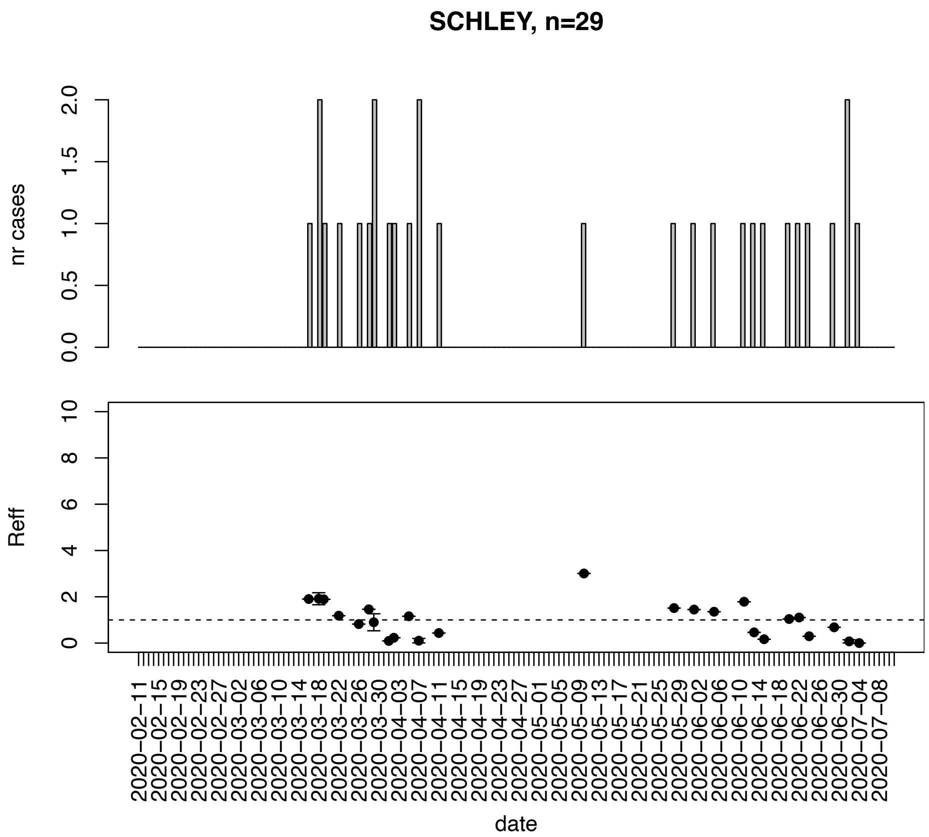


Figure. S123: Epidemic curves and reproduction number estimates until July 13th in Schley county.

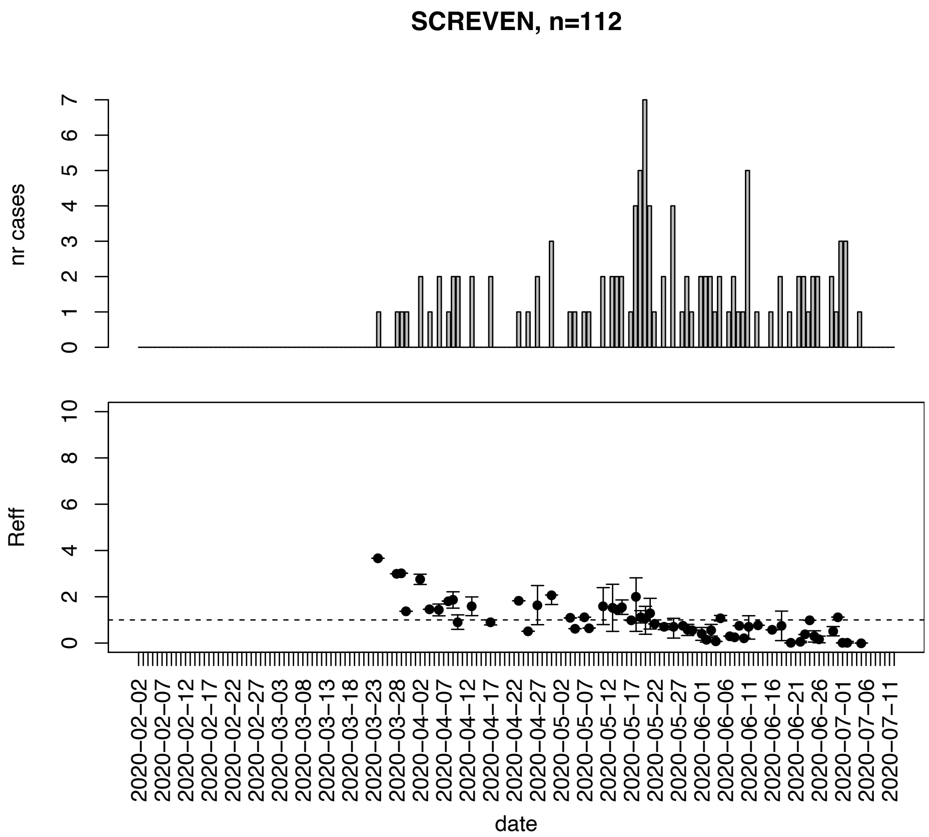


Figure. S124: Epidemic curves and reproduction number estimates until July 13th in Screven county.

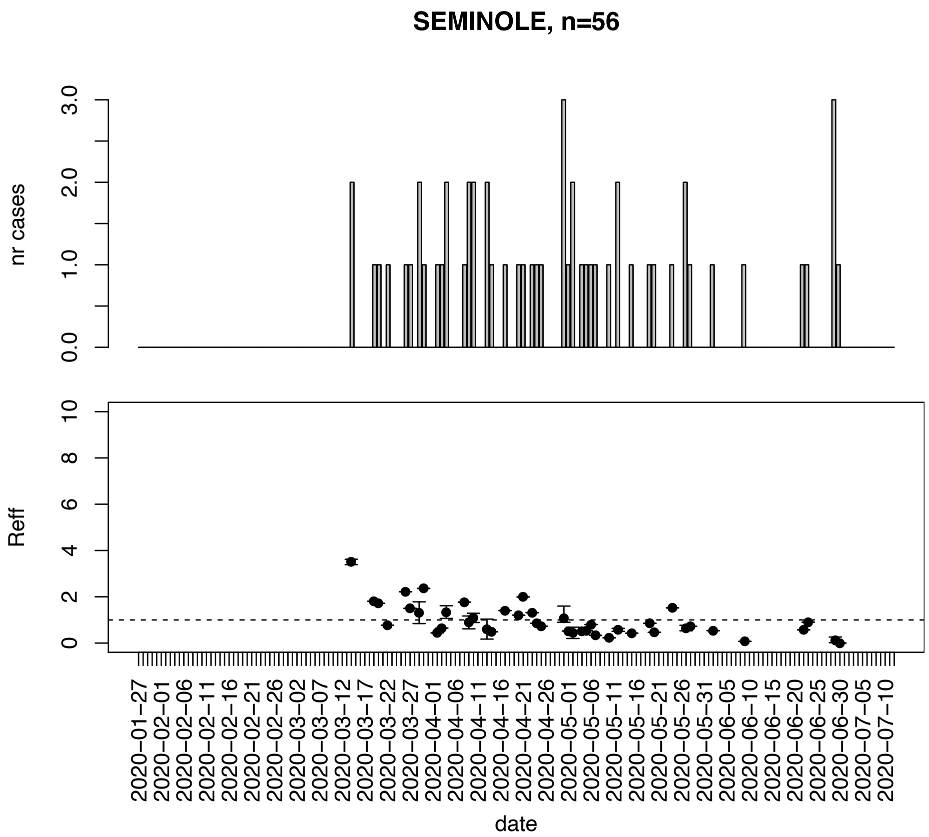


Figure. S125: Epidemic curves and reproduction number estimates until July 13th in Seminole county.

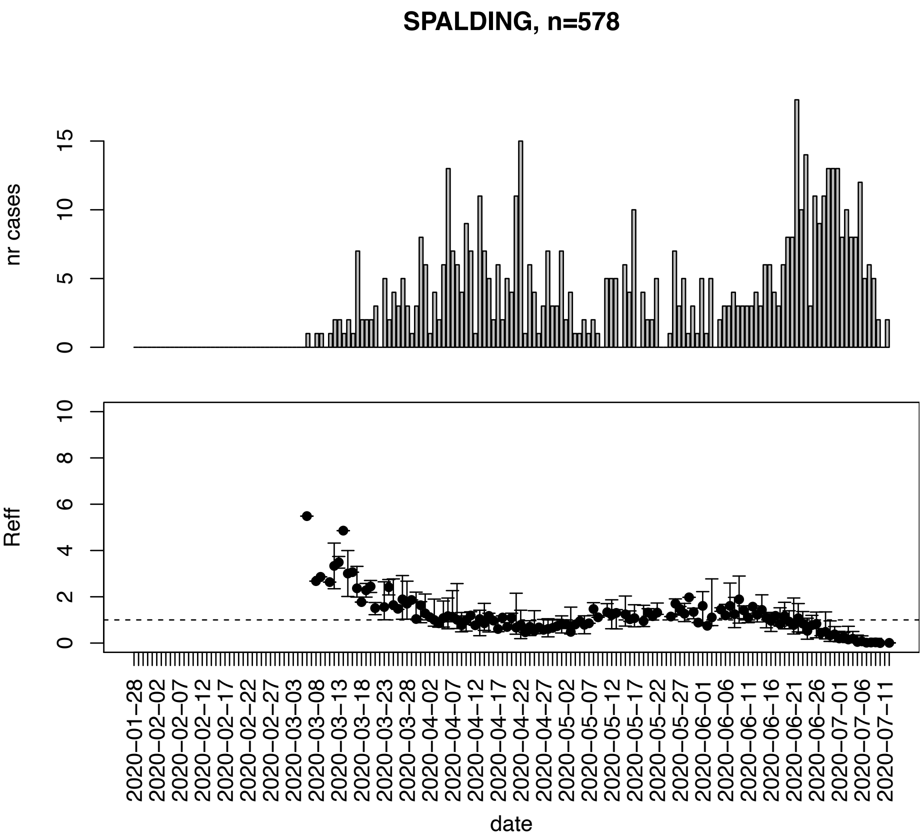


Figure. S126: Epidemic curves and reproduction number estimates until July 13th in Spalding county.

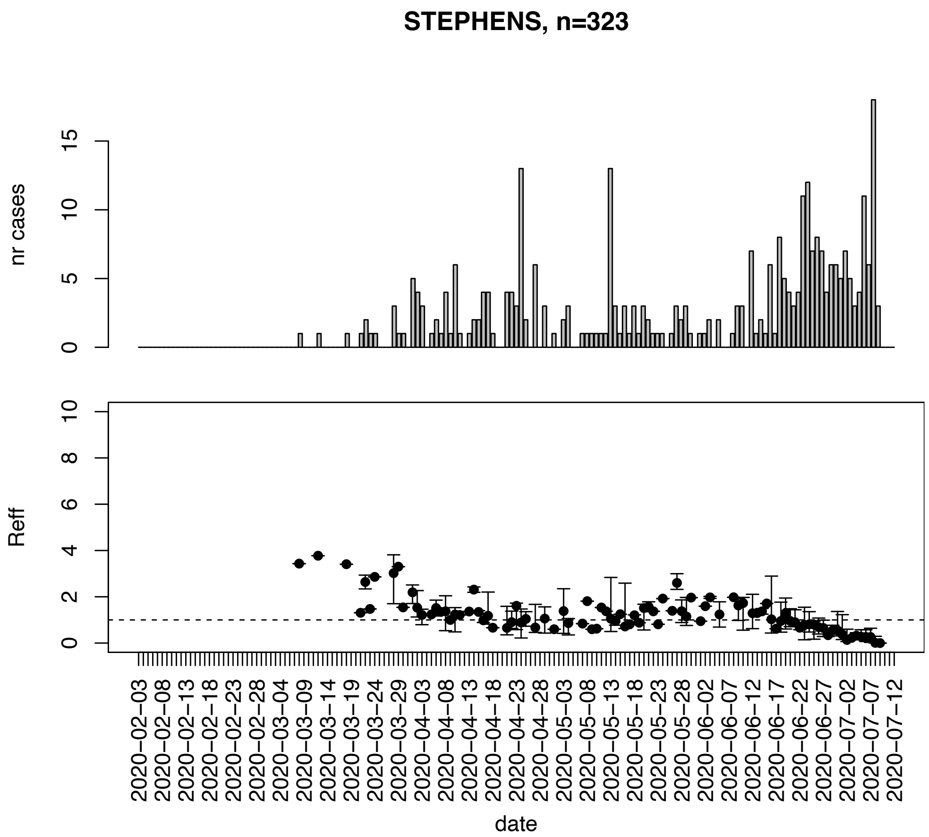


Figure. S127: Epidemic curves and reproduction number estimates until July 13th in Stephens county.

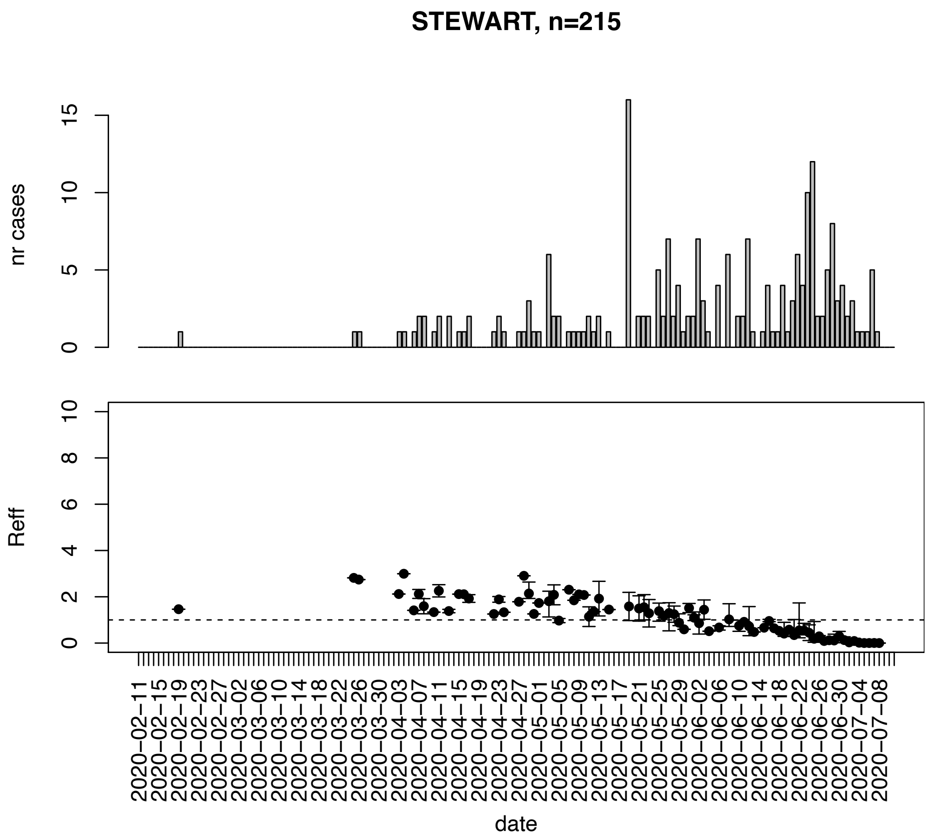


Figure. S128: Epidemic curves and reproduction number estimates until July 13th in Stewart county.

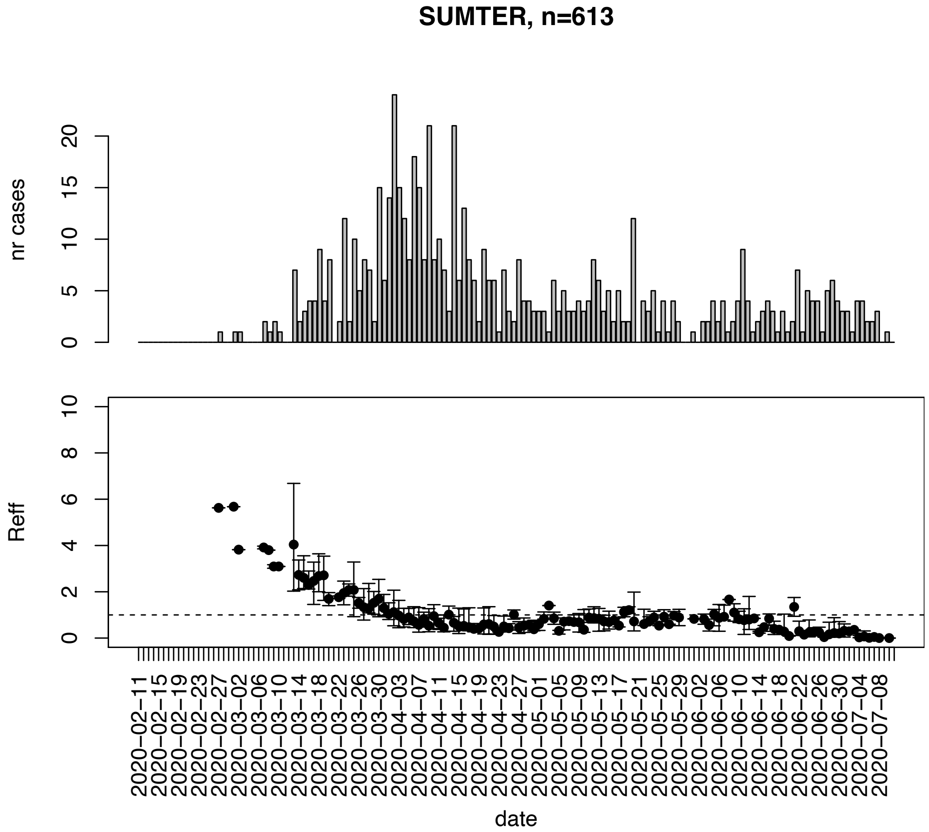


Figure. S129: Epidemic curves and reproduction number estimates until July 13th in Sumter county.

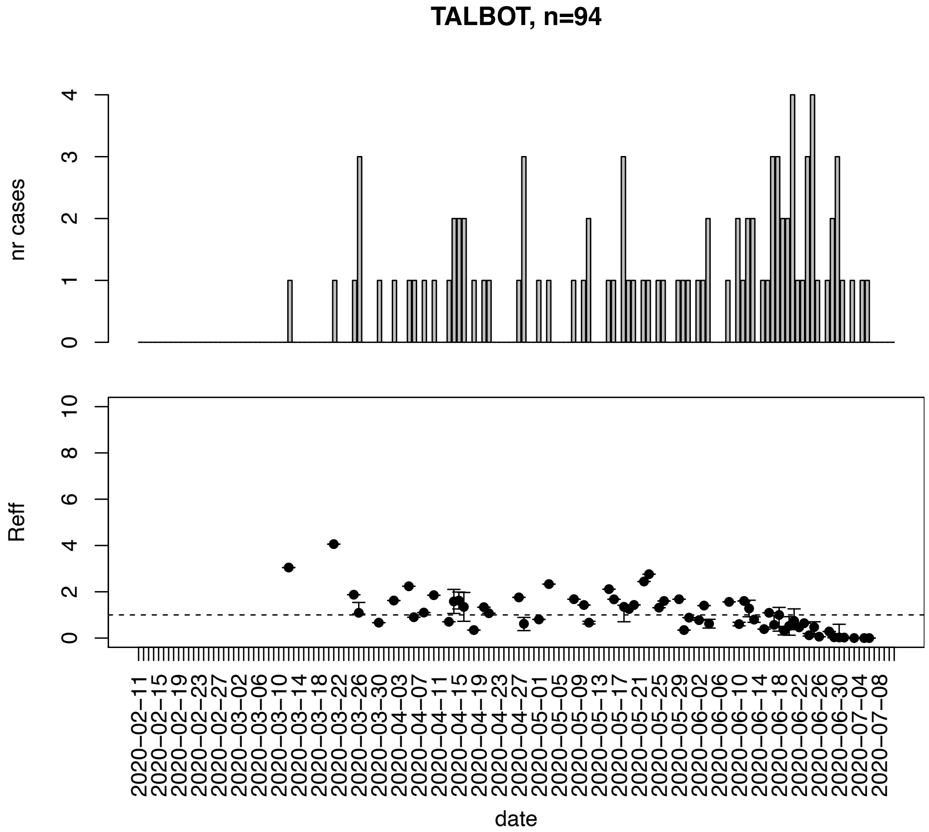


Figure. S130: Epidemic curves and reproduction number estimates until July 13th in Talbot county.

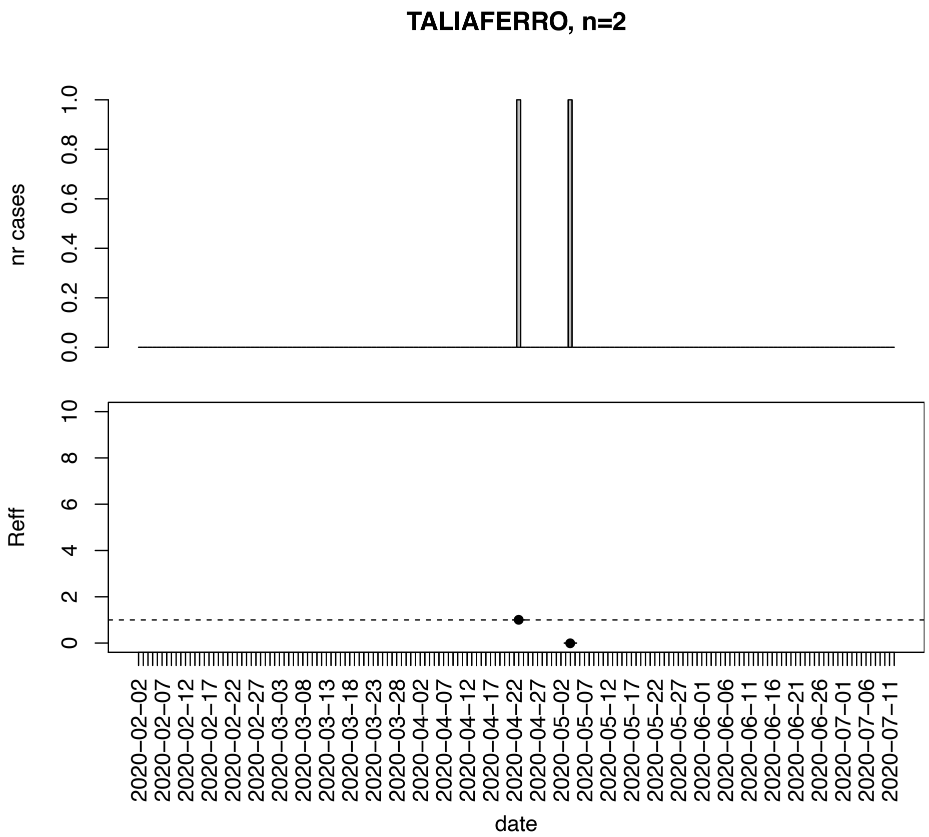


Figure. S131: Epidemic curves and reproduction number estimates until July 13th in Taliaferro county.

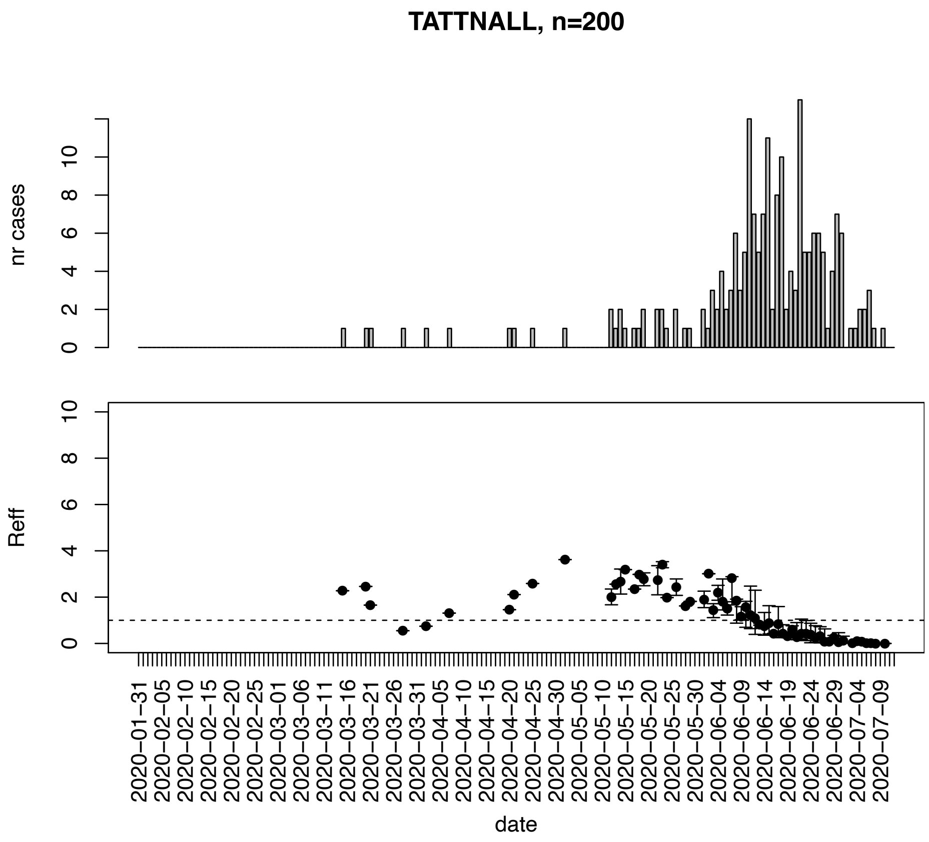


Figure. S132: Epidemic curves and reproduction number estimates until July 13th in Tattnall county.

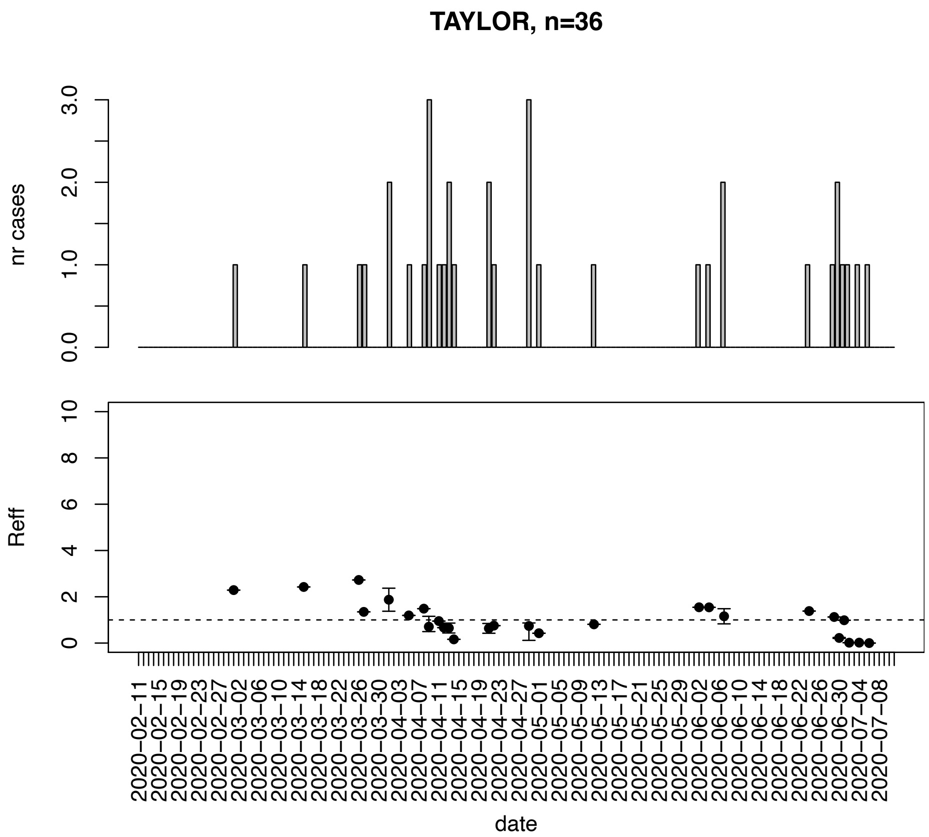


Figure. S133: Epidemic curves and reproduction number estimates until July 13th in Taylor county.

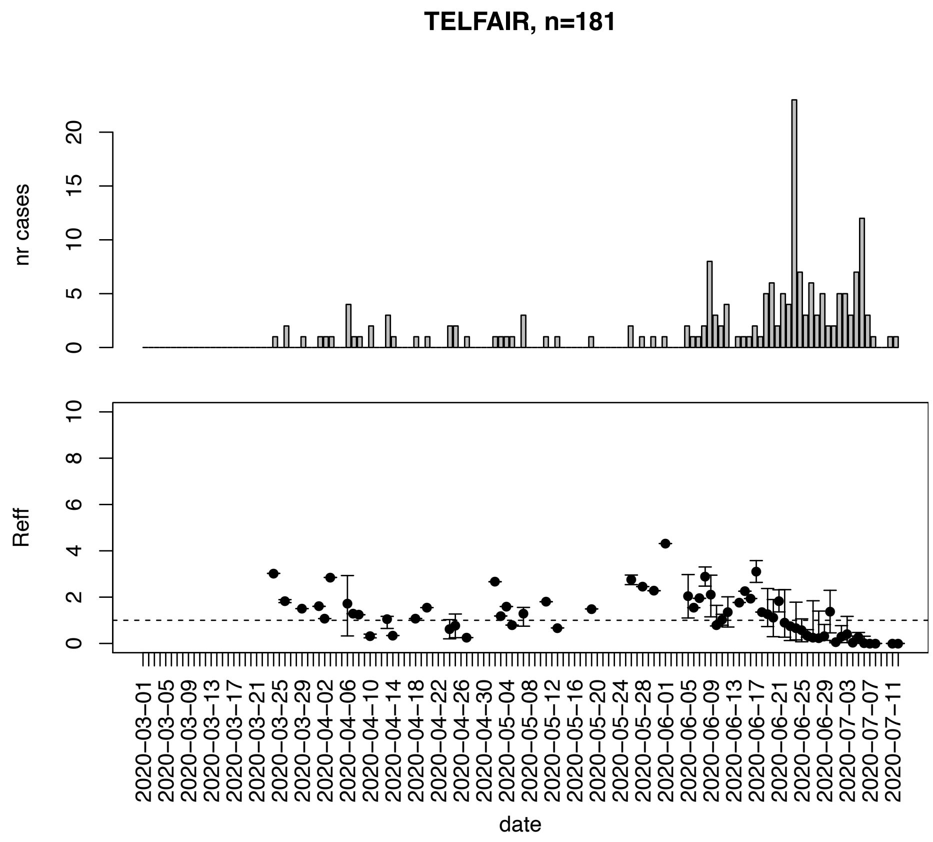


Figure. S134: Epidemic curves and reproduction number estimates until July 13th in Telfair county.

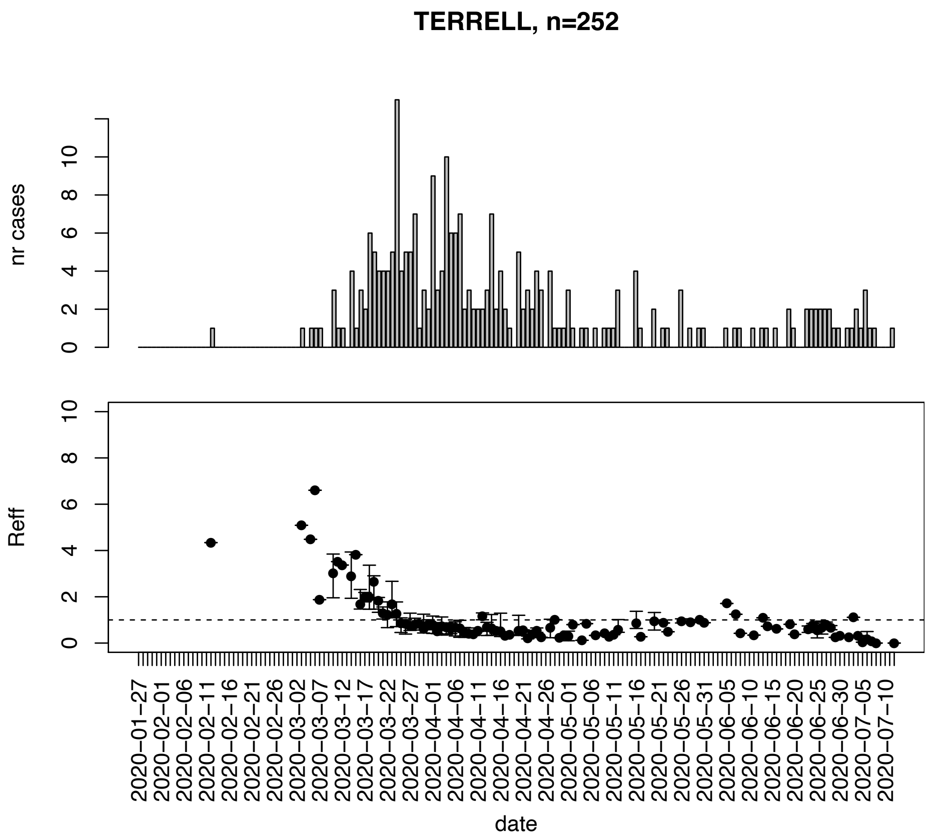


Figure. S135: Epidemic curves and reproduction number estimates until July 13th in Terrell county.

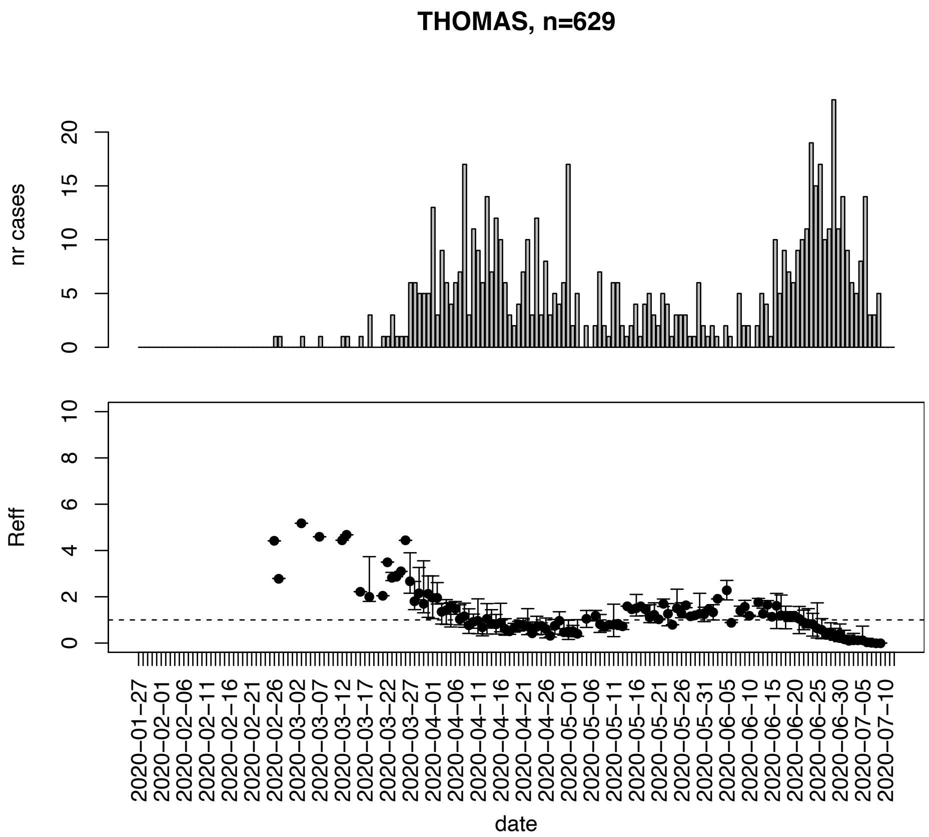


Figure. S136: Epidemic curves and reproduction number estimates until July 13th in Thomas county.

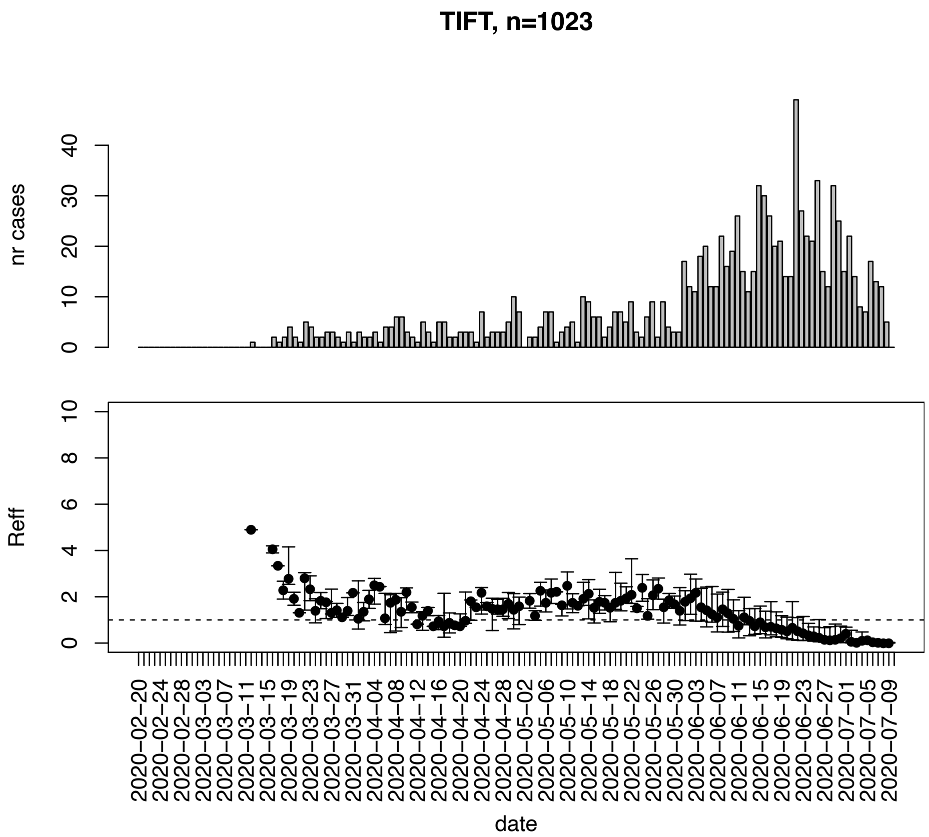


Figure. S137: Epidemic curves and reproduction number estimates until July 13th in Tift county.

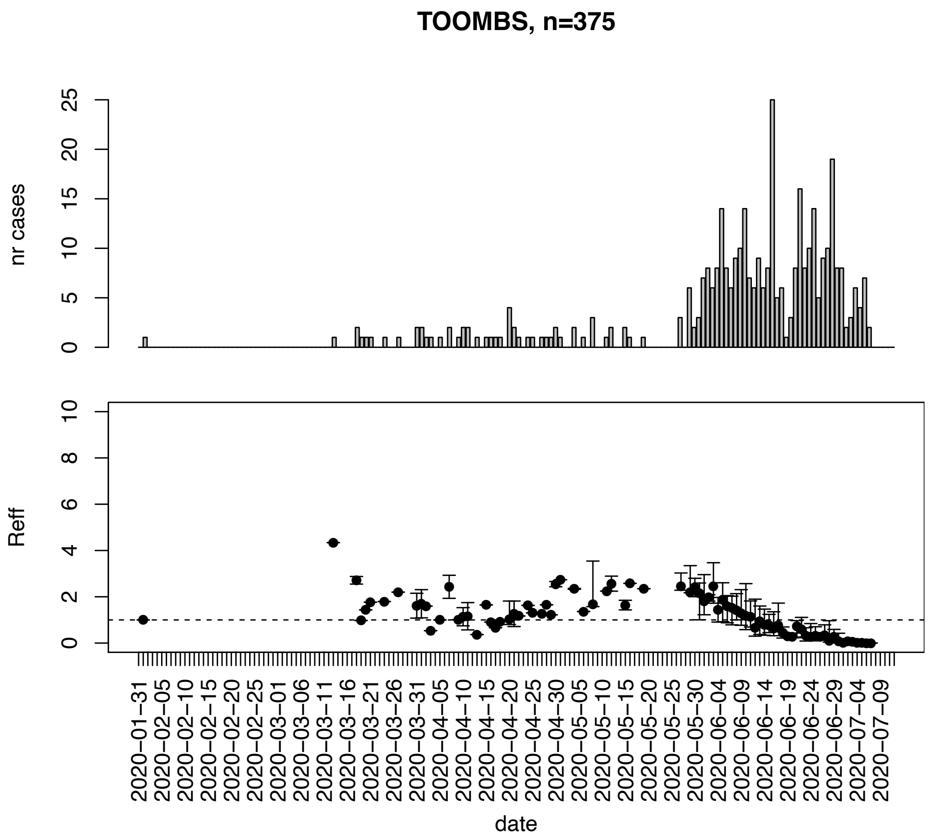


Figure. S138: Epidemic curves and reproduction number estimates until July 13th in Toombs county.

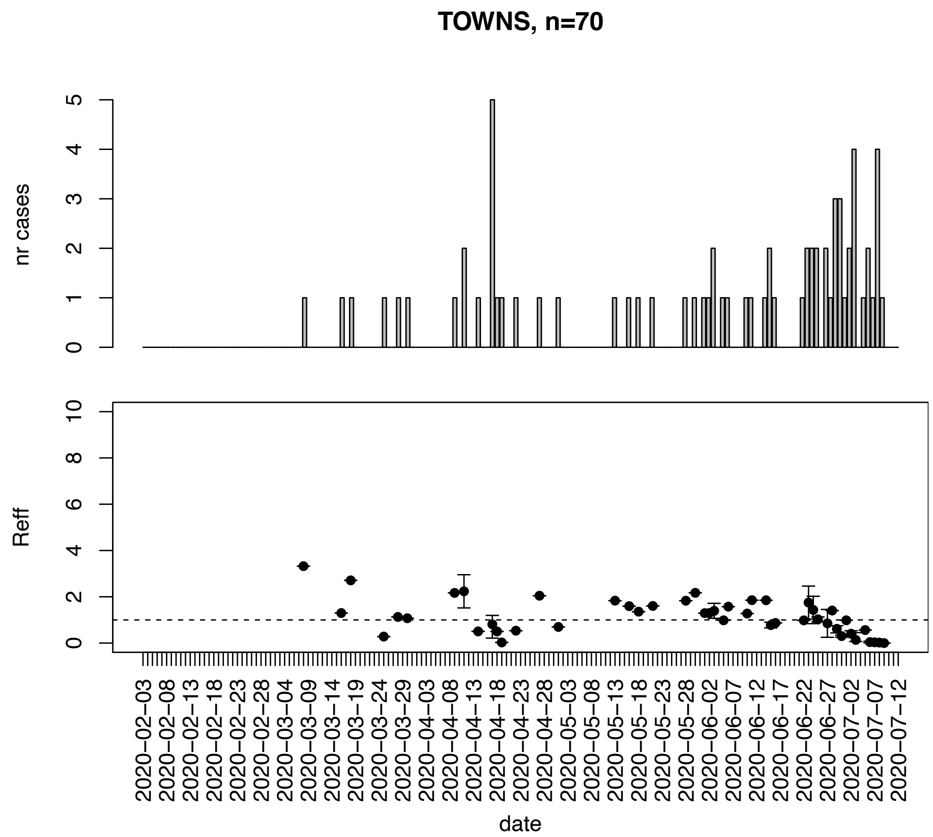


Figure. S139: Epidemic curves and reproduction number estimates until July 13th in Towns county.

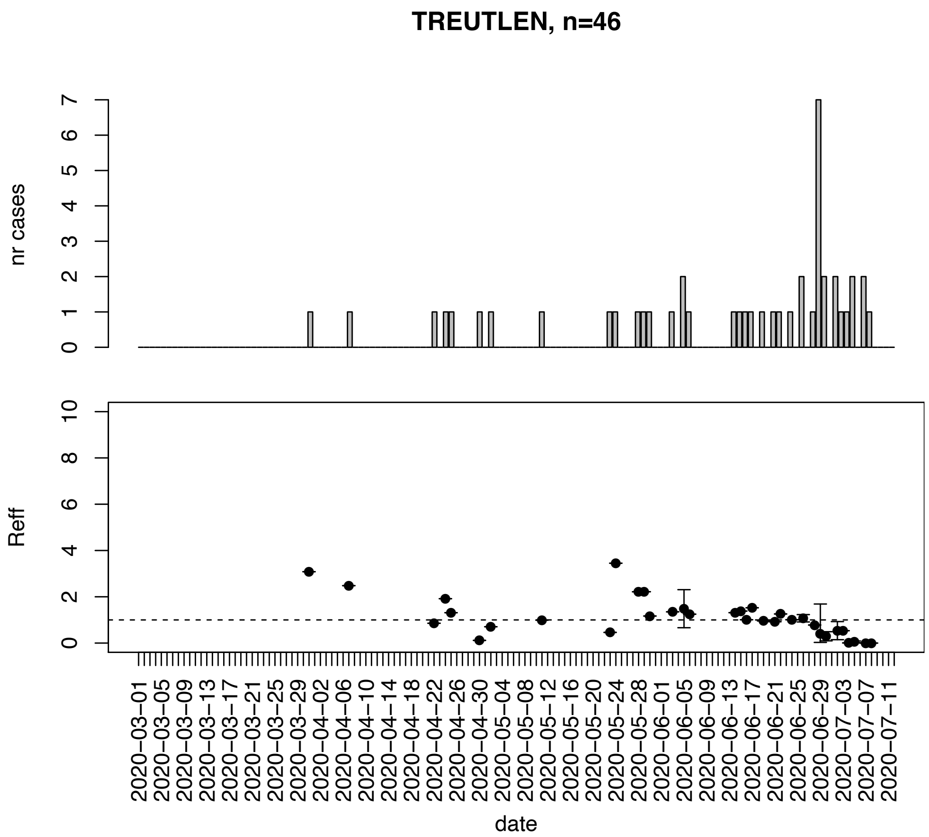


Figure. S140: Epidemic curves and reproduction number estimates until July 13th in Treutlen county.

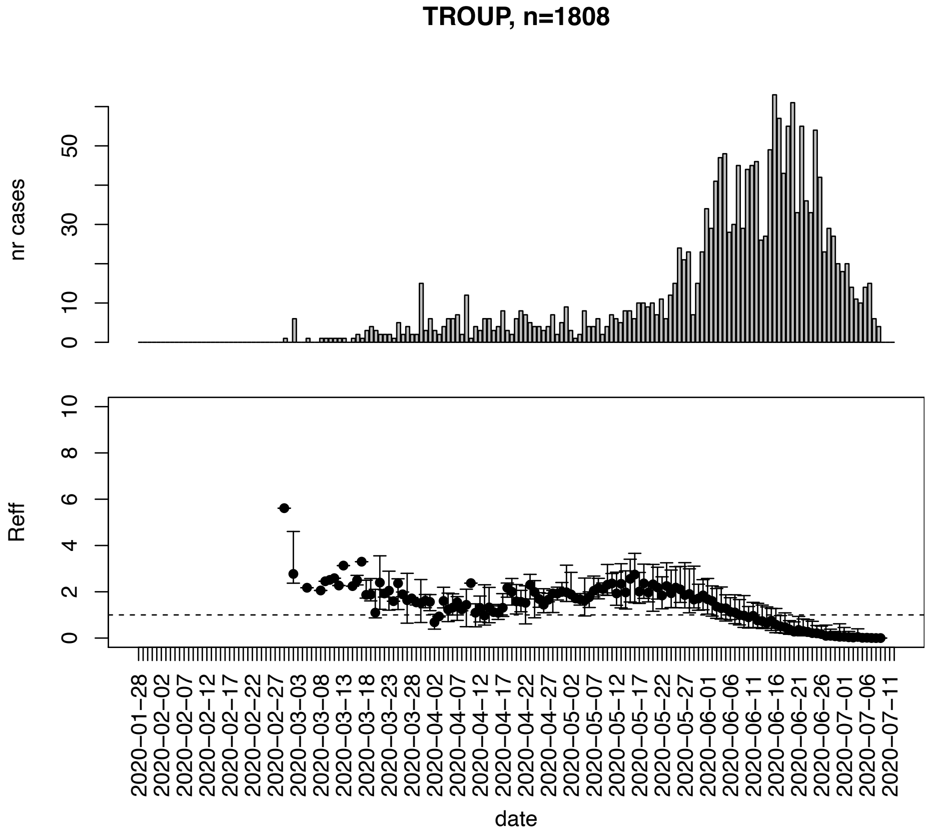


Figure. S141: Epidemic curves and reproduction number estimates until July 13th in Troup county.

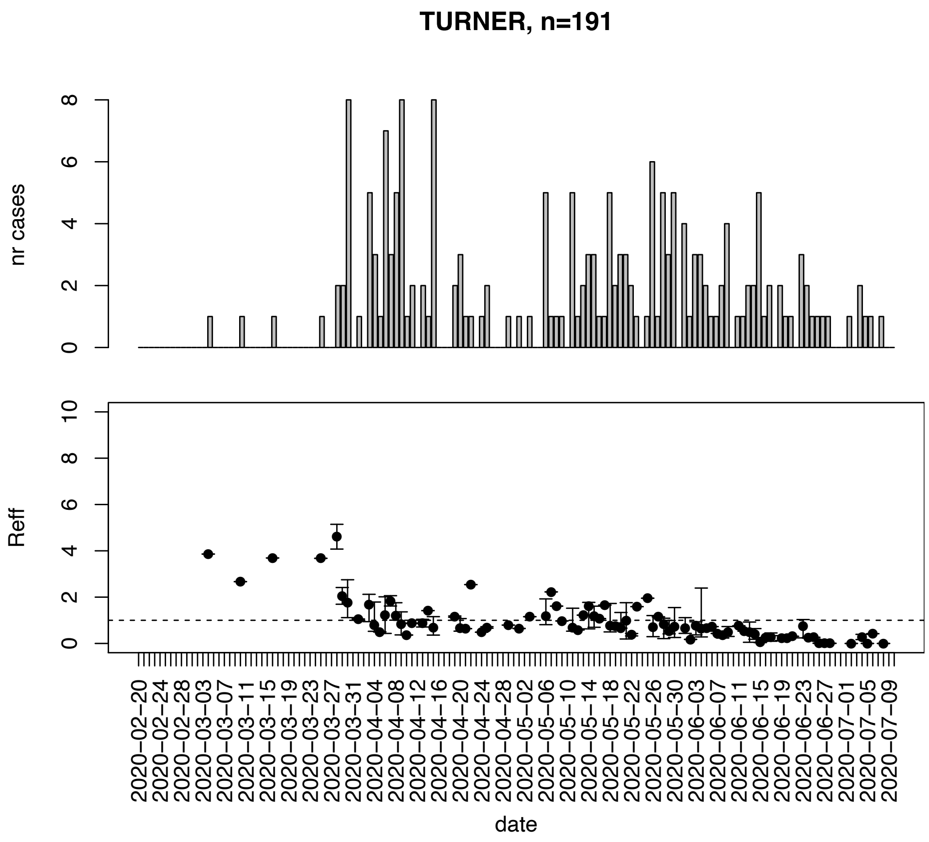


Figure. S142: Epidemic curves and reproduction number estimates until July 13th in Turner county.

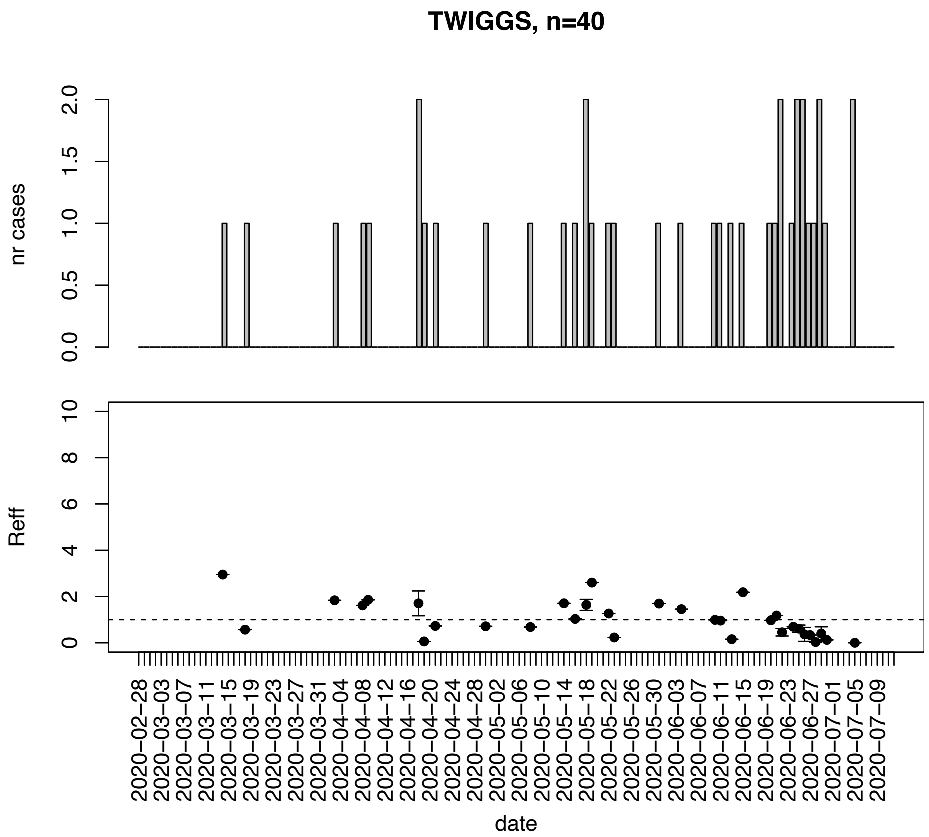


Figure. S143: Epidemic curves and reproduction number estimates until July 13th in Twiggs county.

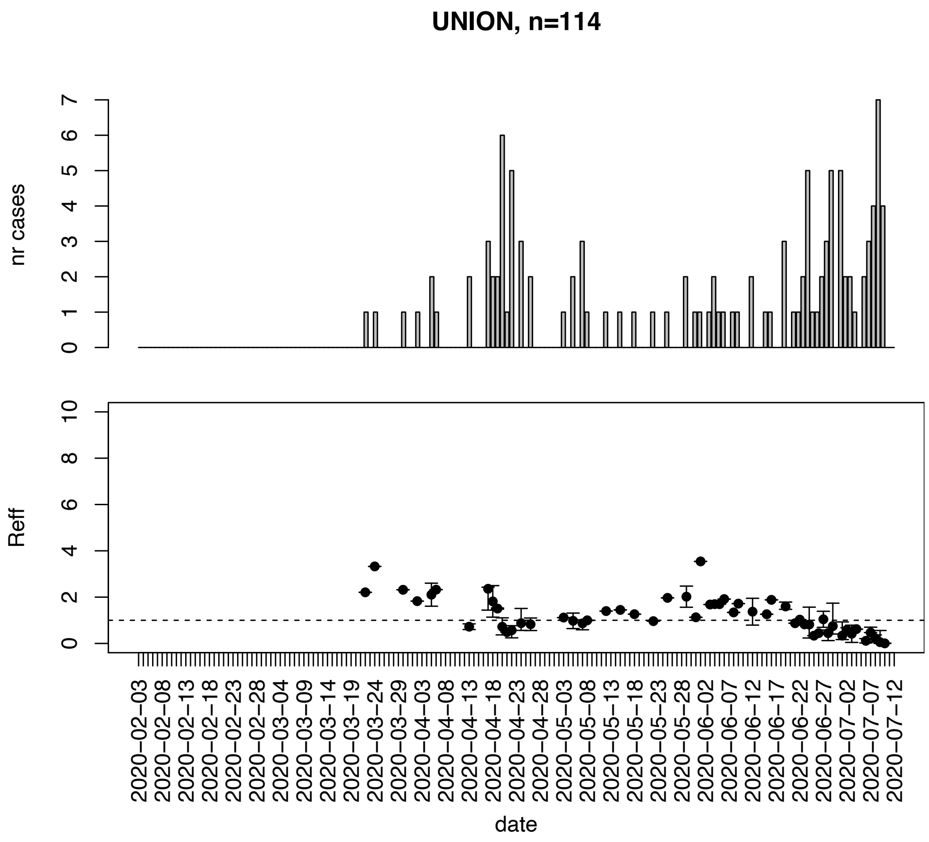


Figure. S144: Epidemic curves and reproduction number estimates until July 13th in Union county.

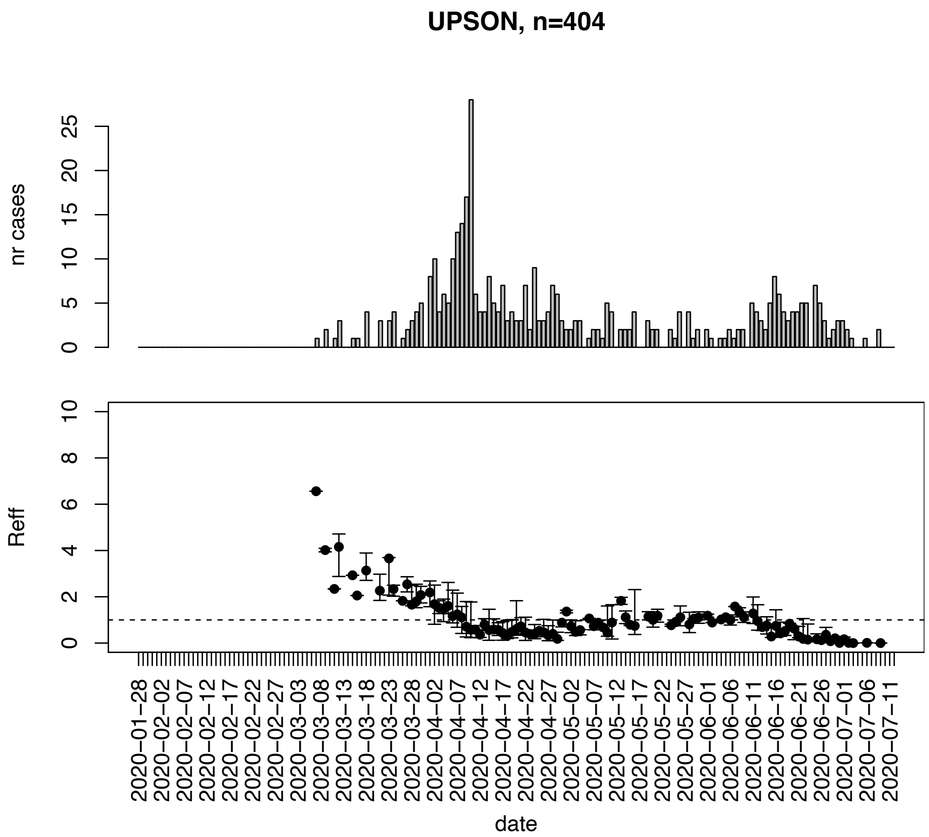


Figure. S145: Epidemic curves and reproduction number estimates until July 13th in Upson county.

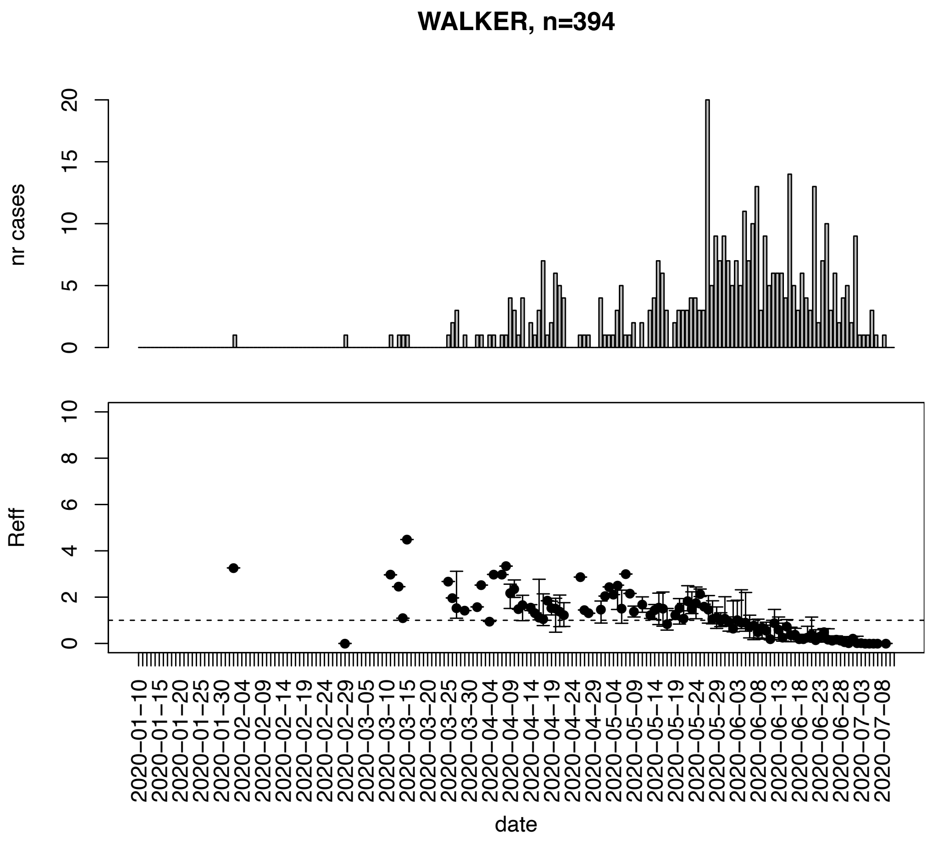


Figure. S146: Epidemic curves and reproduction number estimates until July 13th in Walker county.

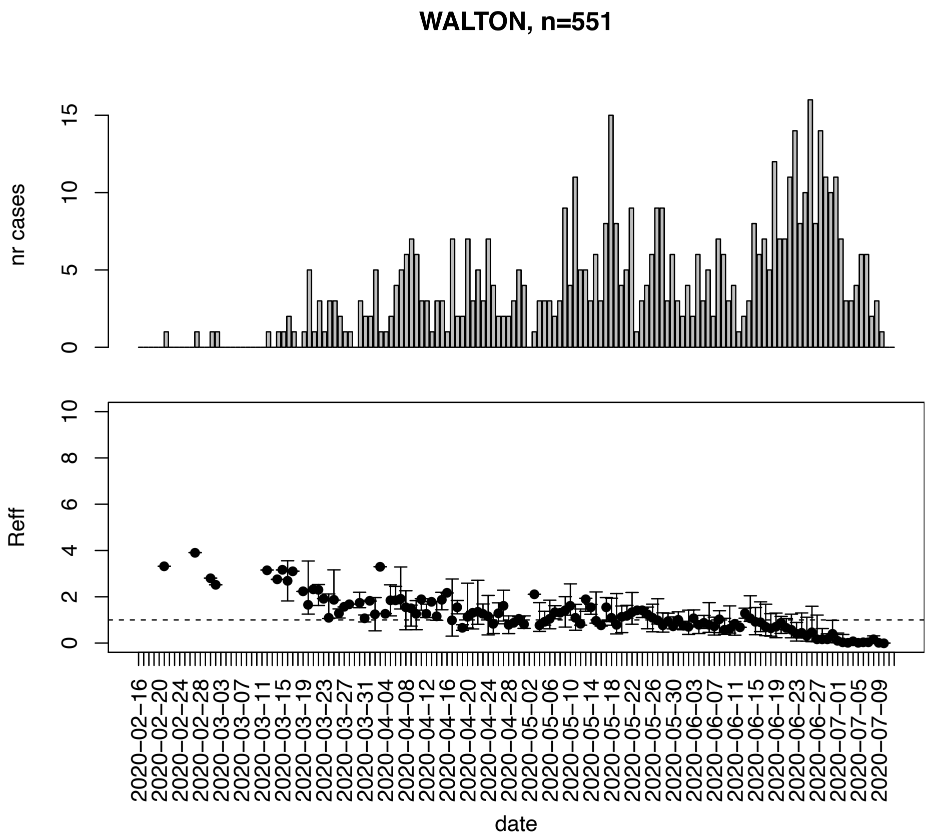


Figure. S147: Epidemic curves and reproduction number estimates until July 13th in Walton county.

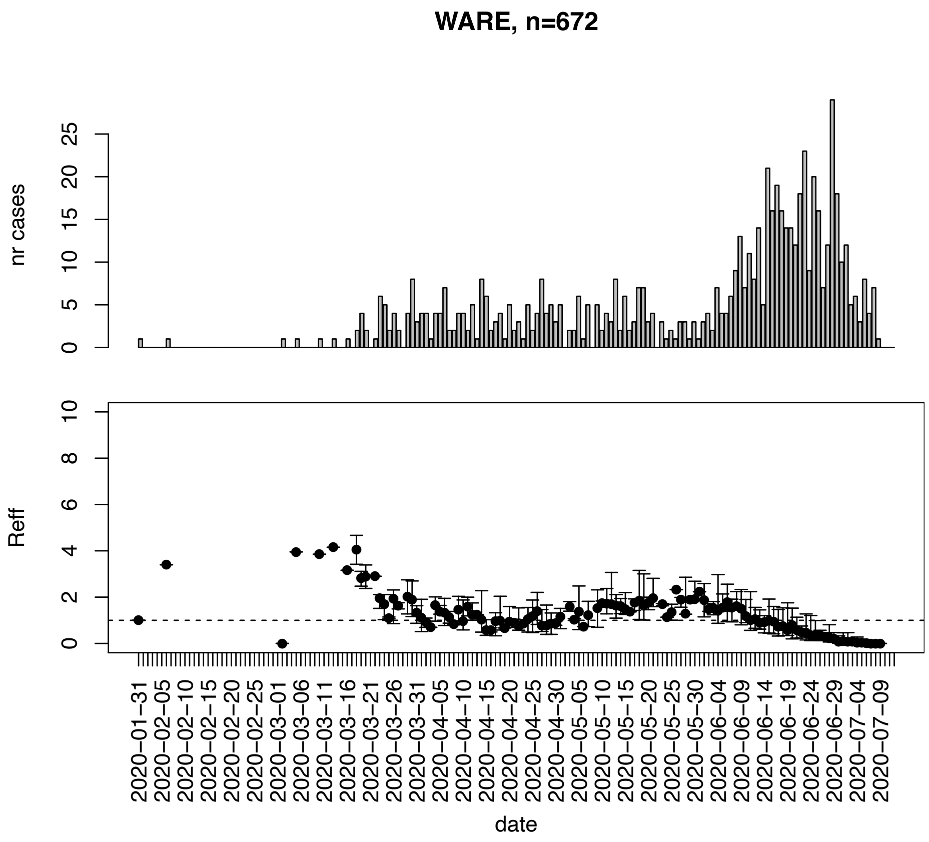


Figure. S148: Epidemic curves and reproduction number estimates until July 13th in Ware county.

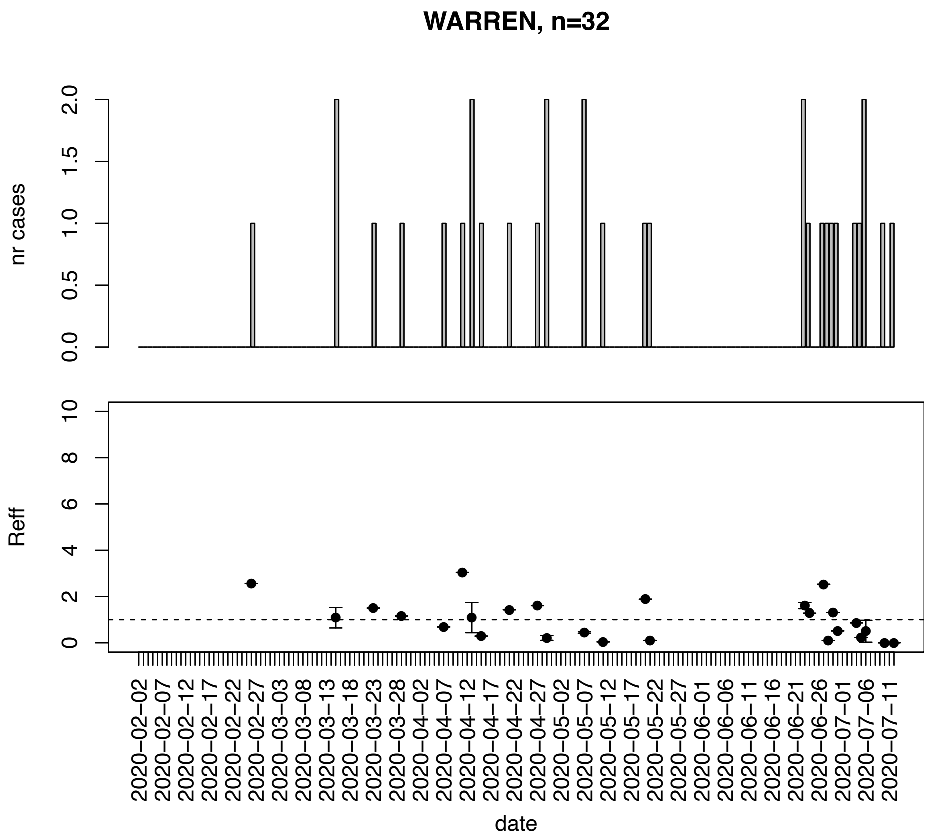


Figure. S149: Epidemic curves and reproduction number estimates until July 13th in Warren county.

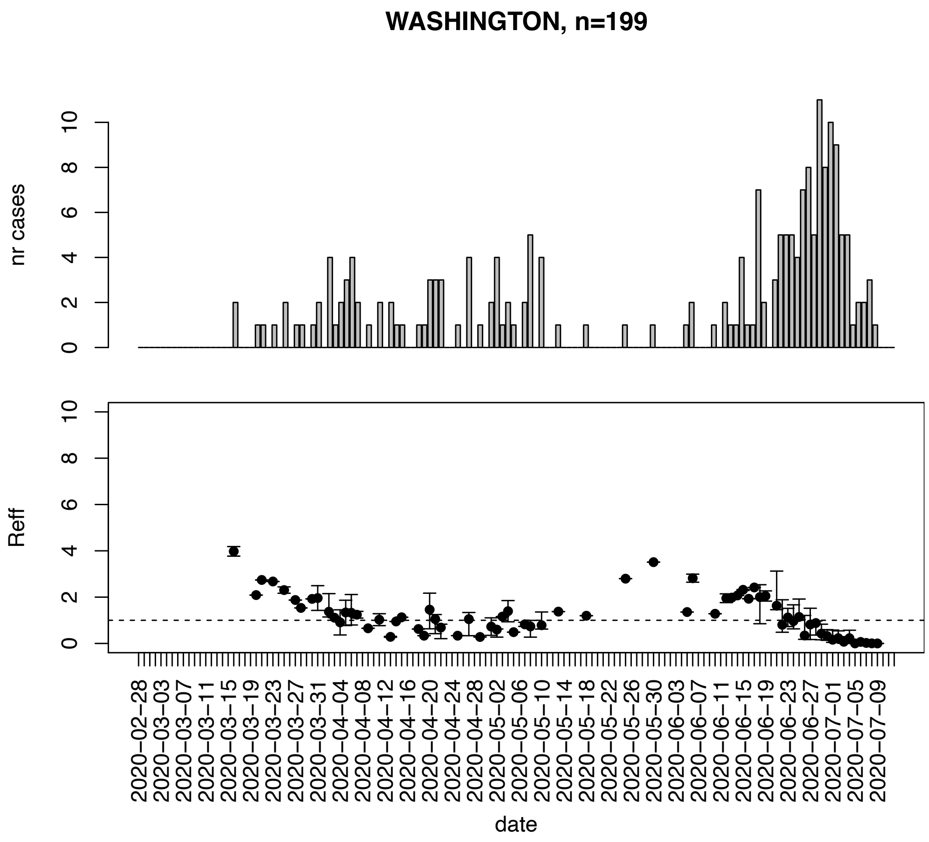


Figure. S150: Epidemic curves and reproduction number estimates until July 13th in Washington county.

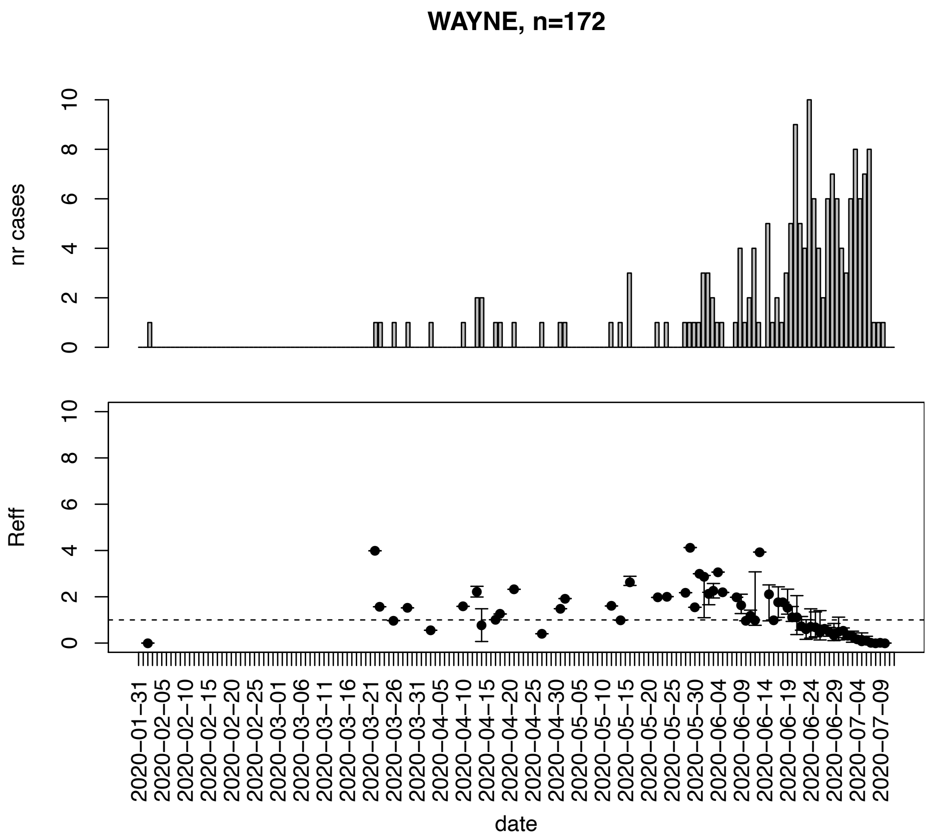


Figure. S151: Epidemic curves and reproduction number estimates until July 13th in Wayne county.

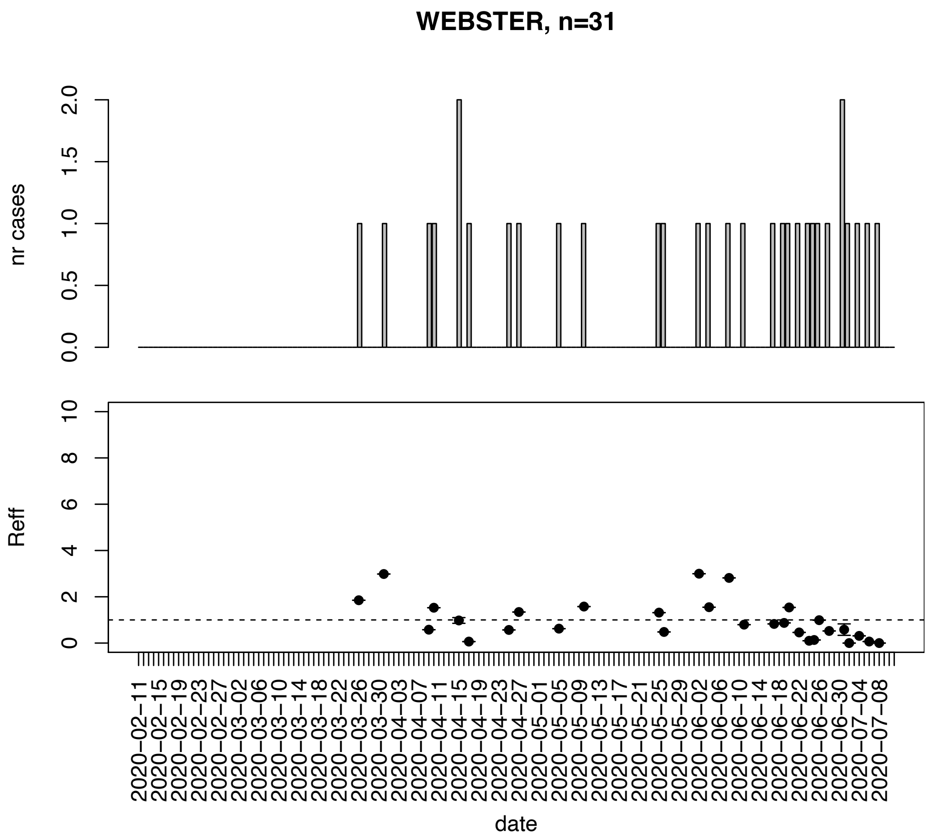


Figure. S152: Epidemic curves and reproduction number estimates until July 13th in Webster county.

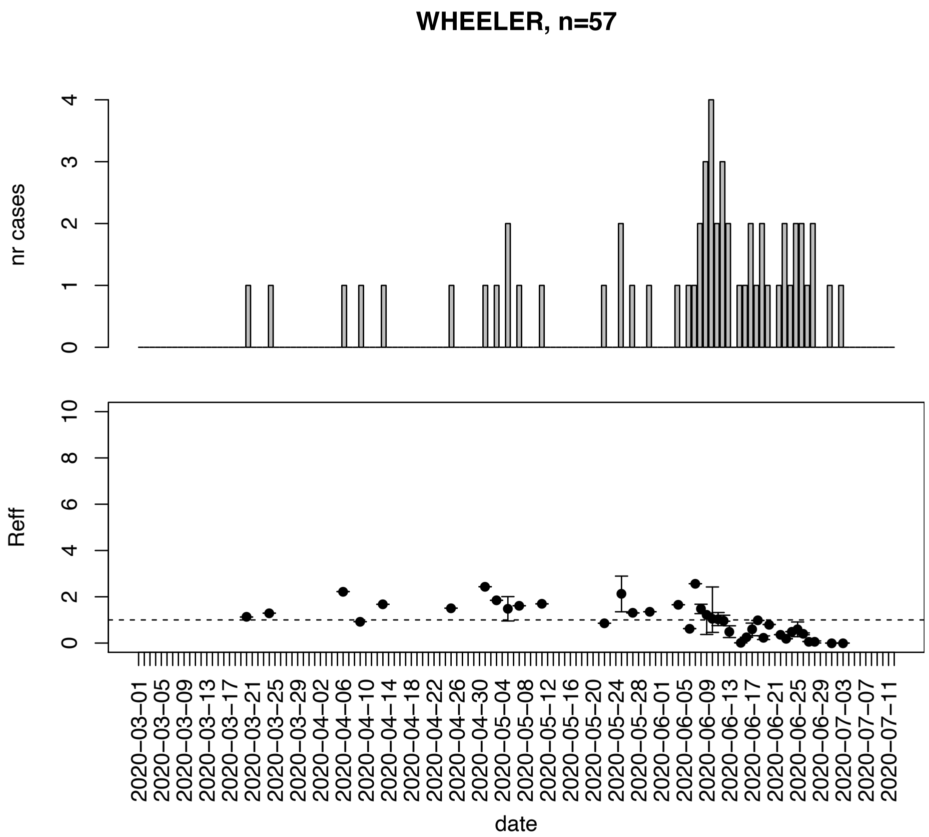


Figure. S153: Epidemic curves and reproduction number estimates until July 13th in Wheeler county.

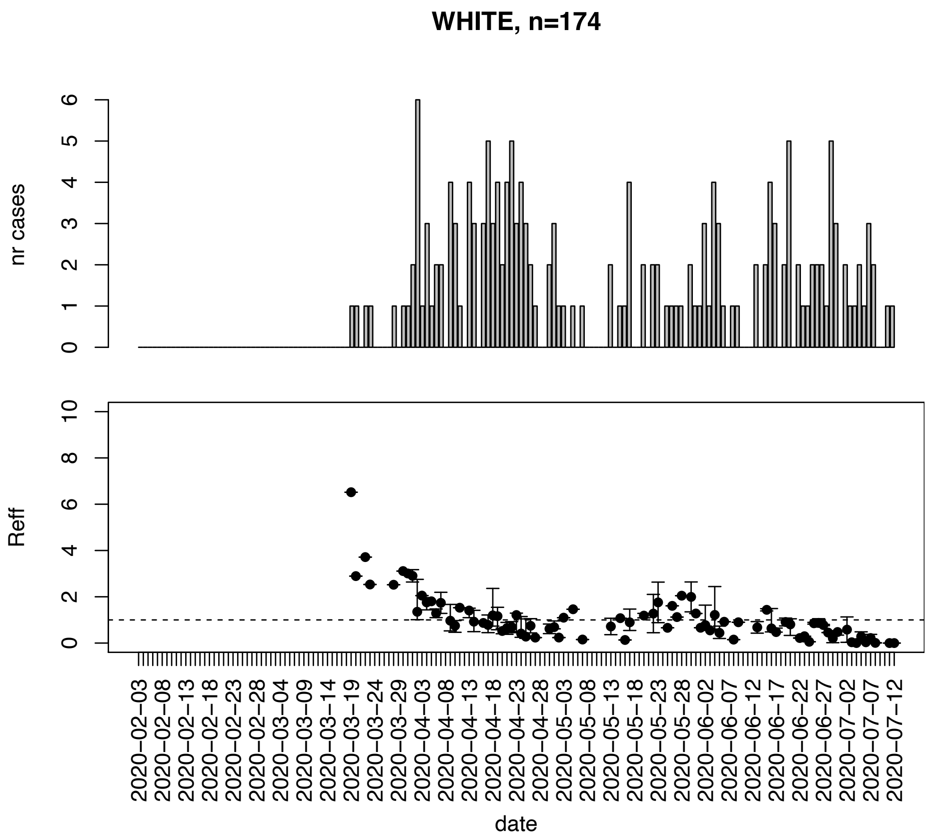


Figure. S154: Epidemic curves and reproduction number estimates until July 13th in White county.

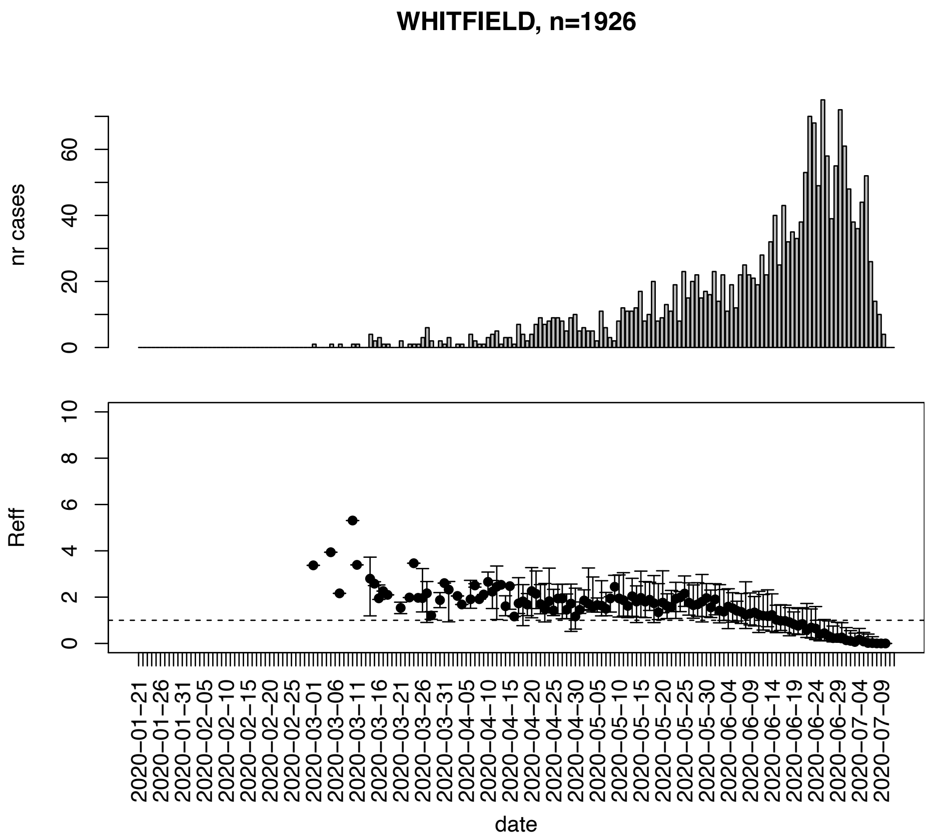


Figure. S155: Epidemic curves and reproduction number estimates until July 13th in Whitfield county.

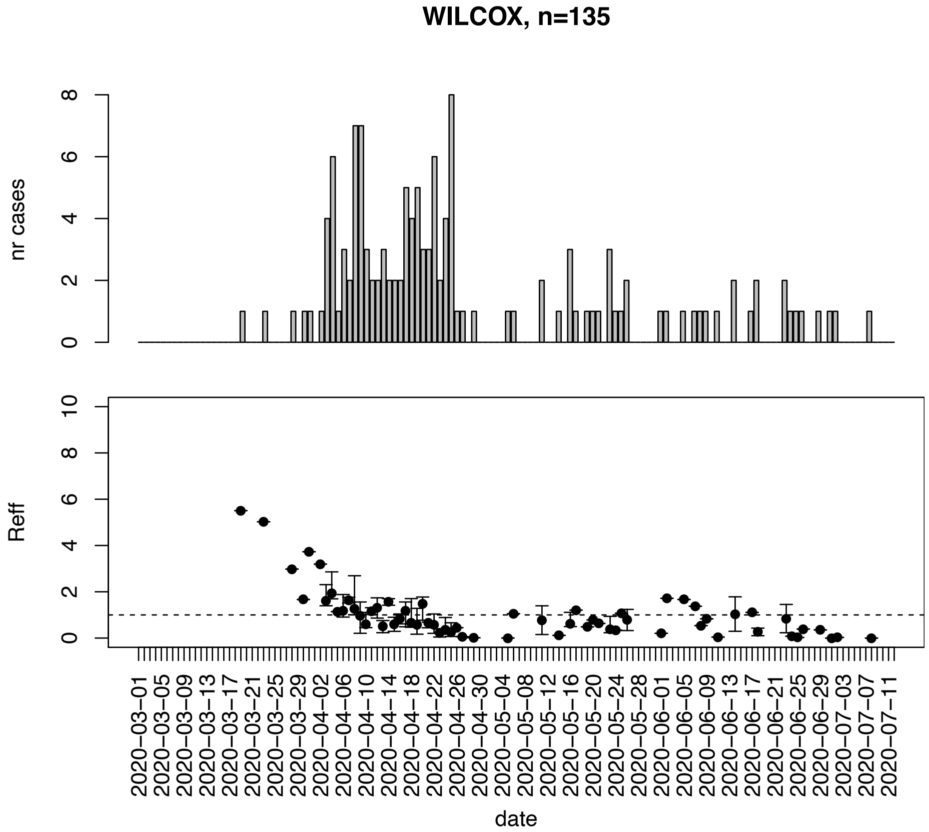


Figure. S156: Epidemic curves and reproduction number estimates until July 13th in Wilcox county.

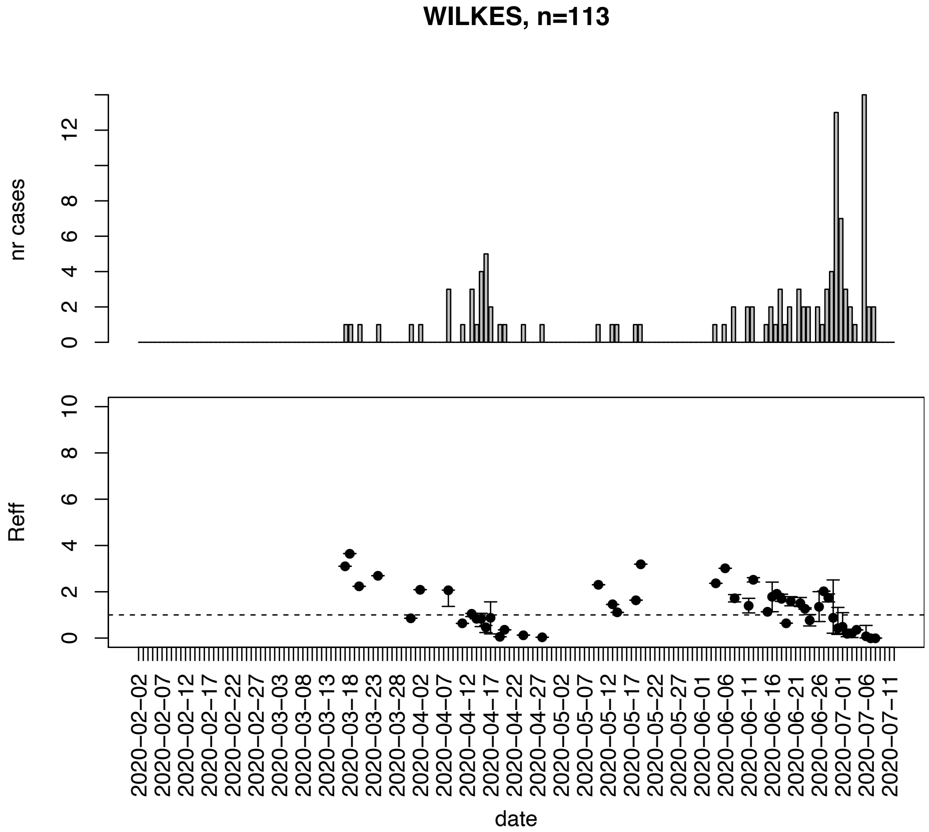


Figure. S157: Epidemic curves and reproduction number estimates until July 13th in Wilkes county.

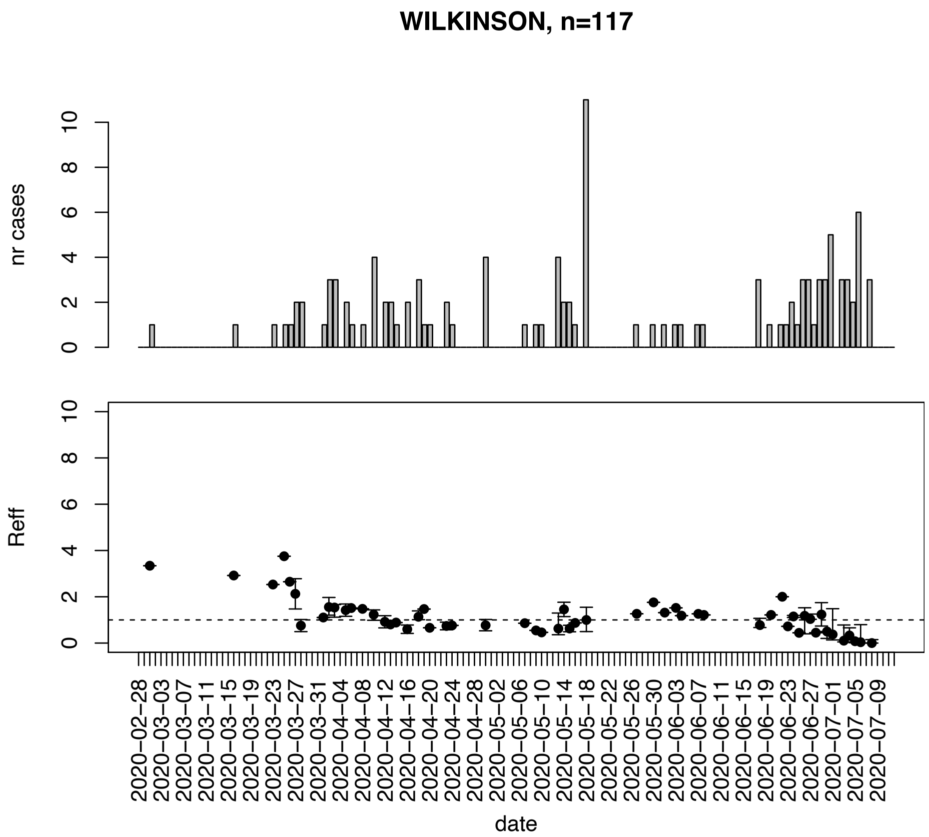


Figure. S158: Epidemic curves and reproduction number estimates until July 13th in Wilkinson county.

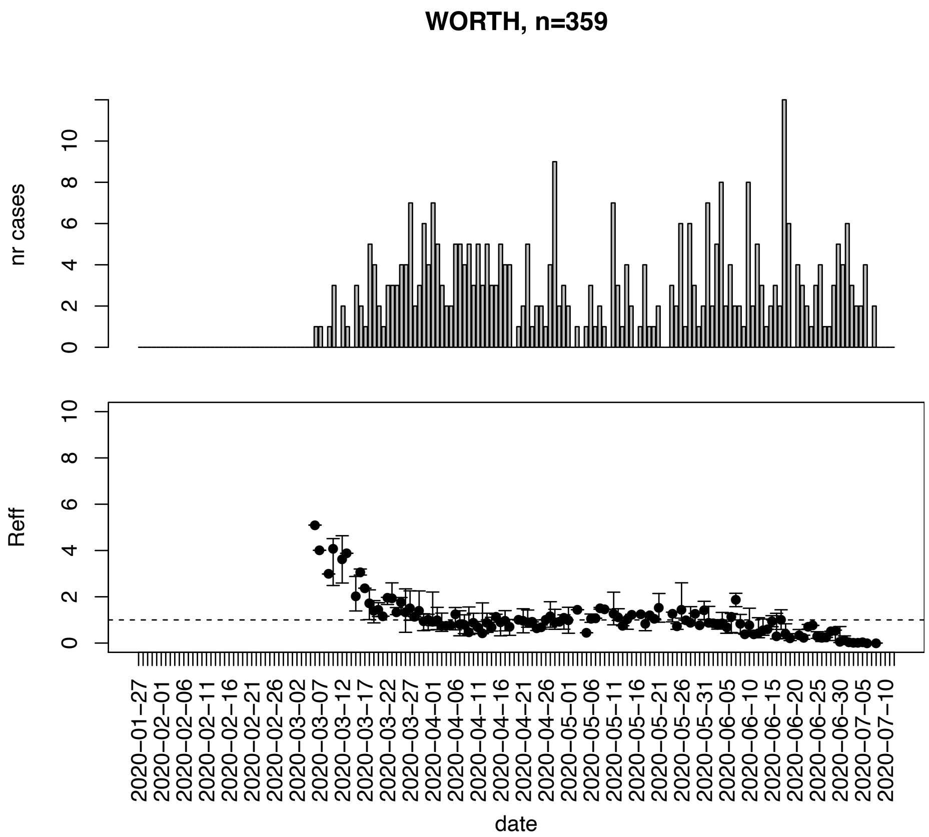


Figure. S159: Epidemic curves and reproduction number estimates until July 13th in Worth county.
